# Recorded Time and Risk-Based Selection for a Second Look Before Emergency Department Discharge

**DOI:** 10.64898/2026.11.18.26368460

**Authors:** Agnideep Aich, Carl Preiksaitis, Christian Rose

**Affiliations:** Department of Emergency Medicine, Stanford University School of Medicine, Stanford, California, USA

## Abstract

**IMPORTANCE:** Emergency department admission plans usually prompt discussion with an accepting team, but discharge decisions do not routinely involve an equivalent discussion, and review must fit limited time and capacity.

**OBJECTIVE:** To determine how much time remains after emergency department discharge decisions and how well simple and model-based rules target limited review capacity.

**DESIGN, SETTING, AND PARTICIPANTS:** Retrospective cohort study at Stanford Health Care’s adult emergency department, with 2020–2021 training, 2022 tuning, a 2023 temporal test set, and a 2024 held-out temporal test set, including adults aged 18–100 years. There were 258,247 eligible encounters in 2020–2023 and 77,086 in 2024.

**EXPOSURES:** Recorded time from the decision to reversal, handoff, departure, or clinical decision unit entry, capped at 60 minutes. Simulations used fixed 2022 thresholds and daily caps of 6 discharge and 3 admission 10-minute reviews.

**MAIN OUTCOMES AND MEASURES:** The composite comprised 7-day return ending in admission, qualifying critical care within 24 hours, or death within 7 days. Event reach was the proportion of event encounters selected with sufficient recorded time. Replication was prespecified as a Risk-only minus Prior-visits-first difference with a 95% lower bound above 0.

**RESULTS:** The 2024 encounters involved 51,587 patients (median encounter age, 53 years [IQR, 35–70]; 42,333 encounters [54.9%] involving female patients). Events occurred in 1,066 of 50,675 discharge encounters and 1,125 of 26,411 admission encounters. Discharge windows were at least 10 minutes in 84.5% overall and 86.4% with events. Discharge event reach was 3.2% with Random, 5.9% with Emergency Severity Index–first, 15.8% with Prior-visits-first, and 19.4% with Risk-only selection. The Risk-only minus Prior-visits-first difference was 3.66 percentage points (nominal 95% CI, −0.18 to 6.26; empirical coverage, 93.4%), not meeting the replication criterion; 2023 reach was 19.5% versus 16.0%.

**CONCLUSIONS AND RELEVANCE:** Most discharge decisions had a recorded window of at least 10 minutes. For discharges, selection by prior visits or predicted risk reached several times more events than random or triage-based selection, and the model did not reliably outperform prior visits. Prospective studies should test whether brief, targeted review improves outcomes.

**Key Points:** *Question:* After an emergency department discharge decision, is there time for a second look, and how should limited review capacity be targeted?

*Findings:* In this cohort study’s 2024 held-out temporal test set (77,086 encounters), 84.5% of discharge decisions had at least 10 recorded minutes. Event reach was 3.2% with random selection, 5.9% with Emergency Severity Index–first, 15.8% with prior visits, and 19.4% with a risk model; the model’s advantage was not statistically significant and did not meet the replication criterion.

*Meaning:* A prior-visit rule reached about 80% as many events as the model; clinical benefit requires prospective testing.

## Introduction

In 2020–2023, 63% of candidate adult emergency department (ED) encounters at our institution were assigned to the discharge decision-time group. In local practice, admission plans usually prompt discussion with an accepting team. Discharge decisions do not routinely require an equivalent discussion. In a Canadian study, physicians reported relying on clinical judgment for 87.6% of discharges from high-acuity ED areas.^1^ Adverse events follow some discharges, and some ward admissions require intensive care within 24 hours,^2,3^ although neither outcome shows that the initial decision was wrong.

A second clinician’s review can change these decisions: in one UK ED, senior review changed approximately 28% of junior clinicians’ disposition plans.^4^ Delivering review reliably is harder. In the French CHARMED trial, 32% of intervention-group patients were never cross-checked, some possibly because they left between scheduled sessions,^5^ and a UK national audit found consultant or associate-specialist review for 35% of adults in specified high-risk discharge groups and recommended electronic prompts.^6^

With limited capacity, the practical question is which discharges to review. Prediction models estimate short-term outcomes after discharge, return admissions, and escalation after admission,^7–9^ but in the Dutch MARS-ED trial, physicians reported score-related reassessment for 1 of 644 intervention patients.^10^ A model could instead choose which encounters receive another clinician’s review, with its value depending on available capacity and the time remaining before the patient leaves.^11,12^

Boarding research and a federal quality measure describe time after admission decisions,^13,14^ but little is known about time after discharge decisions or how to allocate review. We asked 2 practical questions: whether enough time remains after an attending physician’s discharge decision for a brief second look, and how limited review capacity should be targeted, comparing simple rules with a prediction model. Admission plans served as a contrast, the 2024 held-out temporal test set was analyzed under a prespecified plan, and no reviews were delivered.

## Methods

### Study Design and Population

This retrospective cohort study included Stanford Health Care adult ED encounters during 2020–2024, restricted to recorded ages 18–100 years. Ages above 100 were excluded because they frequently reflect placeholder values. ED records were linked to hospital admission, discharge, and transfer (ADT) records and death dates from the Stanford Medicine Research Data Repository (STARR). The 2020–2023 sources represented September 30, 2026 (UTC); the 2024 sources represented November 6, 2026 (UTC). Vital signs for a 2023 sensitivity analysis came from an October 2026 extraction. Repeat encounters were retained. Eligibility required resolved identity, a qualifying decision, a measurable window, and a known outcome. Reporting followed Strengthening the Reporting of Observational Studies in Epidemiology (STROBE), REporting of studies Conducted using Observational Routinely-collected health Data (RECORD), and Transparent Reporting of a multivariable prediction model for Individual Prognosis Or Diagnosis with artificial intelligence (TRIPOD+AI).^15–17^ The Stanford University Institutional Review Board approved the study (protocol 69107) and waived informed consent.

### Recorded Decisions and Windows

Discharge time zero was the earliest qualifying readiness or conditional-discharge marker, even after an earlier admission plan. In local workflow, these attending-only markers document the attending physician’s discharge decision. Conditional discharge delegates completion to the nurse once specified criteria are met. The individual user was not analyzed. Discharge markers after the first recorded handoff did not qualify. Otherwise, the earliest qualifying admission-plan marker set time zero. A decision to admit is provisional and may be recorded by a resident or attending. The admission order is usually signed immediately after discussion with the accepting team; its timestamp may approximate that discussion, whose actual timing was unmeasured. Final disposition did not redefine groups (eMethods; eTable 1; eTable 2; eTable 3).

Markers had to fall between arrival and departure or earlier clinical decision unit (CDU) entry. The CDU provides ED-based observation, distinct from hospital admission or hospital observation; admission plans ending there remained in the admission group. Untimed or out-of-interval markers were disregarded; conflicting directions at discharge time zero were excluded. Windows ended at reversal, recorded handoff, departure, or CDU entry, capped at 60 minutes. Valid zero windows remained eligible. Missing handoffs used departure or CDU entry as a flagged fallback. Reversals did not themselves constitute outcomes (eTable 4).

### Electronic Outcomes

The composite comprised adult ED return arriving within 7 days and ultimately ending in inpatient admission, qualifying critical care within 24 hours, or death within 7 days of time zero. Follow-up continued beyond the review window. Returns had to follow ED departure and hospital discharge when recorded. Admission-plan critical care required progression from acute or intermediate care; discharge-group events could occur during the index encounter or a return. Baseline critical care did not count. Death required the entire recorded day to lie after time zero and within 7 days. Any positive component established an event; otherwise, an unknown component made the composite unknown (eTable 5; eTable 6).

### Prediction Models

Arrival years defined training (2020–2021), tuning (2022), the temporal test set (2023), and the held-out temporal test set (2024) (eFigure 1). Decisions had to fall within their assigned period; training and tuning encounters were excluded if 7-day follow-up reached the next period. Patients could contribute across periods.

Predictors included age, decision type and timing, prior completed adult ED visits within 90 days, available history, laboratory counts and results, Emergency Severity Index (ESI), arrival mode, and chief complaint. Laboratory orders and numeric results required availability by time zero (eTable 7; eTable 8; eTable 9; eFigure 2). Triage recording times were unavailable; availability was assumed. Recorded sex, race, ethnicity, language, and interpreter need described the cohort and subgroups, without entering prediction models.

Separate models compared ridge logistic regression and gradient-boosted trees, using training-only preprocessing, 2022 log-loss selection, and 2022 intercept recalibration (eMethods; 2023 calibration in eFigure 3). A 2023 sensitivity added six first recorded vital signs between arrival and time zero; missing or invalid first values remained missing (eTable 10; eTable 11). Models, preprocessing, thresholds, and capacities were applied unchanged in 2024, without refitting or recalibration.

### Review Selection

Risk-only prioritized predicted event risk. Risk-plus-window multiplied risk by the conditional probability of sufficient time given an event, estimated from training and tuning events (eTable 12; eTable 13; eTable 14). ESI-first prioritized levels 1–5; Age-first prioritized older age; Prior-visits-first prioritized more prior visits. Missing values ranked last. Simple logistic used age, arrival-to-decision time, prior visits, ESI, and arrival mode.

Sequential thresholds flagged the highest-scoring 5% of 2022 encounters for 10-minute reviews and 10% for 5-minute reviews. Ties used fixed seeded random numbers; Random flagged by those numbers alone. Flagged encounters were selected in decision order, with simultaneous decisions sharing remaining capacity fractionally. Daily capacities, derived from 2022 volume and 50 review-minutes per 100 encounters, were 3 admission and 6 discharge reviews for 10-minute reviews, doubled for 5-minute reviews. Capacity reset at midnight without carryover and applied over all 365 days in 2023 and 366 in 2024. Sensitivities used a 10% flag rate for 10-minute reviews and start delays of 5 or 10 minutes (eTable 15; eTable 16; eTable 17; eTable 18).

Event reach was the proportion of composite-event encounters selected with enough recorded time for delay plus review duration. Attempts without sufficient time consumed capacity. Attempts per event reached did not measure events prevented. Staffing and queues were not modeled. Full-day rankings using later decisions and annual pooling were retrospective 2023 benchmarks (eTable 19–eTable 21).

### Statistical Analysis

Random, Risk-only, and Risk-plus-window selection, the 5-, 10-, and 15-minute review durations, and the 60-minute cap were specified before any test-year evaluation; budgets, full-day allocation, and window-model details were finalized after initial 2023 results. Post hoc 2020–2023 analyses included the simpler comparators, sequential allocation, recorded transitions, subgroup comparisons, window-model and interval diagnostics, the vital-sign sensitivity analysis, and the unknown-outcome comparison. Recorded transitions compared first resident-plan markers with the next and last qualifying statuses and final disposition; they neither identify nor bound attending overrides (eTable 22–eTable 26).

The decision-time definition was revised on clinical workflow grounds after initial 2023 analyses. The 2024 primary analyses and replication criterion were then specified in an analysis plan posted to the Open Science Framework before extraction and since registered (https://osf.io/4sjne), and 2024 was analyzed once with models, thresholds, and capacities fixed. The primary 2024 analyses were descriptive discharge-window sufficiency at 10 minutes and Risk-only minus Prior-visits-first reach with 10-minute reviews, 5% tuning thresholds, and zero delay. The replication criterion required a difference above 0 with a 95% lower bound above 0. Patient-cluster bootstrap intervals used 2,000 paired resamples, with fitted models fixed and selection repeated; at least 1,900 finite estimates were required. Nested diagnostics used 1,000 outer and 2,000 inner samples to assess containment of each year’s full-cohort estimate, not population coverage. Single-rule reach was descriptive; paired intervals were nominal and reported with empirical coverage (eTable 27; eTable 28; eTable 29). Counts of 1–10 and selected cells were withheld. Subgroups required at least 20 events and 11 encounters. Window-association tests were 2-sided. Software versions are in eMethods.

## Results

### Study Population and Windows

In 2020–2023, 258,247 of 284,627 candidates were eligible: 119,662 training, 65,871 tuning, and 72,714 temporal test encounters. In 2024, 77,086 of 83,002 candidates were eligible, involving 51,587 patients (Figure 1; eTable 30). Median encounter age in 2024 was 53 years (IQR, 35–70), and 42,333 encounters (54.9%) involved female patients. ESI level 4 decreased from 22.8% in training to 11.3% in 2023 and 10.5% in 2024 (Table 1; eTable 31; eTable 32; eTable 33).

**Figure 1.**
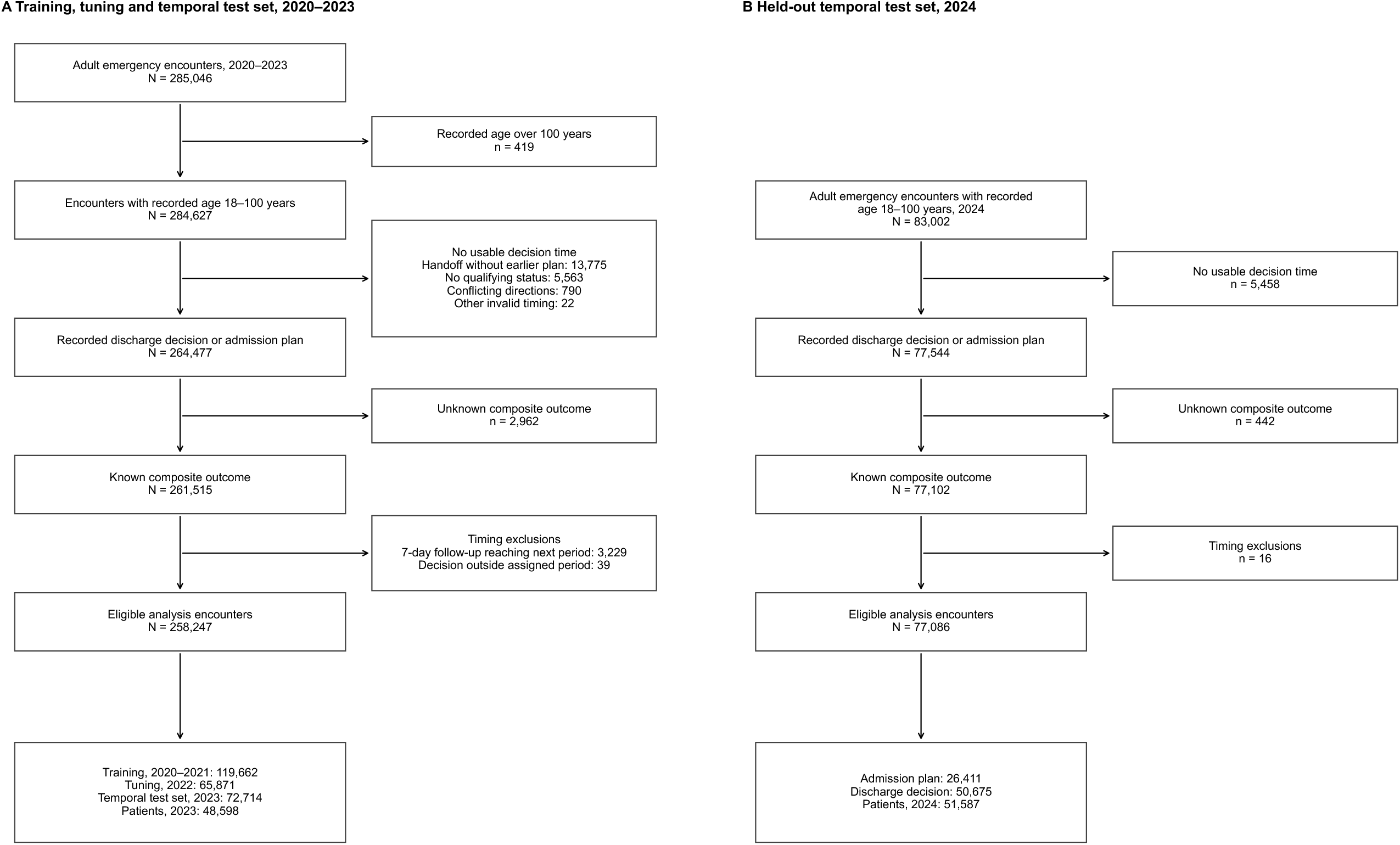
Study Population. Counts are encounters unless patients are specified. A, 2020–2023: exclusions are sequential: recorded age above 100 years, decision time, outcome ascertainment, then temporal eligibility. Unknown outcomes are excluded before timing exclusions. The 7–day boundary rule applies to training and tuning encounters. B, 2024: candidates had recorded ages 18–100 years. Decision-time and timing exclusions are shown as totals. Follow-up could extend into January 2025. Patients may contribute to more than one period.

**Table 1.** Characteristics of Eligible ED Encounters.

| Characteristic | 2020–2021 | 2022 | 2023 | 2024 |
|---|---|---|---|---|
| **Cohort** |  |  |  |  |
| Encounters | 119,662 | 65,871 | 72,714 | 77,086 |
| Unique patients | 72,980 | 44,477 | 48,598 | 51,587 |
| **Age, y** |  |  |  |  |
| Median (IQR) | 49.0 (33.0–66.0) | 52.0 (34.0–69.0) | 52.0 (34.0–70.0) | 53.0 (35.0–70.0) |
| Available | 119,662 (100.0) | 65,871 (100.0) | 72,714 (100.0) | 77,086 (100.0) |
| Missing | 0 (0.0) | 0 (0.0) | 0 (0.0) | 0 (0.0) |
| **Sex** |  |  |  |  |
| Female | 63,276 (52.9) | 36,066 (54.8) | 40,252 (55.4) | 42,333 (54.9) |
| Male, non-binary or unknown sex | 56,386 (47.1) | 29,805 (45.2) | 32,462 (44.6) | 34,753 (45.1) |
| Missing | 0 (0.0) | 0 (0.0) | 0 (0.0) | 0 (0.0) |
| **Race** |  |  |  |  |
| American Indian or Alaska Native | 287 (0.2) | 164 (0.2) | 181 (0.2) | 224 (0.3) |
| Asian | 17,151 (14.3) | 10,760 (16.3) | 12,317 (16.9) | 13,040 (16.9) |
| Black or African American | 7,518 (6.3) | 4,112 (6.2) | 4,484 (6.2) | 4,634 (6.0) |
| Multiple recorded labels | 3,305 (2.8) | 2,021 (3.1) | 2,201 (3.0) | 2,206 (2.9) |
| Native Hawaiian or Other Pacific Islander | 2,612 (2.2) | 1,328 (2.0) | 1,479 (2.0) | 1,496 (1.9) |
| Other | 40,986 (34.3) | 21,096 (32.0) | 22,877 (31.5) | 23,961 (31.1) |
| White | 46,471 (38.8) | 25,550 (38.8) | 27,904 (38.4) | 29,209 (37.9) |
| Declines to state | 1,206 (1.0) | 740 (1.1) | 1,109 (1.5) | 2,094 (2.7) |
| Missing | 49 (0.0) | 48 (0.1) | 118 (0.2) | 148 (0.2) |
| Unknown | — | — | — | — |
| **Ethnicity** |  |  |  |  |
| Hispanic/Latino | 34,847 (29.1) | 19,126 (29.0) | 21,762 (29.9) | 23,318 (30.2) |
| Non-Hispanic/non-Latino | 83,682 (69.9) | 46,078 (70.0) | 49,962 (68.7) | 52,032 (67.5) |
| Declines to state | 848 (0.7) | 510 (0.8) | 770 (1.1) | 1,418 (1.8) |
| Missing | 45 (0.0) | 27 (0.0) | 110 (0.2) | 143 (0.2) |
| Unknown | 240 (0.2) | 130 (0.2) | 110 (0.2) | 175 (0.2) |
| **Language** |  |  |  |  |
| English | 97,755 (81.7) | 52,914 (80.3) | 57,088 (78.5) | 60,270 (78.2) |
| Farsi | 539 (0.5) | 256 (0.4) | 306 (0.4) | — |
| Mandarin | 1,429 (1.2) | 990 (1.5) | 1,216 (1.7) | 1,213 (1.6) |
| Spanish | 15,312 (12.8) | 8,623 (13.1) | 10,317 (14.2) | 11,248 (14.6) |
| Vietnamese | 1,118 (0.9) | 851 (1.3) | 1,093 (1.5) | 1,072 (1.4) |
| Other or missing | 3,509 (2.9) | 2,237 (3.4) | 2,694 (3.7) | 2,911 (3.8) |
| Unknown | 0 (0.0) | 0 (0.0) | 0 (0.0) | — |
| **Interpreter need** |  |  |  |  |
| Not required | 100,112 (83.7) | 54,331 (82.5) | 58,885 (81.0) | 62,127 (80.6) |
| Required | 19,535 (16.3) | 11,521 (17.5) | 13,757 (18.9) | 14,842 (19.3) |
| Missing | 15 (0.0) | 19 (0.0) | 72 (0.1) | 117 (0.2) |
| **ESI** |  |  |  |  |
| 1 | 884 (0.7) | 413 (0.6) | 407 (0.6) | 396 (0.5) |
| 2 | 22,077 (18.4) | 15,054 (22.9) | 18,346 (25.2) | 21,016 (27.3) |
| 3 | 66,762 (55.8) | 39,961 (60.7) | 44,760 (61.6) | 46,678 (60.6) |
| 4 | 27,239 (22.8) | 9,517 (14.4) | 8,250 (11.3) | 8,110 (10.5) |
| 5 | — | — | — | — |
| Missing | — | — | — | — |
| **Arrival mode** |  |  |  |  |
| Ambulance, American Medical Response | 12,145 (10.1) | 7,647 (11.6) | 9,433 (13.0) | 10,669 (13.8) |
| Ambulance, Palo Alto Paramedics | 3,777 (3.2) | 2,228 (3.4) | 2,034 (2.8) | 1,830 (2.4) |
| Ambulance | 1,544 (1.3) | 685 (1.0) | 902 (1.2) | 1,496 (1.9) |
| Car/bus | 97,259 (81.3) | 52,698 (80.0) | 57,872 (79.6) | 60,842 (78.9) |
| Walk | 1,094 (0.9) | 737 (1.1) | 828 (1.1) | 979 (1.3) |
| Other or missing | 3,843 (3.2) | 1,876 (2.8) | 1,645 (2.3) | 1,270 (1.6) |

Values are No. (%) unless indicated. Periods are training (2020–2021), tuning (2022), the temporal test set (2023), and the held-out temporal test set (2024). Repeat encounters are retained; patients can contribute to more than one period. Age is recorded without imputation. Male, non-binary, and unknown sex are pooled; missing sex remains separate. The recorded White, non-Hispanic race row is omitted, with its encounters retained in denominators. Other is a recorded race category; language and arrival categories pool remaining and missing values. The separate Unknown language category occurs only in 2024. Arrival-mode labels are recorded encounter-table categories; provider categories remain as recorded. Counts of 1–10, corresponding percentages, and selected cells from which they could be calculated are not shown (—). ED indicates emergency department; ESI, Emergency Severity Index; IQR, interquartile range.

In 2024, events occurred in 1,066 of 50,675 discharge encounters and 1,125 of 26,411 admission encounters. Discharge windows were at least 10 minutes in 84.5% overall, 86.4% with events, and 84.5% without; corresponding 2023 proportions were 85.4%, 84.0%, and 85.4%. Admission proportions overall were 96.3% in 2024 and 96.2% in 2023 (Table 2; eFigure 4). Across 2020–2023 recorded-decision encounters, discharge decision-to-window-end time was 22 minutes (IQR, 12–41); uncapped admission-plan-to-handoff time was 104 minutes (IQR, 49–186) among recorded-handoff encounters (eTable 34).

**Table 2.** Recorded Windows Sufficient for Review in the Two Test Sets.

| Review, min | Measure | Discharge | Admission |
|---|---|---|---|
| **2023 temporal test set, estimate (95% CI)** |  |  |  |
| 5 | With event, % | 91.29 (89.36 to 93.21) | 97.84 (96.98 to 98.63) |
| 5 | Without event, % | 95.61 (95.39 to 95.82) | 98.03 (97.85 to 98.21) |
| 5 | Difference, percentage points | −4.32 (−6.26 to −2.38) | −0.19 (−1.07 to 0.63) |
| 10 | With event, % | 83.99 (81.63 to 86.26) | 96.13 (95.00 to 97.18) |
| 10 | Without event, % | 85.40 (85.06 to 85.74) | 96.21 (95.96 to 96.45) |
| 10 | Difference, percentage points | −1.41 (−3.80 to 0.89) | −0.08 (−1.23 to 1.02) |
| 15 | With event, % | 74.44 (71.71 to 77.13) | 93.62 (92.06 to 95.01) |
| 15 | Without event, % | 69.96 (69.53 to 70.39) | 93.87 (93.56 to 94.18) |
| 15 | Difference, percentage points | 4.48 (1.68 to 7.15) | −0.26 (−1.82 to 1.17) |
| **2024 held-out temporal test set, descriptive percentage** |  |  |  |
| 5 | All encounters, % | 94.7 | 98.2 |
| 5 | With event, % | 93.3 | 98.2 |
| 5 | Without event, % | 94.8 | 98.2 |
| 10 | All encounters, % | 84.5 | 96.3 |
| 10 | With event, % | 86.4 | 96.3 |
| 10 | Without event, % | 84.5 | 96.3 |
| 15 | All encounters, % | 68.9 | 94.0 |
| 15 | With event, % | 76.1 | 94.2 |
| 15 | Without event, % | 68.8 | 94.0 |

Values describe recorded windows at least as long as the review. The 2023 event-minus-nonevent differences use paired patient-cluster resamples; parentheses contain 95% percentile confidence intervals from 2,000 resamples. The 2024 values are descriptive proportions; corresponding intervals and event-minus-nonevent differences were not estimated. Recorded opportunity does not establish clinician availability. CI indicates confidence interval.

### Sequential Allocation

In 2024, discharge reach was 19.4% for Risk-only, 15.8% for Prior-visits-first, 11.9% for Simple logistic, 6.3% for Age-first, 5.9% for ESI-first, and 3.2% for Random (Table 3). Risk-only reached 207 events with 2,018 attempts (9.7 per event reached) and Prior-visits-first 168 with 2,081 (12.4 per event); the difference in event reach was 3.66 percentage points (nominal 95% CI, −0.18 to 6.26; empirical coverage, 93.4%), and the replication criterion was not met. In 2023, reach was 19.5% versus 16.0% (difference, 3.5 points; nominal 95% CI, 1.1–6.5; coverage, 96.0%), with 8.9 and 11.7 attempts per event (contrasts in eTable 35 and eTable 36). A 10-minute start delay reduced 2024 Risk-only discharge reach to 16.8% (eTable 17). Admission reach in 2024 was 11.2%, 7.6%, and 10.6% for Risk-only, Prior-visits-first, and Simple logistic (eTable 17).

**Table 3.** Sequential Selection With 10-Minute Reviews and No Start Delay.

| Selection rule | Measure | Discharge | Admission |
|---|---|---|---|
| **2023 temporal test set** |  |  |  |
| Risk-only | Attempts used | 1,857 | 890 |
| Risk-only | Events reached | 208 | 112 |
| Risk-only | Event reach, % | 19.5 | 10.1 |
| Prior-visits-first | Attempts used | 1,994 | 920 |
| Prior-visits-first | Events reached | 171 | 86 |
| Prior-visits-first | Event reach, % | 16.0 | 7.7 |
| ESI-first | Attempts used | 2,011 | 890 |
| ESI-first | Events reached | 54 | 81 |
| ESI-first | Event reach, % | 5.1 | 7.3 |
| Random | Attempts used | 1,865 | 854 |
| Random | Events reached | 29 | 34 |
| Random | Event reach, % | 2.7 | 3.1 |
| Age-first | Attempts used | 1,953 | 868 |
| Age-first | Events reached | 78 | 35 |
| Age-first | Event reach, % | 7.3 | 3.1 |
| Simple logistic | Attempts used | 2,006 | 944 |
| Simple logistic | Events reached | 160 | 125 |
| Simple logistic | Event reach, % | 15.0 | 11.2 |
| Risk-plus-window | Attempts used | 1,856 | 890 |
| Risk-plus-window | Events reached | 208 | 115 |
| Risk-plus-window | Event reach, % | 19.5 | 10.3 |
| **2024 held-out temporal test set** |  |  |  |
| Risk-only | Attempts used | 2,018 | 901 |
| Risk-only | Events reached | 207 | 126 |
| Risk-only | Event reach, % | 19.4 | 11.2 |
| Prior-visits-first | Attempts used | 2,081 | 943 |
| Prior-visits-first | Events reached | 168 | 86 |
| Prior-visits-first | Event reach, % | 15.8 | 7.6 |
| ESI-first | Attempts used | 2,081 | 925 |
| ESI-first | Events reached | 62.5 | 70 |
| ESI-first | Event reach, % | 5.9 | 6.2 |
| Random | Attempts used | 1,973 | 898 |
| Random | Events reached | 34 | 40.5 |
| Random | Event reach, % | 3.2 | 3.6 |
| Age-first | Attempts used | 2,069 | 890 |
| Age-first | Events reached | 67 | 54 |
| Age-first | Event reach, % | 6.3 | 4.8 |
| Simple logistic | Attempts used | 2,132 | 927 |
| Simple logistic | Events reached | 127 | 119 |
| Simple logistic | Event reach, % | 11.9 | 10.6 |
| Risk-plus-window | Attempts used | 2,028 | 910 |
| Risk-plus-window | Events reached | 208 | 123 |
| Risk-plus-window | Event reach, % | 19.5 | 10.9 |

Daily capacity was 6 discharge and 3 admission attempts: 2,190 and 1,095 annually in 2023, and 2,196 and 1,098 in 2024. All rules used the same planned capacity and fixed thresholds targeting 5% of tuning encounters. Attempts used could differ. Event reach is the proportion of all event encounters selected with sufficient recorded time. Attempts without sufficient time still consume capacity. Fractional event counts reflect shared selection at simultaneous decision times. These descriptive estimates do not measure events prevented. ESI indicates Emergency Severity Index.

With 5-minute reviews and 10% tuning thresholds, discharge Risk-only and Prior-visits-first reach was 32.6% and 26.9% in 2024, compared with 33.1% and 28.5% in 2023. Duration, flag fractions, and capacity all differed from the 10-minute scenario. With 10-minute reviews, 5% tuning thresholds, and zero delay, discharge Risk-plus-window reach was 19.5% in both years. Among 2023 discharge encounters with events, the window model’s area under the receiver operating characteristic curve (AUROC) was 0.750, with predicted probabilities of 0.761 and 0.984 at the 5th and 95th percentiles. Across eligible 2023 discharge encounters, the scores had a Spearman correlation of 0.997 and the rules shared 1,750 selections.

### Model Performance and Subgroups

Both primary models selected gradient-boosted trees. Discharge and admission AUROCs were 0.786 (95% CI, 0.770–0.801) and 0.676 (95% CI, 0.660–0.692) in 2024, versus 0.788 and 0.659 in 2023. In 2024, calibration slopes were 0.982 and 1.110, and observed-to-expected ratios were 0.93 in both groups. Fixed 2022 thresholds flagged 5.6% of 2024 discharge encounters for Risk-only and 8.0% for Simple logistic (target, 5%) (Table 4; eTable 37; eTable 38; eTable 39; eTable 18; eFigure 5). With vital signs, 2023 AUROCs were 0.787 and 0.671; this sensitivity was not evaluated in 2024.

**Table 4.** Primary Model Performance in the Two Test Sets.

| **Measure** | **2023 discharge** | **2023 admission** | **2024 discharge** | **2024 admission** |
|---|---|---|---|---|
| AUROC | 0.788 (0.773 to 0.802) | 0.659 (0.641 to 0.678) | 0.786 (0.770 to 0.801) | 0.676 (0.660 to 0.692) |
| Average precision | 0.093 (0.081 to 0.108) | 0.097 (0.085 to 0.113) | 0.086 (0.074 to 0.101) | 0.094 (0.084 to 0.106) |
| Brier score | 0.021 (0.019 to 0.022) | 0.043 (0.040 to 0.045) | 0.020 (0.019 to 0.022) | 0.040 (0.038 to 0.042) |
| Log loss | 0.093 (0.088 to 0.098) | 0.178 (0.169 to 0.186) | 0.090 (0.085 to 0.096) | 0.168 (0.161 to 0.175) |
| Observed-to-expected ratio | 1.037 (0.964 to 1.107) | 1.006 (0.941 to 1.067) | 0.928 (0.861 to 1.001) | 0.933 (0.884 to 0.984) |
| Calibration intercept | 0.039 (−0.039 to 0.109) | 0.006 (−0.065 to 0.069) | −0.080 (−0.160 to 0.001) | −0.074 (−0.131 to −0.018) |
| Calibration slope | 0.994 (0.934 to 1.052) | 1.012 (0.901 to 1.128) | 0.982 (0.921 to 1.045) | 1.110 (1.008 to 1.213) |

Both models used gradient-boosted trees, selected and intercept-recalibrated using 2022 tuning data, without fitting or recalibration on either test year. Parentheses contain patient-cluster percentile 95% confidence intervals from 2,000 resamples in each test year. The calibration intercept fixes slope at 1; the calibration slope is estimated jointly with an intercept. AUROC indicates area under the receiver operating characteristic curve.

Under sequential discharge selection, Risk-only reach for ages 18–39 versus 65 years or older was 11.6% versus 26.6% in 2023 and 13.5% versus 26.5% in 2024; Prior-visits-first reach in 2024 was 15.1% and 15.3%. Risk-only reach for Black versus White patients was 19.6% versus 19.8% in 2023 and 15.4% versus 18.5% in 2024 (eTable 40; eTable 41).

### Recorded Transitions and Other Analyses

Across 2020–2023, 12,070 of 96,249 admission plans (12.5%) had a recorded change to a discharge decision, and 2,400 of 183,127 discharge decisions (1.3%) to an admission plan or handoff by departure or CDU entry (eTable 42); early admission plans may reverse because pending results return. The last qualifying status was classified as “Direction change” for 1,176 of 75,077 resident-discharge encounters (1.6%) and 2,233 of 25,314 resident-admission encounters (8.8%). Among encounters whose next qualifying status was In Process, the first later qualifying status other than In Process was opposite to the resident plan in about 14% and 41%, respectively (eTable 24 and eTable 26). These are recorded changes, not verified attending overrides. Final discharge occurred in 8,167 admission-group encounters with a CDU stay (eTable 4).

Returns ending in admission occurred in 1,050 of 1,068 discharge composite-event encounters in 2023; components could overlap (eTable 43). Additional analyses appear in eTable 44–eTable 61, eFigure 6– eFigure 13, and eResults.

## Discussion

This study found 3 things. First, the recorded window was at least 10 minutes for 85.4% of discharge decisions in the 2023 temporal test set and 84.5% in the 2024 held-out temporal test set. Second, in 2024, selection by prior visits or predicted risk reached about 3 to 6 times as many discharge events as Random or ESI-first selection. Third, Risk-only exceeded Prior-visits-first by 3.5 percentage points in 2023 and 3.7 in 2024. The 2024 interval included zero, and the prespecified replication criterion was not met. Across 2020–2023, recorded direction changes followed 1.3% of discharge decisions and 12.5% of admission plans. Review selection was simulated. The study did not measure harm prevented.

The results describe a design that could be tested prospectively. The simulation budgeted up to six 10-minute discharge reviews (60 minutes) per day among roughly 140 eligible discharge decisions. Prior-visits-first and Risk-only reached 168 and 207 events in 2024, about 3 to 4 per week. An electronic health record prompt could fire when the attending records readiness or conditional discharge, flag patients by prior-visit count without a model, and request a 5-to 10-minute review. ESI-first reached 5.9% of 2024 discharge events. Most eligible discharge encounters (82.9%) had ESI level 2 or 3, and triage severity alone could not distinguish among patients at the same level.

Why not use the model? Its additional reach was modest and uncertain, and it carries costs. The model requires infrastructure and maintenance. With thresholds fixed after targeting 5% in 2022, Risk-only flagged 5.6% and Simple logistic 8.0% of 2024 discharge encounters. Risk-only selected older patients more often. In 2024, Risk-only reached 13.5% of events among patients aged 18 to 39 years and 26.5% among those 65 years or older, whereas Prior-visits-first reached about 15% in both groups. Lower reach does not imply that younger patients would benefit less from review, and these comparisons do not establish subgroup calibration or equitable benefit.

Time was a real constraint. About 15% of 2024 discharge windows were shorter than 10 minutes. A 10-minute delay reduced simulated Risk-only reach from 19.4% to 16.8%. The 5-minute scenario reached more events in both years, but its higher flag fraction and capacity prevent attributing the gain to duration alone, and what a 5-minute review can accomplish was not measured. In 2023, weighting by window probability changed the discharge ranking little (Spearman correlation, 0.997).

The admission contrast needs caution. Pending results may explain some changes. The reasons were unmeasured. Because residents may record plans before or after discussion, these changes cannot establish attending overrides or observed senior review. Weaker admission discrimination may reflect plans recorded before results return or accepting-team reassessment; neither was measured. Admission-group membership also used later status sequences, so it is a retrospective contrast. Longer ED boarding was associated with in-hospital mortality^13^ and early clinical deterioration,^18^ and our findings do not support prolonging ED stays.

Models predicted ED returns within 72 hours or 9 days,^19^ admission or death among patients returning within 72 hours,^20^ escalation or death after ward admission,^21^ or ICU care within 24 hours of admission, including direct ED-to-ICU admission.^22^ Differences in populations, prediction times, and outcomes preclude claims of superior clinical usefulness. Our contribution is evaluating selection at the discharge decision, under a fixed review budget, against simple rules based on routinely available information.^12,23^

Electronic outcomes are screening signals, not adjudicated errors. Electronic-trigger studies found missed diagnostic opportunities in a minority of return admissions, yields that cannot be assumed here.^24,25^ Predicted risk does not identify who would benefit from review,^26^ and CHARMED’s reduction in adverse events came mainly from fewer near misses. The reduction in preventable serious adverse events was not statistically significant.^5^

### Limitations

No study reviews were delivered. Selection was simulated, and the electronic outcomes were not adjudicated for preventability. This single-center study also requires external validation, and patients could appear in more than one period. Recorded timestamps may differ from clinical events,^27^ and transfer of responsibility was not measured. Events outside available records could be missed, and escalation need not represent unplanned deterioration. Excluding unknown outcomes may have introduced selection bias. Thirteen of 284,627 candidate encounters from 2020–2023 belonged to patients whose identifier mappings differed between data releases; this does not establish incorrect labels. Triage availability was assumed, vital-sign times did not establish visibility, and blank vital-sign units were assigned from value distributions.

Simulations assumed fixed review durations and omitted queues, interruptions, and staffing, and full-day rankings used later decisions. Intervals conditioned on fitted models, and poor single-rule coverage and imperfect contrast coverage limit inference. Chart review is needed to establish clinical importance and preventability.

A paramedic expeditor role was associated with shorter ED stays and less diversion, but returns and escalation were not evaluated.^28^ A prospective study should measure review time, staffing, management changes, and patient outcomes.

## Conclusions

Most attending discharge decisions had a recorded window of at least 10 minutes. Selection by prior ED visits or predicted risk reached several times more subsequent events than random or triage-based selection, and a risk model did not reliably outperform the simpler prior-visit rule. A prospective study should test whether brief, targeted review at the discharge decision improves outcomes.

## Data Availability

Patient-level data cannot be shared under institutional data-use agreements. Analysis code and a data dictionary are available from the corresponding author on reasonable request.

## Acknowledgment

This research used data or services provided by STARR, “STAnford medicine Research data Repository,” a clinical data warehouse containing live Epic data from Stanford Health Care, the Stanford Children’s Hospital, the University Healthcare Alliance and Packard Children’s Health Alliance clinics and other auxiliary data from Hospital applications such as radiology PACS. STARR platform is developed and operated by Stanford Medicine Research Technology team and is made possible by funding from Stanford School of Medicine Dean’s Office.

## Author Contributions

Dr Aich had full access to all of the data in the study and takes responsibility for the integrity of the data and the accuracy of the data analysis. *Concept and design:* All authors. *Acquisition, analysis, or interpretation of data:* All authors. *Drafting of the manuscript:* Aich. *Critical review of the manuscript for important intellectual content:* All authors. *Statistical analysis:* Aich. *Supervision:* Preiksaitis, Rose.

## Additional Contributions

Artificial intelligence tools—ChatGPT-6 Astra (OpenAI) and Claude Opus 5.5 (Anthropic)—were used to assist with formatting and editing of the manuscript; the authors reviewed all content and take responsibility for it.

## References

1. Calder LA, Arnason T, Vaillancourt C, Perry JJ, Stiell IG, Forster AJ. How do emergency physicians make discharge decisions? Emerg Med J. 2015;32(1):9–14. doi:10.1136/emermed-2013-202421

2. Forster AJ, Rose NGW, van Walraven C, Stiell I. Adverse events following an emergency department visit. Qual Saf Health Care. 2007;16(1):17–22. doi:10.1136/qshc.2005.017384

3. Delgado MK, Liu V, Pines JM, Kipnis P, Gardner MN, Escobar GJ. Risk factors for unplanned transfer to intensive care within 24 hours of admission from the emergency department in an integrated healthcare system. J Hosp Med. 2013;8(1):13–19. doi:10.1002/jhm.1979

4. White AL, Armstrong PAR, Thakore S. Impact of senior clinical review on patient disposition from the emergency department. Emerg Med J. 2010;27(4):262–265. doi:10.1136/emj.2009.077842

5. Freund Y, Goulet H, Leblanc J, et al. Effect of systematic physician cross-checking on reducing adverse events in the emergency department: the CHARMED cluster randomized trial. JAMA Intern Med. 2018;178(6):812–819. doi:10.1001/jamainternmed.2018.0607

6. Kirkwood D, Dalla Vecchia L. Consultant Sign-off 2022: National Quality Improvement Project. National Report. Royal College of Emergency Medicine. Published April 2023. Accessed October 15, 2026. https://rcem.ac.uk/wp-content/uploads/2023/04/Consultant_Sign_Off_Final_v2.pdf

7. Gabayan GZ, Gould MK, Weiss RE, Chiu VY, Sarkisian CA. A risk score to predict short-term outcomes following emergency department discharge. West J Emerg Med. 2018;19(5):842–848. doi:10.5811/westjem.2018.7.37945

8. Zhang Y, Huang Y, Rosen A, et al. Aspiring to clinical significance: insights from developing and evaluating a machine learning model to predict emergency department return visit admissions. PLOS Digit Health. 2024;3(9):e0000606. doi:10.1371/journal.pdig.0000606

9. Glass G, Hartka TR, Keim-Malpass J, Enfield KB, Clark MT. Dynamic data in the ED predict requirement for ICU transfer following acute care admission. J Clin Monit Comput. 2021;35(3):515–523. doi:10.1007/s10877-020-00500-3

10. van Dam PMEL, van Doorn WPTM, Sevenich L, et al. Machine learning for risk stratification in the emergency department (MARS-ED): a randomized controlled trial. Nat Commun. 2026;17(1):242. doi:10.1038/s41467-025-66947-7

11. Jung K, Kashyap S, Avati A, et al. A framework for making predictive models useful in practice. J Am Med Inform Assoc. 2021;28(6):1149–1158. doi:10.1093/jamia/ocaa318

12. Singh K, Shah NH, Vickers AJ. Assessing the net benefit of machine learning models in the presence of resource constraints. J Am Med Inform Assoc. 2023;30(4):668–673. doi:10.1093/jamia/ocad006

13. Singer AJ, Thode HC Jr, Viccellio P, Pines JM. The association between length of emergency department boarding and mortality. Acad Emerg Med. 2011;18(12):1324–1329. doi:10.1111/j.1553-2712.2011.01236.x

14. Centers for Medicare & Medicaid Services. Median admit decision time to ED departure time for admitted patients (CMS111v11). eCQI Resource Center. Accessed October 15, 2026. https://ecqi.healthit.gov/sites/default/files/ecqm/measures/CMS111v11.html

15. von Elm E, Altman DG, Egger M, Pocock SJ, Gøtzsche PC, Vandenbroucke JP; STROBE Initiative. The Strengthening the Reporting of Observational Studies in Epidemiology (STROBE) statement: guidelines for reporting observational studies. PLoS Med. 2007;4(10):e296. doi:10.1371/journal.pmed.0040296

16. Benchimol EI, Smeeth L, Guttmann A, et al; RECORD Working Committee. The REporting of studies Conducted using Observational Routinely-collected health Data (RECORD) statement. PLoS Med. 2015;12(10):e1001885. doi:10.1371/journal.pmed.1001885

17. Collins GS, Moons KGM, Dhiman P, et al. TRIPOD+AI statement: updated guidance for reporting clinical prediction models that use regression or machine learning methods. BMJ. 2024;385:e078378. doi:10.1136/bmj-2023-078378

18. Rizer NW, Klein E, Copenhaver MS, Zhao X, Kelen GD, Hinson JS. Early clinical deterioration among emergency department boarders: a retrospective analysis. Ann Emerg Med. 2026;87(6):681–693. doi:10.1016/j.annemergmed.2026.01.023

19. Hong WS, Haimovich AD, Taylor RA. Predicting 72-hour and 9-day return to the emergency department using machine learning. JAMIA Open. 2019;2(3):346–352. doi:10.1093/jamiaopen/ooz019

20. Sung CW, Ho J, Fan CY, et al. Prediction of high-risk emergency department revisits from a machine-learning algorithm: a proof-of-concept study. BMJ Health Care Inform. 2024;31(1):e100859. doi:10.1136/bmjhci-2023-100859

21. Sutherland ME, Yarmis SJ, Lemkin DL, Winters ME, Dezman ZDW. National Early Warning Score is modestly predictive of care escalation after emergency department-to-floor admission. J Emerg Med. 2020;58(6):882–891. doi:10.1016/j.jemermed.2020.03.032

22. Nguyen M, Corbin CK, Eulalio T, et al. Developing machine learning models to personalize care levels among emergency room patients for hospital admission. J Am Med Inform Assoc. 2021;28(11):2423–2432. doi:10.1093/jamia/ocab118

23. Romero-Brufau S, Huddleston JM, Escobar GJ, Liebow M. Why the C-statistic is not informative to evaluate early warning scores and what metrics to use. Crit Care. 2015;19(1):285. doi:10.1186/s13054-015-0999-1

24. Marks CM, Gibney S, Stenson B, et al. Screening for missed opportunities for diagnosis in the ED using eTriggers and large language models. JAMA Netw Open. 2026;9(6):e2620939. doi:10.1001/jamanetworkopen.2026.20939

25. Vaghani V, Gupta A, Mir U, et al. Implementation of electronic triggers to identify diagnostic errors in emergency departments. JAMA Intern Med. 2025;185(2):143–151. doi:10.1001/jamainternmed.2024.6214

26. Marafino BJ, Schuler A, Liu VX, Escobar GJ, Baiocchi M. Predicting preventable hospital readmissions with causal machine learning. Health Serv Res. 2020;55(6):993–1002. doi:10.1111/1475-6773.13586

27. Gordon BD, Flottemesch TJ, Asplin BR. Accuracy of staff-initiated emergency department tracking system timestamps in identifying actual event times. Ann Emerg Med. 2008;52(5):504–511. doi:10.1016/j.annemergmed.2007.11.036

28. Handel DA, Ma OJ, Workman J, Fu R. Impact of an expeditor on emergency department patient throughput. West J Emerg Med. 2011;12(2):198–203. https://pmc.ncbi.nlm.nih.gov/articles/PMC3099607/ PMID: 21691526; PMCID: PMC3099607

