## Supplement for "Recorded Time and Risk-Based Selection for a Second Look Before Emergency Department Discharge"

Contents

eMethods	2

eResults	11

eTable 1. Operational Codes	12

eTable 2. Time-Zero Assignment by Year	13

eTable 3. Decision-Time Group Denominators	14

eTable 4. Recorded Reversals, Missing Handoffs, and Observation Stays	14

eTable 5. Electronic Outcome Ascertainment	15

eTable 6. Composite Outcome Coverage	15

eTable 7. Candidate Numeric Laboratory Components	15

eTable 8. Numeric Laboratory Predictor Availability in Training, 2020–2021	16

eTable 9. Predictor Missingness in 2020–2023	17

eTable 10. Usable First Vital Signs and Measurement-to-Decision Intervals in 2020–2023	17

eTable 11. Vital-Sign Sensitivity: Prediction and Ranking Allocation in 2023	18

eTable 12. Conditional Window-Model Performance in 2022	19

eTable 13. Conditional Window-Model Discrimination and Predictions in 2023	21

eTable 14. Risk-Only and Risk-Plus-Window Selection Overlap in 2023	21

eTable 15. Sequential Thresholds and Planned Capacities	21

eTable 16. Sequential Selection Across Review Durations, Thresholds, and Delays in 2023	23

eTable 17. Sequential Selection Across Scenarios in 2024	27

eTable 18. Observed Flag Fractions in 2024	31

eTable 19. Full-Day and Annual Ranking at the Main Budget in 2023	34

eTable 20. Selection, Reach, and Lift Across Ranking Budgets in 2023	35

eTable 21. Review Yield Across Ranking Budgets in 2023	43

eTable 22. Definitions of Recorded Resident-Plan Transitions	48

eTable 23. Next Recorded Status After Resident-Plan Markers, 2020–2023	49

eTable 24. Last Qualifying Status After Resident-Plan Markers, 2020–2023	49

eTable 25. Last Resident-Plan Status and Final ED Disposition, 2020–2023	50

eTable 26. Recorded Status After a Return to In Process, 2020–2023	52

eTable 27. Empirical Coverage of Sequential Reach and Paired Contrasts in 2023	53

eTable 28. Empirical Coverage of Sequential Intervals in 2024	53

eTable 29. Empirical Coverage of Full-Day and Annual Ranking Intervals in 2023	54

eTable 30. Eligible Cohorts and Event Rates by Period	55

eTable 31. Recorded Demographic Characteristics by Period	56

eTable 32. Clinical Characteristics by Decision Group in 2023	57

eTable 33. Clinical Characteristics by Decision Group in 2024	57

eTable 34. Uncapped Recorded Decision, Handoff, and Departure Intervals in 2020–2023	58

eTable 35. Sequential Event-Reach Contrasts in 2023	59

eTable 36. Sequential Event-Reach Contrasts in 2024	62

eTable 37. Prediction Sensitivity Analyses in 2023	66

eTable 38. Five-Variable Logistic Model Performance in 2023	67

eTable 39. Model Performance and Prediction Sensitivities in 2024	68

eTable 40. Sequential Selection by Patient Characteristics in 2023	70

eTable 41. Sequential Selection by Patient Characteristics in 2024	75

eTable 42. Eventual Recorded Direction Changes During ED Care, 2020–2023	82

eTable 43. Ranking of Individual Outcome Components in 2023	83

eTable 44. Capped Windows by Recorded Decision Source	83

eTable 45. Recorded Windows Before Model Exclusions in 2023	83

eTable 46. Readiness and Conditional-Discharge Order Among Final Discharges	84

eTable 47. Annual Pooling at the Main Ranking Budget in 2023	84

eTable 48. Descriptive Full-Day and Annual Ranking Contrasts in 2023	84

eTable 49. Risk-Only Capacity and Selection-Order Comparisons in 2023	94

eTable 50. Risk-Only Selection by Daily Demand in 2023	94

eTable 51. Recorded Windows and Outcome Association in 2023	95

eTable 52. Copula Independence Tests in 2023	95

eTable 53. Spline and Copula Descriptions of Outcome Association in 2023	95

eTable 54. Recorded Windows and Predicted Risk in 2023	95

eTable 55. Case Mix by Decision-Time Group	97

eTable 56. Other-Template Record Presence in 2023	103

eTable 57. First Vital-Sign Measurements Outside Screening Bounds	105

eTable 58. Encounters Without a Usable Predecision Vital Sign	107

eTable 59. Resident-Marker Direction and Time-Zero Direction	107

eTable 60. Resident-Discharge Markers Relative to Discharge Decisions	107

eTable 61. Characteristics of Eligible Encounters and Encounters Excluded for an Unknown Composite Outcome	108

eFigure 1. Study Workflow and Temporal Evaluation	110

eFigure 2. Predictor Missingness	111

eFigure 3. Calibration in the 2023 Temporal Test Set	112

eFigure 4. Recorded Windows Sufficient for Review in 2023	113

eFigure 5. Calibration in the 2024 Held-Out Temporal Test Set	114

eFigure 6. Coverage of Full-Day and Annual Ranking Intervals in 2023	115

eFigure 7. Full-Day Ranking With Clinical Comparators in 2023	116

eFigure 8. Full-Day Ranking Event Reach in 2023	117

eFigure 9. Full-Day Ranking Lift in 2023	118

eFigure 10. Annual Ranking Event Reach in 2023	119

eFigure 11. Annual Ranking Lift in 2023	120

eFigure 12. Recorded Windows Before Model Exclusions	121

eFigure 13. Window Length and Outcome Association in 2023	122

eMethods

1. Sources, Identity, and Linkage

The study used adult ED electronic health record data linked to hospital admission, discharge, and transfer (ADT) records and death dates. The 2020–2023 sources represented the September 30, 2026, 02:54:50 UTC snapshot; the 2024 sources represented November 6, 2026, 06:22:36 UTC. Vital signs for the 2023 sensitivity analysis are described in section 14. Clinical timestamps were analyzed in local time. Candidates had recorded ages 18–100 years, first ED department 110100019, and arrivals during 2020–2024.

Patient identity was resolved from patient identifiers associated with the encounter in care-area, ED ADT, and race records. Blank identifiers were ignored. One distinct patient identifier was required; missing or conflicting identities remained unresolved. Crosswalk rows were deduplicated. A usable mapping required one complete mapping per patient and a one-to-one anonymous patient identifier. The coded encounter identifier equaled the source encounter identifier plus the patient-specific encounter-ID offset. Jittered dates were restored by subtracting the patient-specific number of jitter days. Hospital ADT records were linked using the anonymous patient and coded encounter identifiers. Death records were linked by anonymous patient identifier. Hospital ADT extraction covered linked patients from 1 day before arrival through 7 days after valid ED departure; arrival replaced a missing or earlier departure when setting the upper limit. Effective time determined inclusion. If effective time was missing, event time was used; records missing both times were retained for ascertainment checks. Missing or conflicting links were treated as unknown, not as absence of an event.

Checks assessed identifier uniqueness, encounter counts before and after joins, source and local timestamp agreement, duplicate records, and linkage coverage by year. Exact ADT duplicates were removed. Conflicting versions of an ADT event were treated as ambiguous. These checks did not independently validate the clinical meaning of a recorded plan or outcome.

For 2024, ADT and death records used the 2025 STARR release. Encounters were matched to that release by the coded encounter identifier minus a release-level constant. The time shift was ED arrival minus jittered arrival, required to be a whole number of days and stable within each anonymous patient identifier. The coded inpatient identifier equaled its native identifier plus a release-level constant. Existing usable patient mappings were retained when consistent with this linkage. For patients without an existing mapping, all matched encounters had to give one anonymous identifier and shift, no other native patient or unresolved encounter could own that identifier, and every 2024 encounter had to match. Patients with conflicting existing and release-specific identifiers or shifts remained unlinked for 2024. Missing linked outcome components remained unknown; definite events in native ED data could still establish a positive composite. Prior-visit information extended from September 2023, and outcome follow-up extended through January 2025. Native records and projected laboratory records for overlapping 2023 encounters were compared between extracts. These comparisons assessed data consistency, not clinical validity.

2. Cohort and Study Periods

Adult ED encounters were retained as separate observations, including repeat visits. The prediction and allocation cohort required a resolved identity, a qualifying recorded discharge decision or admission plan, a valid review window, and a known composite outcome. The discharge-priority rule below defined mutually exclusive groups; final disposition did not determine assignment. Encounters with no usable decision time were described separately. A separate group was defined for valid CDU entry at or before departure with no qualifying discharge decision, admission plan, or handoff through entry. No 2020–2023 encounters met this definition.

Arrival years defined training (2020–2021), tuning (2022), the temporal test set (2023), and the held-out temporal test set (2024). Time zero had to fall within the assigned period. Training and tuning encounters whose 7-day outcome window reached the next period were excluded. This boundary exclusion did not apply to either test year; follow-up for late-2024 decisions could extend into January 2025. Patients could appear across periods. Preprocessing used training data; selection and recalibration used 2022. The age upper limit applied to index encounters, not to the adult ED return or prior-visit pools. Possible returns included adult ED encounters outside the model cohort; a return did not need a qualifying decision or known composite outcome of its own.

3. Recorded Decision Times and Window Endpoints

Qualifying discharge markers were readiness (6) and conditional discharge (106111); admission-plan markers were Resident Admit Dispo Selected (106108) and Decision to Admit Dispo Selected (Attending) (106116). Admission-status labels do not independently identify the entering clinician. Signed admission order (106107) and readiness for admission (10) were recorded handoff markers. Resident discharge (100030) did not set time zero. In local workflow, readiness and conditional-discharge markers document the attending physician's discharge decision and can only be entered by attending physicians. Conditional discharge delegates completion to the nurse once specified criteria are met. The individual user was not analyzed. A decision to admit is provisional and may be recorded by a resident or attending physician. In local workflow, the admission order is usually signed immediately after discussion with the accepting team. Its timestamp may approximate that discussion, whose actual timing was not directly recorded. This interpretation applies to code 106107, not automatically to code 10, CDU entry, departure, or fallback endpoints. Actual transfer of responsibility was unmeasured.

A marker qualified only when timed and at or after arrival and at or before the earlier of ED departure and first CDU entry. Untimed markers and markers outside that interval were disregarded without automatically excluding an otherwise usable encounter. Repeated markers used their earliest qualifying time. Missing departure, departure before arrival, untimed CDU location, CDU entry before arrival, and ambiguous first CDU entry at or before ED departure prevented a valid time zero. CDU entry used care-area codes 41, 42, and 10616011; an incompatible location recorded simultaneously at the first entry was ambiguous. A CDU waiting-area label alone did not establish entry. The CDU provides ED-based observation, distinct from hospital admission and hospital observation; admission plans ending in CDU entry remained in the admission group.

The earliest discharge decision at or before the first recorded handoff took priority, including after an earlier admission plan. An admission-plan or handoff marker simultaneous with that discharge decision was an unresolved conflict and was excluded. Discharge markers after the first handoff did not establish an ED discharge-review opportunity. Otherwise, an admission plan at or before handoff, or without a recorded handoff, set admission time zero. Encounters with a handoff were excluded when no qualifying plan was recorded at or before the first handoff. Because discharge decisions had priority over earlier admission plans, admission-group membership used the subsequent sequence and is a retrospective contrast rather than a prospectively identifiable group at the initial admission marker.

For discharge decisions, the window ended at the earliest subsequent admission-plan or handoff marker, ED departure, or CDU entry. The subsequent admission-plan or handoff marker was a recorded reversal, not an outcome or proof of deterioration. Discharge–admission–discharge sequences retained the first qualifying discharge decision and ended the window at the first reversal. For admission plans, the endpoint was the earliest recorded handoff, departure, or CDU entry. Missing-handoff admission plans used departure or CDU entry as a flagged fallback; these were not measured handoff windows. Final disposition never removed or reassigned an encounter.

Valid zero-length windows remained eligible. Review windows were capped at 60 minutes for allocation. Caps of 30 and 120 minutes were used only for window and association sensitivities. Uncapped decision-to-handoff and handoff-to-departure descriptions used their stated populations (eTable 34). Windows describe recorded elapsed time, not staff availability. Windows by decision source and readiness/conditional-discharge order are described in eTable 44, eTable 45, and eTable 46.

4. Electronic Outcome Algorithms

The primary outcome was a composite of three recorded events: an adult ED return arriving within 7 days and ultimately ending in inpatient admission, critical care within 24 hours, or death within 7 days. All intervals began at time zero and included their upper limits. A qualifying return had to arrive strictly after the index ED departure and after hospital discharge when that time was recorded. Observation-only returns did not meet the return-admission component. An otherwise time-eligible return with a missing disposition (null or blank) left the return component uncertain unless a qualifying inpatient return established a positive component. If the index encounter ended in admission and its hospital discharge time was missing, an apparent qualifying return was classified as uncertain.

We identified critical care from hospital level-of-care records regardless of patient class. For admission plans, an event required an initial acute or intermediate level followed by critical care within 24 hours after time zero. The starting level was the last active recorded level at time zero or, if none was active, the first recorded level afterward. For discharge decisions, critical care could occur during the index encounter or a subsequent adult ED return within the same 24-hour horizon. Critical care already active at time zero and admission plans whose starting level was critical care did not count as critical-care events. These findings alone did not exclude encounters from the cohort; the composite outcome depended on the return and death components. Blank level-of-care updates carried forward the last active level; discharge or transfer out ended that state unless a simultaneous eligible level update took precedence; conflicting simultaneous levels were ambiguous. When admission, discharge, and transfer records were present but contained no level of care, we classified the component as no recorded critical-care event. A sensitivity analysis treated these cases as uncertain.

Death counted only when the entire recorded calendar day fell strictly after time zero and ended within the 7-day horizon. Dates overlapping either boundary were classified as uncertain. An ED expired status could contribute a positive death outcome only in a sensitivity analysis. Missing links, absent required records, conflicting information, and unresolved timing could leave individual components unknown. The composite was positive if any component was positive. Otherwise, it was unknown if any component was unknown and classified as no recorded event if none was unknown. Unknown composite outcomes were excluded from the primary analyses. A negative classification indicated no qualifying event recorded in the available sources.

An adult ED return with an admission disposition or a database-null disposition at exactly the index departure or hospital discharge time left the return component uncertain unless a qualifying return was also present. Encounters entered in error did not qualify as returns. For an index admission with a known discharge time, a subsequent ED arrival before that discharge was not a qualifying return. The adult ED return pool did not provide complete surveillance of other hospitals.

ADT level-of-care histories used effective timestamps. Original and updated events were eligible; discharge and transfer-out events ended an active state unless a simultaneous eligible level update took precedence. A blank update did not erase a recorded active level. Conflicting simultaneous levels were ambiguous. Admission-plan escalation required critical care strictly after the starting acute or intermediate level, except that an acute or intermediate level already active at time zero used time zero as its starting point. The upper limit was time zero plus 24 hours. No linked ADT records and linked ADT records with no level of care were distinguished: the latter counted as no recorded event in the primary analysis and as uncertain in the conservative sensitivity analysis.

In the primary analysis, an ED expired status without a recorded death date left the death component uncertain. A resolved death date counted only if the day began strictly after time zero and its end was no later than time zero plus 7 days. A death date ending at or before time zero, conflicting death dates, or failed date conversion left the death component uncertain. The ED expired sensitivity used the timed status only within the 7-day horizon.

5. Predictors and Laboratory Availability

Predictors included age, the type of recorded decision, and elapsed time from arrival to time zero. We also included the decision's time of day, day of the week, and month. Prior ED use counted adult ED visits that arrived within the preceding 90 days and ended before time zero. This count was left missing when fewer than 90 days of source data preceded time zero; the available history length was also included. Laboratory predictors comprised order counts, counts of available test components, and selected numeric results. We included the recorded Emergency Severity Index (ESI), arrival mode, and first chief-complaint code. Their recording times were unavailable, so we assumed they represented triage information available at time zero. A sensitivity analysis omitted all three fields. The primary models did not include vital signs. Sex, language, interpreter need, race, and ethnicity were not used as predictors. They were used to describe the cohort and compare allocation across subgroups.

We considered a fixed list of 15 laboratory components in specified units. A component was retained if valid numeric results were available for at least 500 training encounters and at least 1% of training encounters within the admission or discharge group. Numeric results required nonmissing order, result, and verification timestamps with valid chronology, all at or before time zero. Availability was defined by the later of the result and verification timestamps. For each component and unit, we examined the latest record meeting these timing requirements. We accepted only finite, unambiguous numeric values with a verified status. Values reported as inequalities were excluded. If the latest record was unusable, its value remained missing; we did not substitute an older result. Units were not converted.

Decision-record type distinguished the qualifying discharge-readiness, conditional-discharge, resident-admission, or attending-admission markers, including simultaneous same-direction markers. Time of day used sine and cosine terms; weekday and month were categorical. Prior visits had to arrive during the 90 days before time zero, have a valid departure strictly before time zero, and not be entered in error. Less than 90 days of available history produced a missing visit count rather than an observed zero.

Laboratory chronology also required ordering during the ED encounter, with result and verification timestamps no earlier than the order. When collection time was present, it could not precede ED arrival or follow availability. At the latest eligible timestamp, conflicting values or verification states made that component unavailable. Laboratory order counts included distinct orders placed between arrival and time zero; available-component counts used components meeting the timing rules. Neither count required a usable value in the final 15-component numeric set.

6. Model Development and Evaluation

We fitted separate models for admission plans and discharge decisions, comparing ridge logistic regression with histogram-based gradient-boosted trees. All preprocessing parameters were estimated from the training data. Missing numeric values were replaced with training medians, with a separate missingness indicator for each variable. Numeric values were clipped at the training 0.5th and 99.5th percentiles and standardized. Chief-complaint codes were limited to the 25 most frequent codes in the training data. Other codes were grouped as other, and missing codes were assigned a missing category. Categorical predictors were represented by indicator variables. Categories appearing in fewer than 50 training encounters and previously unseen categories were grouped as other, with missing values handled separately. For each decision group, we selected the model with the lowest log loss in 2022 before recalibration. We then fitted an intercept-only adjustment using the 2022 data, retaining the fitted model coefficients and tree structures.

Ridge models used inverse regularization strengths of 0.1, 1, and 10. Tree models used a maximum of 7 or 15 leaves and 100 or 250 boosting iterations. Other tree settings were a learning rate of 0.05, a minimum of 100 encounters per leaf, and an L2 regularization parameter of 10. Early stopping was disabled. Neither model family used class weighting.

We evaluated the selected, recalibrated models in the 2023 temporal test set and the 2024 held-out temporal test set. The fitted primary, simple, and window models and training-derived preprocessing were unchanged for 2024; no model fitting, tuning, or recalibration used 2024 data. We measured discrimination using the area under the receiver operating characteristic curve and average precision. We also reported the Brier score and log loss. Calibration was assessed using the observed-to-expected event ratio, calibration intercept, and calibration slope. Calibration plots compared predicted and observed event rates within risk groups defined by decile cutpoints from the recalibrated 2022 predictions. Calibration intercepts and slopes describe performance in each test set; they were not applied to change the predictions. Model-performance intervals in both test sets used the patient-cluster resampling described in section 11.

The calibration intercept with slope fixed at 1 assessed calibration-in-the-large. A separate logistic model estimated a joint intercept and calibration slope. The prediction output was a probability of the composite recorded outcome, not a probability of preventable harm.

7. Full-Day and Annual Ranking Benchmarks

The full-day and annual ranking benchmarks were evaluated in 2023 only. We compared Random, ESI-first, Risk-only, and Risk-plus-window rules. ESI-first prioritized encounters from ESI level 1 through 5, with missing or invalid levels ranked last. Risk-only prioritized encounters by decreasing predicted event risk. Risk-plus-window multiplied this risk by the estimated probability that the recorded window would accommodate the review, conditional on an event. This second probability was modeled using training encounters with events, with model selection based on 2022 encounters with events. Recorded outcomes and realized window lengths were used to evaluate the rules but did not determine selection. At a selection cutoff, tied encounters shared the remaining review attempts fractionally. Random selection was evaluated using its expected performance.

We evaluated review durations of 5, 10, and 15 minutes and budgets of 25, 50, and 100 review-minutes per 100 eligible encounters. The main scenario used 10-minute reviews and 50 review-minutes per 100 encounters. Within each decision group, each rule received *K*=floor( *nM*/(100*B*)) review attempts, where *n* was the number of eligible 2023 encounters, *M* was the budget per 100 encounters, and *B* was the review duration. Each attempt consumed its full duration, even when the recorded window was too short to complete the review. We assumed that reviews began immediately at the recorded time zero, with no queues or delays.

For the full-day ranking benchmark, we distributed the total attempts across local calendar dates of time zero in proportion to eligible encounter counts. We rounded each day's proportional quota down, then assigned the remaining attempts to dates with the largest fractional remainders. Ties between dates were resolved chronologically. Each rule selected encounters within the resulting daily quotas, with no transfer of attempts between days. These quotas and rankings used completed encounter data and represented a retrospective benchmark. An annual-pooling sensitivity analysis allowed each rule to select encounters across the entire test year using the same total number of attempts.

We defined event reach as the number of selected encounters with an event and a recorded window at least as long as the review, divided by all encounters with an event in the same decision group. We also reported the proportion of encounters selected and the number of events reached per review attempt. Lift was event reach divided by the expected reach under Random selection at the same budget, review duration, and daily or annual allocation. Event reach measured opportunities to complete a review before the recorded endpoint; it did not measure events prevented or patient benefit.

The conditional window models were fitted separately for each decision group and review duration. Candidates were a smoothed constant, (*a*+1)/(*n*+2), where *a* of *n* training events had sufficient windows, and ridge logistic regression with *C*=0.1. Ridge was eligible only when both window classes contained at least 10 training events. Preprocessing used training encounters; model fitting used the event encounters. Predictors comprised the base and triage fields available by time zero, without individual numeric laboratory results or demographic characteristics other than age. Selection minimized 2022 log loss among event encounters, with ties favoring the constant. Intercept recalibration used 2022 event encounters when both window classes were present. Cumulative minima enforced probabilities that did not increase from 5 to 10 to 15 minutes.

Writing *s_i_* for the selected fraction, *Y_i_* for the event indicator, and *W_i_* for the capped window, event reach was

*R_B_*=(∑*_i_s_i_Y_i_*1(*W_i_*≥*B*))/(∑*_i_Y_i_*).

Selection rate was ∑*_i_* *s_i_*/*n*, and events reached per attempt was the numerator of *R_B_* divided by ∑*_i_* *s_i_*. Fractions represented expected selection under random tie resolution, not partial clinical reviews. Reach could differ from sensitivity of selection because a selected event with an insufficient window did not count as reached.

8. Prediction Sensitivities and Simple Comparators

We compared model performance with a constant prediction equal to the training-period event rate within each decision group. We also fitted models without the three triage fields and models without any laboratory predictors, including laboratory order and component counts. Model selection and intercept recalibration were repeated for each variant using the same training and tuning periods.

We evaluated the primary models among patients whose first encounter in the candidate cohort occurred in 2023, retaining all their eligible test encounters. We also reported performance among encounters whose composite outcome remained known when records without level-of-care information were treated as uncertain, for admission in 2023 and both decision groups in 2024. This was a subset defined by outcome ascertainment. Both subset analyses used the primary outcome labels and model predictions without refitting.

Three simple comparators were evaluated: the five-variable logistic model (Simple logistic), Age-first selection, and Prior-visits-first selection. Separate ridge logistic models for admission plans and discharge decisions used age, arrival-to-time-zero interval, prior completed adult ED visits within 90 days, ESI, and arrival mode. The inverse regularization strength was fixed at *C*=1. These models used the same training-only preprocessing, were fitted on 2020–2021 data, and underwent intercept recalibration on 2022 data. Age-first prioritized older patients, and Prior-visits-first prioritized higher visit counts, with missing values ranked last. All three comparators used the same eligible encounters, budgets, daily and annual allocation, and fractional tie handling as the primary rules. Prediction sensitivities and the simple model are shown in eTable 37 and eTable 38. Full-ranking annual intervals, contrasts, and capacity comparisons appear in eTable 47, eTable 48, eTable 49, and eTable 50.

9. Sequential Selection, Review Durations, and Start Delays

This analysis compared Random, ESI-first, Age-first, Prior-visits-first, Simple logistic, Risk-only, and Risk-plus-window under the same capacity and timing rules. Fitted risk and window models remained fixed. ESI ranked valid levels 1–5 before missing or invalid values; missing age or visit counts ranked last. Risk-plus-window used the probability that the window accommodated start delay plus review duration, conditional on an event. Window models were available for completion thresholds of 5, 10, and 15 minutes. Risk-plus-window was not estimated for a 20-minute completion threshold; no substitute model was used.

For each group and scenario, a threshold selected the highest-scoring *k*=ceil( *f* *n*_22_) eligible 2022 encounters, where *f* was the flag fraction. Score ties were resolved with a fixed seeded pseudorandom number assigned by encounter identifier (seed 20260930). The same full-precision score-and-tiebreak thresholds determined 2023 and 2024 flags. The observed flag fraction in a test year was reported without adjustment to match the tuning target. Random used the pseudorandom number alone. This differs from the expected-value Random calculation in the full-day and annual ranking benchmarks.

Daily capacity was *c*=round_half up_{floor( *n*_22_50/(100*B*))/365}, where *B* was review duration in minutes. For 10-minute reviews, 22,505 admission encounters yielded 1,125 tuning-year attempts and 3 reviews daily; 43,366 discharge encounters yielded 2,168 attempts and 6 daily. Planned 2023 capacities were 1,095 and 2,190. For 5-minute reviews, capacities were 6 and 12 daily, or 2,190 and 4,380 annually. For 2024, corresponding annual capacities over 366 days were 1,098 admission and 2,196 discharge attempts for 10-minute reviews, and 2,196 and 4,392 for 5-minute reviews. These were equal planned capacities across rules; attempts actually used could differ.

Flagged encounters were processed in exact recorded decision order within each local date. Simultaneous flagged decisions at the capacity cutoff shared remaining attempts fractionally. Capacity applied on all 365 calendar days in 2023 and all 366 in 2024, reset at midnight, and did not carry over. The main 10-minute scenario used a 5% tuning flag rate; the 5-minute scenario used 10%. A further 10-minute sensitivity used a 10% flag rate. Each scenario was evaluated at 0-, 5-, and 10-minute start delays. A selected event was reached only when its capped window was at least delay plus duration. Every attempt consumed a slot, including attempts with insufficient time. Neither actual window length nor outcomes selected encounters. Queues, interruptions, and available clinicians were not modeled.

We reported planned capacity, attempts used, events reached, and reach in each test year (eTable 16; eTable 17), unused capacity and observed flag rates for 2024 (eTable 17; eTable 18), and attempts per event for 2023 and for the primary 2024 comparison. The 5-minute scenario changed both capacity and thresholds, so differences from 10-minute reviews cannot be assigned solely to duration. A separate zero-delay Risk-only comparison used the 2022 95th-percentile threshold and compared sequential allocation with full-day ranking at constant capacity. This benchmark used a linear-quantile risk threshold without the main simulation's seeded score tiebreak (eTable 49; eTable 50).

10. Windows, Predicted Risk, and Outcome Association

Within each decision group in the 2023 temporal test set, we calculated the proportions of encounters with recorded windows of at least 5, 10, and 15 minutes, separately for encounters with and without the composite event. We reported event-minus-nonevent differences in percentage points. Confidence intervals for these differences came from paired calculations within the same patient-cluster bootstrap resamples. The 2024 window proportions were reported descriptively overall and by event status; window-association models and event-minus-nonevent intervals were not evaluated in 2024.

As a secondary analysis, we modeled the association between window length and the composite event using a Gaussian copula. The window distribution allowed point masses at zero and the cap, with a continuous distribution between them. We first estimated the marginal distributions and then the association parameter, *ρ*. We compared the fit with independence and fitted logistic regression with a natural cubic spline as a flexible alternative. These were descriptive analyses within the test cohort. Window distributions and association parameters were re-estimated in each bootstrap resample. We tested *ρ*=0 using two-sided Wald tests with bootstrap standard errors. Holm adjustment covered the two tests at the 60-minute cap, one for each decision group.

As a secondary description, we compared windows between high-risk and other encounters within each decision group. High risk was defined as a recalibrated prediction at or above the corresponding 2022 90th percentile. We reported differences in mean window length, capped at 60 minutes, and in the proportions with windows of at least 5, 10, and 15 minutes. We also estimated the Spearman correlation between predicted risk and capped window length.

We assessed how the composite risk score ranked each outcome component separately using AUROC, without refitting. Encounters with unknown or excluded component labels were omitted from that component's analysis.

For the Gaussian copula, the binary margin was the observed event prevalence. The window margin used observed probability masses at zero and the cap. Its interior was divided into 12 equal-width bins. Each interior count received one half-count before normalization to the total interior probability. The cumulative distribution was linear within each bin. With margins fixed at their first-stage estimates, we estimated *ρ* by the mixed-measure likelihood over [−0.98,0.98]. Endpoint contributions used copula rectangle probabilities; interior contributions used the corresponding derivative. Boundary estimates and nonestimable fits were recorded. The natural cubic spline comparator used knots at 0, 25%, 50%, 75%, and 100% of the cap and unpenalized logistic regression. These were comparisons of fit within the same test cohort, not evaluations of future prediction performance.

Bootstrap standard errors for *ρ* were the standard deviations of valid replicate estimates. Wald statistics divided the point estimate by its bootstrap standard error. Two-sided normal-reference tests and Holm adjustment were applied to the two primary 60-minute tests. Other caps and decision-source comparisons were secondary. All association parameters depended on the stated model; no causal or tail-dependence interpretation was assigned.

Individual-component ranking used the primary composite risk score, not a component-specific probability. Baseline and direct-critical-care states were excluded from critical-care component ranking even though those encounters could remain in the composite-outcome cohort. Window associations, Holm-adjusted tests, spline comparisons, predicted-risk associations, and component ranking are reported in eTable 51, eTable 52, eTable 53, eTable 54, and eTable 43.

11. Bootstrap Uncertainty and Interval Diagnostics

We used 2,000 patient-cluster bootstrap resamples with seed 20260930. Patients were sampled with replacement with all their eligible test encounters retained. Samples were shared across groups and paired comparisons. Fitted risk and availability models, recalibration, and sequential thresholds and capacities remained fixed. Selection was recomputed within each resample. Full-day and annual benchmarks also recomputed sample-dependent budgets and quotas. Window distributions and association models were re-estimated where applicable. Percentile 95% intervals required at least 1,900 finite estimates; they excluded uncertainty from model development and choice of analysis settings.

Nested diagnostics used 1,000 outer patient-cluster samples and 2,000 inner samples within each. Each sampled patient copy was a distinct cluster for inner resampling. Selection was repeated at both levels. Containment was assessed against the corresponding full-cohort estimate in each test year. Invalid intervals counted as failures; valid-only containment and Monte Carlo standard errors were also reported. The reference estimate was not known population truth.

For full-day and annual ranking, the diagnostic evaluated Random, ESI-first, Risk-only, and Risk-only minus ESI-first at 10-minute reviews and 50 review-minutes per 100 encounters (eTable 29; eFigure 6). Daily results are descriptive points. Annual intervals are shown for the evaluated rules and contrast, with the empirical qualification; they do not validate other budgets or the additional vital-sign intervals.

The sequential diagnostic evaluated Risk-only, Prior-visits-first, and their difference at zero delay for 10-minute/5%-flag and 5-minute/10%-flag scenarios in each group. All 12 statistics had 1,000 valid outer intervals in each year (eTable 27; eTable 28). Single-rule reach and attempts per event are presented as points. The evaluated Risk-only minus Prior-visits-first contrasts have nominal 95% intervals, with empirical coverage reported alongside them. The 2023 contrasts were exploratory. The primary 2024 contrast was discharge Risk-only minus Prior-visits-first reach for 10-minute reviews, 5% tuning flags, and zero delay. Replication required a difference above 0 with a 95% lower bound above 0; fewer than 1,900 finite bootstrap estimates would make this criterion not estimable. Discharge-window sufficiency at 10 minutes was descriptive and had no replication criterion. Other sequential contrasts and delay comparisons are descriptive because their interval coverage was not assessed. These diagnostics do not establish population coverage.

12. Subgroups, Case Mix, and Disclosure Control

Sequential allocation was described by age, sex, race, ethnicity, language, and interpreter need for 10-minute reviews, 5% tuning flags, and zero delay. For each decision group and rule, selection was performed once over all eligible encounters in that year, then summarized by subgroup. Capacity was not divided among subgroups. Nonmissing demographic categories with fewer than 200 training encounters were grouped as Other; missingness was separate. Estimates required at least 20 events and 11 encounters. Age groups were 18–39, 40–64, and 65–100 years. Recorded male, non-binary, and unknown sex were pooled for display, with missing sex separate. Other race categories in cohort tables reflect recorded categories; in allocation tables, Other can also include categories pooled by the training-frequency rule. No race categories were combined for privacy protection. Demographic labels came from recorded electronic health record fields without inference from names. Subgroup discrimination, calibration, and clinical benefit were not evaluated (eTable 40; eTable 41).

Subgroup selection and reach percentages were withheld when they could reveal a count of 1–10 or its complement. Complementary withholding jointly considered category encounter and event denominators, each rule's decision-group totals, and all displayed values. Rows were omitted when needed to protect other cells. The 2024 discharge Hispanic/Latino row was withheld for this reason. Table 1 pools male, non-binary, and unknown sex, omits the recorded White, non-Hispanic race row, and applies complementary withholding. The separately recorded 2024 Unknown language category and a complementary language cell are withheld.

Case mix was described across discharge decisions, admission plans with and without recorded handoff, handoff without an earlier plan, conflicts at discharge time zero, no qualifying status, and other invalid timing. These mutually exclusive groups included all candidates, before outcome and model exclusions. Final disposition was descriptive; excluded groups were not assigned event rates (eTable 55).

We compared recorded characteristics of eligible encounters with those of encounters with an unknown composite outcome. The unknown-outcome group included encounters that also failed temporal eligibility criteria. Encounters excluded solely by temporal eligibility criteria were not shown. Language and arrival-mode categories followed Table 1. Repeat encounters were retained, and patients could contribute to both groups. No hypothesis tests were performed.

Recorded ages were restricted to 18–100 years. Cohort tables used recorded age without imputation; predictive models used the training-derived clipping described in section 6. ESI values 1–5, invalid values, and missingness were distinct. Languages included the five most frequent training categories and Other, with missingness pooled into Other for presentation; a separately recorded Unknown category in 2024 was retained and withheld. For arrival mode, the five most frequent training categories were retained, with remaining and missing codes pooled. Arrival-mode labels came from the encounters table. Denominators included missing records. To protect patient privacy, counts of 1–10, and selected cells from which they could be calculated, are not shown (—). Corresponding percentages were also not shown. Timing quantiles required at least 11 contributing encounters. Zero counts are shown unless the cell or its row is withheld.

13. Analysis Details and Software

The 2020–2023 five-variable model, ESI-first, Age-first, Prior-visits-first, sequential allocation, recorded direction changes and resident-plan transitions, subgroup and case-mix descriptions, component ranking, window-model diagnostics, interval diagnostics, vital-sign sensitivity, and unknown-outcome comparison were post hoc analyses. The 2024 primary analyses and replication criterion are described in section 11. The analysis plan specified that no withheld count could be calculated from other published values; a small number of withheld counts can be calculated from published totals. Analyses used Python 3.14.7 (Python Software Foundation), NumPy 2.5.4, pandas 3.0.6, SciPy 1.18.1, scikit-learn 1.9.1, and DuckDB 1.5.6. Study plotting used Matplotlib 3.11.2. The vital-sign extraction used google-cloud-bigquery 3.46.2 and PyArrow 26.0.0.

14. Vital-Sign Sensitivity Analysis

In a sensitivity analysis, we added heart rate, systolic and diastolic blood pressure, respiratory rate, peripheral oxygen saturation, and temperature to the primary predictors. Vital signs were obtained from the same repository from the October 27, 2026 (UTC), source snapshot. This extraction was matched to the 258,247 eligible 2020–2023 encounters; cohort membership, study periods, time zero, and outcome definitions were unchanged. Encounters without usable measurements remained included.

Measurements came from the “Vitals” and “ED Tech Vitals” flowsheet templates. Template names did not independently establish validation or clinician availability. We matched patient and encounter identifiers through the crosswalk and subtracted each patient's date offset from the source timestamps. In a sample of 1,000 encounters, all patient and encounter identifiers matched, and all corrected arrival timestamps matched the recorded ED arrivals, with a maximum difference of 0 seconds. Every retained measurement also required matching patient and encounter identifiers and a corrected measurement time between that encounter's ED arrival and time zero, inclusive.

For each vital sign, we selected the earliest eligible recorded measurement across the two templates. Identical tied values collapsed to one value. An invalid or conflicting value at the earliest timestamp made that vital missing; a later measurement was not substituted. Measurements before recorded ED arrival or after time zero were excluded. Eligibility used recorded measurement time. Entry or verification timestamps were not available to establish when a value became visible to clinicians.

Oxygen saturation had a recorded percentage unit. For blank-unit entries, units were assigned from stored value distributions and formats: heart rate in beats/min, respiratory rate in breaths/min, blood pressure in mm Hg, and temperature in degrees Fahrenheit. The median temperature was 98.1 in “Vitals” and 97.9 in “ED Tech Vitals.” These distributions supported unit assumptions; they were not independent confirmation from a source dictionary. Paired blood pressures were parsed as systolic/diastolic values. Temperature was converted to degrees Celsius using (*F*−32)×5/9. Usable ranges were inclusive: heart rate, 20–300 beats/min; systolic pressure, 40–300 mm Hg; diastolic pressure, 20–200 mm Hg; respiratory rate, 3–80 breaths/min; oxygen saturation, 30–100%; and temperature, 25–45 °C. Nonfinite, unparsable, out-of-range, or conflicting first values remained missing. Coverage and measurement-to-time-zero intervals are reported in eTable 10.

A supplementary presence check examined the 72,714 encounters in the 2023 temporal test set for nonblank records in other templates during each exact arrival-to-time-zero window when the same vital had no nonblank record in the selected templates. Counts were tabulated separately by template and could overlap across templates. Presence did not establish a usable value, a confirmed unit, or availability to clinicians.

We repeated the primary training-only preprocessing, candidate model settings, 2022 log-loss selection, and 2022 intercept recalibration with the six vital signs and their missing-value indicators added. Performance was assessed in 2023 using AUROC, average precision, Brier score, log loss, observed-to-expected ratio, calibration-in-the-large, and calibration slope. We compared the sensitivity models with the primary predictions on the same encounters. Model selection and recalibration were repeated with the additional predictors; the comparison did not append vital signs to unchanged fitted models.

At the main scenario of 10-minute reviews and 50 review-minutes per 100 encounters, we evaluated Risk-only with vital signs, primary Risk-only, ESI-first, and Random selection using the same daily and annual allocation rules. Daily reach was descriptive. Annual reach contrasts compared Risk-only with vital signs against primary Risk-only and ESI-first. Confidence intervals for performance, paired performance differences, annual reach, and paired annual reach differences used 2,000 patient-cluster bootstrap resamples with seed 20260930. The same resamples were shared across decision groups and comparisons, fitted models remained fixed, and allocation was recomputed. Percentile 95% intervals required at least 1,900 finite estimates and did not include model-development uncertainty (eTable 11). The interval-coverage diagnostic did not evaluate these additional intervals. Counts from other templates are in eTable 56; these may overlap and do not establish usable measurements. Screening and missingness summaries are in eTable 57 and eTable 58. These allocation sensitivities use full-day and annual ranking, not sequential selection.

15. Recorded Resident-Plan Transitions

We separately identified each encounter's first resident-discharge (100030) and first resident-admission (106108) marker during ED care, including arrival and the earlier of departure or CDU entry. An encounter could enter both analyses. The next status was the earliest listed status at or after that resident marker, excluding the same resident code. Simultaneous statuses with the same outcome category retained it; conflicting categories were a tie. No subsequent listed status was kept as a separate category.

After resident discharge, readiness or conditional discharge represented agreement; attending admission or handoff represented the direction-change category; resident admission was a separate resident-revised-plan category. After resident admission, attending admission or handoff represented agreement; readiness or conditional discharge represented direction change; resident discharge was a separate resident-revised-plan category. For both, In Process, CDU admission, and transfer had separate categories (eTable 22). Thus direction change does not include all opposite-direction resident markers. Neither it nor return to In Process establishes an adjudicated override.

Category percentages used all resident-plan encounters as denominator, including ties and no later status. The time-zero-direction comparison was separate: it compared resident-marker direction with time-zero direction, excluding encounters with no usable decision time and discharge decisions with later reversal from its different-direction denominator (eTable 59). The time-zero decision could precede the resident marker. These percentages are not subsequent reversal rates. Absence of a resident marker does not establish absence of resident involvement. Timing summaries and the resident-discharge-to-decision interval are shown in eTable 34 and eTable 60.

We also compared each first resident-plan marker with the last qualifying status at or after it and at or before the earlier of ED departure and CDU entry. Repeated occurrences of the same resident-plan code were eligible only if strictly later than the first marker. Last statuses were classified both by direction relative to the resident plan and by status group. Simultaneous statuses with different groups retained a tie classification even when their direction agreed. Final ED disposition was tabulated separately, because CDU discharge and other events after handoff can differ from the recorded plan. For encounters whose next status was In Process, we classified the next qualifying status other than In Process as same direction, opposite direction, another status, tie, or no later decision (eTable 24; eTable 25; eTable 26). Residents may enter disposition statuses before or after discussing the patient with the attending. Thus neither next-status nor last-status transitions identify or provide a bound on changes attributable to attending review.

16. Eventual Recorded Direction Changes

This 2020–2023 description used all candidate encounters with a qualifying marker, without requiring model eligibility. Markers had to be timed and occur from arrival through the earlier of ED departure and CDU entry. The first admission-plan marker (106108 or 106116) was compared with later readiness or conditional-discharge markers (6 or 106111). The first discharge-decision marker was compared with later admission-plan or handoff markers (106108, 106116, 106107, or 10). A marker at exactly the anchor time was a tie, reported separately even if a later opposite-direction marker existed. Admission-to-discharge changes were stratified by whether they occurred at or before the first handoff, or after handoff. Results were reported by year and pooled (eTable 42). These denominators differ from the discharge-priority time-zero groups. A change could reflect pending test results or other clinical developments and does not establish a second review.

17. Conditional Window-Model Diagnostics

For each decision group and 5-, 10-, and 15-minute completion threshold, we evaluated the fitted window model's AUROC for sufficient time among 2023 encounters with composite events. Corresponding results among all eligible 2023 encounters were secondary. We described predicted probabilities by their mean and 5th, 25th, 50th, 75th, and 95th percentiles. For 10-minute sequential reviews with 5% tuning flags and zero delay, we compared Risk-only and Risk-plus-window score ranks using Spearman correlation and tabulated selections shared by or unique to each rule (eTable 13; eTable 14). Fractional selections retained the allocation weights. These diagnostics were not repeated in 2024.

eResults

Time-Zero Assignments and Exclusions

Among 284,627 candidate encounters, 179,798 had a discharge decision, 83,594 had an admission plan with recorded handoff, and 1,085 had an admission plan without handoff. The 20,150 exclusions for no usable decision time comprised 13,775 handoffs without an earlier plan, 790 conflicts at discharge time zero, 5,563 encounters without a qualifying status, and 22 with other invalid timing (eTable 2; eTable 3; eTable 55). These are populations by decision-time group before outcome and temporal exclusions. No encounters met the separate CDU-placement definition.

Among admission plans without handoff, 877 of 1,085 ended in transfer. Final discharge occurred in 9,188 admission-group encounters, including 8,167 with a CDU stay; final disposition did not reassign them. Of 13,775 handoff-without-plan exclusions, 2,528 had CDU entry, including 2,445 with only handoff before entry (eTable 4). The 9,697 discharge decisions preceded by an admission plan include both resident and attending admission markers.

Recorded Decision Intervals

Across 2020–2023, uncapped admission-plan-to-handoff time was 104 minutes (IQR, 49–186), and handoff-to-departure time was 160 minutes (IQR, 95–347), among 83,594 encounters with recorded handoff. The latter interval is not part of the review window. Discharge decision-to-window-end time was 22 minutes (IQR, 12–41) among 179,798 encounters (eTable 34). Capped windows and the relationship between resident-discharge markers, readiness, and conditional discharge are reported separately (eTable 44; eTable 60; eTable 46). These timing populations differ from model-eligible 2023 encounters.

Outcome Ascertainment

Among 2023 encounters with usable decision times, before final temporal eligibility exclusions, the sensitivity counting qualifying timed ED-expired records classified 1,130 admission-group composite events, compared with 1,112 under the primary definition. Fewer than 11 discharge-group encounters changed classification; exact counts are withheld. This sensitivity population included uncertain primary outcomes and is not the final model cohort. The ordinary composite and component ascertainment appear in eTable 5 and eTable 6.

Sequential Interval Diagnostics

All 12 evaluated statistics had 1,000 valid outer intervals. Coverage of the eight single-rule reach intervals ranged from 34.9% to 75.7%, with every miss below the full-cohort estimate. For Risk-only minus Prior-visits-first, 10-minute coverage was 94.8% for admission and 96.0% for discharge; 5-minute coverage was 93.1% and 93.2%. Corresponding 10-minute contrast misses below/above the reference were 47/5 and 7/33; 5-minute misses were 68/1 and 0/68 (eTable 27). These are counts of resampling intervals, not patients. Containment of an empirical reference does not establish population coverage. The 2023 contrast intervals are exploratory, and other sequential contrasts and delays are descriptive (eTable 35).

Vital-Sign Sensitivity

In 2023, usable first measurements were available for 92.78%–96.54% of admission and 95.11%–97.61% of discharge encounters, depending on the vital sign. No usable predecision vital was available for 636 admission encounters (2.61%) and 974 discharge encounters (2.02%) (eTable 10; eTable 58). Record presence in excluded templates does not establish usable additional measurements; counts can overlap and cannot be summed into a unique-encounter total (eTable 56).

Both sensitivity models selected gradient-boosted trees: a maximum of 15 leaves per tree and 100 boosting iterations for admission, and 7 leaves and 250 iterations for discharge. Both primary models also used trees. All 54 reported intervals had 2,000 finite resamples. Calibration slopes were 1.012 for the primary and 1.022 for the sensitivity admission model, and 0.994 and 1.009 for discharge. Annual reach differences from primary models included zero in both groups. These comparisons do not establish equivalence; model selection and recalibration were repeated with the additional predictors (eTable 11).

Recorded Resident-Plan Transitions

In the pooled 2020–2023 population, the direction-change category comprised 210 of 75,077 encounters with a resident-discharge marker and 361 of 25,314 encounters with a resident-admission marker. An opposite-direction resident marker was the next qualifying status for 148 resident discharge plans and 617 resident admission plans (eTable 22; eTable 23). The different-time-zero-direction proportions, 0.98% and 7.62%, used different denominators and could compare against an earlier time-zero decision; they are not subsequent reversal rates (eTable 59).

Other Analyses

The full-day and annual ranking benchmarks are shown across durations and budgets in eTable 19, eTable 20, eTable 21, eTable 47, and eTable 48, with corresponding plots in eFigure 7, eFigure 8, eFigure 9, eFigure 10, and eFigure 11. Window descriptions before model exclusions appear in eTable 45 and eFigure 12. Model-based window associations and risk-window summaries appear in eTable 51, eTable 52, eTable 53, eTable 54, and eFigure 13. These are associations, not causal effects. Component ranking uses the composite risk score without component-specific refitting (eTable 43).

2024 Held-Out Temporal Test Set

Of 83,002 candidates aged 18–100 years, 5,458 had no usable decision time. There were 77,544 recorded decisions, 442 unknown composite outcomes, and 16 further timing exclusions, leaving 77,086 eligible encounters in 51,587 patients. The cohort included 26,411 admission-plan and 50,675 discharge-decision encounters (Figure 1). Window proportions at 5, 10, and 15 minutes are in Table 2. The clinical characteristics by decision group are in eTable 33; the four-period cohort characteristics are in Table 1.

Risk-only minus Prior-visits-first discharge reach for the primary 10-minute scenario was 3.66 percentage points (nominal 95% CI, −0.18 to 6.26), failing the replication criterion. Both rules used the same planned capacity of 2,196 attempts; Risk-only used 2,018 and Prior-visits-first 2,081. All sequential scenarios and contrasts appear in eTable 17 and eTable 36. Flag fractions are reported without resetting thresholds to match the 2022 target (eTable 18).

The 2024 nested diagnostic had 1,000 valid intervals for every evaluated statistic. At 10 minutes, empirical coverage of the Risk-only minus Prior-visits-first contrast was 93.4% for discharge and 85.6% for admission. At 5 minutes, it was 97.6% and 96.9%, respectively. For discharge at 10 minutes, single-rule coverage was 51.1% for Risk-only and 75.9% for Prior-visits-first; for admission it was 16.7% and 46.2%. These diagnostics support reporting single-rule reach as descriptive points and qualify the nominal contrast intervals (eTable 28).

Primary, simple, and sensitivity model performance in 2024 is described in eTable 39, with primary-model calibration plotted in eFigure 5. All models and 2022 recalibration adjustments remained fixed. The vital-sign sensitivity, full-day and annual ranking benchmarks, and window-association models were not evaluated in 2024. Sequential subgroup results use full-population selection within each decision group (eTable 41).

Eventual Changes and Window-Model Diagnostics

In 2020–2023, 12,070 of 96,249 admission-plan encounters (12.54%) had a later discharge decision, whereas 2,400 of 183,127 discharge-decision encounters (1.31%) had a later admission-plan or handoff marker. Simultaneous opposite-direction markers were classified as ties, not later changes. Admission-to-discharge changes included 10,085 at or before first handoff or without handoff, and 1,985 after handoff (eTable 42). These counts differ from the 9,697 discharge-priority decisions preceded by an admission plan because the marker and time-zero definitions differ.

The last-status direction-change category included 1,176 resident-discharge and 2,233 resident-admission encounters. The next-status, last-status, final-disposition and In Process descriptions use distinct definitions and should not be combined into an attending-review rate (eTable 23; eTable 24; eTable 25; eTable 26).

Among 2023 discharge encounters with events, the 10-minute window model's AUROC was 0.750 and its 5th–95th probability percentiles were 0.761–0.984. Among all discharge encounters, those percentiles were 0.731–0.981. Risk-only and Risk-plus-window shared 1,750 selections, out of 1,857 and 1,856 selections, respectively; their score-rank correlation was 0.997 (eTable 13; eTable 14). The model discriminated sufficient windows among event encounters, but window weighting changed relatively few selections.

| eTable 1. Operational Codes | | |
| --- | --- | --- |
| Field | Code | Use in this study |
| First ED department | 110100019 | Stanford adult ED candidate and return pool. |
| Patient status | 106108 | Resident Admit Dispo Selected; admission-plan marker. |
| Patient status | 106116 | Decision to Admit Dispo Selected (Attending); admission-plan marker. |
| Patient status | 100030 | Resident-discharge marker; transition analysis only, not time zero. |
| Patient status | 106111 | Conditional discharge; qualifying discharge decision. |
| Patient status | 6 | Ready for discharge; qualifying discharge decision. |
| Patient status | 106107 | Signed admission order; recorded handoff marker. |
| Patient status | 10 | Ready for admission; recorded handoff marker. |
| Care area | 41, 42 | CDU entry used in determining the window endpoint. |
| Care area | 10616011 | CDU entry used in determining the window endpoint (CDU overflow; first recorded in 2024). |
| Level of care | 5 | Acute care. |
| Level of care | 9 | Intermediate care. |
| Level of care | 8 | Critical care. |
| ED final disposition | 2 | Return ending in inpatient admission. |
| ED final disposition | 25 | Observation; not a return-admission event. |
| ED final disposition | 18 | Encounter entered in error; excluded from return and prior-visit counts. |
| ED final disposition | 32 | Discharge to police custody; first recorded in 2024; not a return-admission event. |
| Patient status | 106031 | ED expired; positive death evidence only in sensitivity analysis. |
| ADT event subtype | 1, 3 | Original or updated event. |
| ADT event type | 1, 3, 5, 6, 7 | Admission, transfer in, patient update, census, or hospital outpatient event; eligible level updates. |
| ADT event type | 2, 4 | Discharge or transfer out; closes the active state when no simultaneous eligible level update takes precedence. |
| Laboratory verification | 9, Verified | Required code and label for numeric laboratory values. |
| Patient status | 11 | In Process; separate transition category. |
| Patient status | 106109 | CDU admission; separate transition category. |
| Patient status | 14, 106114, 106115 | Transfer statuses; separate transition category. |

Recorded admission-status labels do not independently establish the entering clinician; status timestamps do not independently establish transfer of responsibility. In local workflow, codes 6 and 106111 can only be entered by attending physicians; the individual user was not analyzed. Code 106107 may approximate accepting-team discussion; the other endpoints do not establish its timing or actual transfer of responsibility. Abbreviations: ADT, admission, discharge, and transfer; CDU, clinical decision unit; ED, emergency department.

| eTable 2. Time-Zero Assignment by Year | | |
| --- | --- | --- |
| Year | Decision-time group | Encounters |
| 2020 | Discharge decision | 42,647 |
| 2020 | Admission plan with recorded handoff | 16,788 |
| 2020 | Admission plan without recorded handoff | 230 |
| 2020 | Excluded: handoff without earlier plan | 2,786 |
| 2020 | Excluded: admission plan or handoff at the discharge decision time | — |
| 2020 | Excluded: no qualifying status | 1,224 |
| 2020 | Excluded: departure before arrival | — |
| 2021 | Discharge decision | 42,908 |
| 2021 | Admission plan with recorded handoff | 19,772 |
| 2021 | Admission plan without recorded handoff | 239 |
| 2021 | Excluded: handoff without earlier plan | 3,336 |
| 2021 | Excluded: admission plan or handoff at the discharge decision time | — |
| 2021 | Excluded: no qualifying status | 1,539 |
| 2021 | Excluded: departure before arrival | — |
| 2022 | Discharge decision | 45,371 |
| 2022 | Admission plan with recorded handoff | 22,813 |
| 2022 | Admission plan without recorded handoff | 304 |
| 2022 | Excluded: handoff without earlier plan | 3,660 |
| 2022 | Excluded: admission plan or handoff at the discharge decision time | — |
| 2022 | Excluded: no qualifying status | 1,768 |
| 2022 | Excluded: departure before arrival | — |
| 2023 | Discharge decision | 48,872 |
| 2023 | Admission plan with recorded handoff | 24,221 |
| 2023 | Admission plan without recorded handoff | 312 |
| 2023 | Excluded: handoff without earlier plan | 3,993 |
| 2023 | Excluded: admission plan or handoff at the discharge decision time | — |
| 2023 | Excluded: no qualifying status | 1,032 |
| 2023 | Excluded: departure before arrival | — |

All candidate encounters before outcome and temporal eligibility exclusions. To protect patient privacy, counts of 1–10, and selected cells from which they could be calculated, are not shown (—).

| eTable 3. Decision-Time Group Denominators | | |
| --- | --- | --- |
| Decision-time group | Encounters | Candidates % |
| Discharge decision | 179,798 | 63.2 |
| Admission plan with recorded handoff | 83,594 | 29.4 |
| Admission plan without recorded handoff | 1,085 | 0.4 |
| Handoff without earlier plan | 13,775 | 4.8 |
| Admission plan or handoff at the discharge decision time | 790 | 0.3 |
| No qualifying status | 5,563 | 2.0 |
| Other invalid timing | 22 | 0.0 |

All 284,627 candidates during 2020–2023. Groups are mutually exclusive and precede model eligibility exclusions.

| eTable 4. Recorded Reversals, Missing Handoffs, and Observation Stays | |
| --- | --- |
| Measure | Encounters |
| Discharge decisions | 179,798 |
| Discharge decisions preceded by an admission plan | 9,697 |
| Discharge decisions followed by a later admission plan (window ends there) | 1,413 |
| Discharge decisions followed by a later admission plan and handoff at the same time (window ends there) | 15 |
| Discharge decisions followed by a later handoff (window ends there) | 646 |
| Admission plans | 84,679 |
| Admission plans without a recorded handoff | 1,085 |
| of which final disposition was transfer | 877 |
| Admission plans with a final discharge | 9,188 |
| of which had a CDU location during the ED stay | 8,167 |
| Excluded: handoff without earlier plan | 13,775 |
| of which entered the CDU | 2,528 |
| of which had only a handoff before CDU entry | 2,445 |
| CDU entry without a qualifying discharge decision, admission plan, or handoff through entry | 0 |

All 2020–2023 candidates with the stated decision-time group. Final disposition is descriptive. A reversal is not itself a composite event. No encounters met the CDU-placement definition.

| eTable 5. Electronic Outcome Ascertainment | | | | | |
| --- | --- | --- | --- | --- | --- |
| Decision group | Outcome | Event | Excluded | No recorded event | Unknown |
| Admission | Critical care within 24 hours | 1,278 | 5,159 | 78,101 | 141 |
| Admission | Death within 7 days | 795 | 0 | 83,470 | 414 |
| Admission | Return ending in admission | 2,021 | 0 | 82,635 | 23 |
| Discharge | Critical care within 24 hours | — | — | — | — |
| Discharge | Death within 7 days | — | — | — | — |
| Discharge | Return ending in admission | 3,456 | 0 | 176,212 | 130 |

All 2020–2023 encounters with qualifying decisions before model exclusions. The composite comprised an adult ED return arriving within 7 days of time zero and ultimately ending in inpatient admission, qualifying critical care within 24 hours, or death within 7 days. Unknown components were not recoded as absent. Baseline or direct critical care did not satisfy the critical-care component but did not prevent other components from establishing an event. To protect patient privacy, counts of 1–10, and selected cells from which they could be calculated, are not shown (—).

| eTable 6. Composite Outcome Coverage | | | | | |
| --- | --- | --- | --- | --- | --- |
| Year | Decision group | Decision-time group | No recorded event | Event | Unknown |
| 2020 | Admission | Recorded decision | 16,083 | 849 | 86 |
| 2020 | Discharge | Recorded decision | 41,609 | 701 | 337 |
| 2020 | No time zero | No usable decision time | 0 | 0 | 4,233 |
| 2021 | Admission | Recorded decision | 18,935 | 977 | 99 |
| 2021 | Discharge | Recorded decision | 41,549 | 815 | 544 |
| 2021 | No time zero | No usable decision time | 0 | 0 | 5,041 |
| 2022 | Admission | Recorded decision | 21,939 | 1,065 | 113 |
| 2022 | Discharge | Recorded decision | 43,331 | 932 | 1,108 |
| 2022 | No time zero | No usable decision time | 0 | 0 | 5,624 |
| 2023 | Admission | Recorded decision | — | — | — |
| 2023 | Discharge | Recorded decision | — | — | — |
| 2023 | No time zero | No usable decision time | 0 | 0 | 5,252 |

Populations by decision-time group before model eligibility exclusions. The composite comprised an adult ED return arriving within 7 days of time zero and ultimately ending in inpatient admission, qualifying critical care within 24 hours, or death within 7 days. Encounters with no usable decision time were not assigned outcomes. To protect patient privacy, counts of 1–10, and selected cells from which they could be calculated, are not shown (—).

| eTable 7. Candidate Numeric Laboratory Components | | |
| --- | --- | --- |
| Component | Source identifier | Recorded unit |
| White blood cells | 1230100000 | K/uL |
| Hemoglobin | 1230100002 | g/dL |
| Platelets | 1230100008 | K/uL |
| Sodium | 1230100706 | mmol/L |
| Potassium | 1230100707 | mmol/L |
| Glucose | 1230100708 | mg/dL |
| Blood urea nitrogen | 1230100781 | mg/dL |
| Creatinine | 1230100709 | mg/dL |
| Chloride | 1230100729 | mmol/L |
| Total carbon dioxide | 1230100717 | mmol/L |
| Calcium | 1230100782 | mg/dL |
| Albumin | 1230100715 | g/dL |
| Aspartate aminotransferase | 1230100714 | U/L |
| Alanine aminotransferase | 1230100713 | U/L |
| Total bilirubin | 1230100711 | mg/dL |

Candidate components were retained only when the training availability criteria were met. Units were not converted.

| eTable 8. Numeric Laboratory Predictor Availability in Training, 2020–2021 | | | | |
| --- | --- | --- | --- | --- |
| Decision group | Laboratory component | Recorded unit | Available values | Encounters |
| Admission | WHITE BLOOD CELLS (WBC) | K/uL | 22,750 | 36,417 |
| Admission | HEMOGLOBIN (HGB) | g/dL | 22,739 | 36,417 |
| Admission | PLATELET COUNT (PLT) | K/uL | 22,735 | 36,417 |
| Admission | SODIUM | mmol/L | 17,717 | 36,417 |
| Admission | POTASSIUM | mmol/L | 17,701 | 36,417 |
| Admission | GLUCOSE | mg/dL | 17,717 | 36,417 |
| Admission | BLOOD UREA NITROGEN (BUN) | mg/dL | 17,715 | 36,417 |
| Admission | CREATININE | mg/dL | 17,688 | 36,417 |
| Admission | CHLORIDE | mmol/L | 17,689 | 36,417 |
| Admission | CO2 | mmol/L | 17,688 | 36,417 |
| Admission | CALCIUM | mg/dL | 17,691 | 36,417 |
| Admission | ALBUMIN | g/dL | 17,490 | 36,417 |
| Admission | AST (SGOT) | U/L | 17,487 | 36,417 |
| Admission | ALT (SGPT) | U/L | 17,465 | 36,417 |
| Admission | BILIRUBIN, TOTAL | mg/dL | 16,870 | 36,417 |
| Discharge | WHITE BLOOD CELLS (WBC) | K/uL | 42,684 | 83,245 |
| Discharge | HEMOGLOBIN (HGB) | g/dL | 42,680 | 83,245 |
| Discharge | PLATELET COUNT (PLT) | K/uL | 42,667 | 83,245 |
| Discharge | SODIUM | mmol/L | 41,447 | 83,245 |
| Discharge | POTASSIUM | mmol/L | 41,460 | 83,245 |
| Discharge | GLUCOSE | mg/dL | 41,445 | 83,245 |
| Discharge | BLOOD UREA NITROGEN (BUN) | mg/dL | 41,449 | 83,245 |
| Discharge | CREATININE | mg/dL | 41,401 | 83,245 |
| Discharge | CHLORIDE | mmol/L | 41,401 | 83,245 |
| Discharge | CO2 | mmol/L | 41,400 | 83,245 |
| Discharge | CALCIUM | mg/dL | 41,401 | 83,245 |
| Discharge | ALBUMIN | g/dL | 40,597 | 83,245 |
| Discharge | AST (SGOT) | U/L | 40,594 | 83,245 |
| Discharge | ALT (SGPT) | U/L | 40,570 | 83,245 |
| Discharge | BILIRUBIN, TOTAL | mg/dL | 38,297 | 83,245 |

Training encounters within each decision group. Retention required at least 500 available values and at least 1% of group encounters. Laboratory units were not converted.

| eTable 9. Predictor Missingness in 2020–2023 | | | |
| --- | --- | --- | --- |
| Predictor | 2020–2021 | 2022 | 2023 |
| Age | 0 | 0 | 0 |
| Arrival mode | 0.07 | 0.09 | 0.06 |
| Available laboratory components | 0 | 0 | 0 |
| Chief-complaint code | 0.21 | 0.17 | 0.04 |
| Decision-record type | 0 | 0 | 0 |
| Arrival-to-decision time | 0 | 0 | 0 |
| Emergency Severity Index | 1 | 0.45 | 0.41 |
| Days of available history | 0 | 0 | 0 |
| Decision direction (admission or discharge) | 0 | 0 | 0 |
| Laboratory orders | 0 | 0 | 0 |
| Decision hour (cosine) | 0 | 0 | 0 |
| Decision hour (sine) | 0 | 0 | 0 |
| Decision month | 0 | 0 | 0 |
| Decision weekday | 0 | 0 | 0 |
| Prior ED visits in 90 days | 10.8 | 0 | 0 |

Values are percentages among retained recorded-decision encounters, including those with unknown outcomes. Individual laboratory-value availability is shown separately. Triage availability at time zero was assumed.

| eTable 10. Usable First Vital Signs and Measurement-to-Decision Intervals in 2020–2023 | | | | | |
| --- | --- | --- | --- | --- | --- |
| Year | Decision group | Vital sign | Encounters | Usable, No. (%) | Minutes, median (IQR) |
| 2020 | Admission | Heart rate | 16,932 | 15,786 (93.23) | 104 (52–176) |
| 2020 | Admission | Systolic blood pressure | 16,932 | 15,655 (92.46) | 105 (52–177) |
| 2020 | Admission | Diastolic blood pressure | 16,932 | 15,652 (92.44) | 105 (52–177) |
| 2020 | Admission | Respiratory rate | 16,932 | 15,722 (92.85) | 104 (52–176) |
| 2020 | Admission | Peripheral oxygen saturation | 16,932 | 15,671 (92.55) | 104 (52–177) |
| 2020 | Admission | Temperature | 16,932 | 14,842 (87.66) | 107 (53–179) |
| 2020 | Discharge | Heart rate | 42,310 | 39,476 (93.30) | 138 (40–236) |
| 2020 | Discharge | Systolic blood pressure | 42,310 | 37,402 (88.40) | 146 (48–242) |
| 2020 | Discharge | Diastolic blood pressure | 42,310 | 37,396 (88.39) | 146 (48–242) |
| 2020 | Discharge | Respiratory rate | 42,310 | 39,257 (92.78) | 138 (40–237) |
| 2020 | Discharge | Peripheral oxygen saturation | 42,310 | 39,340 (92.98) | 138 (40–236) |
| 2020 | Discharge | Temperature | 42,310 | 38,093 (90.03) | 137 (38–236) |
| 2021 | Admission | Heart rate | 19,485 | 18,472 (94.80) | 108 (54–184) |
| 2021 | Admission | Systolic blood pressure | 19,485 | 18,363 (94.24) | 108 (54–184) |
| 2021 | Admission | Diastolic blood pressure | 19,485 | 18,357 (94.21) | 108 (54–184) |
| 2021 | Admission | Respiratory rate | 19,485 | 18,386 (94.36) | 108 (54–184) |
| 2021 | Admission | Peripheral oxygen saturation | 19,485 | 18,342 (94.13) | 108 (54–184) |
| 2021 | Admission | Temperature | 19,485 | 17,403 (89.31) | 111 (57–187) |
| 2021 | Discharge | Heart rate | 40,935 | 38,899 (95.03) | 184 (100–277) |
| 2021 | Discharge | Systolic blood pressure | 40,935 | 36,774 (89.84) | 193 (115–283) |
| 2021 | Discharge | Diastolic blood pressure | 40,935 | 36,770 (89.83) | 193 (115–283) |
| 2021 | Discharge | Respiratory rate | 40,935 | 38,749 (94.66) | 184 (100–277) |
| 2021 | Discharge | Peripheral oxygen saturation | 40,935 | 38,853 (94.91) | 184 (99–277) |
| 2021 | Discharge | Temperature | 40,935 | 37,507 (91.63) | 186 (100–278) |
| 2022 | Admission | Heart rate | 22,505 | 21,328 (94.77) | 118 (59–208) |
| 2022 | Admission | Systolic blood pressure | 22,505 | 21,254 (94.44) | 118 (59–208) |
| 2022 | Admission | Diastolic blood pressure | 22,505 | 21,244 (94.40) | 118 (60–208) |
| 2022 | Admission | Respiratory rate | 22,505 | 21,205 (94.22) | 118 (59–208) |
| 2022 | Admission | Peripheral oxygen saturation | 22,505 | 21,234 (94.35) | 118 (59–208) |
| 2022 | Admission | Temperature | 22,505 | 20,134 (89.46) | 121 (62–212) |
| 2022 | Discharge | Heart rate | 43,366 | 41,421 (95.51) | 221 (136–323) |
| 2022 | Discharge | Systolic blood pressure | 43,366 | 40,445 (93.26) | 224 (142–325) |
| 2022 | Discharge | Diastolic blood pressure | 43,366 | 40,437 (93.25) | 224 (142–325) |
| 2022 | Discharge | Respiratory rate | 43,366 | 41,283 (95.20) | 221 (135–323) |
| 2022 | Discharge | Peripheral oxygen saturation | 43,366 | 41,392 (95.45) | 220 (135–322) |
| 2022 | Discharge | Temperature | 43,366 | 39,809 (91.80) | 223 (138–325) |
| 2023 | Admission | Heart rate | 24,401 | 23,557 (96.54) | 129 (66–229) |
| 2023 | Admission | Systolic blood pressure | 24,401 | 23,477 (96.21) | 130 (66–229) |
| 2023 | Admission | Diastolic blood pressure | 24,401 | 23,470 (96.18) | 130 (66–229) |
| 2023 | Admission | Respiratory rate | 24,401 | 23,428 (96.01) | 130 (66–229) |
| 2023 | Admission | Peripheral oxygen saturation | 24,401 | 23,447 (96.09) | 129 (66–229) |
| 2023 | Admission | Temperature | 24,401 | 22,640 (92.78) | 132 (68–231) |
| 2023 | Discharge | Heart rate | 48,313 | 47,157 (97.61) | 242 (155–348) |
| 2023 | Discharge | Systolic blood pressure | 48,313 | 47,120 (97.53) | 242 (155–349) |
| 2023 | Discharge | Diastolic blood pressure | 48,313 | 47,116 (97.52) | 242 (155–349) |
| 2023 | Discharge | Respiratory rate | 48,313 | 47,054 (97.39) | 242 (155–348) |
| 2023 | Discharge | Peripheral oxygen saturation | 48,313 | 47,057 (97.40) | 242 (154–348) |
| 2023 | Discharge | Temperature | 48,313 | 45,949 (95.11) | 243 (156–350) |

All eligible encounters were retained. First measurements were between arrival and time zero, inclusive. Invalid first values were not replaced. Minutes describe time from a usable first measurement to time zero, rounded to whole minutes. Recorded measurement time does not establish clinician visibility.

| eTable 11. Vital-Sign Sensitivity: Prediction and Ranking Allocation in 2023 | | | |
| --- | --- | --- | --- |
| Measure | Model or comparison | Admission plans | Discharge decisions |
| AUROC | Primary model | 0.659 (0.641 to 0.678) | 0.788 (0.773 to 0.802) |
| AUROC | Model with vitals | 0.671 (0.653 to 0.688) | 0.787 (0.771 to 0.802) |
| AUROC | With vitals minus primary | 0.011 (0.005 to 0.018) | −0.001 (−0.007 to 0.005) |
| Average precision | Primary model | 0.097 (0.085 to 0.113) | 0.093 (0.081 to 0.108) |
| Average precision | Model with vitals | 0.103 (0.091 to 0.119) | 0.096 (0.083 to 0.112) |
| Average precision | With vitals minus primary | 0.006 (−0.003 to 0.016) | 0.003 (−0.002 to 0.010) |
| Brier score | Primary model | 0.043 (0.040 to 0.045) | 0.021 (0.019 to 0.022) |
| Brier score | Model with vitals | 0.042 (0.040 to 0.045) | 0.021 (0.019 to 0.022) |
| Brier score | With vitals minus primary | −0.00015 (−0.00029 to −0.00003) | −0.00004 (−0.00010 to 0.00003) |
| Log loss | Primary model | 0.178 (0.169 to 0.186) | 0.093 (0.088 to 0.098) |
| Log loss | Model with vitals | 0.176 (0.168 to 0.185) | 0.093 (0.088 to 0.098) |
| Log loss | With vitals minus primary | −0.0012 (−0.0020 to −0.0005) | −0.0001 (−0.0006 to 0.0004) |
| Observed-to-expected ratio | Primary model | 1.006 (0.941 to 1.067) | 1.037 (0.964 to 1.107) |
| Observed-to-expected ratio | Model with vitals | 0.992 (0.929 to 1.053) | 1.024 (0.951 to 1.092) |
| Observed-to-expected ratio | With vitals minus primary | −0.013 (−0.018 to −0.009) | −0.013 (−0.020 to −0.007) |
| Calibration-in-the-large | Primary model | 0.006 (−0.065 to 0.069) | 0.039 (−0.039 to 0.109) |
| Calibration-in-the-large | Model with vitals | −0.008 (−0.078 to 0.056) | 0.025 (−0.054 to 0.094) |
| Calibration-in-the-large | With vitals minus primary | −0.014 (−0.019 to −0.009) | −0.014 (−0.020 to −0.007) |
| Calibration slope | Primary model | 1.012 (0.901 to 1.128) | 0.994 (0.934 to 1.052) |
| Calibration slope | Model with vitals | 1.022 (0.922 to 1.126) | 1.009 (0.951 to 1.068) |
| Calibration slope | With vitals minus primary | 0.009 (−0.037 to 0.058) | 0.015 (−0.006 to 0.037) |
| Daily event reach, % | Risk-only with vitals | 16.37 | 24.06 |
| Daily event reach, % | Primary Risk-only | 14.39 | 23.69 |
| Daily event reach, % | ESI-first | 9.81 | 6.70 |
| Daily event reach, % | Random | 4.80 | 4.20 |
| Annual event reach, % | Risk-only with vitals | 15.83 (13.84 to 18.09) | 24.72 (22.11 to 27.36) |
| Annual event reach, % | Primary Risk-only | 14.39 (12.25 to 16.37) | 25.00 (22.20 to 27.82) |
| Annual event reach, % | ESI-first | 9.89 (8.69 to 11.29) | 6.69 (5.95 to 7.48) |
| Annual event reach, % | Random | 4.81 (4.75 to 4.86) | 4.20 (4.08 to 4.31) |
| Annual reach difference, percentage points | With vitals minus primary | 1.44 (−0.18 to 3.52) | −0.28 (−1.91 to 1.49) |
| Annual reach difference, percentage points | With vitals minus ESI-first | 5.94 (3.85 to 8.30) | 18.03 (15.46 to 20.60) |

Sensitivity analysis. Allocation uses 10-minute reviews and 50 review-minutes per 100 encounters. Parentheses contain 95% patient-cluster percentile confidence intervals from 2,000 resamples; at least 1,900 finite estimates were required. Fitted prediction models remained fixed. Model selection and recalibration were repeated with six vital signs added. Both primary and sensitivity models used gradient-boosted trees. Daily and annual rows refer to full-day/annual ranking, not sequential selection. Daily comparisons are descriptive. Brier differences use five decimal places and log-loss differences four. Event reach is the proportion of all composite-event encounters selected with sufficient recorded review time. Review attempts without enough recorded time still count against the budget. Reach does not measure events prevented.

| eTable 12. Conditional Window-Model Performance in 2022 | | | | |
| --- | --- | --- | --- | --- |
| Decision group | Review (min) | Tuning events | Observed sufficient (%) | Predicted sufficient (%) |
| Admission | 5 | 1,041 | 98.94 | 98.94 |
| Admission | 10 | 1,041 | 97.12 | 97.12 |
| Admission | 15 | 1,041 | 95.49 | 95.44 |
| Discharge | 5 | 907 | 95.59 | 95.59 |
| Discharge | 10 | 907 | 91.18 | 91.16 |
| Discharge | 15 | 907 | 80.93 | 80.89 |

Probabilities of sufficient recorded time conditional on a composite event. Fitting used 2020–2021 and selection/recalibration used 2022.

| eTable 13. Conditional Window-Model Discrimination and Predictions in 2023 | | | | | | | | | |
| --- | --- | --- | --- | --- | --- | --- | --- | --- | --- |
| Population | Completion, min | Encounters | AUROC | Mean | 5th | 25th | 50th | 75th | 95th |
| Discharge | | | | | | | | | |
| With event | 5 | 1,068 | 0.790 | 0.955 | 0.860 | 0.942 | 0.970 | 0.985 | 0.995 |
| With event | 10 | 1,068 | 0.750 | 0.910 | 0.761 | 0.882 | 0.931 | 0.961 | 0.984 |
| With event | 15 | 1,068 | 0.773 | 0.812 | 0.620 | 0.744 | 0.812 | 0.906 | 0.962 |
| All eligible | 5 | 48,313 | 0.665 | 0.948 | 0.850 | 0.931 | 0.963 | 0.982 | 0.994 |
| All eligible | 10 | 48,313 | 0.644 | 0.892 | 0.731 | 0.854 | 0.911 | 0.950 | 0.981 |
| All eligible | 15 | 48,313 | 0.690 | 0.762 | 0.500 | 0.691 | 0.772 | 0.856 | 0.949 |
| Admission | | | | | | | | | |
| With event | 5 | 1,112 | 0.505 | 0.989 | 0.976 | 0.987 | 0.991 | 0.993 | 0.995 |
| With event | 10 | 1,112 | 0.596 | 0.969 | 0.938 | 0.963 | 0.974 | 0.981 | 0.988 |
| With event | 15 | 1,112 | 0.626 | 0.951 | 0.888 | 0.941 | 0.961 | 0.972 | 0.983 |
| All eligible | 5 | 24,401 | 0.618 | 0.988 | 0.972 | 0.986 | 0.990 | 0.993 | 0.995 |
| All eligible | 10 | 24,401 | 0.623 | 0.965 | 0.925 | 0.958 | 0.970 | 0.979 | 0.986 |
| All eligible | 15 | 24,401 | 0.627 | 0.948 | 0.878 | 0.938 | 0.959 | 0.971 | 0.982 |

The model predicts sufficient recorded time conditional on a composite event. Discrimination among event encounters is the target evaluation; all-encounter values are secondary. Quantiles describe predicted probabilities, not window lengths. Each model was fitted using training events and selected and recalibrated using tuning events. AUROC indicates area under the receiver operating characteristic curve.

| eTable 14. Risk-Only and Risk-Plus-Window Selection Overlap in 2023 | | | | | | |
| --- | --- | --- | --- | --- | --- | --- |
| Group | Risk-only | Risk-plus-window | Both rules | Risk-only alone | Risk-plus-window alone | Spearman correlation |
| Admission | 890 | 890 | 866 | 24 | 24 | 0.999 |
| Discharge | 1,857 | 1,856 | 1,750 | 107 | 106 | 0.997 |

Ten-minute sequential reviews, 5% tuning flags, and zero delay. Counts are selected encounter fractions, including fractional allocations at simultaneous decision times. Correlation compares predicted event risk with risk multiplied by predicted conditional window sufficiency. Event-overlap counts are not shown to protect small cells.

| eTable 15. Sequential Thresholds and Planned Capacities | | | | | | | | |
| --- | --- | --- | --- | --- | --- | --- | --- | --- |
| Group | Review, min | Flag rate, % | Rule | Delay, min | Daily capacity | Annual capacity | Score threshold | Tiebreak threshold |
| Admission | 10 | 5 | Random | All | 3 | 1,095 | 0 | 0.9525 |
| Admission | 10 | 5 | ESI-first | All | 3 | 1,095 | −2 | 0.9203 |
| Admission | 10 | 5 | Age-first | All | 3 | 1,095 | 91 | 0.1365 |
| Admission | 10 | 5 | Prior-visits-first | All | 3 | 1,095 | 3 | 0.6642 |
| Admission | 10 | 5 | Simple logistic | All | 3 | 1,095 | 0.0888 | 0.8469 |
| Admission | 10 | 5 | Risk-only | All | 3 | 1,095 | 0.0993 | 0.3088 |
| Admission | 10 | 5 | Risk-plus-window | 0 | 3 | 1,095 | 0.0971 | 0.2921 |
| Admission | 10 | 5 | Risk-plus-window | 5 | 3 | 1,095 | 0.0957 | 0.2041 |
| Admission | 10 | 5 | Risk-plus-window | 10 | 3 | 1,095 | Not estimated | Not estimated |
| Admission | 10 | 10 | Random | All | 3 | 1,095 | 0 | 0.9010 |
| Admission | 10 | 10 | ESI-first | All | 3 | 1,095 | −2 | 0.7955 |
| Admission | 10 | 10 | Age-first | All | 3 | 1,095 | 86 | 0.9236 |
| Admission | 10 | 10 | Prior-visits-first | All | 3 | 1,095 | 2 | 0.7008 |
| Admission | 10 | 10 | Simple logistic | All | 3 | 1,095 | 0.0729 | 0.3034 |
| Admission | 10 | 10 | Risk-only | All | 3 | 1,095 | 0.0842 | 0.0769 |
| Admission | 10 | 10 | Risk-plus-window | 0 | 3 | 1,095 | 0.0822 | 0.9051 |
| Admission | 10 | 10 | Risk-plus-window | 5 | 3 | 1,095 | 0.0810 | 0.5025 |
| Admission | 10 | 10 | Risk-plus-window | 10 | 3 | 1,095 | Not estimated | Not estimated |
| Admission | 5 | 10 | Random | All | 6 | 2,190 | 0 | 0.9010 |
| Admission | 5 | 10 | ESI-first | All | 6 | 2,190 | −2 | 0.7955 |
| Admission | 5 | 10 | Age-first | All | 6 | 2,190 | 86 | 0.9236 |
| Admission | 5 | 10 | Prior-visits-first | All | 6 | 2,190 | 2 | 0.7008 |
| Admission | 5 | 10 | Simple logistic | All | 6 | 2,190 | 0.0729 | 0.3034 |
| Admission | 5 | 10 | Risk-only | All | 6 | 2,190 | 0.0842 | 0.0769 |
| Admission | 5 | 10 | Risk-plus-window | 0 | 6 | 2,190 | 0.0834 | 0.2919 |
| Admission | 5 | 10 | Risk-plus-window | 5 | 6 | 2,190 | 0.0822 | 0.9051 |
| Admission | 5 | 10 | Risk-plus-window | 10 | 6 | 2,190 | 0.0810 | 0.5025 |
| Discharge | 10 | 5 | Random | All | 6 | 2,190 | 0 | 0.9505 |
| Discharge | 10 | 5 | ESI-first | All | 6 | 2,190 | −2 | 0.6406 |
| Discharge | 10 | 5 | Age-first | All | 6 | 2,190 | 82 | 0.5622 |
| Discharge | 10 | 5 | Prior-visits-first | All | 6 | 2,190 | 3 | 0.2748 |
| Discharge | 10 | 5 | Simple logistic | All | 6 | 2,190 | 0.0519 | 0.8069 |
| Discharge | 10 | 5 | Risk-only | All | 6 | 2,190 | 0.0755 | 0.4760 |
| Discharge | 10 | 5 | Risk-plus-window | 0 | 6 | 2,190 | 0.0698 | 0.5797 |
| Discharge | 10 | 5 | Risk-plus-window | 5 | 6 | 2,190 | 0.0631 | 0.8740 |
| Discharge | 10 | 5 | Risk-plus-window | 10 | 6 | 2,190 | Not estimated | Not estimated |
| Discharge | 10 | 10 | Random | All | 6 | 2,190 | 0 | 0.9013 |
| Discharge | 10 | 10 | ESI-first | All | 6 | 2,190 | −2 | 0.2743 |
| Discharge | 10 | 10 | Age-first | All | 6 | 2,190 | 75 | 0.6909 |
| Discharge | 10 | 10 | Prior-visits-first | All | 6 | 2,190 | 2 | 0.1120 |
| Discharge | 10 | 10 | Simple logistic | All | 6 | 2,190 | 0.0406 | 0.0193 |
| Discharge | 10 | 10 | Risk-only | All | 6 | 2,190 | 0.0486 | 0.5938 |
| Discharge | 10 | 10 | Risk-plus-window | 0 | 6 | 2,190 | 0.0447 | 0.0870 |
| Discharge | 10 | 10 | Risk-plus-window | 5 | 6 | 2,190 | 0.0399 | 0.5890 |
| Discharge | 10 | 10 | Risk-plus-window | 10 | 6 | 2,190 | Not estimated | Not estimated |
| Discharge | 5 | 10 | Random | All | 12 | 4,380 | 0 | 0.9013 |
| Discharge | 5 | 10 | ESI-first | All | 12 | 4,380 | −2 | 0.2743 |
| Discharge | 5 | 10 | Age-first | All | 12 | 4,380 | 75 | 0.6909 |
| Discharge | 5 | 10 | Prior-visits-first | All | 12 | 4,380 | 2 | 0.1120 |
| Discharge | 5 | 10 | Simple logistic | All | 12 | 4,380 | 0.0406 | 0.0193 |
| Discharge | 5 | 10 | Risk-only | All | 12 | 4,380 | 0.0486 | 0.5938 |
| Discharge | 5 | 10 | Risk-plus-window | 0 | 12 | 4,380 | 0.0467 | 0.1813 |
| Discharge | 5 | 10 | Risk-plus-window | 5 | 12 | 4,380 | 0.0447 | 0.0870 |
| Discharge | 5 | 10 | Risk-plus-window | 10 | 12 | 4,380 | 0.0399 | 0.5890 |

Thresholds were fixed using 2022. Printed thresholds are rounded for description; selection used full precision. For ESI-first, the score is the negative ESI level, with missing or invalid levels ranked last. A threshold of −2 corresponds to ESI 2; tied scores use the stated tiebreak threshold. At an exactly equal score, the fixed seeded tiebreak must meet the tiebreak threshold. Random uses its tiebreak alone. Risk-plus-window thresholds depend on delay plus duration; the 20-minute completion scenario was not estimated. Simple logistic indicates the five-variable logistic model. ESI indicates Emergency Severity Index.

| eTable 16. Sequential Selection Across Review Durations, Thresholds, and Delays in 2023 | | | | | | | | | |
| --- | --- | --- | --- | --- | --- | --- | --- | --- | --- |
| Group | Review, min | Flag rate, % | Delay, min | Rule | Planned attempts | Attempts used | Events reached | Reach, % | Attempts per event |
| Admission | 10 | 5 | 0 | Random | 1,095 | 854 | 34 | 3.06 | 25.1 |
| Admission | 10 | 5 | 0 | ESI-first | 1,095 | 890 | 81 | 7.28 | 11.0 |
| Admission | 10 | 5 | 0 | Age-first | 1,095 | 868 | 35 | 3.15 | 24.8 |
| Admission | 10 | 5 | 0 | Prior-visits-first | 1,095 | 920 | 86 | 7.73 | 10.7 |
| Admission | 10 | 5 | 0 | Simple logistic | 1,095 | 944 | 125 | 11.24 | 7.6 |
| Admission | 10 | 5 | 0 | Risk-only | 1,095 | 890 | 112 | 10.07 | 7.9 |
| Admission | 10 | 5 | 0 | Risk-plus-window | 1,095 | 890 | 115 | 10.34 | 7.7 |
| Admission | 10 | 5 | 5 | Random | 1,095 | 854 | 34 | 3.06 | 25.1 |
| Admission | 10 | 5 | 5 | ESI-first | 1,095 | 890 | 81 | 7.28 | 11.0 |
| Admission | 10 | 5 | 5 | Age-first | 1,095 | 868 | 35 | 3.15 | 24.8 |
| Admission | 10 | 5 | 5 | Prior-visits-first | 1,095 | 920 | 82 | 7.37 | 11.2 |
| Admission | 10 | 5 | 5 | Simple logistic | 1,095 | 944 | 123 | 11.06 | 7.7 |
| Admission | 10 | 5 | 5 | Risk-only | 1,095 | 890 | 105 | 9.44 | 8.5 |
| Admission | 10 | 5 | 5 | Risk-plus-window | 1,095 | 880 | 109 | 9.80 | 8.1 |
| Admission | 10 | 5 | 10 | Random | 1,095 | 854 | 34 | 3.06 | 25.1 |
| Admission | 10 | 5 | 10 | ESI-first | 1,095 | 890 | 80 | 7.19 | 11.1 |
| Admission | 10 | 5 | 10 | Age-first | 1,095 | 868 | 33 | 2.97 | 26.3 |
| Admission | 10 | 5 | 10 | Prior-visits-first | 1,095 | 920 | 82 | 7.37 | 11.2 |
| Admission | 10 | 5 | 10 | Simple logistic | 1,095 | 944 | 123 | 11.06 | 7.7 |
| Admission | 10 | 5 | 10 | Risk-only | 1,095 | 890 | 104 | 9.35 | 8.6 |
| Admission | 10 | 5 | 10 | Risk-plus-window | 1,095 | Not estimated | Not estimated | Not estimated | Not estimated |
| Admission | 10 | 10 | 0 | Random | 1,095 | 1,071 | 50 | 4.50 | 21.4 |
| Admission | 10 | 10 | 0 | ESI-first | 1,095 | 1,074 | 80 | 7.19 | 13.4 |
| Admission | 10 | 10 | 0 | Age-first | 1,095 | 1,075 | 64 | 5.76 | 16.8 |
| Admission | 10 | 10 | 0 | Prior-visits-first | 1,095 | 1,084 | 98 | 8.81 | 11.1 |
| Admission | 10 | 10 | 0 | Simple logistic | 1,095 | 1,076 | 120.5 | 10.84 | 8.9 |
| Admission | 10 | 10 | 0 | Risk-only | 1,095 | 1,079 | 137.5 | 12.37 | 7.8 |
| Admission | 10 | 10 | 0 | Risk-plus-window | 1,095 | 1,079 | 135.5 | 12.19 | 8.0 |
| Admission | 10 | 10 | 5 | Random | 1,095 | 1,071 | 50 | 4.50 | 21.4 |
| Admission | 10 | 10 | 5 | ESI-first | 1,095 | 1,074 | 79 | 7.10 | 13.6 |
| Admission | 10 | 10 | 5 | Age-first | 1,095 | 1,075 | 64 | 5.76 | 16.8 |
| Admission | 10 | 10 | 5 | Prior-visits-first | 1,095 | 1,084 | 94 | 8.45 | 11.5 |
| Admission | 10 | 10 | 5 | Simple logistic | 1,095 | 1,076 | 117.5 | 10.57 | 9.2 |
| Admission | 10 | 10 | 5 | Risk-only | 1,095 | 1,079 | 131.5 | 11.83 | 8.2 |
| Admission | 10 | 10 | 5 | Risk-plus-window | 1,095 | 1,075 | 124.5 | 11.20 | 8.6 |
| Admission | 10 | 10 | 10 | Random | 1,095 | 1,071 | 49 | 4.41 | 21.9 |
| Admission | 10 | 10 | 10 | ESI-first | 1,095 | 1,074 | 79 | 7.10 | 13.6 |
| Admission | 10 | 10 | 10 | Age-first | 1,095 | 1,075 | 62 | 5.58 | 17.3 |
| Admission | 10 | 10 | 10 | Prior-visits-first | 1,095 | 1,084 | 93 | 8.36 | 11.7 |
| Admission | 10 | 10 | 10 | Simple logistic | 1,095 | 1,076 | 116.5 | 10.48 | 9.2 |
| Admission | 10 | 10 | 10 | Risk-only | 1,095 | 1,079 | 130.5 | 11.74 | 8.3 |
| Admission | 10 | 10 | 10 | Risk-plus-window | 1,095 | Not estimated | Not estimated | Not estimated | Not estimated |
| Admission | 5 | 10 | 0 | Random | 2,190 | 1,875 | 82.5 | 7.42 | 22.7 |
| Admission | 5 | 10 | 0 | ESI-first | 2,190 | 1,972 | 143.5 | 12.90 | 13.7 |
| Admission | 5 | 10 | 0 | Age-first | 2,190 | 1,938 | 104.5 | 9.40 | 18.5 |
| Admission | 5 | 10 | 0 | Prior-visits-first | 2,190 | 1,940 | 158 | 14.21 | 12.3 |
| Admission | 5 | 10 | 0 | Simple logistic | 2,190 | 1,910 | 203.5 | 18.30 | 9.4 |
| Admission | 5 | 10 | 0 | Risk-only | 2,190 | 1,895 | 218 | 19.60 | 8.7 |
| Admission | 5 | 10 | 0 | Risk-plus-window | 2,190 | 1,892 | 217 | 19.51 | 8.7 |
| Admission | 5 | 10 | 5 | Random | 2,190 | 1,875 | 82.5 | 7.42 | 22.7 |
| Admission | 5 | 10 | 5 | ESI-first | 2,190 | 1,972 | 141.5 | 12.72 | 13.9 |
| Admission | 5 | 10 | 5 | Age-first | 2,190 | 1,938 | 101.5 | 9.13 | 19.1 |
| Admission | 5 | 10 | 5 | Prior-visits-first | 2,190 | 1,940 | 155 | 13.94 | 12.5 |
| Admission | 5 | 10 | 5 | Simple logistic | 2,190 | 1,910 | 199.5 | 17.94 | 9.6 |
| Admission | 5 | 10 | 5 | Risk-only | 2,190 | 1,895 | 213 | 19.15 | 8.9 |
| Admission | 5 | 10 | 5 | Risk-plus-window | 2,190 | 1,894 | 213 | 19.15 | 8.9 |
| Admission | 5 | 10 | 10 | Random | 2,190 | 1,875 | 81.5 | 7.33 | 23.0 |
| Admission | 5 | 10 | 10 | ESI-first | 2,190 | 1,972 | 140.5 | 12.63 | 14.0 |
| Admission | 5 | 10 | 10 | Age-first | 2,190 | 1,938 | 101.5 | 9.13 | 19.1 |
| Admission | 5 | 10 | 10 | Prior-visits-first | 2,190 | 1,940 | 150 | 13.49 | 12.9 |
| Admission | 5 | 10 | 10 | Simple logistic | 2,190 | 1,910 | 195.5 | 17.58 | 9.8 |
| Admission | 5 | 10 | 10 | Risk-only | 2,190 | 1,895 | 206 | 18.53 | 9.2 |
| Admission | 5 | 10 | 10 | Risk-plus-window | 2,190 | 1,889 | 205 | 18.44 | 9.2 |
| Discharge | 10 | 5 | 0 | Random | 2,190 | 1,865 | 29 | 2.72 | 64.3 |
| Discharge | 10 | 5 | 0 | ESI-first | 2,190 | 2,011 | 54 | 5.06 | 37.2 |
| Discharge | 10 | 5 | 0 | Age-first | 2,190 | 1,953 | 78 | 7.30 | 25.0 |
| Discharge | 10 | 5 | 0 | Prior-visits-first | 2,190 | 1,994 | 171 | 16.01 | 11.7 |
| Discharge | 10 | 5 | 0 | Simple logistic | 2,190 | 2,006 | 160 | 14.98 | 12.5 |
| Discharge | 10 | 5 | 0 | Risk-only | 2,190 | 1,857 | 208 | 19.48 | 8.9 |
| Discharge | 10 | 5 | 0 | Risk-plus-window | 2,190 | 1,856 | 208 | 19.48 | 8.9 |
| Discharge | 10 | 5 | 5 | Random | 2,190 | 1,865 | 26 | 2.43 | 71.7 |
| Discharge | 10 | 5 | 5 | ESI-first | 2,190 | 2,011 | 52 | 4.87 | 38.7 |
| Discharge | 10 | 5 | 5 | Age-first | 2,190 | 1,953 | 74 | 6.93 | 26.4 |
| Discharge | 10 | 5 | 5 | Prior-visits-first | 2,190 | 1,994 | 149 | 13.95 | 13.4 |
| Discharge | 10 | 5 | 5 | Simple logistic | 2,190 | 2,006 | 148 | 13.86 | 13.6 |
| Discharge | 10 | 5 | 5 | Risk-only | 2,190 | 1,857 | 192 | 17.98 | 9.7 |
| Discharge | 10 | 5 | 5 | Risk-plus-window | 2,190 | 1,845 | 190 | 17.79 | 9.7 |
| Discharge | 10 | 5 | 10 | Random | 2,190 | 1,865 | 22 | 2.06 | 84.8 |
| Discharge | 10 | 5 | 10 | ESI-first | 2,190 | 2,011 | 46 | 4.31 | 43.7 |
| Discharge | 10 | 5 | 10 | Age-first | 2,190 | 1,953 | 67 | 6.27 | 29.1 |
| Discharge | 10 | 5 | 10 | Prior-visits-first | 2,190 | 1,994 | 137 | 12.83 | 14.6 |
| Discharge | 10 | 5 | 10 | Simple logistic | 2,190 | 2,006 | 136 | 12.73 | 14.8 |
| Discharge | 10 | 5 | 10 | Risk-only | 2,190 | 1,857 | 171 | 16.01 | 10.9 |
| Discharge | 10 | 5 | 10 | Risk-plus-window | 2,190 | Not estimated | Not estimated | Not estimated | Not estimated |
| Discharge | 10 | 10 | 0 | Random | 2,190 | 2,178 | 44 | 4.12 | 49.5 |
| Discharge | 10 | 10 | 0 | ESI-first | 2,190 | 2,187 | 72 | 6.74 | 30.4 |
| Discharge | 10 | 10 | 0 | Age-first | 2,190 | 2,179 | 69 | 6.46 | 31.6 |
| Discharge | 10 | 10 | 0 | Prior-visits-first | 2,190 | 2,183 | 145 | 13.58 | 15.1 |
| Discharge | 10 | 10 | 0 | Simple logistic | 2,190 | 2,172 | 132.5 | 12.41 | 16.4 |
| Discharge | 10 | 10 | 0 | Risk-only | 2,190 | 2,169 | 197.5 | 18.49 | 11.0 |
| Discharge | 10 | 10 | 0 | Risk-plus-window | 2,190 | 2,163 | 196.5 | 18.40 | 11.0 |
| Discharge | 10 | 10 | 5 | Random | 2,190 | 2,178 | 42 | 3.93 | 51.9 |
| Discharge | 10 | 10 | 5 | ESI-first | 2,190 | 2,187 | 65 | 6.09 | 33.6 |
| Discharge | 10 | 10 | 5 | Age-first | 2,190 | 2,179 | 65 | 6.09 | 33.5 |
| Discharge | 10 | 10 | 5 | Prior-visits-first | 2,190 | 2,183 | 132 | 12.36 | 16.5 |
| Discharge | 10 | 10 | 5 | Simple logistic | 2,190 | 2,172 | 118 | 11.05 | 18.4 |
| Discharge | 10 | 10 | 5 | Risk-only | 2,190 | 2,169 | 182.5 | 17.09 | 11.9 |
| Discharge | 10 | 10 | 5 | Risk-plus-window | 2,190 | 2,170 | 185 | 17.32 | 11.7 |
| Discharge | 10 | 10 | 10 | Random | 2,190 | 2,178 | 34 | 3.18 | 64.1 |
| Discharge | 10 | 10 | 10 | ESI-first | 2,190 | 2,187 | 54 | 5.06 | 40.5 |
| Discharge | 10 | 10 | 10 | Age-first | 2,190 | 2,179 | 57 | 5.34 | 38.2 |
| Discharge | 10 | 10 | 10 | Prior-visits-first | 2,190 | 2,183 | 115 | 10.77 | 19.0 |
| Discharge | 10 | 10 | 10 | Simple logistic | 2,190 | 2,172 | 109 | 10.21 | 19.9 |
| Discharge | 10 | 10 | 10 | Risk-only | 2,190 | 2,169 | 158.5 | 14.84 | 13.7 |
| Discharge | 10 | 10 | 10 | Risk-plus-window | 2,190 | Not estimated | Not estimated | Not estimated | Not estimated |
| Discharge | 5 | 10 | 0 | Random | 4,380 | 3,982 | 84 | 7.87 | 47.4 |
| Discharge | 5 | 10 | 0 | ESI-first | 4,380 | 4,201 | 135 | 12.64 | 31.1 |
| Discharge | 5 | 10 | 0 | Age-first | 4,380 | 4,111 | 135 | 12.64 | 30.5 |
| Discharge | 5 | 10 | 0 | Prior-visits-first | 4,380 | 4,051 | 304 | 28.46 | 13.3 |
| Discharge | 5 | 10 | 0 | Simple logistic | 4,380 | 4,113 | 278 | 26.03 | 14.8 |
| Discharge | 5 | 10 | 0 | Risk-only | 4,380 | 3,924 | 354 | 33.15 | 11.1 |
| Discharge | 5 | 10 | 0 | Risk-plus-window | 4,380 | 3,916 | 357 | 33.43 | 11.0 |
| Discharge | 5 | 10 | 5 | Random | 4,380 | 3,982 | 78 | 7.30 | 51.1 |
| Discharge | 5 | 10 | 5 | ESI-first | 4,380 | 4,201 | 124 | 11.61 | 33.9 |
| Discharge | 5 | 10 | 5 | Age-first | 4,380 | 4,111 | 128 | 11.99 | 32.1 |
| Discharge | 5 | 10 | 5 | Prior-visits-first | 4,380 | 4,051 | 289 | 27.06 | 14.0 |
| Discharge | 5 | 10 | 5 | Simple logistic | 4,380 | 4,113 | 263 | 24.63 | 15.6 |
| Discharge | 5 | 10 | 5 | Risk-only | 4,380 | 3,924 | 331 | 30.99 | 11.9 |
| Discharge | 5 | 10 | 5 | Risk-plus-window | 4,380 | 3,917 | 330 | 30.90 | 11.9 |
| Discharge | 5 | 10 | 10 | Random | 4,380 | 3,982 | 71 | 6.65 | 56.1 |
| Discharge | 5 | 10 | 10 | ESI-first | 4,380 | 4,201 | 112 | 10.49 | 37.5 |
| Discharge | 5 | 10 | 10 | Age-first | 4,380 | 4,111 | 121 | 11.33 | 34.0 |
| Discharge | 5 | 10 | 10 | Prior-visits-first | 4,380 | 4,051 | 257 | 24.06 | 15.8 |
| Discharge | 5 | 10 | 10 | Simple logistic | 4,380 | 4,113 | 238 | 22.28 | 17.3 |
| Discharge | 5 | 10 | 10 | Risk-only | 4,380 | 3,924 | 304 | 28.46 | 12.9 |
| Discharge | 5 | 10 | 10 | Risk-plus-window | 4,380 | 3,929 | 300 | 28.09 | 13.1 |

Descriptive point estimates. Fractional event counts reflect shared capacity at simultaneous decision cutoffs. Thresholds and planned capacities used 2022. All encounters remained in each scenario; insufficient windows prevented completion, not selection. Event reach is the proportion of all composite-event encounters selected with sufficient recorded review time. Review attempts without enough recorded time still count against the budget. Reach does not measure events prevented. Simple logistic indicates the five-variable logistic model. ESI indicates Emergency Severity Index.

| eTable 17. Sequential Selection Across Scenarios in 2024 | | | | | | | | |
| --- | --- | --- | --- | --- | --- | --- | --- | --- |
| Review, min | Tuning flags, % | Delay, min | Rule | Capacity | Attempts | Unused | Events reached | Reach, % |
| Discharge | | | | | | | | |
| 10 | 5 | 0 | Random | 2,196 | 1,973 | 223 | 34 | 3.2 |
| 10 | 5 | 0 | ESI-first | 2,196 | 2,081 | 115 | 62.5 | 5.9 |
| 10 | 5 | 0 | Age-first | 2,196 | 2,069 | 127 | 67 | 6.3 |
| 10 | 5 | 0 | Prior-visits-first | 2,196 | 2,081 | 115 | 168 | 15.8 |
| 10 | 5 | 0 | Simple logistic | 2,196 | 2,132 | 64 | 127 | 11.9 |
| 10 | 5 | 0 | Risk-only | 2,196 | 2,018 | 178 | 207 | 19.4 |
| 10 | 5 | 0 | Risk-plus-window | 2,196 | 2,028 | 168 | 208 | 19.5 |
| 10 | 5 | 5 | Random | 2,196 | 1,973 | 223 | 28 | 2.6 |
| 10 | 5 | 5 | ESI-first | 2,196 | 2,081 | 115 | 54.5 | 5.1 |
| 10 | 5 | 5 | Age-first | 2,196 | 2,069 | 127 | 60 | 5.6 |
| 10 | 5 | 5 | Prior-visits-first | 2,196 | 2,081 | 115 | 153 | 14.4 |
| 10 | 5 | 5 | Simple logistic | 2,196 | 2,132 | 64 | 116 | 10.9 |
| 10 | 5 | 5 | Risk-only | 2,196 | 2,018 | 178 | 194 | 18.2 |
| 10 | 5 | 5 | Risk-plus-window | 2,196 | 2,026 | 170 | 199 | 18.7 |
| 10 | 5 | 10 | Random | 2,196 | 1,973 | 223 | 22 | 2.1 |
| 10 | 5 | 10 | ESI-first | 2,196 | 2,081 | 115 | 48.5 | 4.5 |
| 10 | 5 | 10 | Age-first | 2,196 | 2,069 | 127 | 54 | 5.1 |
| 10 | 5 | 10 | Prior-visits-first | 2,196 | 2,081 | 115 | 136 | 12.8 |
| 10 | 5 | 10 | Simple logistic | 2,196 | 2,132 | 64 | 104 | 9.8 |
| 10 | 5 | 10 | Risk-only | 2,196 | 2,018 | 178 | 179 | 16.8 |
| 10 | 5 | 10 | Risk-plus-window | 2,196 | NE | NE | NE | NE |
| 10 | 10 | 0 | Random | 2,196 | — | — | 39 | 3.7 |
| 10 | 10 | 0 | ESI-first | 2,196 | — | — | 59 | 5.5 |
| 10 | 10 | 0 | Age-first | 2,196 | — | — | 73 | 6.8 |
| 10 | 10 | 0 | Prior-visits-first | 2,196 | 2,196 | 0 | 141.5 | 13.3 |
| 10 | 10 | 0 | Simple logistic | 2,196 | — | — | 113.5 | 10.6 |
| 10 | 10 | 0 | Risk-only | 2,196 | — | — | 167 | 15.7 |
| 10 | 10 | 0 | Risk-plus-window | 2,196 | — | — | 164 | 15.4 |
| 10 | 10 | 5 | Random | 2,196 | — | — | 35 | 3.3 |
| 10 | 10 | 5 | ESI-first | 2,196 | — | — | 54 | 5.1 |
| 10 | 10 | 5 | Age-first | 2,196 | — | — | 67 | 6.3 |
| 10 | 10 | 5 | Prior-visits-first | 2,196 | 2,196 | 0 | 131.5 | 12.3 |
| 10 | 10 | 5 | Simple logistic | 2,196 | — | — | 101.5 | 9.5 |
| 10 | 10 | 5 | Risk-only | 2,196 | — | — | 156 | 14.6 |
| 10 | 10 | 5 | Risk-plus-window | 2,196 | — | — | 156 | 14.6 |
| 10 | 10 | 10 | Random | 2,196 | — | — | 31 | 2.9 |
| 10 | 10 | 10 | ESI-first | 2,196 | — | — | 50 | 4.7 |
| 10 | 10 | 10 | Age-first | 2,196 | — | — | 59 | 5.5 |
| 10 | 10 | 10 | Prior-visits-first | 2,196 | 2,196 | 0 | 116.5 | 10.9 |
| 10 | 10 | 10 | Simple logistic | 2,196 | — | — | 91.5 | 8.6 |
| 10 | 10 | 10 | Risk-only | 2,196 | — | — | 143 | 13.4 |
| 10 | 10 | 10 | Risk-plus-window | 2,196 | NE | NE | NE | NE |
| 5 | 10 | 0 | Random | 4,392 | 4,098 | 294 | 76 | 7.1 |
| 5 | 10 | 0 | ESI-first | 4,392 | 4,294 | 98 | 119 | 11.2 |
| 5 | 10 | 0 | Age-first | 4,392 | 4,235 | 157 | 132 | 12.4 |
| 5 | 10 | 0 | Prior-visits-first | 4,392 | 4,190 | 202 | 287 | 26.9 |
| 5 | 10 | 0 | Simple logistic | 4,392 | 4,308 | 84 | 222 | 20.8 |
| 5 | 10 | 0 | Risk-only | 4,392 | 4,201 | 191 | 348 | 32.6 |
| 5 | 10 | 0 | Risk-plus-window | 4,392 | 4,209 | 183 | 342 | 32.1 |
| 5 | 10 | 5 | Random | 4,392 | 4,098 | 294 | 67 | 6.3 |
| 5 | 10 | 5 | ESI-first | 4,392 | 4,294 | 98 | 112 | 10.5 |
| 5 | 10 | 5 | Age-first | 4,392 | 4,235 | 157 | 123 | 11.5 |
| 5 | 10 | 5 | Prior-visits-first | 4,392 | 4,190 | 202 | 267 | 25.0 |
| 5 | 10 | 5 | Simple logistic | 4,392 | 4,308 | 84 | 213 | 20.0 |
| 5 | 10 | 5 | Risk-only | 4,392 | 4,201 | 191 | 332 | 31.1 |
| 5 | 10 | 5 | Risk-plus-window | 4,392 | 4,207 | 185 | 327 | 30.7 |
| 5 | 10 | 10 | Random | 4,392 | 4,098 | 294 | 60 | 5.6 |
| 5 | 10 | 10 | ESI-first | 4,392 | 4,294 | 98 | 101 | 9.5 |
| 5 | 10 | 10 | Age-first | 4,392 | 4,235 | 157 | 114 | 10.7 |
| 5 | 10 | 10 | Prior-visits-first | 4,392 | 4,190 | 202 | 243 | 22.8 |
| 5 | 10 | 10 | Simple logistic | 4,392 | 4,308 | 84 | 194 | 18.2 |
| 5 | 10 | 10 | Risk-only | 4,392 | 4,201 | 191 | 306 | 28.7 |
| 5 | 10 | 10 | Risk-plus-window | 4,392 | 4,214 | 178 | 296 | 27.8 |
| Admission | | | | | | | | |
| 10 | 5 | 0 | Random | 1,098 | 898 | 200 | 40.5 | 3.6 |
| 10 | 5 | 0 | ESI-first | 1,098 | 925 | 173 | 70 | 6.2 |
| 10 | 5 | 0 | Age-first | 1,098 | 890 | 208 | 54 | 4.8 |
| 10 | 5 | 0 | Prior-visits-first | 1,098 | 943 | 155 | 86 | 7.6 |
| 10 | 5 | 0 | Simple logistic | 1,098 | 927 | 171 | 119 | 10.6 |
| 10 | 5 | 0 | Risk-only | 1,098 | 901 | 197 | 126 | 11.2 |
| 10 | 5 | 0 | Risk-plus-window | 1,098 | 910 | 188 | 123 | 10.9 |
| 10 | 5 | 5 | Random | 1,098 | 898 | 200 | 38.5 | 3.4 |
| 10 | 5 | 5 | ESI-first | 1,098 | 925 | 173 | 69 | 6.1 |
| 10 | 5 | 5 | Age-first | 1,098 | 890 | 208 | 53 | 4.7 |
| 10 | 5 | 5 | Prior-visits-first | 1,098 | 943 | 155 | 82 | 7.3 |
| 10 | 5 | 5 | Simple logistic | 1,098 | 927 | 171 | 118 | 10.5 |
| 10 | 5 | 5 | Risk-only | 1,098 | 901 | 197 | 126 | 11.2 |
| 10 | 5 | 5 | Risk-plus-window | 1,098 | 906 | 192 | 123 | 10.9 |
| 10 | 5 | 10 | Random | 1,098 | 898 | 200 | 38.5 | 3.4 |
| 10 | 5 | 10 | ESI-first | 1,098 | 925 | 173 | 69 | 6.1 |
| 10 | 5 | 10 | Age-first | 1,098 | 890 | 208 | 50 | 4.4 |
| 10 | 5 | 10 | Prior-visits-first | 1,098 | 943 | 155 | 79 | 7.0 |
| 10 | 5 | 10 | Simple logistic | 1,098 | 927 | 171 | 117 | 10.4 |
| 10 | 5 | 10 | Risk-only | 1,098 | 901 | 197 | 126 | 11.2 |
| 10 | 5 | 10 | Risk-plus-window | 1,098 | NE | NE | NE | NE |
| 10 | 10 | 0 | Random | 1,098 | 1,085 | 13 | 51 | 4.5 |
| 10 | 10 | 0 | ESI-first | 1,098 | — | — | 74 | 6.6 |
| 10 | 10 | 0 | Age-first | 1,098 | 1,080 | 18 | 62 | 5.5 |
| 10 | 10 | 0 | Prior-visits-first | 1,098 | 1,085 | 13 | 84 | 7.5 |
| 10 | 10 | 0 | Simple logistic | 1,098 | — | — | 113 | 10.0 |
| 10 | 10 | 0 | Risk-only | 1,098 | 1,084 | 14 | 130.5 | 11.6 |
| 10 | 10 | 0 | Risk-plus-window | 1,098 | 1,086 | 12 | 130.5 | 11.6 |
| 10 | 10 | 5 | Random | 1,098 | 1,085 | 13 | 50 | 4.4 |
| 10 | 10 | 5 | ESI-first | 1,098 | — | — | 74 | 6.6 |
| 10 | 10 | 5 | Age-first | 1,098 | 1,080 | 18 | 62 | 5.5 |
| 10 | 10 | 5 | Prior-visits-first | 1,098 | 1,085 | 13 | 82 | 7.3 |
| 10 | 10 | 5 | Simple logistic | 1,098 | — | — | 111 | 9.9 |
| 10 | 10 | 5 | Risk-only | 1,098 | 1,084 | 14 | 129.5 | 11.5 |
| 10 | 10 | 5 | Risk-plus-window | 1,098 | 1,084 | 14 | 129.5 | 11.5 |
| 10 | 10 | 10 | Random | 1,098 | 1,085 | 13 | 50 | 4.4 |
| 10 | 10 | 10 | ESI-first | 1,098 | — | — | 72 | 6.4 |
| 10 | 10 | 10 | Age-first | 1,098 | 1,080 | 18 | 59 | 5.2 |
| 10 | 10 | 10 | Prior-visits-first | 1,098 | 1,085 | 13 | 78 | 6.9 |
| 10 | 10 | 10 | Simple logistic | 1,098 | — | — | 109 | 9.7 |
| 10 | 10 | 10 | Risk-only | 1,098 | 1,084 | 14 | 129.5 | 11.5 |
| 10 | 10 | 10 | Risk-plus-window | 1,098 | NE | NE | NE | NE |
| 5 | 10 | 0 | Random | 2,196 | 1,976 | 220 | 100 | 8.9 |
| 5 | 10 | 0 | ESI-first | 2,196 | 2,048 | 148 | 134 | 11.9 |
| 5 | 10 | 0 | Age-first | 2,196 | 1,969 | 227 | 106 | 9.4 |
| 5 | 10 | 0 | Prior-visits-first | 2,196 | 1,980 | 216 | 157 | 14.0 |
| 5 | 10 | 0 | Simple logistic | 2,196 | 1,982 | 214 | 206 | 18.3 |
| 5 | 10 | 0 | Risk-only | 2,196 | 1,957 | 239 | 215 | 19.1 |
| 5 | 10 | 0 | Risk-plus-window | 2,196 | 1,959 | 237 | 216 | 19.2 |
| 5 | 10 | 5 | Random | 2,196 | 1,976 | 220 | 98 | 8.7 |
| 5 | 10 | 5 | ESI-first | 2,196 | 2,048 | 148 | 129 | 11.5 |
| 5 | 10 | 5 | Age-first | 2,196 | 1,969 | 227 | 105 | 9.3 |
| 5 | 10 | 5 | Prior-visits-first | 2,196 | 1,980 | 216 | 154 | 13.7 |
| 5 | 10 | 5 | Simple logistic | 2,196 | 1,982 | 214 | 202 | 18.0 |
| 5 | 10 | 5 | Risk-only | 2,196 | 1,957 | 239 | 213 | 18.9 |
| 5 | 10 | 5 | Risk-plus-window | 2,196 | 1,960 | 236 | 217 | 19.3 |
| 5 | 10 | 10 | Random | 2,196 | 1,976 | 220 | 95 | 8.4 |
| 5 | 10 | 10 | ESI-first | 2,196 | 2,048 | 148 | 127 | 11.3 |
| 5 | 10 | 10 | Age-first | 2,196 | 1,969 | 227 | 103 | 9.2 |
| 5 | 10 | 10 | Prior-visits-first | 2,196 | 1,980 | 216 | 150 | 13.3 |
| 5 | 10 | 10 | Simple logistic | 2,196 | 1,982 | 214 | 198 | 17.6 |
| 5 | 10 | 10 | Risk-only | 2,196 | 1,957 | 239 | 211 | 18.8 |
| 5 | 10 | 10 | Risk-plus-window | 2,196 | 1,959 | 237 | 219 | 19.5 |

All 366 calendar days; capacities and thresholds are fixed from 2022. Single-rule estimates are descriptive. Attempt fractions can occur at simultaneous decision times. NE indicates not estimated: Risk-plus-window has no model for a 20-minute completion threshold. Event reach does not measure events prevented. Counts of 1–10, corresponding percentages, and selected cells from which they could be calculated are not shown (—).

| eTable 18. Observed Flag Fractions in 2024 | | | | | | |
| --- | --- | --- | --- | --- | --- | --- |
| Review, min | Tuning target, % | Delay, min | Rule | Encounters | Flagged | Flagged, % |
| Discharge | | | | | | |
| 10 | 5 | All | Random | 50,675 | 2,476 | 4.89 |
| 10 | 5 | All | ESI-first | 50,675 | 3,205 | 6.32 |
| 10 | 5 | All | Age-first | 50,675 | 2,939 | 5.80 |
| 10 | 5 | All | Prior-visits-first | 50,675 | 2,918 | 5.76 |
| 10 | 5 | All | Simple logistic | 50,675 | 4,053 | 8.00 |
| 10 | 5 | All | Risk-only | 50,675 | 2,815 | 5.56 |
| 10 | 5 | 0 | Risk-plus-window | 50,675 | 2,853 | 5.63 |
| 10 | 5 | 5 | Risk-plus-window | 50,675 | 2,854 | 5.63 |
| 10 | 10 | All | Random | 50,675 | 4,996 | 9.86 |
| 10 | 10 | All | ESI-first | 50,675 | 6,364 | 12.56 |
| 10 | 10 | All | Age-first | 50,675 | 5,791 | 11.43 |
| 10 | 10 | All | Prior-visits-first | 50,675 | 5,228 | 10.32 |
| 10 | 10 | All | Simple logistic | 50,675 | 7,589 | 14.98 |
| 10 | 10 | All | Risk-only | 50,675 | 5,671 | 11.19 |
| 10 | 10 | 0 | Risk-plus-window | 50,675 | 5,693 | 11.23 |
| 10 | 10 | 5 | Risk-plus-window | 50,675 | 5,752 | 11.35 |
| 5 | 10 | All | Random | 50,675 | 4,996 | 9.86 |
| 5 | 10 | All | ESI-first | 50,675 | 6,364 | 12.56 |
| 5 | 10 | All | Age-first | 50,675 | 5,791 | 11.43 |
| 5 | 10 | All | Prior-visits-first | 50,675 | 5,228 | 10.32 |
| 5 | 10 | All | Simple logistic | 50,675 | 7,589 | 14.98 |
| 5 | 10 | All | Risk-only | 50,675 | 5,671 | 11.19 |
| 5 | 10 | 0 | Risk-plus-window | 50,675 | 5,701 | 11.25 |
| 5 | 10 | 5 | Risk-plus-window | 50,675 | 5,693 | 11.23 |
| 5 | 10 | 10 | Risk-plus-window | 50,675 | 5,752 | 11.35 |
| Admission | | | | | | |
| 10 | 5 | All | Random | 26,411 | 1,284 | 4.86 |
| 10 | 5 | All | ESI-first | 26,411 | 1,359 | 5.15 |
| 10 | 5 | All | Age-first | 26,411 | 1,234 | 4.67 |
| 10 | 5 | All | Prior-visits-first | 26,411 | 1,340 | 5.07 |
| 10 | 5 | All | Simple logistic | 26,411 | 1,296 | 4.91 |
| 10 | 5 | All | Risk-only | 26,411 | 1,260 | 4.77 |
| 10 | 5 | 0 | Risk-plus-window | 26,411 | 1,265 | 4.79 |
| 10 | 5 | 5 | Risk-plus-window | 26,411 | 1,248 | 4.73 |
| 10 | 10 | All | Random | 26,411 | 2,572 | 9.74 |
| 10 | 10 | All | ESI-first | 26,411 | 2,903 | 10.99 |
| 10 | 10 | All | Age-first | 26,411 | 2,599 | 9.84 |
| 10 | 10 | All | Prior-visits-first | 26,411 | 2,559 | 9.69 |
| 10 | 10 | All | Simple logistic | 26,411 | 2,547 | 9.64 |
| 10 | 10 | All | Risk-only | 26,411 | 2,509 | 9.50 |
| 10 | 10 | 0 | Risk-plus-window | 26,411 | 2,533 | 9.59 |
| 10 | 10 | 5 | Risk-plus-window | 26,411 | 2,513 | 9.51 |
| 5 | 10 | All | Random | 26,411 | 2,572 | 9.74 |
| 5 | 10 | All | ESI-first | 26,411 | 2,903 | 10.99 |
| 5 | 10 | All | Age-first | 26,411 | 2,599 | 9.84 |
| 5 | 10 | All | Prior-visits-first | 26,411 | 2,559 | 9.69 |
| 5 | 10 | All | Simple logistic | 26,411 | 2,547 | 9.64 |
| 5 | 10 | All | Risk-only | 26,411 | 2,509 | 9.50 |
| 5 | 10 | 0 | Risk-plus-window | 26,411 | 2,516 | 9.53 |
| 5 | 10 | 5 | Risk-plus-window | 26,411 | 2,533 | 9.59 |
| 5 | 10 | 10 | Risk-plus-window | 26,411 | 2,513 | 9.51 |

Thresholds and seeded tiebreaks come from 2022 and were not adjusted to the 2024 flag fraction. For rules other than Risk-plus-window, the score and flags do not depend on delay. Counts of 1–10, corresponding percentages, and selected cells from which they could be calculated are not shown (—).

| eTable 19. Full-Day and Annual Ranking at the Main Budget in 2023 | | | | | |
| --- | --- | --- | --- | --- | --- |
| Decision group | Rule | Daily reach % | Annual reach % | Daily lift | Annual lift |
| Admission | Random | 4.80 | 4.81 | 1.00 | 1.00 |
| Admission | ESI-first | 9.81 | 9.89 | 2.04 | 2.06 |
| Admission | Age-first | 4.32 | 4.67 | 0.90 | 0.97 |
| Admission | Prior-visits-first | 10.49 | 10.75 | 2.19 | 2.24 |
| Admission | Simple logistic | 12.68 | 14.30 | 2.64 | 2.97 |
| Admission | Risk-only | 14.39 | 14.39 | 3.00 | 2.99 |
| Admission | Risk-plus-window | 14.66 | 14.12 | 3.05 | 2.94 |
| Discharge | Random | 4.20 | 4.20 | 1.00 | 1.00 |
| Discharge | ESI-first | 6.70 | 6.69 | 1.60 | 1.59 |
| Discharge | Age-first | 8.43 | 8.31 | 2.01 | 1.98 |
| Discharge | Prior-visits-first | 19.32 | 19.18 | 4.60 | 4.57 |
| Discharge | Simple logistic | 18.45 | 17.60 | 4.39 | 4.19 |
| Discharge | Risk-only | 23.69 | 25.00 | 5.64 | 5.95 |
| Discharge | Risk-plus-window | 23.69 | 24.72 | 5.64 | 5.89 |

Ten-minute reviews and 50 review-minutes per 100 encounters. Full-day ranks include later decisions; annual pooling includes the full year. These are descriptive benchmarks without queues or delays. Event reach is the proportion of all composite-event encounters selected with sufficient recorded review time. Review attempts without enough recorded time still count against the budget. Reach does not measure events prevented. Lift is reach relative to expected-value Random. Simple logistic indicates the five-variable logistic model. ESI indicates Emergency Severity Index.

| eTable 20. Selection, Reach, and Lift Across Ranking Budgets in 2023 | | | | | | | |
| --- | --- | --- | --- | --- | --- | --- | --- |
| Decision group | Allocation | Review min | Minutes per 100 | Rule | Selected % | Reach % | Lift |
| Admission | Full-day ranking | 5 | 25 | Random | 5.00 | 4.88 | 1.00 |
| Admission | Full-day ranking | 5 | 25 | ESI-first | 5.00 | 9.96 | 2.04 |
| Admission | Full-day ranking | 5 | 25 | Risk-only | 5.00 | 14.75 | 3.02 |
| Admission | Full-day ranking | 5 | 25 | Simple logistic | 5.00 | 12.95 | 2.65 |
| Admission | Full-day ranking | 5 | 25 | Age-first | 5.00 | 4.50 | 0.92 |
| Admission | Full-day ranking | 5 | 25 | Prior-visits-first | 5.00 | 10.76 | 2.20 |
| Admission | Full-day ranking | 5 | 50 | Random | 10.00 | 9.79 | 1.00 |
| Admission | Full-day ranking | 5 | 50 | ESI-first | 10.00 | 15.60 | 1.59 |
| Admission | Full-day ranking | 5 | 50 | Risk-only | 10.00 | 23.65 | 2.42 |
| Admission | Full-day ranking | 5 | 50 | Simple logistic | 10.00 | 22.93 | 2.34 |
| Admission | Full-day ranking | 5 | 50 | Age-first | 10.00 | 10.49 | 1.07 |
| Admission | Full-day ranking | 5 | 50 | Prior-visits-first | 10.00 | 18.59 | 1.90 |
| Admission | Full-day ranking | 5 | 100 | Random | 20.00 | 19.58 | 1.00 |
| Admission | Full-day ranking | 5 | 100 | ESI-first | 20.00 | 26.77 | 1.37 |
| Admission | Full-day ranking | 5 | 100 | Risk-only | 20.00 | 39.84 | 2.03 |
| Admission | Full-day ranking | 5 | 100 | Simple logistic | 20.00 | 36.96 | 1.89 |
| Admission | Full-day ranking | 5 | 100 | Age-first | 20.00 | 20.72 | 1.06 |
| Admission | Full-day ranking | 5 | 100 | Prior-visits-first | 20.00 | 30.48 | 1.56 |
| Admission | Full-day ranking | 10 | 25 | Random | 2.50 | 2.39 | 1.00 |
| Admission | Full-day ranking | 10 | 25 | ESI-first | 2.50 | 6.30 | 2.64 |
| Admission | Full-day ranking | 10 | 25 | Risk-only | 2.50 | 8.45 | 3.54 |
| Admission | Full-day ranking | 10 | 25 | Simple logistic | 2.50 | 7.28 | 3.05 |
| Admission | Full-day ranking | 10 | 25 | Age-first | 2.50 | 1.86 | 0.78 |
| Admission | Full-day ranking | 10 | 25 | Prior-visits-first | 2.50 | 5.83 | 2.44 |
| Admission | Full-day ranking | 10 | 50 | Random | 5.00 | 4.80 | 1.00 |
| Admission | Full-day ranking | 10 | 50 | ESI-first | 5.00 | 9.81 | 2.04 |
| Admission | Full-day ranking | 10 | 50 | Risk-only | 5.00 | 14.39 | 3.00 |
| Admission | Full-day ranking | 10 | 50 | Simple logistic | 5.00 | 12.68 | 2.64 |
| Admission | Full-day ranking | 10 | 50 | Age-first | 5.00 | 4.32 | 0.90 |
| Admission | Full-day ranking | 10 | 50 | Prior-visits-first | 5.00 | 10.49 | 2.19 |
| Admission | Full-day ranking | 10 | 100 | Random | 10.00 | 9.62 | 1.00 |
| Admission | Full-day ranking | 10 | 100 | ESI-first | 10.00 | 15.36 | 1.60 |
| Admission | Full-day ranking | 10 | 100 | Risk-only | 10.00 | 23.02 | 2.39 |
| Admission | Full-day ranking | 10 | 100 | Simple logistic | 10.00 | 22.48 | 2.34 |
| Admission | Full-day ranking | 10 | 100 | Age-first | 10.00 | 10.22 | 1.06 |
| Admission | Full-day ranking | 10 | 100 | Prior-visits-first | 10.00 | 18.32 | 1.90 |
| Admission | Full-day ranking | 15 | 25 | Random | 1.66 | 1.56 | 1.00 |
| Admission | Full-day ranking | 15 | 25 | ESI-first | 1.66 | 4.60 | 2.95 |
| Admission | Full-day ranking | 15 | 25 | Risk-only | 1.66 | 5.76 | 3.69 |
| Admission | Full-day ranking | 15 | 25 | Simple logistic | 1.66 | 5.04 | 3.23 |
| Admission | Full-day ranking | 15 | 25 | Age-first | 1.66 | 1.08 | 0.69 |
| Admission | Full-day ranking | 15 | 25 | Prior-visits-first | 1.66 | 3.95 | 2.53 |
| Admission | Full-day ranking | 15 | 50 | Random | 3.33 | 3.12 | 1.00 |
| Admission | Full-day ranking | 15 | 50 | ESI-first | 3.33 | 7.72 | 2.47 |
| Admission | Full-day ranking | 15 | 50 | Risk-only | 3.33 | 10.16 | 3.25 |
| Admission | Full-day ranking | 15 | 50 | Simple logistic | 3.33 | 9.44 | 3.02 |
| Admission | Full-day ranking | 15 | 50 | Age-first | 3.33 | 2.44 | 0.78 |
| Admission | Full-day ranking | 15 | 50 | Prior-visits-first | 3.33 | 7.26 | 2.33 |
| Admission | Full-day ranking | 15 | 100 | Random | 6.66 | 6.24 | 1.00 |
| Admission | Full-day ranking | 15 | 100 | ESI-first | 6.66 | 11.58 | 1.86 |
| Admission | Full-day ranking | 15 | 100 | Risk-only | 6.66 | 17.09 | 2.74 |
| Admission | Full-day ranking | 15 | 100 | Simple logistic | 6.66 | 16.19 | 2.59 |
| Admission | Full-day ranking | 15 | 100 | Age-first | 6.66 | 6.44 | 1.03 |
| Admission | Full-day ranking | 15 | 100 | Prior-visits-first | 6.66 | 12.83 | 2.06 |
| Admission | Annual ranking | 5 | 25 | Random | 5.00 | 4.89 | 1.00 |
| Admission | Annual ranking | 5 | 25 | ESI-first | 5.00 | 10.04 | 2.05 |
| Admission | Annual ranking | 5 | 25 | Risk-only | 5.00 | 14.66 | 3.00 |
| Admission | Annual ranking | 5 | 25 | Simple logistic | 5.00 | 14.57 | 2.98 |
| Admission | Annual ranking | 5 | 25 | Age-first | 5.00 | 4.85 | 0.99 |
| Admission | Annual ranking | 5 | 25 | Prior-visits-first | 5.00 | 11.02 | 2.25 |
| Admission | Annual ranking | 5 | 50 | Random | 10.00 | 9.78 | 1.00 |
| Admission | Annual ranking | 5 | 50 | ESI-first | 10.00 | 15.63 | 1.60 |
| Admission | Annual ranking | 5 | 50 | Risk-only | 10.00 | 25.54 | 2.61 |
| Admission | Annual ranking | 5 | 50 | Simple logistic | 10.00 | 22.84 | 2.33 |
| Admission | Annual ranking | 5 | 50 | Age-first | 10.00 | 10.99 | 1.12 |
| Admission | Annual ranking | 5 | 50 | Prior-visits-first | 10.00 | 18.54 | 1.90 |
| Admission | Annual ranking | 5 | 100 | Random | 20.00 | 19.57 | 1.00 |
| Admission | Annual ranking | 5 | 100 | ESI-first | 20.00 | 26.81 | 1.37 |
| Admission | Annual ranking | 5 | 100 | Risk-only | 20.00 | 39.93 | 2.04 |
| Admission | Annual ranking | 5 | 100 | Simple logistic | 20.00 | 37.23 | 1.90 |
| Admission | Annual ranking | 5 | 100 | Age-first | 20.00 | 20.34 | 1.04 |
| Admission | Annual ranking | 5 | 100 | Prior-visits-first | 20.00 | 30.81 | 1.57 |
| Admission | Annual ranking | 10 | 25 | Random | 2.50 | 2.40 | 1.00 |
| Admission | Annual ranking | 10 | 25 | ESI-first | 2.50 | 7.14 | 2.97 |
| Admission | Annual ranking | 10 | 25 | Risk-only | 2.50 | 8.63 | 3.59 |
| Admission | Annual ranking | 10 | 25 | Simple logistic | 2.50 | 8.00 | 3.33 |
| Admission | Annual ranking | 10 | 25 | Age-first | 2.50 | 1.88 | 0.78 |
| Admission | Annual ranking | 10 | 25 | Prior-visits-first | 2.50 | 6.82 | 2.84 |
| Admission | Annual ranking | 10 | 50 | Random | 5.00 | 4.81 | 1.00 |
| Admission | Annual ranking | 10 | 50 | ESI-first | 5.00 | 9.89 | 2.06 |
| Admission | Annual ranking | 10 | 50 | Risk-only | 5.00 | 14.39 | 2.99 |
| Admission | Annual ranking | 10 | 50 | Simple logistic | 5.00 | 14.30 | 2.97 |
| Admission | Annual ranking | 10 | 50 | Age-first | 5.00 | 4.67 | 0.97 |
| Admission | Annual ranking | 10 | 50 | Prior-visits-first | 5.00 | 10.75 | 2.24 |
| Admission | Annual ranking | 10 | 100 | Random | 10.00 | 9.61 | 1.00 |
| Admission | Annual ranking | 10 | 100 | ESI-first | 10.00 | 15.40 | 1.60 |
| Admission | Annual ranking | 10 | 100 | Risk-only | 10.00 | 25.00 | 2.60 |
| Admission | Annual ranking | 10 | 100 | Simple logistic | 10.00 | 22.48 | 2.34 |
| Admission | Annual ranking | 10 | 100 | Age-first | 10.00 | 10.72 | 1.12 |
| Admission | Annual ranking | 10 | 100 | Prior-visits-first | 10.00 | 18.27 | 1.90 |
| Admission | Annual ranking | 15 | 25 | Random | 1.66 | 1.56 | 1.00 |
| Admission | Annual ranking | 15 | 25 | ESI-first | 1.66 | 6.21 | 3.99 |
| Admission | Annual ranking | 15 | 25 | Risk-only | 1.66 | 6.21 | 3.98 |
| Admission | Annual ranking | 15 | 25 | Simple logistic | 1.66 | 5.40 | 3.46 |
| Admission | Annual ranking | 15 | 25 | Age-first | 1.66 | 1.04 | 0.67 |
| Admission | Annual ranking | 15 | 25 | Prior-visits-first | 1.66 | 4.73 | 3.03 |
| Admission | Annual ranking | 15 | 50 | Random | 3.33 | 3.12 | 1.00 |
| Admission | Annual ranking | 15 | 50 | ESI-first | 3.33 | 8.01 | 2.57 |
| Admission | Annual ranking | 15 | 50 | Risk-only | 3.33 | 10.25 | 3.29 |
| Admission | Annual ranking | 15 | 50 | Simple logistic | 3.33 | 9.80 | 3.14 |
| Admission | Annual ranking | 15 | 50 | Age-first | 3.33 | 2.93 | 0.94 |
| Admission | Annual ranking | 15 | 50 | Prior-visits-first | 3.33 | 7.69 | 2.47 |
| Admission | Annual ranking | 15 | 100 | Random | 6.66 | 6.24 | 1.00 |
| Admission | Annual ranking | 15 | 100 | ESI-first | 6.66 | 11.60 | 1.86 |
| Admission | Annual ranking | 15 | 100 | Risk-only | 6.66 | 17.54 | 2.81 |
| Admission | Annual ranking | 15 | 100 | Simple logistic | 6.66 | 17.27 | 2.77 |
| Admission | Annual ranking | 15 | 100 | Age-first | 6.66 | 6.68 | 1.07 |
| Admission | Annual ranking | 15 | 100 | Prior-visits-first | 6.66 | 12.92 | 2.07 |
| Discharge | Full-day ranking | 5 | 25 | Random | 5.00 | 4.56 | 1.00 |
| Discharge | Full-day ranking | 5 | 25 | ESI-first | 5.00 | 7.38 | 1.62 |
| Discharge | Full-day ranking | 5 | 25 | Risk-only | 5.00 | 25.28 | 5.54 |
| Discharge | Full-day ranking | 5 | 25 | Simple logistic | 5.00 | 19.48 | 4.27 |
| Discharge | Full-day ranking | 5 | 25 | Age-first | 5.00 | 8.83 | 1.93 |
| Discharge | Full-day ranking | 5 | 25 | Prior-visits-first | 5.00 | 20.52 | 4.50 |
| Discharge | Full-day ranking | 5 | 50 | Random | 10.00 | 9.12 | 1.00 |
| Discharge | Full-day ranking | 5 | 50 | ESI-first | 10.00 | 14.65 | 1.61 |
| Discharge | Full-day ranking | 5 | 50 | Risk-only | 10.00 | 39.42 | 4.32 |
| Discharge | Full-day ranking | 5 | 50 | Simple logistic | 10.00 | 31.65 | 3.47 |
| Discharge | Full-day ranking | 5 | 50 | Age-first | 10.00 | 15.97 | 1.75 |
| Discharge | Full-day ranking | 5 | 50 | Prior-visits-first | 10.00 | 31.34 | 3.44 |
| Discharge | Full-day ranking | 5 | 100 | Random | 20.00 | 18.26 | 1.00 |
| Discharge | Full-day ranking | 5 | 100 | ESI-first | 20.00 | 26.99 | 1.48 |
| Discharge | Full-day ranking | 5 | 100 | Risk-only | 20.00 | 57.49 | 3.15 |
| Discharge | Full-day ranking | 5 | 100 | Simple logistic | 20.00 | 46.35 | 2.54 |
| Discharge | Full-day ranking | 5 | 100 | Age-first | 20.00 | 31.04 | 1.70 |
| Discharge | Full-day ranking | 5 | 100 | Prior-visits-first | 20.00 | 45.84 | 2.51 |
| Discharge | Full-day ranking | 10 | 25 | Random | 2.50 | 2.10 | 1.00 |
| Discharge | Full-day ranking | 10 | 25 | ESI-first | 2.50 | 3.46 | 1.64 |
| Discharge | Full-day ranking | 10 | 25 | Risk-only | 2.50 | 16.20 | 7.71 |
| Discharge | Full-day ranking | 10 | 25 | Simple logistic | 2.50 | 11.14 | 5.30 |
| Discharge | Full-day ranking | 10 | 25 | Age-first | 2.50 | 4.28 | 2.03 |
| Discharge | Full-day ranking | 10 | 25 | Prior-visits-first | 2.50 | 10.37 | 4.94 |
| Discharge | Full-day ranking | 10 | 50 | Random | 5.00 | 4.20 | 1.00 |
| Discharge | Full-day ranking | 10 | 50 | ESI-first | 5.00 | 6.70 | 1.60 |
| Discharge | Full-day ranking | 10 | 50 | Risk-only | 5.00 | 23.69 | 5.64 |
| Discharge | Full-day ranking | 10 | 50 | Simple logistic | 5.00 | 18.45 | 4.39 |
| Discharge | Full-day ranking | 10 | 50 | Age-first | 5.00 | 8.43 | 2.01 |
| Discharge | Full-day ranking | 10 | 50 | Prior-visits-first | 5.00 | 19.32 | 4.60 |
| Discharge | Full-day ranking | 10 | 100 | Random | 10.00 | 8.39 | 1.00 |
| Discharge | Full-day ranking | 10 | 100 | ESI-first | 10.00 | 13.32 | 1.59 |
| Discharge | Full-day ranking | 10 | 100 | Risk-only | 10.00 | 36.70 | 4.37 |
| Discharge | Full-day ranking | 10 | 100 | Simple logistic | 10.00 | 30.06 | 3.58 |
| Discharge | Full-day ranking | 10 | 100 | Age-first | 10.00 | 14.88 | 1.77 |
| Discharge | Full-day ranking | 10 | 100 | Prior-visits-first | 10.00 | 29.83 | 3.55 |
| Discharge | Full-day ranking | 15 | 25 | Random | 1.67 | 1.25 | 1.00 |
| Discharge | Full-day ranking | 15 | 25 | ESI-first | 1.67 | 2.12 | 1.70 |
| Discharge | Full-day ranking | 15 | 25 | Risk-only | 1.67 | 11.24 | 8.98 |
| Discharge | Full-day ranking | 15 | 25 | Simple logistic | 1.67 | 7.02 | 5.61 |
| Discharge | Full-day ranking | 15 | 25 | Age-first | 1.67 | 2.78 | 2.22 |
| Discharge | Full-day ranking | 15 | 25 | Prior-visits-first | 1.67 | 6.27 | 5.01 |
| Discharge | Full-day ranking | 15 | 50 | Random | 3.33 | 2.48 | 1.00 |
| Discharge | Full-day ranking | 15 | 50 | ESI-first | 3.33 | 4.05 | 1.63 |
| Discharge | Full-day ranking | 15 | 50 | Risk-only | 3.33 | 18.35 | 7.39 |
| Discharge | Full-day ranking | 15 | 50 | Simple logistic | 3.33 | 12.36 | 4.98 |
| Discharge | Full-day ranking | 15 | 50 | Age-first | 3.33 | 5.34 | 2.15 |
| Discharge | Full-day ranking | 15 | 50 | Prior-visits-first | 3.33 | 11.18 | 4.50 |
| Discharge | Full-day ranking | 15 | 100 | Random | 6.66 | 4.96 | 1.00 |
| Discharge | Full-day ranking | 15 | 100 | ESI-first | 6.66 | 7.96 | 1.60 |
| Discharge | Full-day ranking | 15 | 100 | Risk-only | 6.66 | 26.03 | 5.24 |
| Discharge | Full-day ranking | 15 | 100 | Simple logistic | 6.66 | 20.22 | 4.07 |
| Discharge | Full-day ranking | 15 | 100 | Age-first | 6.66 | 9.70 | 1.95 |
| Discharge | Full-day ranking | 15 | 100 | Prior-visits-first | 6.66 | 20.47 | 4.12 |
| Discharge | Annual ranking | 5 | 25 | Random | 5.00 | 4.56 | 1.00 |
| Discharge | Annual ranking | 5 | 25 | ESI-first | 5.00 | 7.29 | 1.60 |
| Discharge | Annual ranking | 5 | 25 | Risk-only | 5.00 | 26.78 | 5.87 |
| Discharge | Annual ranking | 5 | 25 | Simple logistic | 5.00 | 18.35 | 4.02 |
| Discharge | Annual ranking | 5 | 25 | Age-first | 5.00 | 8.80 | 1.93 |
| Discharge | Annual ranking | 5 | 25 | Prior-visits-first | 5.00 | 20.31 | 4.45 |
| Discharge | Annual ranking | 5 | 50 | Random | 10.00 | 9.13 | 1.00 |
| Discharge | Annual ranking | 5 | 50 | ESI-first | 10.00 | 14.52 | 1.59 |
| Discharge | Annual ranking | 5 | 50 | Risk-only | 10.00 | 40.26 | 4.41 |
| Discharge | Annual ranking | 5 | 50 | Simple logistic | 10.00 | 30.90 | 3.38 |
| Discharge | Annual ranking | 5 | 50 | Age-first | 10.00 | 16.62 | 1.82 |
| Discharge | Annual ranking | 5 | 50 | Prior-visits-first | 10.00 | 32.38 | 3.55 |
| Discharge | Annual ranking | 5 | 100 | Random | 20.00 | 18.26 | 1.00 |
| Discharge | Annual ranking | 5 | 100 | ESI-first | 20.00 | 27.11 | 1.48 |
| Discharge | Annual ranking | 5 | 100 | Risk-only | 20.00 | 57.49 | 3.15 |
| Discharge | Annual ranking | 5 | 100 | Simple logistic | 20.00 | 45.51 | 2.49 |
| Discharge | Annual ranking | 5 | 100 | Age-first | 20.00 | 31.35 | 1.72 |
| Discharge | Annual ranking | 5 | 100 | Prior-visits-first | 20.00 | 45.85 | 2.51 |
| Discharge | Annual ranking | 10 | 25 | Random | 2.50 | 2.10 | 1.00 |
| Discharge | Annual ranking | 10 | 25 | ESI-first | 2.50 | 3.38 | 1.61 |
| Discharge | Annual ranking | 10 | 25 | Risk-only | 2.50 | 15.17 | 7.23 |
| Discharge | Annual ranking | 10 | 25 | Simple logistic | 2.50 | 12.17 | 5.80 |
| Discharge | Annual ranking | 10 | 25 | Age-first | 2.50 | 4.33 | 2.07 |
| Discharge | Annual ranking | 10 | 25 | Prior-visits-first | 2.50 | 11.27 | 5.37 |
| Discharge | Annual ranking | 10 | 50 | Random | 5.00 | 4.20 | 1.00 |
| Discharge | Annual ranking | 10 | 50 | ESI-first | 5.00 | 6.69 | 1.59 |
| Discharge | Annual ranking | 10 | 50 | Risk-only | 5.00 | 25.00 | 5.95 |
| Discharge | Annual ranking | 10 | 50 | Simple logistic | 5.00 | 17.60 | 4.19 |
| Discharge | Annual ranking | 10 | 50 | Age-first | 5.00 | 8.31 | 1.98 |
| Discharge | Annual ranking | 10 | 50 | Prior-visits-first | 5.00 | 19.18 | 4.57 |
| Discharge | Annual ranking | 10 | 100 | Random | 10.00 | 8.40 | 1.00 |
| Discharge | Annual ranking | 10 | 100 | ESI-first | 10.00 | 13.31 | 1.58 |
| Discharge | Annual ranking | 10 | 100 | Risk-only | 10.00 | 37.45 | 4.46 |
| Discharge | Annual ranking | 10 | 100 | Simple logistic | 10.00 | 29.40 | 3.50 |
| Discharge | Annual ranking | 10 | 100 | Age-first | 10.00 | 15.56 | 1.85 |
| Discharge | Annual ranking | 10 | 100 | Prior-visits-first | 10.00 | 30.94 | 3.68 |
| Discharge | Annual ranking | 15 | 25 | Random | 1.67 | 1.24 | 1.00 |
| Discharge | Annual ranking | 15 | 25 | ESI-first | 1.67 | 2.07 | 1.67 |
| Discharge | Annual ranking | 15 | 25 | Risk-only | 1.67 | 10.39 | 8.38 |
| Discharge | Annual ranking | 15 | 25 | Simple logistic | 1.67 | 8.33 | 6.72 |
| Discharge | Annual ranking | 15 | 25 | Age-first | 1.67 | 2.65 | 2.14 |
| Discharge | Annual ranking | 15 | 25 | Prior-visits-first | 1.67 | 7.06 | 5.69 |
| Discharge | Annual ranking | 15 | 50 | Random | 3.33 | 2.48 | 1.00 |
| Discharge | Annual ranking | 15 | 50 | ESI-first | 3.33 | 4.07 | 1.64 |
| Discharge | Annual ranking | 15 | 50 | Risk-only | 3.33 | 17.32 | 6.98 |
| Discharge | Annual ranking | 15 | 50 | Simple logistic | 3.33 | 12.17 | 4.91 |
| Discharge | Annual ranking | 15 | 50 | Age-first | 3.33 | 5.12 | 2.06 |
| Discharge | Annual ranking | 15 | 50 | Prior-visits-first | 3.33 | 12.37 | 4.99 |
| Discharge | Annual ranking | 15 | 100 | Random | 6.66 | 4.96 | 1.00 |
| Discharge | Annual ranking | 15 | 100 | ESI-first | 6.66 | 8.05 | 1.62 |
| Discharge | Annual ranking | 15 | 100 | Risk-only | 6.66 | 26.78 | 5.40 |
| Discharge | Annual ranking | 15 | 100 | Simple logistic | 6.66 | 19.94 | 4.02 |
| Discharge | Annual ranking | 15 | 100 | Age-first | 6.66 | 9.67 | 1.95 |
| Discharge | Annual ranking | 15 | 100 | Prior-visits-first | 6.66 | 20.50 | 4.13 |
| Admission | Annual ranking | 5 | 25 | Risk-plus-window | 5.00 | 14.57 | 2.98 |
| Admission | Annual ranking | 5 | 50 | Risk-plus-window | 10.00 | 25.36 | 2.59 |
| Admission | Annual ranking | 5 | 100 | Risk-plus-window | 20.00 | 40.02 | 2.05 |
| Admission | Annual ranking | 10 | 25 | Risk-plus-window | 2.50 | 8.54 | 3.55 |
| Admission | Annual ranking | 10 | 50 | Risk-plus-window | 5.00 | 14.12 | 2.94 |
| Admission | Annual ranking | 10 | 100 | Risk-plus-window | 10.00 | 24.73 | 2.57 |
| Admission | Annual ranking | 15 | 25 | Risk-plus-window | 1.66 | 6.38 | 4.10 |
| Admission | Annual ranking | 15 | 50 | Risk-plus-window | 3.33 | 10.34 | 3.32 |
| Admission | Annual ranking | 15 | 100 | Risk-plus-window | 6.66 | 17.27 | 2.77 |
| Discharge | Annual ranking | 5 | 25 | Risk-plus-window | 5.00 | 26.69 | 5.85 |
| Discharge | Annual ranking | 5 | 50 | Risk-plus-window | 10.00 | 40.36 | 4.42 |
| Discharge | Annual ranking | 5 | 100 | Risk-plus-window | 20.00 | 56.84 | 3.11 |
| Discharge | Annual ranking | 10 | 25 | Risk-plus-window | 2.50 | 15.82 | 7.54 |
| Discharge | Annual ranking | 10 | 50 | Risk-plus-window | 5.00 | 24.72 | 5.89 |
| Discharge | Annual ranking | 10 | 100 | Risk-plus-window | 10.00 | 37.45 | 4.46 |
| Discharge | Annual ranking | 15 | 25 | Risk-plus-window | 1.67 | 11.42 | 9.21 |
| Discharge | Annual ranking | 15 | 50 | Risk-plus-window | 3.33 | 18.16 | 7.32 |
| Discharge | Annual ranking | 15 | 100 | Risk-plus-window | 6.66 | 26.31 | 5.30 |
| Admission | Full-day ranking | 5 | 25 | Risk-plus-window | 5.00 | 14.84 | 3.04 |
| Admission | Full-day ranking | 5 | 50 | Risk-plus-window | 10.00 | 23.56 | 2.41 |
| Admission | Full-day ranking | 5 | 100 | Risk-plus-window | 20.00 | 39.84 | 2.03 |
| Admission | Full-day ranking | 10 | 25 | Risk-plus-window | 2.50 | 8.36 | 3.50 |
| Admission | Full-day ranking | 10 | 50 | Risk-plus-window | 5.00 | 14.66 | 3.05 |
| Admission | Full-day ranking | 10 | 100 | Risk-plus-window | 10.00 | 22.93 | 2.38 |
| Admission | Full-day ranking | 15 | 25 | Risk-plus-window | 1.66 | 5.85 | 3.74 |
| Admission | Full-day ranking | 15 | 50 | Risk-plus-window | 3.33 | 9.98 | 3.20 |
| Admission | Full-day ranking | 15 | 100 | Risk-plus-window | 6.66 | 16.82 | 2.70 |
| Discharge | Full-day ranking | 5 | 25 | Risk-plus-window | 5.00 | 24.72 | 5.42 |
| Discharge | Full-day ranking | 5 | 50 | Risk-plus-window | 10.00 | 38.76 | 4.25 |
| Discharge | Full-day ranking | 5 | 100 | Risk-plus-window | 20.00 | 56.93 | 3.12 |
| Discharge | Full-day ranking | 10 | 25 | Risk-plus-window | 2.50 | 16.29 | 7.75 |
| Discharge | Full-day ranking | 10 | 50 | Risk-plus-window | 5.00 | 23.69 | 5.64 |
| Discharge | Full-day ranking | 10 | 100 | Risk-plus-window | 10.00 | 36.33 | 4.33 |
| Discharge | Full-day ranking | 15 | 25 | Risk-plus-window | 1.67 | 11.70 | 9.36 |
| Discharge | Full-day ranking | 15 | 50 | Risk-plus-window | 3.33 | 18.16 | 7.32 |
| Discharge | Full-day ranking | 15 | 100 | Risk-plus-window | 6.66 | 26.31 | 5.30 |

Full-day and annual ranking of eligible 2023 encounters. Durations and budgets are shown in each row. Descriptive points. Event reach is the proportion of all composite-event encounters selected with sufficient recorded review time. Review attempts without enough recorded time still count against the budget. Reach does not measure events prevented. Lift is relative to expected-value Random at the same budget. Simple logistic indicates the five-variable logistic model. ESI indicates Emergency Severity Index.

| eTable 21. Review Yield Across Ranking Budgets in 2023 | | | | | | |
| --- | --- | --- | --- | --- | --- | --- |
| Decision group | Allocation | Review min | Minutes per 100 | Rule | Events per review % | Reviews per event |
| Admission | Full-day ranking | 5 | 25 | Random | 4.45 | 22.47 |
| Admission | Full-day ranking | 5 | 25 | ESI-first | 9.08 | 11.01 |
| Admission | Full-day ranking | 5 | 25 | Risk-only | 13.44 | 7.44 |
| Admission | Full-day ranking | 5 | 25 | Simple logistic | 11.80 | 8.47 |
| Admission | Full-day ranking | 5 | 25 | Age-first | 4.11 | 24.36 |
| Admission | Full-day ranking | 5 | 25 | Prior-visits-first | 9.80 | 10.20 |
| Admission | Full-day ranking | 5 | 50 | Random | 4.46 | 22.41 |
| Admission | Full-day ranking | 5 | 50 | ESI-first | 7.11 | 14.07 |
| Admission | Full-day ranking | 5 | 50 | Risk-only | 10.78 | 9.28 |
| Admission | Full-day ranking | 5 | 50 | Simple logistic | 10.45 | 9.57 |
| Admission | Full-day ranking | 5 | 50 | Age-first | 4.78 | 20.91 |
| Admission | Full-day ranking | 5 | 50 | Prior-visits-first | 8.47 | 11.80 |
| Admission | Full-day ranking | 5 | 100 | Random | 4.46 | 22.41 |
| Admission | Full-day ranking | 5 | 100 | ESI-first | 6.10 | 16.39 |
| Admission | Full-day ranking | 5 | 100 | Risk-only | 9.08 | 11.02 |
| Admission | Full-day ranking | 5 | 100 | Simple logistic | 8.42 | 11.87 |
| Admission | Full-day ranking | 5 | 100 | Age-first | 4.72 | 21.18 |
| Admission | Full-day ranking | 5 | 100 | Prior-visits-first | 6.95 | 14.40 |
| Admission | Full-day ranking | 10 | 25 | Random | 4.35 | 22.98 |
| Admission | Full-day ranking | 10 | 25 | ESI-first | 11.49 | 8.70 |
| Admission | Full-day ranking | 10 | 25 | Risk-only | 15.41 | 6.49 |
| Admission | Full-day ranking | 10 | 25 | Simple logistic | 13.28 | 7.53 |
| Admission | Full-day ranking | 10 | 25 | Age-first | 3.39 | 29.52 |
| Admission | Full-day ranking | 10 | 25 | Prior-visits-first | 10.63 | 9.40 |
| Admission | Full-day ranking | 10 | 50 | Random | 4.37 | 22.86 |
| Admission | Full-day ranking | 10 | 50 | ESI-first | 8.94 | 11.18 |
| Admission | Full-day ranking | 10 | 50 | Risk-only | 13.11 | 7.63 |
| Admission | Full-day ranking | 10 | 50 | Simple logistic | 11.56 | 8.65 |
| Admission | Full-day ranking | 10 | 50 | Age-first | 3.94 | 25.37 |
| Admission | Full-day ranking | 10 | 50 | Prior-visits-first | 9.56 | 10.46 |
| Admission | Full-day ranking | 10 | 100 | Random | 4.38 | 22.81 |
| Admission | Full-day ranking | 10 | 100 | ESI-first | 7.00 | 14.29 |
| Admission | Full-day ranking | 10 | 100 | Risk-only | 10.49 | 9.53 |
| Admission | Full-day ranking | 10 | 100 | Simple logistic | 10.25 | 9.76 |
| Admission | Full-day ranking | 10 | 100 | Age-first | 4.66 | 21.47 |
| Admission | Full-day ranking | 10 | 100 | Prior-visits-first | 8.35 | 11.98 |
| Admission | Full-day ranking | 15 | 25 | Random | 4.28 | 23.38 |
| Admission | Full-day ranking | 15 | 25 | ESI-first | 12.61 | 7.93 |
| Admission | Full-day ranking | 15 | 25 | Risk-only | 15.76 | 6.34 |
| Admission | Full-day ranking | 15 | 25 | Simple logistic | 13.79 | 7.25 |
| Admission | Full-day ranking | 15 | 25 | Age-first | 2.96 | 33.83 |
| Admission | Full-day ranking | 15 | 25 | Prior-visits-first | 10.82 | 9.24 |
| Admission | Full-day ranking | 15 | 50 | Random | 4.27 | 23.41 |
| Admission | Full-day ranking | 15 | 50 | ESI-first | 10.56 | 9.47 |
| Admission | Full-day ranking | 15 | 50 | Risk-only | 13.90 | 7.19 |
| Admission | Full-day ranking | 15 | 50 | Simple logistic | 12.92 | 7.74 |
| Admission | Full-day ranking | 15 | 50 | Age-first | 3.34 | 29.93 |
| Admission | Full-day ranking | 15 | 50 | Prior-visits-first | 9.93 | 10.07 |
| Admission | Full-day ranking | 15 | 100 | Random | 4.27 | 23.44 |
| Admission | Full-day ranking | 15 | 100 | ESI-first | 7.92 | 12.62 |
| Admission | Full-day ranking | 15 | 100 | Risk-only | 11.69 | 8.56 |
| Admission | Full-day ranking | 15 | 100 | Simple logistic | 11.07 | 9.03 |
| Admission | Full-day ranking | 15 | 100 | Age-first | 4.41 | 22.69 |
| Admission | Full-day ranking | 15 | 100 | Prior-visits-first | 8.77 | 11.40 |
| Admission | Annual ranking | 5 | 25 | Random | 4.46 | 22.43 |
| Admission | Annual ranking | 5 | 25 | ESI-first | 9.15 | 10.93 |
| Admission | Annual ranking | 5 | 25 | Risk-only | 13.36 | 7.48 |
| Admission | Annual ranking | 5 | 25 | Simple logistic | 13.28 | 7.53 |
| Admission | Annual ranking | 5 | 25 | Age-first | 4.42 | 22.62 |
| Admission | Annual ranking | 5 | 25 | Prior-visits-first | 10.04 | 9.96 |
| Admission | Annual ranking | 5 | 50 | Random | 4.46 | 22.43 |
| Admission | Annual ranking | 5 | 50 | ESI-first | 7.12 | 14.04 |
| Admission | Annual ranking | 5 | 50 | Risk-only | 11.64 | 8.59 |
| Admission | Annual ranking | 5 | 50 | Simple logistic | 10.41 | 9.61 |
| Admission | Annual ranking | 5 | 50 | Age-first | 5.01 | 19.97 |
| Admission | Annual ranking | 5 | 50 | Prior-visits-first | 8.45 | 11.83 |
| Admission | Annual ranking | 5 | 100 | Random | 4.46 | 22.43 |
| Admission | Annual ranking | 5 | 100 | ESI-first | 6.11 | 16.37 |
| Admission | Annual ranking | 5 | 100 | Risk-only | 9.10 | 10.99 |
| Admission | Annual ranking | 5 | 100 | Simple logistic | 8.48 | 11.79 |
| Admission | Annual ranking | 5 | 100 | Age-first | 4.63 | 21.58 |
| Admission | Annual ranking | 5 | 100 | Prior-visits-first | 7.02 | 14.24 |
| Admission | Annual ranking | 10 | 25 | Random | 4.38 | 22.83 |
| Admission | Annual ranking | 10 | 25 | ESI-first | 13.01 | 7.69 |
| Admission | Annual ranking | 10 | 25 | Risk-only | 15.74 | 6.35 |
| Admission | Annual ranking | 10 | 25 | Simple logistic | 14.59 | 6.85 |
| Admission | Annual ranking | 10 | 25 | Age-first | 3.43 | 29.16 |
| Admission | Annual ranking | 10 | 25 | Prior-visits-first | 12.43 | 8.04 |
| Admission | Annual ranking | 10 | 50 | Random | 4.38 | 22.83 |
| Admission | Annual ranking | 10 | 50 | ESI-first | 9.01 | 11.09 |
| Admission | Annual ranking | 10 | 50 | Risk-only | 13.11 | 7.63 |
| Admission | Annual ranking | 10 | 50 | Simple logistic | 13.03 | 7.67 |
| Admission | Annual ranking | 10 | 50 | Age-first | 4.26 | 23.49 |
| Admission | Annual ranking | 10 | 50 | Prior-visits-first | 9.80 | 10.21 |
| Admission | Annual ranking | 10 | 100 | Random | 4.38 | 22.83 |
| Admission | Annual ranking | 10 | 100 | ESI-first | 7.02 | 14.25 |
| Admission | Annual ranking | 10 | 100 | Risk-only | 11.39 | 8.78 |
| Admission | Annual ranking | 10 | 100 | Simple logistic | 10.25 | 9.76 |
| Admission | Annual ranking | 10 | 100 | Age-first | 4.89 | 20.47 |
| Admission | Annual ranking | 10 | 100 | Prior-visits-first | 8.33 | 12.01 |
| Admission | Annual ranking | 15 | 25 | Random | 4.27 | 23.44 |
| Admission | Annual ranking | 15 | 25 | ESI-first | 17.01 | 5.88 |
| Admission | Annual ranking | 15 | 25 | Risk-only | 17.00 | 5.88 |
| Admission | Annual ranking | 15 | 25 | Simple logistic | 14.78 | 6.77 |
| Admission | Annual ranking | 15 | 25 | Age-first | 2.85 | 35.10 |
| Admission | Annual ranking | 15 | 25 | Prior-visits-first | 12.94 | 7.73 |
| Admission | Annual ranking | 15 | 50 | Random | 4.27 | 23.44 |
| Admission | Annual ranking | 15 | 50 | ESI-first | 10.96 | 9.13 |
| Admission | Annual ranking | 15 | 50 | Risk-only | 14.02 | 7.13 |
| Admission | Annual ranking | 15 | 50 | Simple logistic | 13.41 | 7.46 |
| Admission | Annual ranking | 15 | 50 | Age-first | 4.01 | 24.95 |
| Admission | Annual ranking | 15 | 50 | Prior-visits-first | 10.52 | 9.50 |
| Admission | Annual ranking | 15 | 100 | Random | 4.27 | 23.44 |
| Admission | Annual ranking | 15 | 100 | ESI-first | 7.94 | 12.60 |
| Admission | Annual ranking | 15 | 100 | Risk-only | 11.99 | 8.34 |
| Admission | Annual ranking | 15 | 100 | Simple logistic | 11.81 | 8.47 |
| Admission | Annual ranking | 15 | 100 | Age-first | 4.57 | 21.88 |
| Admission | Annual ranking | 15 | 100 | Prior-visits-first | 8.84 | 11.31 |
| Discharge | Full-day ranking | 5 | 25 | Random | 2.02 | 49.54 |
| Discharge | Full-day ranking | 5 | 25 | ESI-first | 3.26 | 30.63 |
| Discharge | Full-day ranking | 5 | 25 | Risk-only | 11.18 | 8.94 |
| Discharge | Full-day ranking | 5 | 25 | Simple logistic | 8.61 | 11.61 |
| Discharge | Full-day ranking | 5 | 25 | Age-first | 3.91 | 25.61 |
| Discharge | Full-day ranking | 5 | 25 | Prior-visits-first | 9.07 | 11.02 |
| Discharge | Full-day ranking | 5 | 50 | Random | 2.02 | 49.59 |
| Discharge | Full-day ranking | 5 | 50 | ESI-first | 3.24 | 30.87 |
| Discharge | Full-day ranking | 5 | 50 | Risk-only | 8.71 | 11.48 |
| Discharge | Full-day ranking | 5 | 50 | Simple logistic | 7.00 | 14.29 |
| Discharge | Full-day ranking | 5 | 50 | Age-first | 3.53 | 28.33 |
| Discharge | Full-day ranking | 5 | 50 | Prior-visits-first | 6.93 | 14.43 |
| Discharge | Full-day ranking | 5 | 100 | Random | 2.02 | 49.54 |
| Discharge | Full-day ranking | 5 | 100 | ESI-first | 2.98 | 33.52 |
| Discharge | Full-day ranking | 5 | 100 | Risk-only | 6.35 | 15.74 |
| Discharge | Full-day ranking | 5 | 100 | Simple logistic | 5.12 | 19.52 |
| Discharge | Full-day ranking | 5 | 100 | Age-first | 3.43 | 29.15 |
| Discharge | Full-day ranking | 5 | 100 | Prior-visits-first | 5.07 | 19.74 |
| Discharge | Full-day ranking | 10 | 25 | Random | 1.86 | 53.78 |
| Discharge | Full-day ranking | 10 | 25 | ESI-first | 3.06 | 32.70 |
| Discharge | Full-day ranking | 10 | 25 | Risk-only | 14.33 | 6.98 |
| Discharge | Full-day ranking | 10 | 25 | Simple logistic | 9.86 | 10.14 |
| Discharge | Full-day ranking | 10 | 25 | Age-first | 3.78 | 26.43 |
| Discharge | Full-day ranking | 10 | 25 | Prior-visits-first | 9.18 | 10.90 |
| Discharge | Full-day ranking | 10 | 50 | Random | 1.86 | 53.84 |
| Discharge | Full-day ranking | 10 | 50 | ESI-first | 2.96 | 33.76 |
| Discharge | Full-day ranking | 10 | 50 | Risk-only | 10.48 | 9.55 |
| Discharge | Full-day ranking | 10 | 50 | Simple logistic | 8.16 | 12.26 |
| Discharge | Full-day ranking | 10 | 50 | Age-first | 3.73 | 26.84 |
| Discharge | Full-day ranking | 10 | 50 | Prior-visits-first | 8.54 | 11.70 |
| Discharge | Full-day ranking | 10 | 100 | Random | 1.86 | 53.90 |
| Discharge | Full-day ranking | 10 | 100 | ESI-first | 2.94 | 33.97 |
| Discharge | Full-day ranking | 10 | 100 | Risk-only | 8.11 | 12.32 |
| Discharge | Full-day ranking | 10 | 100 | Simple logistic | 6.64 | 15.05 |
| Discharge | Full-day ranking | 10 | 100 | Age-first | 3.29 | 30.41 |
| Discharge | Full-day ranking | 10 | 100 | Prior-visits-first | 6.60 | 15.16 |
| Discharge | Full-day ranking | 15 | 25 | Random | 1.66 | 60.25 |
| Discharge | Full-day ranking | 15 | 25 | ESI-first | 2.82 | 35.51 |
| Discharge | Full-day ranking | 15 | 25 | Risk-only | 14.91 | 6.71 |
| Discharge | Full-day ranking | 15 | 25 | Simple logistic | 9.32 | 10.73 |
| Discharge | Full-day ranking | 15 | 25 | Age-first | 3.69 | 27.13 |
| Discharge | Full-day ranking | 15 | 25 | Prior-visits-first | 8.31 | 12.03 |
| Discharge | Full-day ranking | 15 | 50 | Random | 1.65 | 60.71 |
| Discharge | Full-day ranking | 15 | 50 | ESI-first | 2.69 | 37.23 |
| Discharge | Full-day ranking | 15 | 50 | Risk-only | 12.17 | 8.21 |
| Discharge | Full-day ranking | 15 | 50 | Simple logistic | 8.20 | 12.20 |
| Discharge | Full-day ranking | 15 | 50 | Age-first | 3.55 | 28.20 |
| Discharge | Full-day ranking | 15 | 50 | Prior-visits-first | 7.42 | 13.48 |
| Discharge | Full-day ranking | 15 | 100 | Random | 1.65 | 60.73 |
| Discharge | Full-day ranking | 15 | 100 | ESI-first | 2.64 | 37.86 |
| Discharge | Full-day ranking | 15 | 100 | Risk-only | 8.63 | 11.58 |
| Discharge | Full-day ranking | 15 | 100 | Simple logistic | 6.71 | 14.91 |
| Discharge | Full-day ranking | 15 | 100 | Age-first | 3.22 | 31.10 |
| Discharge | Full-day ranking | 15 | 100 | Prior-visits-first | 6.79 | 14.73 |
| Discharge | Annual ranking | 5 | 25 | Random | 2.02 | 49.55 |
| Discharge | Annual ranking | 5 | 25 | ESI-first | 3.22 | 31.03 |
| Discharge | Annual ranking | 5 | 25 | Risk-only | 11.84 | 8.44 |
| Discharge | Annual ranking | 5 | 25 | Simple logistic | 8.12 | 12.32 |
| Discharge | Annual ranking | 5 | 25 | Age-first | 3.89 | 25.71 |
| Discharge | Annual ranking | 5 | 25 | Prior-visits-first | 8.98 | 11.14 |
| Discharge | Annual ranking | 5 | 50 | Random | 2.02 | 49.55 |
| Discharge | Annual ranking | 5 | 50 | ESI-first | 3.21 | 31.15 |
| Discharge | Annual ranking | 5 | 50 | Risk-only | 8.90 | 11.23 |
| Discharge | Annual ranking | 5 | 50 | Simple logistic | 6.83 | 14.64 |
| Discharge | Annual ranking | 5 | 50 | Age-first | 3.67 | 27.22 |
| Discharge | Annual ranking | 5 | 50 | Prior-visits-first | 7.16 | 13.97 |
| Discharge | Annual ranking | 5 | 100 | Random | 2.02 | 49.55 |
| Discharge | Annual ranking | 5 | 100 | ESI-first | 3.00 | 33.37 |
| Discharge | Annual ranking | 5 | 100 | Risk-only | 6.35 | 15.74 |
| Discharge | Annual ranking | 5 | 100 | Simple logistic | 5.03 | 19.88 |
| Discharge | Annual ranking | 5 | 100 | Age-first | 3.46 | 28.86 |
| Discharge | Annual ranking | 5 | 100 | Prior-visits-first | 5.07 | 19.73 |
| Discharge | Annual ranking | 10 | 25 | Random | 1.86 | 53.86 |
| Discharge | Annual ranking | 10 | 25 | ESI-first | 2.99 | 33.46 |
| Discharge | Annual ranking | 10 | 25 | Risk-only | 13.42 | 7.45 |
| Discharge | Annual ranking | 10 | 25 | Simple logistic | 10.77 | 9.28 |
| Discharge | Annual ranking | 10 | 25 | Age-first | 3.84 | 26.07 |
| Discharge | Annual ranking | 10 | 25 | Prior-visits-first | 9.98 | 10.02 |
| Discharge | Annual ranking | 10 | 50 | Random | 1.86 | 53.86 |
| Discharge | Annual ranking | 10 | 50 | ESI-first | 2.96 | 33.81 |
| Discharge | Annual ranking | 10 | 50 | Risk-only | 11.06 | 9.04 |
| Discharge | Annual ranking | 10 | 50 | Simple logistic | 7.78 | 12.85 |
| Discharge | Annual ranking | 10 | 50 | Age-first | 3.67 | 27.22 |
| Discharge | Annual ranking | 10 | 50 | Prior-visits-first | 8.48 | 11.79 |
| Discharge | Annual ranking | 10 | 100 | Random | 1.86 | 53.86 |
| Discharge | Annual ranking | 10 | 100 | ESI-first | 2.94 | 33.98 |
| Discharge | Annual ranking | 10 | 100 | Risk-only | 8.28 | 12.08 |
| Discharge | Annual ranking | 10 | 100 | Simple logistic | 6.50 | 15.39 |
| Discharge | Annual ranking | 10 | 100 | Age-first | 3.44 | 29.08 |
| Discharge | Annual ranking | 10 | 100 | Prior-visits-first | 6.84 | 14.62 |
| Discharge | Annual ranking | 15 | 25 | Random | 1.65 | 60.77 |
| Discharge | Annual ranking | 15 | 25 | ESI-first | 2.75 | 36.34 |
| Discharge | Annual ranking | 15 | 25 | Risk-only | 13.79 | 7.25 |
| Discharge | Annual ranking | 15 | 25 | Simple logistic | 11.06 | 9.04 |
| Discharge | Annual ranking | 15 | 25 | Age-first | 3.52 | 28.42 |
| Discharge | Annual ranking | 15 | 25 | Prior-visits-first | 9.36 | 10.68 |
| Discharge | Annual ranking | 15 | 50 | Random | 1.65 | 60.77 |
| Discharge | Annual ranking | 15 | 50 | ESI-first | 2.70 | 37.06 |
| Discharge | Annual ranking | 15 | 50 | Risk-only | 11.49 | 8.70 |
| Discharge | Annual ranking | 15 | 50 | Simple logistic | 8.07 | 12.38 |
| Discharge | Annual ranking | 15 | 50 | Age-first | 3.40 | 29.45 |
| Discharge | Annual ranking | 15 | 50 | Prior-visits-first | 8.20 | 12.19 |
| Discharge | Annual ranking | 15 | 100 | Random | 1.65 | 60.77 |
| Discharge | Annual ranking | 15 | 100 | ESI-first | 2.67 | 37.43 |
| Discharge | Annual ranking | 15 | 100 | Risk-only | 8.88 | 11.26 |
| Discharge | Annual ranking | 15 | 100 | Simple logistic | 6.61 | 15.12 |
| Discharge | Annual ranking | 15 | 100 | Age-first | 3.21 | 31.17 |
| Discharge | Annual ranking | 15 | 100 | Prior-visits-first | 6.80 | 14.70 |
| Admission | Annual ranking | 5 | 25 | Risk-plus-window | 13.28 | 7.53 |
| Admission | Annual ranking | 5 | 50 | Risk-plus-window | 11.56 | 8.65 |
| Admission | Annual ranking | 5 | 100 | Risk-plus-window | 9.12 | 10.97 |
| Admission | Annual ranking | 10 | 25 | Risk-plus-window | 15.57 | 6.42 |
| Admission | Annual ranking | 10 | 50 | Risk-plus-window | 12.87 | 7.77 |
| Admission | Annual ranking | 10 | 100 | Risk-plus-window | 11.27 | 8.87 |
| Admission | Annual ranking | 15 | 25 | Risk-plus-window | 17.49 | 5.72 |
| Admission | Annual ranking | 15 | 50 | Risk-plus-window | 14.15 | 7.07 |
| Admission | Annual ranking | 15 | 100 | Risk-plus-window | 11.81 | 8.47 |
| Discharge | Annual ranking | 5 | 25 | Risk-plus-window | 11.80 | 8.47 |
| Discharge | Annual ranking | 5 | 50 | Risk-plus-window | 8.92 | 11.21 |
| Discharge | Annual ranking | 5 | 100 | Risk-plus-window | 6.28 | 15.92 |
| Discharge | Annual ranking | 10 | 25 | Risk-plus-window | 14.00 | 7.14 |
| Discharge | Annual ranking | 10 | 50 | Risk-plus-window | 10.93 | 9.15 |
| Discharge | Annual ranking | 10 | 100 | Risk-plus-window | 8.28 | 12.08 |
| Discharge | Annual ranking | 15 | 25 | Risk-plus-window | 15.16 | 6.60 |
| Discharge | Annual ranking | 15 | 50 | Risk-plus-window | 12.05 | 8.30 |
| Discharge | Annual ranking | 15 | 100 | Risk-plus-window | 8.73 | 11.46 |
| Admission | Full-day ranking | 5 | 25 | Risk-plus-window | 13.52 | 7.39 |
| Admission | Full-day ranking | 5 | 50 | Risk-plus-window | 10.74 | 9.31 |
| Admission | Full-day ranking | 5 | 100 | Risk-plus-window | 9.08 | 11.02 |
| Admission | Full-day ranking | 10 | 25 | Risk-plus-window | 15.25 | 6.56 |
| Admission | Full-day ranking | 10 | 50 | Risk-plus-window | 13.36 | 7.48 |
| Admission | Full-day ranking | 10 | 100 | Risk-plus-window | 10.45 | 9.57 |
| Admission | Full-day ranking | 15 | 25 | Risk-plus-window | 16.01 | 6.25 |
| Admission | Full-day ranking | 15 | 50 | Risk-plus-window | 13.65 | 7.32 |
| Admission | Full-day ranking | 15 | 100 | Risk-plus-window | 11.50 | 8.70 |
| Discharge | Full-day ranking | 5 | 25 | Risk-plus-window | 10.93 | 9.15 |
| Discharge | Full-day ranking | 5 | 50 | Risk-plus-window | 8.57 | 11.67 |
| Discharge | Full-day ranking | 5 | 100 | Risk-plus-window | 6.29 | 15.89 |
| Discharge | Full-day ranking | 10 | 25 | Risk-plus-window | 14.42 | 6.94 |
| Discharge | Full-day ranking | 10 | 50 | Risk-plus-window | 10.48 | 9.55 |
| Discharge | Full-day ranking | 10 | 100 | Risk-plus-window | 8.03 | 12.45 |
| Discharge | Full-day ranking | 15 | 25 | Risk-plus-window | 15.53 | 6.44 |
| Discharge | Full-day ranking | 15 | 50 | Risk-plus-window | 12.05 | 8.30 |
| Discharge | Full-day ranking | 15 | 100 | Risk-plus-window | 8.73 | 11.46 |

Full-day and annual rankings; descriptive points. Reviews per event means attempts divided by events reached, not a number needed to prevent harm. Event reach is the proportion of all composite-event encounters selected with sufficient recorded review time. Review attempts without enough recorded time still count against the budget. Reach does not measure events prevented. Simple logistic indicates the five-variable logistic model. ESI indicates Emergency Severity Index.

| eTable 22. Definitions of Recorded Resident-Plan Transitions | | | |
| --- | --- | --- | --- |
| Starting marker | Next code | Recorded next-status label | Category |
| Resident admission | 106116 | Decision to Admit Dispo Selected (Attending) | Agreement |
| Resident admission | 10 | Ready for Admit | Agreement |
| Resident admission | 106107 | Admit Order Signed | Agreement |
| Resident admission | 106109 | Decision to Admit - CDU | CDU admission |
| Resident admission | 106111 | Conditional Discharge | Direction change |
| Resident admission | 6 | Ready for Discharge | Direction change |
| Resident admission | 106114 | Resident Transfer Dispo Selected | Transfer |
| Resident admission | 106115 | Transfer to LPCH | Transfer |
| Resident admission | 14 | Ready for Transfer | Transfer |
| Resident admission | 100030 | Resident Discharge Dispo Selected | Resident revised plan |
| Resident admission | 11 | In Process | Returned to in process |
| Resident discharge | 106111 | Conditional Discharge | Agreement |
| Resident discharge | 6 | Ready for Discharge | Agreement |
| Resident discharge | 106109 | Decision to Admit - CDU | CDU admission |
| Resident discharge | 10 | Ready for Admit | Direction change |
| Resident discharge | 106107 | Admit Order Signed | Direction change |
| Resident discharge | 106116 | Decision to Admit Dispo Selected (Attending) | Direction change |
| Resident discharge | 106114 | Resident Transfer Dispo Selected | Transfer |
| Resident discharge | 106115 | Transfer to LPCH | Transfer |
| Resident discharge | 14 | Ready for Transfer | Transfer |
| Resident discharge | 106108 | Resident Admit Dispo Selected | Resident revised plan |
| Resident discharge | 11 | In Process | Returned to in process |

The earliest qualifying next status at or after the resident marker is used. Repeated copies of the same starting code are excluded. Simultaneous statuses with different categories are ties. Direction change and resident-revised-plan categories remain distinct. Status names do not independently identify the clinician entering each admission marker. Recorded transitions are not adjudicated attending overrides.

| eTable 23. Next Recorded Status After Resident-Plan Markers, 2020–2023 | | |
| --- | --- | --- |
| Next-status category | Resident discharge | Resident admission |
| All resident-plan encounters | 75,077 | 25,314 |
| Agreement | 71,344 (95.03) | 21,803 (86.13) |
| Direction-change category | 210 (0.28) | 361 (1.43) |
| Resident-revised-plan category | 148 (0.20) | 617 (2.44) |
| Return to In Process | 2,651 (3.53) | 1,802 (7.12) |
| CDU admission | 0 (0.00) | 0 (0.00) |
| Transfer | 42 (0.06) | 411 (1.62) |
| Tied categories | 548 (0.73) | 280 (1.11) |
| No later listed status | 134 (0.18) | 40 (0.16) |

Values are No. (%) using all encounters with the stated resident marker, including ties and no next status. An encounter may contribute to both columns. Direction-change excludes the separate resident-revised-plan category, which contains opposite-direction resident markers. No category establishes an adjudicated attending override.

| eTable 24. Last Qualifying Status After Resident-Plan Markers, 2020–2023 | | |
| --- | --- | --- |
| Direction relative to plan | Last status group | Encounters |
| Resident discharge | | |
| In Process | In Process | 85 |
| No later status | No later status | 134 |
| Opposite direction | Direction change | 1,176 |
| Opposite direction | Opposite resident plan | — |
| Other status | Transfer | 43 |
| Same direction | Agreement | 73,132 |
| Same direction | Repeated resident plan | — |
| Same direction | Tie | — |
| Tie | Tie | — |
| Resident admission | | |
| In Process | In Process | 275 |
| No later status | No later status | 37 |
| Opposite direction | Direction change | 2,233 |
| Opposite direction | Opposite resident plan | — |
| Opposite direction | Tie | — |
| Other status | Transfer | 268 |
| Same direction | Agreement | 22,117 |
| Same direction | Repeated resident plan | — |
| Same direction | Tie | — |
| Tie | Tie | — |

Pooled encounters with a first qualifying resident-plan marker. The last qualifying status at or after the marker and by departure or CDU entry is used, excluding the anchor itself. Later repeats of the same resident code remain eligible. Direction and status group are separate: different simultaneous groups can have the same direction. Encounters may enter both resident-plan analyses. Counts of 1–10, corresponding percentages, and selected cells from which they could be calculated are not shown (—). Changes do not identify or bound attending review.

| eTable 25. Last Resident-Plan Status and Final ED Disposition, 2020–2023 | | | |
| --- | --- | --- | --- |
| Direction | Last status group | Final disposition | Encounters |
| Resident discharge | | | |
| In Process | In Process | Admit | — |
| In Process | In Process | Discharge | 63 |
| In Process | In Process | Other | — |
| In Process | In Process | Unmapped | — |
| No later status | No later status | Admit | — |
| No later status | No later status | Discharge | 83 |
| No later status | No later status | Other | — |
| No later status | No later status | Transfer | — |
| No later status | No later status | Unmapped | — |
| Opposite direction | Direction change | Admit | 845 |
| Opposite direction | Direction change | Discharge | 296 |
| Opposite direction | Direction change | Other | — |
| Opposite direction | Direction change | Transfer | — |
| Opposite direction | Direction change | Unmapped | — |
| Opposite direction | Opposite resident plan | Discharge | — |
| Other status | Transfer | Admit | — |
| Other status | Transfer | Transfer | — |
| Other status | Transfer | Unmapped | — |
| Same direction | Agreement | Admit | — |
| Same direction | Agreement | Discharge | 72,778 |
| Same direction | Agreement | Other | — |
| Same direction | Agreement | Transfer | — |
| Same direction | Agreement | Unmapped | — |
| Same direction | Repeated resident plan | Discharge | — |
| Same direction | Repeated resident plan | Other | — |
| Same direction | Tie | Discharge | — |
| Tie | Tie | Admit | — |
| Tie | Tie | Discharge | — |
| Tie | Tie | Other | — |
| Tie | Tie | Transfer | — |
| Tie | Tie | Unmapped | — |
| Resident admission | | | |
| In Process | In Process | Admit | — |
| In Process | In Process | Discharge | 203 |
| In Process | In Process | Other | — |
| In Process | In Process | Unmapped | — |
| No later status | No later status | Admit | — |
| No later status | No later status | Discharge | — |
| No later status | No later status | Error | — |
| No later status | No later status | Other | 12 |
| No later status | No later status | Transfer | — |
| No later status | No later status | Unmapped | — |
| Opposite direction | Direction change | Admit | — |
| Opposite direction | Direction change | Discharge | 2,100 |
| Opposite direction | Direction change | Other | — |
| Opposite direction | Direction change | Unmapped | — |
| Opposite direction | Opposite resident plan | Admit | — |
| Opposite direction | Opposite resident plan | Discharge | — |
| Opposite direction | Opposite resident plan | Other | — |
| Opposite direction | Tie | Discharge | — |
| Other status | Transfer | Admit | — |
| Other status | Transfer | Discharge | — |
| Other status | Transfer | Other | — |
| Other status | Transfer | Transfer | — |
| Other status | Transfer | Unmapped | — |
| Same direction | Agreement | Admit | 20,146 |
| Same direction | Agreement | Discharge | 1,751 |
| Same direction | Agreement | Other | 36 |
| Same direction | Agreement | Transfer | 11 |
| Same direction | Agreement | Unmapped | 173 |
| Same direction | Repeated resident plan | Admit | — |
| Same direction | Repeated resident plan | Other | — |
| Same direction | Tie | Admit | — |
| Same direction | Tie | Discharge | — |
| Tie | Tie | Admit | — |
| Tie | Tie | Discharge | — |
| Tie | Tie | Other | — |
| Tie | Tie | Transfer | — |
| Tie | Tie | Unmapped | — |

The pooled resident-plan populations match the last-status table. Final disposition is recorded separately and can reflect CDU care or events after handoff; it does not redefine the original plan or model decision group. Counts of 1–10, corresponding percentages, and selected cells from which they could be calculated are not shown (—). ED indicates emergency department; CDU, clinical decision unit.

| eTable 26. Recorded Status After a Return to In Process, 2020–2023 | | |
| --- | --- | --- |
| Direction relative to resident plan | Following status group | Encounters |
| Resident discharge | | |
| No later decision | No later decision | 13 |
| Opposite direction | Direction change | 293 |
| Opposite direction | Multiple groups | — |
| Opposite direction | Opposite resident plan | 65 |
| Other status | Transfer | — |
| Same direction | Agreement | 1,123 |
| Same direction | Multiple groups | 71 |
| Same direction | Repeated resident plan | 1,063 |
| Tie | Multiple groups | — |
| Resident admission | | |
| No later decision | No later decision | — |
| Opposite direction | Direction change | 242 |
| Opposite direction | Multiple groups | 23 |
| Opposite direction | Opposite resident plan | 477 |
| Other status | Transfer | — |
| Same direction | Agreement | 727 |
| Same direction | Multiple groups | 13 |
| Same direction | Repeated resident plan | 296 |
| Tie | Multiple groups | — |

Among resident-plan encounters whose next qualifying status was In Process, this table describes the first later qualifying status other than In Process by departure or CDU entry. Multiple groups at the same time can share one direction; conflicting directions form a tie. These counts are not attributable to attending review. Counts of 1–10, corresponding percentages, and selected cells from which they could be calculated are not shown (—).

| eTable 27. Empirical Coverage of Sequential Reach and Paired Contrasts in 2023 | | | | | | | | |
| --- | --- | --- | --- | --- | --- | --- | --- | --- |
| Group | Review, min | Statistic | Reference, % or pp | Coverage, % | MCSE, pp | Below reference | Above reference | Mean shift, pp |
| Admission | 10 | Risk-only | 10.07 | 39.1 | 1.54 | 609 | 0 | −0.92 |
| Admission | 10 | Prior-visits-first | 7.73 | 54.0 | 1.58 | 460 | 0 | −0.79 |
| Admission | 10 | Risk-only minus Prior-visits-first | 2.34 | 94.8 | 0.70 | 47 | 5 | −0.13 |
| Admission | 5 | Risk-only | 19.60 | 50.8 | 1.58 | 492 | 0 | −1.22 |
| Admission | 5 | Prior-visits-first | 14.21 | 75.7 | 1.36 | 243 | 0 | −0.64 |
| Admission | 5 | Risk-only minus Prior-visits-first | 5.40 | 93.1 | 0.80 | 68 | 1 | −0.57 |
| Discharge | 10 | Risk-only | 19.48 | 46.0 | 1.58 | 540 | 0 | −1.43 |
| Discharge | 10 | Prior-visits-first | 16.01 | 36.0 | 1.52 | 640 | 0 | −1.72 |
| Discharge | 10 | Risk-only minus Prior-visits-first | 3.46 | 96.0 | 0.62 | 7 | 33 | 0.28 |
| Discharge | 5 | Risk-only | 33.15 | 64.2 | 1.52 | 358 | 0 | −1.29 |
| Discharge | 5 | Prior-visits-first | 28.46 | 34.9 | 1.51 | 651 | 0 | −2.32 |
| Discharge | 5 | Risk-only minus Prior-visits-first | 4.68 | 93.2 | 0.80 | 0 | 68 | 1.03 |

Nested diagnostic with 1,000 outer samples and 2,000 inner resamples. All intervals were valid. Zero start delay; 10-minute reviews used a 5% flag rate, 5-minute reviews a 10% flag rate. Reference values are full-cohort estimates, not known population truths. Counts describe resampling intervals. Abbreviations: MCSE, Monte Carlo standard error; pp, percentage points.

| eTable 28. Empirical Coverage of Sequential Intervals in 2024 | | | | | | | | |
| --- | --- | --- | --- | --- | --- | --- | --- | --- |
| Group | Review, min | Statistic | Reference, % or pp | Coverage, % | MCSE, pp | Valid | Below reference | Above reference |
| Admission | 10 | Risk-only | 11.20 | 16.7 | 1.18 | 1,000 | 833 | 0 |
| Admission | 10 | Prior-visits-first | 7.64 | 46.2 | 1.58 | 1,000 | 538 | 0 |
| Admission | 10 | Risk-only minus Prior-visits-first | 3.56 | 85.6 | 1.11 | 1,000 | 142 | 2 |
| Admission | 5 | Risk-only | 19.11 | 69.1 | 1.46 | 1,000 | 309 | 0 |
| Admission | 5 | Prior-visits-first | 13.96 | 54.6 | 1.57 | 1,000 | 454 | 0 |
| Admission | 5 | Risk-only minus Prior-visits-first | 5.16 | 96.9 | 0.55 | 1,000 | 11 | 20 |
| Discharge | 10 | Risk-only | 19.42 | 51.1 | 1.58 | 1,000 | 489 | 0 |
| Discharge | 10 | Prior-visits-first | 15.76 | 75.9 | 1.35 | 1,000 | 241 | 0 |
| Discharge | 10 | Risk-only minus Prior-visits-first | 3.66 | 93.4 | 0.79 | 1,000 | 60 | 6 |
| Discharge | 5 | Risk-only | 32.65 | 62.9 | 1.53 | 1,000 | 371 | 0 |
| Discharge | 5 | Prior-visits-first | 26.92 | 70.9 | 1.44 | 1,000 | 291 | 0 |
| Discharge | 5 | Risk-only minus Prior-visits-first | 5.72 | 97.6 | 0.48 | 1,000 | 20 | 4 |

Nested patient-cluster diagnostic: 1,000 outer and 2,000 inner samples, with selection repeated at both levels. Zero delay; 10-minute reviews use 5% tuning flags and 5-minute reviews use 10%. Reference values are the full-2024-cohort estimates, not population truths. All 1,000 intervals per statistic were valid. Counts of intervals are simulation outputs, not patient counts. MCSE indicates Monte Carlo standard error; pp, percentage points.

| eTable 29. Empirical Coverage of Full-Day and Annual Ranking Intervals in 2023 | | | | | | | | | |
| --- | --- | --- | --- | --- | --- | --- | --- | --- | --- |
| Allocation | Decision group | Statistic | Reference % or pp | Coverage % | MCSE pp | Invalid | Below reference | Above reference | Mean shift pp |
| Daily | Admission | Random | 4.80 | 98.1 | 0.43 | 0 | 3 | 16 | 0.01 |
| Daily | Admission | ESI-first | 9.81 | 69.2 | 1.46 | 0 | 308 | 0 | −0.36 |
| Daily | Admission | Risk-only | 14.39 | 78.5 | 1.30 | 0 | 215 | 0 | −0.59 |
| Daily | Admission | Risk-only minus ESI-first | 4.58 | 95.5 | 0.66 | 0 | 40 | 5 | −0.23 |
| Daily | Discharge | Random | 4.20 | 95.1 | 0.68 | 0 | 21 | 28 | 0.00 |
| Daily | Discharge | ESI-first | 6.70 | 95.7 | 0.64 | 0 | 27 | 16 | 0.02 |
| Daily | Discharge | Risk-only | 23.69 | 95.8 | 0.63 | 0 | 39 | 3 | 0.04 |
| Daily | Discharge | Risk-only minus ESI-first | 16.99 | 95.2 | 0.68 | 0 | 46 | 2 | 0.02 |
| Annual | Admission | Random | 4.81 | 95.1 | 0.68 | 0 | 21 | 28 | −0.00 |
| Annual | Admission | ESI-first | 9.89 | 95.4 | 0.66 | 0 | 27 | 19 | 0.01 |
| Annual | Admission | Risk-only | 14.39 | 94.4 | 0.73 | 0 | 40 | 16 | −0.07 |
| Annual | Admission | Risk-only minus ESI-first | 4.50 | 94.9 | 0.70 | 0 | 37 | 14 | −0.08 |
| Annual | Discharge | Random | 4.20 | 94.6 | 0.71 | 0 | 21 | 33 | 0.00 |
| Annual | Discharge | ESI-first | 6.69 | 95.7 | 0.64 | 0 | 28 | 15 | 0.00 |
| Annual | Discharge | Risk-only | 25.00 | 95.0 | 0.69 | 0 | 26 | 24 | 0.07 |
| Annual | Discharge | Risk-only minus ESI-first | 18.31 | 95.0 | 0.69 | 0 | 26 | 24 | 0.06 |

Nested patient-cluster diagnostic: 1,000 outer samples and 2,000 inner samples. Reference values are full-cohort estimates; invalid intervals count as failures. Counts describe intervals, not patients. Empirical containment does not establish population coverage. Abbreviations: MCSE, Monte Carlo standard error; pp, percentage points.

| eTable 30. Eligible Cohorts and Event Rates by Period | | | | |
| --- | --- | --- | --- | --- |
| Period | Decision group | Encounters | Events | Event rate (%) |
| 2020–2021 | Admission | 36,417 | 1,799 | 4.94 |
| 2020–2021 | Discharge | 83,245 | 1,495 | 1.80 |
| 2022 | Admission | 22,505 | 1,041 | 4.63 |
| 2022 | Discharge | 43,366 | 907 | 2.09 |
| 2023 | Admission | 24,401 | 1,112 | 4.56 |
| 2023 | Discharge | 48,313 | 1,068 | 2.21 |
| 2024 | Admission | 26,411 | 1,125 | 4.26 |
| 2024 | Discharge | 50,675 | 1,066 | 2.10 |

The composite comprised an adult ED return arriving within 7 days of time zero and ultimately ending in inpatient admission, qualifying critical care within 24 hours, or death within 7 days. Training: 2020–2021; tuning: 2022; temporal test set: 2023; held-out temporal test set: 2024. Patients could appear across periods. Eligibility follows decision-time, known-outcome, and temporal exclusions, including time zero outside its assigned period and the 7-day development boundary.

| eTable 31. Recorded Demographic Characteristics by Period | | | | | |
| --- | --- | --- | --- | --- | --- |
| Characteristic | Category | 2020–2021 | 2022 | 2023 | 2024 |
| Sex | Female | 63,276 (52.9) | 36,066 (54.8) | 40,252 (55.4) | 42,333 (54.9) |
| Sex | Male, non-binary or unknown sex | 56,386 (47.1) | 29,805 (45.2) | 32,462 (44.6) | 34,753 (45.1) |
| Sex | Missing | 0 (0.0) | 0 (0.0) | 0 (0.0) | 0 (0.0) |
| Race | American Indian or Alaska Native | 287 (0.2) | 164 (0.2) | 181 (0.2) | 224 (0.3) |
| Race | Asian | 17,151 (14.3) | 10,760 (16.3) | 12,317 (16.9) | 13,040 (16.9) |
| Race | Black or African American | 7,518 (6.3) | 4,112 (6.2) | 4,484 (6.2) | 4,634 (6.0) |
| Race | Multiple recorded labels | 3,305 (2.8) | 2,021 (3.1) | 2,201 (3.0) | 2,206 (2.9) |
| Race | Native Hawaiian or Other Pacific Islander | 2,612 (2.2) | 1,328 (2.0) | 1,479 (2.0) | 1,496 (1.9) |
| Race | Other | 40,986 (34.3) | 21,096 (32.0) | 22,877 (31.5) | 23,961 (31.1) |
| Race | White | 46,471 (38.8) | 25,550 (38.8) | 27,904 (38.4) | 29,209 (37.9) |
| Race | Declines to state | 1,206 (1.0) | 740 (1.1) | 1,109 (1.5) | 2,094 (2.7) |
| Race | Missing | 49 (0.0) | 48 (0.1) | 118 (0.2) | 148 (0.2) |
| Race | Unknown | — | — | — | — |
| Ethnicity | Hispanic/Latino | 34,847 (29.1) | 19,126 (29.0) | 21,762 (29.9) | 23,318 (30.2) |
| Ethnicity | Non-Hispanic/non-Latino | 83,682 (69.9) | 46,078 (70.0) | 49,962 (68.7) | 52,032 (67.5) |
| Ethnicity | Declines to state | 848 (0.7) | 510 (0.8) | 770 (1.1) | 1,418 (1.8) |
| Ethnicity | Missing | 45 (0.0) | 27 (0.0) | 110 (0.2) | 143 (0.2) |
| Ethnicity | Unknown | 240 (0.2) | 130 (0.2) | 110 (0.2) | 175 (0.2) |
| Language | English | 97,755 (81.7) | 52,914 (80.3) | 57,088 (78.5) | 60,270 (78.2) |
| Language | Farsi | 539 (0.5) | 256 (0.4) | 306 (0.4) | — |
| Language | Mandarin | 1,429 (1.2) | 990 (1.5) | 1,216 (1.7) | 1,213 (1.6) |
| Language | Spanish | 15,312 (12.8) | 8,623 (13.1) | 10,317 (14.2) | 11,248 (14.6) |
| Language | Vietnamese | 1,118 (0.9) | 851 (1.3) | 1,093 (1.5) | 1,072 (1.4) |
| Language | Other or missing | 3,509 (2.9) | 2,237 (3.4) | 2,694 (3.7) | 2,911 (3.8) |
| Language | Unknown | 0 (0.0) | 0 (0.0) | 0 (0.0) | — |
| Interpreter need | Not required | 100,112 (83.7) | 54,331 (82.5) | 58,885 (81.0) | 62,127 (80.6) |
| Interpreter need | Required | 19,535 (16.3) | 11,521 (17.5) | 13,757 (18.9) | 14,842 (19.3) |
| Interpreter need | Missing | 15 (0.0) | 19 (0.0) | 72 (0.1) | 117 (0.2) |

Values are No. (%) and correspond to Table 1. Recorded categories and the same pooling and withholding are retained. Counts of 1–10, corresponding percentages, and selected cells from which they could be calculated are not shown (—).

| eTable 32. Clinical Characteristics by Decision Group in 2023 | | |
| --- | --- | --- |
| Characteristic | 2023 Admission | 2023 Discharge |
| Cohort |  |  |
| Encounters | 24,401 | 48,313 |
| Unique patients | 17,364 | 35,896 |
| Age, y |  |  |
| Median (IQR) | 66 (49–78) | 45 (31–63) |
| Available | 24,401 (100.0) | 48,313 (100.0) |
| Missing | 0 (0.0) | 0 (0.0) |
| ESI |  |  |
| 1 | 364 (1.5) | 43 (0.1) |
| 2 | 10,557 (43.3) | 7,789 (16.1) |
| 3 | 13,120 (53.8) | 31,640 (65.5) |
| 4 | 255 (1.0) | 7,995 (16.5) |
| 5 | — | — |
| Missing | — | — |
| Arrival mode |  |  |
| Car/bus | 16,874 (69.2) | 40,998 (84.9) |
| Walk | 245 (1.0) | 583 (1.2) |
| Ambulance, American Medical Response | 4,770 (19.5) | 4,663 (9.7) |
| Ambulance, Palo Alto Paramedics | 1,064 (4.4) | 970 (2.0) |
| Ambulance | 472 (1.9) | 430 (0.9) |
| Other or missing | 976 (4.0) | 669 (1.4) |

Values are No. (%) or median (IQR). Age is recorded age without imputation. ESI categories are levels 1–5 and Missing. No separate invalid category is displayed. Arrival modes are recorded electronic health record categories; ambulance categories are shown separately as recorded. To protect patient privacy, counts of 1–10, and selected cells from which they could be calculated, are not shown (—).

| eTable 33. Clinical Characteristics by Decision Group in 2024 | | | |
| --- | --- | --- | --- |
| Characteristic | Category | 2024 Admission | 2024 Discharge |
| Cohort | Encounters | 26,411 | 50,675 |
| Cohort | Unique patients | 18,773 | 37,780 |
| Age, y | Median (IQR) | 65.0 (49.0–78.0) | 45.0 (31.0–64.0) |
| Age, y | Available | 26,411 (100.0) | 50,675 (100.0) |
| Age, y | Missing | 0 (0.0) | 0 (0.0) |
| ESI | 1 | 355 (1.3) | 41 (0.1) |
| ESI | 2 | 12,357 (46.8) | 8,659 (17.1) |
| ESI | 3 | 13,314 (50.4) | 33,364 (65.8) |
| ESI | 4 | 259 (1.0) | 7,851 (15.5) |
| ESI | 5 | — | — |
| ESI | Missing | — | — |
| Arrival mode | Ambulance, American Medical Response | 5,368 (20.3) | 5,301 (10.5) |
| Arrival mode | Ambulance, Palo Alto Paramedics | 957 (3.6) | 873 (1.7) |
| Arrival mode | Ambulance | 853 (3.2) | 643 (1.3) |
| Arrival mode | Car/bus | 18,130 (68.6) | 42,712 (84.3) |
| Arrival mode | Walk | 372 (1.4) | 607 (1.2) |
| Arrival mode | Other or missing | 731 (2.8) | 539 (1.1) |

Eligible 2024 encounters. Values are No. (%) unless stated otherwise. The decision groups partition encounters; patients may appear in both. Denominators include missing records. Age uses available values and linear-interpolated quartiles. Categories match Table 1. Counts of 1–10, corresponding percentages, and selected cells from which they could be calculated are not shown (—). IQR indicates interquartile range; ESI, Emergency Severity Index.

| eTable 34. Uncapped Recorded Decision, Handoff, and Departure Intervals in 2020–2023 | | | | |
| --- | --- | --- | --- | --- |
| Period | Interval | Encounters | Median (IQR), min | Mean, min |
| 2020 | Handoff to ED departure | 16,788 | 158.0 (100.0–305.0) | 324.9 |
| 2020 | Admission plan to ED departure | 16,788 | 307.0 (207.0–489.0) | 459.2 |
| 2020 | Admission plan to handoff | 16,788 | 103.0 (50.0–181.0) | 134.3 |
| 2020 | Admission plan to departure/CDU fallback | 230 | 293.0 (173.5–579.0) | 581.6 |
| 2020 | Discharge decision to ED departure | 42,647 | 20.0 (9.0–39.0) | 40.4 |
| 2020 | Discharge decision to window end | 42,647 | 20.0 (9.0–39.0) | 34.9 |
| 2021 | Handoff to ED departure | 19,772 | 157.0 (99.0–302.0) | 332.9 |
| 2021 | Admission plan to ED departure | 19,772 | 306.0 (205.0–491.0) | 469.5 |
| 2021 | Admission plan to handoff | 19,772 | 102.0 (49.0–182.0) | 136.6 |
| 2021 | Admission plan to departure/CDU fallback | 239 | 351.0 (179.0–883.5) | 711.0 |
| 2021 | Discharge decision to ED departure | 42,908 | 23.0 (13.0–43.0) | 46.9 |
| 2021 | Discharge decision to window end | 42,908 | 23.0 (13.0–43.0) | 40.5 |
| 2022 | Handoff to ED departure | 22,813 | 161.0 (95.0–360.0) | 354.7 |
| 2022 | Admission plan to ED departure | 22,813 | 318.0 (207.0–548.0) | 497.2 |
| 2022 | Admission plan to handoff | 22,813 | 106.0 (49.0–186.0) | 142.5 |
| 2022 | Admission plan to departure/CDU fallback | 304 | 387.5 (199.0–872.0) | 784.9 |
| 2022 | Discharge decision to ED departure | 45,371 | 23.0 (13.0–42.0) | 47.6 |
| 2022 | Discharge decision to window end | 45,371 | 23.0 (13.0–42.0) | 41.4 |
| 2023 | Handoff to ED departure | 24,221 | 163.0 (89.0–421.0) | 364.8 |
| 2023 | Admission plan to ED departure | 24,221 | 331.0 (204.0–602.0) | 507.7 |
| 2023 | Admission plan to handoff | 24,221 | 105.0 (48.0–192.0) | 142.9 |
| 2023 | Admission plan to departure/CDU fallback | 312 | 426.5 (220.8–1111.0) | 878.5 |
| 2023 | Discharge decision to ED departure | 48,872 | 22.0 (13.0–40.0) | 47.5 |
| 2023 | Discharge decision to window end | 48,872 | 22.0 (13.0–39.0) | 41.2 |
| All | Handoff to ED departure | 83,594 | 160.0 (95.0–347.0) | 346.5 |
| All | Admission plan to ED departure | 83,594 | 316.0 (206.0–535.0) | 486.1 |
| All | Admission plan to handoff | 83,594 | 104.0 (49.0–186.0) | 139.6 |
| All | Admission plan to departure/CDU fallback | 1,085 | 365.0 (194.0–889.0) | 752.4 |
| All | Discharge decision to ED departure | 179,798 | 22.0 (12.0–41.0) | 45.7 |
| All | Discharge decision to window end | 179,798 | 22.0 (12.0–41.0) | 39.6 |

Populations by decision-time group before outcome and model exclusions. Admission-plan-to-handoff and handoff-to-departure intervals include only recorded-handoff encounters. Handoff-to-departure is not part of the review window. Fallback windows do not establish observed handoff time. Quantiles require at least 11 contributing encounters. The signed admission-order timestamp may approximate accepting-team discussion in local workflow. This interpretation does not automatically apply to readiness-for-admission or fallback endpoints; actual discussion and responsibility-transfer times were unmeasured.

| eTable 35. Sequential Event-Reach Contrasts in 2023 | | | | | | |
| --- | --- | --- | --- | --- | --- | --- |
| Group | Review, min | Flag rate, % | Delay, min | Contrast | Difference, pp | Nominal 95% interval, pp |
| Admission | 10 | 5 | 0 | Risk-only minus Random | 7.01 | Not reported |
| Admission | 10 | 5 | 0 | Risk-only minus ESI-first | 2.79 | Not reported |
| Admission | 10 | 5 | 0 | Risk-only minus Age-first | 6.92 | Not reported |
| Admission | 10 | 5 | 0 | Risk-only minus Prior-visits-first | 2.34 | 0.19 to 4.20 |
| Admission | 10 | 5 | 0 | Risk-only minus Simple logistic | −1.17 | Not reported |
| Admission | 10 | 5 | 0 | Risk-plus-window minus Risk-only | 0.27 | Not reported |
| Admission | 10 | 5 | 5 | Risk-only minus Random | 6.38 | Not reported |
| Admission | 10 | 5 | 5 | Risk-only minus ESI-first | 2.16 | Not reported |
| Admission | 10 | 5 | 5 | Risk-only minus Age-first | 6.29 | Not reported |
| Admission | 10 | 5 | 5 | Risk-only minus Prior-visits-first | 2.07 | Not reported |
| Admission | 10 | 5 | 5 | Risk-only minus Simple logistic | −1.62 | Not reported |
| Admission | 10 | 5 | 5 | Risk-plus-window minus Risk-only | 0.36 | Not reported |
| Admission | 10 | 5 | 10 | Risk-only minus Random | 6.29 | Not reported |
| Admission | 10 | 5 | 10 | Risk-only minus ESI-first | 2.16 | Not reported |
| Admission | 10 | 5 | 10 | Risk-only minus Age-first | 6.38 | Not reported |
| Admission | 10 | 5 | 10 | Risk-only minus Prior-visits-first | 1.98 | Not reported |
| Admission | 10 | 5 | 10 | Risk-only minus Simple logistic | −1.71 | Not reported |
| Admission | 10 | 5 | 10 | Risk-plus-window minus Risk-only | Not estimated | Not reported |
| Admission | 10 | 10 | 0 | Risk-only minus Random | 7.87 | Not reported |
| Admission | 10 | 10 | 0 | Risk-only minus ESI-first | 5.17 | Not reported |
| Admission | 10 | 10 | 0 | Risk-only minus Age-first | 6.61 | Not reported |
| Admission | 10 | 10 | 0 | Risk-only minus Prior-visits-first | 3.55 | Not reported |
| Admission | 10 | 10 | 0 | Risk-only minus Simple logistic | 1.53 | Not reported |
| Admission | 10 | 10 | 0 | Risk-plus-window minus Risk-only | −0.18 | Not reported |
| Admission | 10 | 10 | 5 | Risk-only minus Random | 7.33 | Not reported |
| Admission | 10 | 10 | 5 | Risk-only minus ESI-first | 4.72 | Not reported |
| Admission | 10 | 10 | 5 | Risk-only minus Age-first | 6.07 | Not reported |
| Admission | 10 | 10 | 5 | Risk-only minus Prior-visits-first | 3.37 | Not reported |
| Admission | 10 | 10 | 5 | Risk-only minus Simple logistic | 1.26 | Not reported |
| Admission | 10 | 10 | 5 | Risk-plus-window minus Risk-only | −0.63 | Not reported |
| Admission | 10 | 10 | 10 | Risk-only minus Random | 7.33 | Not reported |
| Admission | 10 | 10 | 10 | Risk-only minus ESI-first | 4.63 | Not reported |
| Admission | 10 | 10 | 10 | Risk-only minus Age-first | 6.16 | Not reported |
| Admission | 10 | 10 | 10 | Risk-only minus Prior-visits-first | 3.37 | Not reported |
| Admission | 10 | 10 | 10 | Risk-only minus Simple logistic | 1.26 | Not reported |
| Admission | 10 | 10 | 10 | Risk-plus-window minus Risk-only | Not estimated | Not reported |
| Admission | 5 | 10 | 0 | Risk-only minus Random | 12.19 | Not reported |
| Admission | 5 | 10 | 0 | Risk-only minus ESI-first | 6.70 | Not reported |
| Admission | 5 | 10 | 0 | Risk-only minus Age-first | 10.21 | Not reported |
| Admission | 5 | 10 | 0 | Risk-only minus Prior-visits-first | 5.40 | 2.12 to 7.36 |
| Admission | 5 | 10 | 0 | Risk-only minus Simple logistic | 1.30 | Not reported |
| Admission | 5 | 10 | 0 | Risk-plus-window minus Risk-only | −0.09 | Not reported |
| Admission | 5 | 10 | 5 | Risk-only minus Random | 11.74 | Not reported |
| Admission | 5 | 10 | 5 | Risk-only minus ESI-first | 6.43 | Not reported |
| Admission | 5 | 10 | 5 | Risk-only minus Age-first | 10.03 | Not reported |
| Admission | 5 | 10 | 5 | Risk-only minus Prior-visits-first | 5.22 | Not reported |
| Admission | 5 | 10 | 5 | Risk-only minus Simple logistic | 1.21 | Not reported |
| Admission | 5 | 10 | 5 | Risk-plus-window minus Risk-only | 0.00 | Not reported |
| Admission | 5 | 10 | 10 | Risk-only minus Random | 11.20 | Not reported |
| Admission | 5 | 10 | 10 | Risk-only minus ESI-first | 5.89 | Not reported |
| Admission | 5 | 10 | 10 | Risk-only minus Age-first | 9.40 | Not reported |
| Admission | 5 | 10 | 10 | Risk-only minus Prior-visits-first | 5.04 | Not reported |
| Admission | 5 | 10 | 10 | Risk-only minus Simple logistic | 0.94 | Not reported |
| Admission | 5 | 10 | 10 | Risk-plus-window minus Risk-only | −0.09 | Not reported |
| Discharge | 10 | 5 | 0 | Risk-only minus Random | 16.76 | Not reported |
| Discharge | 10 | 5 | 0 | Risk-only minus ESI-first | 14.42 | Not reported |
| Discharge | 10 | 5 | 0 | Risk-only minus Age-first | 12.17 | Not reported |
| Discharge | 10 | 5 | 0 | Risk-only minus Prior-visits-first | 3.46 | 1.08 to 6.51 |
| Discharge | 10 | 5 | 0 | Risk-only minus Simple logistic | 4.49 | Not reported |
| Discharge | 10 | 5 | 0 | Risk-plus-window minus Risk-only | 0.00 | Not reported |
| Discharge | 10 | 5 | 5 | Risk-only minus Random | 15.54 | Not reported |
| Discharge | 10 | 5 | 5 | Risk-only minus ESI-first | 13.11 | Not reported |
| Discharge | 10 | 5 | 5 | Risk-only minus Age-first | 11.05 | Not reported |
| Discharge | 10 | 5 | 5 | Risk-only minus Prior-visits-first | 4.03 | Not reported |
| Discharge | 10 | 5 | 5 | Risk-only minus Simple logistic | 4.12 | Not reported |
| Discharge | 10 | 5 | 5 | Risk-plus-window minus Risk-only | −0.19 | Not reported |
| Discharge | 10 | 5 | 10 | Risk-only minus Random | 13.95 | Not reported |
| Discharge | 10 | 5 | 10 | Risk-only minus ESI-first | 11.70 | Not reported |
| Discharge | 10 | 5 | 10 | Risk-only minus Age-first | 9.74 | Not reported |
| Discharge | 10 | 5 | 10 | Risk-only minus Prior-visits-first | 3.18 | Not reported |
| Discharge | 10 | 5 | 10 | Risk-only minus Simple logistic | 3.28 | Not reported |
| Discharge | 10 | 5 | 10 | Risk-plus-window minus Risk-only | Not estimated | Not reported |
| Discharge | 10 | 10 | 0 | Risk-only minus Random | 14.37 | Not reported |
| Discharge | 10 | 10 | 0 | Risk-only minus ESI-first | 11.75 | Not reported |
| Discharge | 10 | 10 | 0 | Risk-only minus Age-first | 12.03 | Not reported |
| Discharge | 10 | 10 | 0 | Risk-only minus Prior-visits-first | 4.92 | Not reported |
| Discharge | 10 | 10 | 0 | Risk-only minus Simple logistic | 6.09 | Not reported |
| Discharge | 10 | 10 | 0 | Risk-plus-window minus Risk-only | −0.09 | Not reported |
| Discharge | 10 | 10 | 5 | Risk-only minus Random | 13.16 | Not reported |
| Discharge | 10 | 10 | 5 | Risk-only minus ESI-first | 11.00 | Not reported |
| Discharge | 10 | 10 | 5 | Risk-only minus Age-first | 11.00 | Not reported |
| Discharge | 10 | 10 | 5 | Risk-only minus Prior-visits-first | 4.73 | Not reported |
| Discharge | 10 | 10 | 5 | Risk-only minus Simple logistic | 6.04 | Not reported |
| Discharge | 10 | 10 | 5 | Risk-plus-window minus Risk-only | 0.23 | Not reported |
| Discharge | 10 | 10 | 10 | Risk-only minus Random | 11.66 | Not reported |
| Discharge | 10 | 10 | 10 | Risk-only minus ESI-first | 9.78 | Not reported |
| Discharge | 10 | 10 | 10 | Risk-only minus Age-first | 9.50 | Not reported |
| Discharge | 10 | 10 | 10 | Risk-only minus Prior-visits-first | 4.07 | Not reported |
| Discharge | 10 | 10 | 10 | Risk-only minus Simple logistic | 4.63 | Not reported |
| Discharge | 10 | 10 | 10 | Risk-plus-window minus Risk-only | Not estimated | Not reported |
| Discharge | 5 | 10 | 0 | Risk-only minus Random | 25.28 | Not reported |
| Discharge | 5 | 10 | 0 | Risk-only minus ESI-first | 20.51 | Not reported |
| Discharge | 5 | 10 | 0 | Risk-only minus Age-first | 20.51 | Not reported |
| Discharge | 5 | 10 | 0 | Risk-only minus Prior-visits-first | 4.68 | 2.52 to 8.96 |
| Discharge | 5 | 10 | 0 | Risk-only minus Simple logistic | 7.12 | Not reported |
| Discharge | 5 | 10 | 0 | Risk-plus-window minus Risk-only | 0.28 | Not reported |
| Discharge | 5 | 10 | 5 | Risk-only minus Random | 23.69 | Not reported |
| Discharge | 5 | 10 | 5 | Risk-only minus ESI-first | 19.38 | Not reported |
| Discharge | 5 | 10 | 5 | Risk-only minus Age-first | 19.01 | Not reported |
| Discharge | 5 | 10 | 5 | Risk-only minus Prior-visits-first | 3.93 | Not reported |
| Discharge | 5 | 10 | 5 | Risk-only minus Simple logistic | 6.37 | Not reported |
| Discharge | 5 | 10 | 5 | Risk-plus-window minus Risk-only | −0.09 | Not reported |
| Discharge | 5 | 10 | 10 | Risk-only minus Random | 21.82 | Not reported |
| Discharge | 5 | 10 | 10 | Risk-only minus ESI-first | 17.98 | Not reported |
| Discharge | 5 | 10 | 10 | Risk-only minus Age-first | 17.13 | Not reported |
| Discharge | 5 | 10 | 10 | Risk-only minus Prior-visits-first | 4.40 | Not reported |
| Discharge | 5 | 10 | 10 | Risk-only minus Simple logistic | 6.18 | Not reported |
| Discharge | 5 | 10 | 10 | Risk-plus-window minus Risk-only | −0.37 | Not reported |

Differences are paired points. Exploratory nominal intervals are shown only for zero-delay Risk-only minus Prior-visits-first in the two scenarios examined by the coverage diagnostic. All other contrasts are descriptive. These are not confirmatory significance tests. Abbreviation: pp, percentage points.

| eTable 36. Sequential Event-Reach Contrasts in 2024 | | | | |
| --- | --- | --- | --- | --- |
| Review, min | Tuning flags, % | Delay, min | Contrast | Difference, percentage points (nominal 95% CI where shown) |
| Discharge | | | | |
| 10 | 5 | 0 | Risk-only minus Random | 16.23 |
| 10 | 5 | 0 | Risk-only minus ESI-first | 13.56 |
| 10 | 5 | 0 | Risk-only minus Age-first | 13.13 |
| 10 | 5 | 0 | Risk-only minus Prior-visits-first | 3.66 (−0.18 to 6.26) |
| 10 | 5 | 0 | Risk-only minus Simple logistic | 7.50 |
| 10 | 5 | 0 | Risk-plus-window minus Risk-only | 0.09 |
| 10 | 5 | 5 | Risk-only minus Random | 15.57 |
| 10 | 5 | 5 | Risk-only minus ESI-first | 13.09 |
| 10 | 5 | 5 | Risk-only minus Age-first | 12.57 |
| 10 | 5 | 5 | Risk-only minus Prior-visits-first | 3.85 |
| 10 | 5 | 5 | Risk-only minus Simple logistic | 7.32 |
| 10 | 5 | 5 | Risk-plus-window minus Risk-only | 0.47 |
| 10 | 5 | 10 | Risk-only minus Random | 14.73 |
| 10 | 5 | 10 | Risk-only minus ESI-first | 12.24 |
| 10 | 5 | 10 | Risk-only minus Age-first | 11.73 |
| 10 | 5 | 10 | Risk-only minus Prior-visits-first | 4.03 |
| 10 | 5 | 10 | Risk-only minus Simple logistic | 7.04 |
| 10 | 5 | 10 | Risk-plus-window minus Risk-only | NE |
| 10 | 10 | 0 | Risk-only minus Random | 12.01 |
| 10 | 10 | 0 | Risk-only minus ESI-first | 10.13 |
| 10 | 10 | 0 | Risk-only minus Age-first | 8.82 |
| 10 | 10 | 0 | Risk-only minus Prior-visits-first | 2.39 |
| 10 | 10 | 0 | Risk-only minus Simple logistic | 5.02 |
| 10 | 10 | 0 | Risk-plus-window minus Risk-only | −0.28 |
| 10 | 10 | 5 | Risk-only minus Random | 11.35 |
| 10 | 10 | 5 | Risk-only minus ESI-first | 9.57 |
| 10 | 10 | 5 | Risk-only minus Age-first | 8.35 |
| 10 | 10 | 5 | Risk-only minus Prior-visits-first | 2.30 |
| 10 | 10 | 5 | Risk-only minus Simple logistic | 5.11 |
| 10 | 10 | 5 | Risk-plus-window minus Risk-only | 0.00 |
| 10 | 10 | 10 | Risk-only minus Random | 10.51 |
| 10 | 10 | 10 | Risk-only minus ESI-first | 8.72 |
| 10 | 10 | 10 | Risk-only minus Age-first | 7.88 |
| 10 | 10 | 10 | Risk-only minus Prior-visits-first | 2.49 |
| 10 | 10 | 10 | Risk-only minus Simple logistic | 4.83 |
| 10 | 10 | 10 | Risk-plus-window minus Risk-only | NE |
| 5 | 10 | 0 | Risk-only minus Random | 25.52 |
| 5 | 10 | 0 | Risk-only minus ESI-first | 21.48 |
| 5 | 10 | 0 | Risk-only minus Age-first | 20.26 |
| 5 | 10 | 0 | Risk-only minus Prior-visits-first | 5.72 (2.10 to 8.94) |
| 5 | 10 | 0 | Risk-only minus Simple logistic | 11.82 |
| 5 | 10 | 0 | Risk-plus-window minus Risk-only | −0.56 |
| 5 | 10 | 5 | Risk-only minus Random | 24.86 |
| 5 | 10 | 5 | Risk-only minus ESI-first | 20.64 |
| 5 | 10 | 5 | Risk-only minus Age-first | 19.61 |
| 5 | 10 | 5 | Risk-only minus Prior-visits-first | 6.10 |
| 5 | 10 | 5 | Risk-only minus Simple logistic | 11.16 |
| 5 | 10 | 5 | Risk-plus-window minus Risk-only | −0.47 |
| 5 | 10 | 10 | Risk-only minus Random | 23.08 |
| 5 | 10 | 10 | Risk-only minus ESI-first | 19.23 |
| 5 | 10 | 10 | Risk-only minus Age-first | 18.01 |
| 5 | 10 | 10 | Risk-only minus Prior-visits-first | 5.91 |
| 5 | 10 | 10 | Risk-only minus Simple logistic | 10.51 |
| 5 | 10 | 10 | Risk-plus-window minus Risk-only | −0.94 |
| Admission | | | | |
| 10 | 5 | 0 | Risk-only minus Random | 7.60 |
| 10 | 5 | 0 | Risk-only minus ESI-first | 4.98 |
| 10 | 5 | 0 | Risk-only minus Age-first | 6.40 |
| 10 | 5 | 0 | Risk-only minus Prior-visits-first | 3.56 (1.14 to 4.94) |
| 10 | 5 | 0 | Risk-only minus Simple logistic | 0.62 |
| 10 | 5 | 0 | Risk-plus-window minus Risk-only | −0.27 |
| 10 | 5 | 5 | Risk-only minus Random | 7.78 |
| 10 | 5 | 5 | Risk-only minus ESI-first | 5.07 |
| 10 | 5 | 5 | Risk-only minus Age-first | 6.49 |
| 10 | 5 | 5 | Risk-only minus Prior-visits-first | 3.91 |
| 10 | 5 | 5 | Risk-only minus Simple logistic | 0.71 |
| 10 | 5 | 5 | Risk-plus-window minus Risk-only | −0.27 |
| 10 | 5 | 10 | Risk-only minus Random | 7.78 |
| 10 | 5 | 10 | Risk-only minus ESI-first | 5.07 |
| 10 | 5 | 10 | Risk-only minus Age-first | 6.76 |
| 10 | 5 | 10 | Risk-only minus Prior-visits-first | 4.18 |
| 10 | 5 | 10 | Risk-only minus Simple logistic | 0.80 |
| 10 | 5 | 10 | Risk-plus-window minus Risk-only | NE |
| 10 | 10 | 0 | Risk-only minus Random | 7.07 |
| 10 | 10 | 0 | Risk-only minus ESI-first | 5.02 |
| 10 | 10 | 0 | Risk-only minus Age-first | 6.09 |
| 10 | 10 | 0 | Risk-only minus Prior-visits-first | 4.13 |
| 10 | 10 | 0 | Risk-only minus Simple logistic | 1.56 |
| 10 | 10 | 0 | Risk-plus-window minus Risk-only | 0.00 |
| 10 | 10 | 5 | Risk-only minus Random | 7.07 |
| 10 | 10 | 5 | Risk-only minus ESI-first | 4.93 |
| 10 | 10 | 5 | Risk-only minus Age-first | 6.00 |
| 10 | 10 | 5 | Risk-only minus Prior-visits-first | 4.22 |
| 10 | 10 | 5 | Risk-only minus Simple logistic | 1.64 |
| 10 | 10 | 5 | Risk-plus-window minus Risk-only | 0.00 |
| 10 | 10 | 10 | Risk-only minus Random | 7.07 |
| 10 | 10 | 10 | Risk-only minus ESI-first | 5.11 |
| 10 | 10 | 10 | Risk-only minus Age-first | 6.27 |
| 10 | 10 | 10 | Risk-only minus Prior-visits-first | 4.58 |
| 10 | 10 | 10 | Risk-only minus Simple logistic | 1.82 |
| 10 | 10 | 10 | Risk-plus-window minus Risk-only | NE |
| 5 | 10 | 0 | Risk-only minus Random | 10.22 |
| 5 | 10 | 0 | Risk-only minus ESI-first | 7.20 |
| 5 | 10 | 0 | Risk-only minus Age-first | 9.69 |
| 5 | 10 | 0 | Risk-only minus Prior-visits-first | 5.16 (3.06 to 8.18) |
| 5 | 10 | 0 | Risk-only minus Simple logistic | 0.80 |
| 5 | 10 | 0 | Risk-plus-window minus Risk-only | 0.09 |
| 5 | 10 | 5 | Risk-only minus Random | 10.22 |
| 5 | 10 | 5 | Risk-only minus ESI-first | 7.47 |
| 5 | 10 | 5 | Risk-only minus Age-first | 9.60 |
| 5 | 10 | 5 | Risk-only minus Prior-visits-first | 5.24 |
| 5 | 10 | 5 | Risk-only minus Simple logistic | 0.98 |
| 5 | 10 | 5 | Risk-plus-window minus Risk-only | 0.36 |
| 5 | 10 | 10 | Risk-only minus Random | 10.31 |
| 5 | 10 | 10 | Risk-only minus ESI-first | 7.47 |
| 5 | 10 | 10 | Risk-only minus Age-first | 9.60 |
| 5 | 10 | 10 | Risk-only minus Prior-visits-first | 5.42 |
| 5 | 10 | 10 | Risk-only minus Simple logistic | 1.16 |
| 5 | 10 | 10 | Risk-plus-window minus Risk-only | 0.71 |

Nominal 95% confidence intervals are shown only for zero-delay Risk-only minus Prior-visits-first contrasts with 10-minute reviews and 5% tuning flags or 5-minute reviews and 10% tuning flags, for both decision groups. Intervals use 2,000 paired patient-cluster resamples, with models fixed and selection repeated. The primary 10-minute discharge contrast did not meet the replication criterion; empirical interval coverage was 93.4% for discharge and 85.6% for admission. All other contrasts are descriptive point estimates. NE indicates not estimated; CI, confidence interval.

| eTable 37. Prediction Sensitivity Analyses in 2023 | | | | |
| --- | --- | --- | --- | --- |
| Decision group | Model | Population | Measure | Estimate (95% CI) |
| Admission | Without laboratory values | All encounters | AUROC | 0.635 (0.616 to 0.654) |
| Admission | Without laboratory values | All encounters | Average precision | 0.087 (0.076 to 0.103) |
| Admission | Without laboratory values | All encounters | Brier score | 0.043 (0.040 to 0.045) |
| Admission | Without laboratory values | All encounters | Calibration slope | 0.949 (0.830 to 1.070) |
| Admission | Without triage fields | All encounters | AUROC | 0.655 (0.638 to 0.674) |
| Admission | Without triage fields | All encounters | Average precision | 0.087 (0.077 to 0.100) |
| Admission | Without triage fields | All encounters | Brier score | 0.043 (0.040 to 0.045) |
| Admission | Without triage fields | All encounters | Calibration slope | 1.101 (0.978 to 1.235) |
| Admission | Full model | All encounters | AUROC | 0.659 (0.641 to 0.678) |
| Admission | Full model | All encounters | Average precision | 0.097 (0.085 to 0.113) |
| Admission | Full model | All encounters | Brier score | 0.043 (0.040 to 0.045) |
| Admission | Full model | All encounters | Calibration slope | 1.012 (0.901 to 1.128) |
| Admission | Full model | Recorded hospital level | AUROC | 0.628 (0.610 to 0.648) |
| Admission | Full model | Recorded hospital level | Average precision | 0.108 (0.096 to 0.125) |
| Admission | Full model | Recorded hospital level | Brier score | 0.054 (0.051 to 0.058) |
| Admission | Full model | Recorded hospital level | Calibration slope | 0.834 (0.719 to 0.958) |
| Admission | Full model | New patients | AUROC | 0.662 (0.637 to 0.689) |
| Admission | Full model | New patients | Average precision | 0.105 (0.088 to 0.129) |
| Admission | Full model | New patients | Brier score | 0.040 (0.036 to 0.043) |
| Admission | Full model | New patients | Calibration slope | 1.093 (0.931 to 1.269) |
| Admission | Training prevalence | All encounters | AUROC | 0.500 (0.500 to 0.500) |
| Admission | Training prevalence | All encounters | Average precision | 0.046 (0.043 to 0.049) |
| Admission | Training prevalence | All encounters | Brier score | 0.044 (0.041 to 0.046) |
| Discharge | Without laboratory values | All encounters | AUROC | 0.759 (0.743 to 0.775) |
| Discharge | Without laboratory values | All encounters | Average precision | 0.073 (0.063 to 0.085) |
| Discharge | Without laboratory values | All encounters | Brier score | 0.021 (0.020 to 0.023) |
| Discharge | Without laboratory values | All encounters | Calibration slope | 1.086 (1.008 to 1.160) |
| Discharge | Without triage fields | All encounters | AUROC | 0.776 (0.759 to 0.792) |
| Discharge | Without triage fields | All encounters | Average precision | 0.092 (0.080 to 0.107) |
| Discharge | Without triage fields | All encounters | Brier score | 0.021 (0.019 to 0.022) |
| Discharge | Without triage fields | All encounters | Calibration slope | 1.039 (0.975 to 1.103) |
| Discharge | Full model | All encounters | AUROC | 0.788 (0.773 to 0.802) |
| Discharge | Full model | All encounters | Average precision | 0.093 (0.081 to 0.108) |
| Discharge | Full model | All encounters | Brier score | 0.021 (0.019 to 0.022) |
| Discharge | Full model | All encounters | Calibration slope | 0.994 (0.934 to 1.052) |
| Discharge | Full model | New patients | AUROC | 0.789 (0.765 to 0.812) |
| Discharge | Full model | New patients | Average precision | 0.073 (0.059 to 0.093) |
| Discharge | Full model | New patients | Brier score | 0.014 (0.013 to 0.016) |
| Discharge | Full model | New patients | Calibration slope | 1.042 (0.955 to 1.134) |
| Discharge | Training prevalence | All encounters | AUROC | 0.500 (0.500 to 0.500) |
| Discharge | Training prevalence | All encounters | Average precision | 0.022 (0.020 to 0.024) |
| Discharge | Training prevalence | All encounters | Brier score | 0.022 (0.020 to 0.023) |

Parentheses contain 95% patient-cluster percentile confidence intervals from 2,000 resamples; at least 1,900 finite estimates were required. Fitted prediction models remained fixed. Recorded-level subset results concern admission encounters whose outcome remained known when absent level-of-care information was treated as uncertain. They are not a representative alternative population.

| eTable 38. Five-Variable Logistic Model Performance in 2023 | | | | | |
| --- | --- | --- | --- | --- | --- |
| Decision group | Model | AUROC | Average precision | Brier score | Log loss |
| Admission | Simple logistic | 0.634 | 0.087 | 0.0429 | 0.1796 |
| Admission | Risk-only | 0.659 | 0.097 | 0.0427 | 0.1776 |
| Discharge | Simple logistic | 0.722 | 0.066 | 0.0212 | 0.0987 |
| Discharge | Risk-only | 0.788 | 0.093 | 0.0208 | 0.0932 |

Comparator fitted using ridge logistic regression. Points only. Age, arrival-to-decision time, prior completed adult ED visits within 90 days, ESI, and arrival mode were used. Training-only preprocessing and 2022 intercept recalibration were applied. Simple logistic indicates the five-variable logistic model. ESI indicates Emergency Severity Index.

| eTable 39. Model Performance and Prediction Sensitivities in 2024 | | | | | | | | | | | | | |
| --- | --- | --- | --- | --- | --- | --- | --- | --- | --- | --- | --- | --- | --- |
| Model | Subset | Prediction | Encounters | Events | Observed | Predicted | AUROC | Average precision | Brier | Log loss | O/E | Cal. intercept | Cal. slope |
| Discharge | | | | | | | | | | | | | |
| Primary | All | 2022 recalibration | 50,675 | 1,066 | 0.021 | 0.023 | 0.786 | 0.086 | 0.020 | 0.090 | 0.928 | −0.080 | 0.982 |
| Primary | Known conservative labels | 2022 recalibration | 1,445 | 1,066 | 0.738 | 0.055 | 0.652 | 0.824 | 0.654 | 2.398 | 13.340 | 4.590 | 0.492 |
| Primary | All | Unrecalibrated | 50,675 | 1,066 | 0.021 | 0.022 | 0.786 | 0.086 | 0.020 | 0.090 | 0.935 | −0.071 | 0.982 |
| Without triage | All | 2022 recalibration | 50,675 | 1,066 | 0.021 | 0.022 | 0.781 | 0.084 | 0.020 | 0.091 | 0.937 | −0.069 | 1.042 |
| Without triage | All | Unrecalibrated | 50,675 | 1,066 | 0.021 | 0.023 | 0.781 | 0.084 | 0.020 | 0.091 | 0.933 | −0.074 | 1.042 |
| Without laboratory predictors | All | 2022 recalibration | 50,675 | 1,066 | 0.021 | 0.024 | 0.761 | 0.072 | 0.020 | 0.092 | 0.886 | −0.127 | 1.098 |
| Without laboratory predictors | All | Unrecalibrated | 50,675 | 1,066 | 0.021 | 0.025 | 0.761 | 0.072 | 0.020 | 0.093 | 0.840 | −0.183 | 1.098 |
| Training prevalence | All | Constant | 50,675 | 1,066 | 0.021 | 0.018 | 0.500 | 0.021 | 0.021 | 0.102 | 1.171 | 0.161 | NE |
| Simple logistic | All | 2022 recalibration | 50,675 | 1,066 | 0.021 | 0.025 | 0.723 | 0.058 | 0.020 | 0.096 | 0.853 | −0.166 | 0.998 |
| Admission | | | | | | | | | | | | | |
| Primary | All | 2022 recalibration | 26,411 | 1,125 | 0.043 | 0.046 | 0.676 | 0.094 | 0.040 | 0.168 | 0.933 | −0.074 | 1.110 |
| Primary | Known conservative labels | 2022 recalibration | 20,478 | 1,125 | 0.055 | 0.049 | 0.643 | 0.104 | 0.051 | 0.206 | 1.127 | 0.128 | 0.926 |
| Primary | All | Unrecalibrated | 26,411 | 1,125 | 0.043 | 0.048 | 0.676 | 0.094 | 0.040 | 0.168 | 0.884 | −0.131 | 1.110 |
| Without triage | All | 2022 recalibration | 26,411 | 1,125 | 0.043 | 0.045 | 0.669 | 0.081 | 0.040 | 0.169 | 0.940 | −0.066 | 1.187 |
| Without triage | All | Unrecalibrated | 26,411 | 1,125 | 0.043 | 0.048 | 0.669 | 0.081 | 0.040 | 0.169 | 0.896 | −0.117 | 1.187 |
| Without laboratory predictors | All | 2022 recalibration | 26,411 | 1,125 | 0.043 | 0.047 | 0.666 | 0.086 | 0.040 | 0.169 | 0.913 | −0.097 | 1.110 |
| Without laboratory predictors | All | Unrecalibrated | 26,411 | 1,125 | 0.043 | 0.050 | 0.666 | 0.086 | 0.040 | 0.169 | 0.856 | −0.166 | 1.110 |
| Training prevalence | All | Constant | 26,411 | 1,125 | 0.043 | 0.049 | 0.500 | 0.043 | 0.041 | 0.177 | 0.862 | −0.155 | NE |
| Simple logistic | All | 2022 recalibration | 26,411 | 1,125 | 0.043 | 0.046 | 0.660 | 0.086 | 0.040 | 0.169 | 0.922 | −0.086 | 1.160 |

Point estimates; 95% confidence intervals for the primary model are in Table 4. Observed and predicted values are event proportions. All fitted models and recalibration adjustments use training and tuning data only; no 2024 recalibration was applied. Known conservative labels identifies encounters whose composite remains known if missing level-of-care information is classified as uncertain; original labels and predictions are retained. Calibration intercept fixes slope at 1; slope is jointly estimated with an intercept. NE indicates not estimable; AUROC, area under the receiver operating characteristic curve; O/E, observed-to-expected event ratio.

| eTable 40. Sequential Selection by Patient Characteristics in 2023 | | | | |
| --- | --- | --- | --- | --- |
| Characteristic | Category | Rule | Selected, % | Event reach, % |
| Discharge | | | | |
| Sex | Female | Random | 3.97 | 2.57 |
| Sex | Female | ESI-first | 3.95 | 4.77 |
| Sex | Female | Age-first | 4.31 | 8.81 |
| Sex | Female | Prior-visits-first | 3.24 | 14.31 |
| Sex | Female | Simple logistic | 3.86 | 13.76 |
| Sex | Female | Risk-only | 3.28 | 19.82 |
| Sex | Female | Risk-plus-window | 3.26 | 19.45 |
| Sex | Male or unknown sex | Random | 3.72 | 2.87 |
| Sex | Male or unknown sex | ESI-first | 4.45 | 5.35 |
| Sex | Male or unknown sex | Age-first | 3.69 | 5.74 |
| Sex | Male or unknown sex | Prior-visits-first | 5.33 | 17.78 |
| Sex | Male or unknown sex | Simple logistic | 4.55 | 16.25 |
| Sex | Male or unknown sex | Risk-only | 4.61 | 19.12 |
| Sex | Male or unknown sex | Risk-plus-window | 4.62 | 19.50 |
| Age | 18–39 | Random | 3.81 | — |
| Age | 18–39 | ESI-first | 3.74 | 5.81 |
| Age | 18–39 | Age-first | 0.00 | 0.00 |
| Age | 18–39 | Prior-visits-first | 3.31 | 18.99 |
| Age | 18–39 | Simple logistic | 0.82 | 10.85 |
| Age | 18–39 | Risk-only | 1.25 | 11.63 |
| Age | 18–39 | Risk-plus-window | 1.24 | 12.02 |
| Age | 40–64 | Random | 3.93 | — |
| Age | 40–64 | ESI-first | 4.32 | 3.86 |
| Age | 40–64 | Age-first | 0.00 | 0.00 |
| Age | 40–64 | Prior-visits-first | 5.04 | 18.07 |
| Age | 40–64 | Simple logistic | 3.20 | 12.53 |
| Age | 40–64 | Risk-only | 3.71 | 17.59 |
| Age | 40–64 | Risk-plus-window | 3.66 | 17.35 |
| Age | 65 or older | Random | 3.84 | 3.54 |
| Age | 65 or older | ESI-first | 4.69 | 5.82 |
| Age | 65 or older | Age-first | 17.74 | 19.75 |
| Age | 65 or older | Prior-visits-first | 4.18 | 11.90 |
| Age | 65 or older | Simple logistic | 11.71 | 20.25 |
| Age | 65 or older | Risk-only | 8.76 | 26.58 |
| Age | 65 or older | Risk-plus-window | 8.85 | 26.58 |
| Race | Asian | Random | 3.69 | — |
| Race | Asian | ESI-first | 4.03 | — |
| Race | Asian | Age-first | 5.69 | — |
| Race | Asian | Prior-visits-first | 1.37 | — |
| Race | Asian | Simple logistic | 3.48 | 13.67 |
| Race | Asian | Risk-only | 3.75 | 20.14 |
| Race | Asian | Risk-plus-window | 3.84 | 21.58 |
| Race | Black or African American | Random | 4.03 | — |
| Race | Black or African American | ESI-first | 3.90 | — |
| Race | Black or African American | Age-first | 2.06 | — |
| Race | Black or African American | Prior-visits-first | 12.63 | 26.80 |
| Race | Black or African American | Simple logistic | 7.59 | 22.68 |
| Race | Black or African American | Risk-only | 6.54 | 19.59 |
| Race | Black or African American | Risk-plus-window | 6.45 | 19.59 |
| Race | Declines to state | Random | 3.87 | 0.00 |
| Race | Declines to state | ESI-first | 3.93 | — |
| Race | Declines to state | Age-first | — | — |
| Race | Declines to state | Prior-visits-first | 2.86 | — |
| Race | Declines to state | Simple logistic | 1.79 | — |
| Race | Declines to state | Risk-only | 2.03 | — |
| Race | Declines to state | Risk-plus-window | 2.03 | — |
| Race | Multiple recorded labels | Random | 3.58 | 0.00 |
| Race | Multiple recorded labels | ESI-first | 6.81 | — |
| Race | Multiple recorded labels | Age-first | 3.37 | — |
| Race | Multiple recorded labels | Prior-visits-first | 5.98 | 40.00 |
| Race | Multiple recorded labels | Simple logistic | 6.81 | — |
| Race | Multiple recorded labels | Risk-only | 4.68 | — |
| Race | Multiple recorded labels | Risk-plus-window | 4.95 | — |
| Race | Other | Random | 3.73 | — |
| Race | Other | ESI-first | 3.50 | — |
| Race | Other | Age-first | 1.68 | 5.54 |
| Race | Other | Prior-visits-first | 3.94 | 16.94 |
| Race | Other | Simple logistic | 2.73 | 13.03 |
| Race | Other | Risk-only | 2.90 | 16.61 |
| Race | Other | Risk-plus-window | 2.87 | 15.96 |
| Race | White | Random | 4.10 | 2.92 |
| Race | White | ESI-first | 4.76 | 4.94 |
| Race | White | Age-first | 6.52 | 7.64 |
| Race | White | Prior-visits-first | 4.04 | 14.38 |
| Race | White | Simple logistic | 5.35 | 13.71 |
| Race | White | Risk-only | 4.34 | 19.78 |
| Race | White | Risk-plus-window | 4.33 | 19.55 |
| Ethnicity | Hispanic/Latino | Random | 3.82 | — |
| Ethnicity | Hispanic/Latino | ESI-first | 3.44 | — |
| Ethnicity | Hispanic/Latino | Age-first | — | 4.28 |
| Ethnicity | Hispanic/Latino | Prior-visits-first | — | 18.42 |
| Ethnicity | Hispanic/Latino | Simple logistic | — | — |
| Ethnicity | Hispanic/Latino | Risk-only | 3.09 | — |
| Ethnicity | Hispanic/Latino | Risk-plus-window | 3.04 | — |
| Ethnicity | Non-Hispanic/non-Latino | Random | 3.87 | — |
| Ethnicity | Non-Hispanic/non-Latino | ESI-first | 4.52 | — |
| Ethnicity | Non-Hispanic/non-Latino | Age-first | 5.43 | 8.66 |
| Ethnicity | Non-Hispanic/non-Latino | Prior-visits-first | 4.14 | 15.31 |
| Ethnicity | Non-Hispanic/non-Latino | Simple logistic | 4.91 | 15.05 |
| Ethnicity | Non-Hispanic/non-Latino | Risk-only | 4.28 | 20.37 |
| Ethnicity | Non-Hispanic/non-Latino | Risk-plus-window | 4.30 | 20.51 |
| Language | English | Random | 3.95 | — |
| Language | English | ESI-first | 4.31 | — |
| Language | English | Age-first | 3.67 | 6.08 |
| Language | English | Prior-visits-first | 4.49 | 17.34 |
| Language | English | Simple logistic | 4.12 | 15.15 |
| Language | English | Risk-only | 3.73 | 19.17 |
| Language | English | Risk-plus-window | 3.73 | 19.17 |
| Language | Spanish | Random | 3.86 | — |
| Language | Spanish | ESI-first | 3.41 | — |
| Language | Spanish | Age-first | 2.39 | — |
| Language | Spanish | Prior-visits-first | 2.75 | — |
| Language | Spanish | Simple logistic | 2.98 | 12.00 |
| Language | Spanish | Risk-only | 3.02 | 16.80 |
| Language | Spanish | Risk-plus-window | 2.98 | 16.00 |
| Interpreter need | Not required | Random | 3.92 | — |
| Interpreter need | Not required | ESI-first | 4.28 | — |
| Interpreter need | Not required | Age-first | 3.74 | 6.07 |
| Interpreter need | Not required | Prior-visits-first | 4.42 | 17.10 |
| Interpreter need | Not required | Simple logistic | 4.11 | 15.19 |
| Interpreter need | Not required | Risk-only | 3.75 | 19.57 |
| Interpreter need | Not required | Risk-plus-window | 3.75 | 19.57 |
| Interpreter need | Required | Random | — | — |
| Interpreter need | Required | ESI-first | — | — |
| Interpreter need | Required | Age-first | — | 13.41 |
| Interpreter need | Required | Prior-visits-first | 2.90 | 10.61 |
| Interpreter need | Required | Simple logistic | 4.35 | 13.97 |
| Interpreter need | Required | Risk-only | 4.27 | 18.99 |
| Interpreter need | Required | Risk-plus-window | 4.25 | 18.99 |
| Admission | | | | |
| Sex | Female | Random | 3.58 | 3.56 |
| Sex | Female | ESI-first | 3.31 | 6.52 |
| Sex | Female | Age-first | 4.31 | 3.95 |
| Sex | Female | Prior-visits-first | 3.54 | 7.71 |
| Sex | Female | Simple logistic | 3.42 | 10.28 |
| Sex | Female | Risk-only | 3.34 | 10.08 |
| Sex | Female | Risk-plus-window | 3.36 | 10.87 |
| Sex | Male or unknown sex | Random | 3.42 | 2.64 |
| Sex | Male or unknown sex | ESI-first | 4.00 | 7.92 |
| Sex | Male or unknown sex | Age-first | 2.76 | 2.48 |
| Sex | Male or unknown sex | Prior-visits-first | 4.01 | 7.76 |
| Sex | Male or unknown sex | Simple logistic | 4.34 | 12.05 |
| Sex | Male or unknown sex | Risk-only | 3.97 | 10.07 |
| Sex | Male or unknown sex | Risk-plus-window | 3.95 | 9.90 |
| Age | 18–39 | Random | 3.23 | — |
| Age | 18–39 | ESI-first | 4.15 | 8.33 |
| Age | 18–39 | Age-first | 0.00 | 0.00 |
| Age | 18–39 | Prior-visits-first | 4.61 | 14.74 |
| Age | 18–39 | Simple logistic | 2.67 | 15.38 |
| Age | 18–39 | Risk-only | 2.34 | 10.26 |
| Age | 18–39 | Risk-plus-window | 2.21 | 9.62 |
| Age | 40–64 | Random | 3.65 | — |
| Age | 40–64 | ESI-first | 3.42 | 7.20 |
| Age | 40–64 | Age-first | 0.00 | 0.00 |
| Age | 40–64 | Prior-visits-first | 4.98 | 9.80 |
| Age | 40–64 | Simple logistic | 3.91 | 10.95 |
| Age | 40–64 | Risk-only | 3.54 | 12.10 |
| Age | 40–64 | Risk-plus-window | 3.47 | 11.82 |
| Age | 65 or older | Random | 3.48 | 2.96 |
| Age | 65 or older | ESI-first | 3.64 | 7.06 |
| Age | 65 or older | Age-first | 6.81 | 5.75 |
| Age | 65 or older | Prior-visits-first | 2.77 | 4.76 |
| Age | 65 or older | Simple logistic | 4.19 | 10.34 |
| Age | 65 or older | Risk-only | 4.10 | 8.87 |
| Age | 65 or older | Risk-plus-window | 4.17 | 9.69 |
| Race | Asian | Random | 3.17 | — |
| Race | Asian | ESI-first | 3.93 | 9.13 |
| Race | Asian | Age-first | 3.75 | — |
| Race | Asian | Prior-visits-first | 2.32 | — |
| Race | Asian | Simple logistic | 2.89 | 7.83 |
| Race | Asian | Risk-only | 3.58 | 11.30 |
| Race | Asian | Risk-plus-window | 3.62 | 12.17 |
| Race | Black or African American | Random | 4.27 | — |
| Race | Black or African American | ESI-first | 4.63 | — |
| Race | Black or African American | Age-first | 0.94 | 0.00 |
| Race | Black or African American | Prior-visits-first | 9.27 | 23.08 |
| Race | Black or African American | Simple logistic | 8.40 | 25.64 |
| Race | Black or African American | Risk-only | 4.71 | — |
| Race | Black or African American | Risk-plus-window | 4.78 | — |
| Race | Multiple recorded labels | Random | 3.48 | — |
| Race | Multiple recorded labels | ESI-first | 6.02 | — |
| Race | Multiple recorded labels | Age-first | — | — |
| Race | Multiple recorded labels | Prior-visits-first | 6.16 | — |
| Race | Multiple recorded labels | Simple logistic | 5.49 | — |
| Race | Multiple recorded labels | Risk-only | 5.09 | — |
| Race | Multiple recorded labels | Risk-plus-window | 4.82 | — |
| Race | Native Hawaiian or Other Pacific Islander | Random | 3.75 | — |
| Race | Native Hawaiian or Other Pacific Islander | ESI-first | — | — |
| Race | Native Hawaiian or Other Pacific Islander | Age-first | — | 0.00 |
| Race | Native Hawaiian or Other Pacific Islander | Prior-visits-first | 4.38 | — |
| Race | Native Hawaiian or Other Pacific Islander | Simple logistic | 4.17 | — |
| Race | Native Hawaiian or Other Pacific Islander | Risk-only | 4.58 | — |
| Race | Native Hawaiian or Other Pacific Islander | Risk-plus-window | 4.58 | — |
| Race | Other | Random | 3.58 | — |
| Race | Other | ESI-first | 3.22 | 5.29 |
| Race | Other | Age-first | 1.79 | — |
| Race | Other | Prior-visits-first | 4.16 | 4.85 |
| Race | Other | Simple logistic | 3.44 | 6.17 |
| Race | Other | Risk-only | 3.74 | 10.57 |
| Race | Other | Risk-plus-window | 3.65 | 10.57 |
| Race | White | Random | 3.48 | 2.24 |
| Race | White | ESI-first | 3.57 | 6.73 |
| Race | White | Age-first | 4.90 | 4.29 |
| Race | White | Prior-visits-first | 3.20 | 6.73 |
| Race | White | Simple logistic | 3.74 | 11.02 |
| Race | White | Risk-only | 3.30 | 7.76 |
| Race | White | Risk-plus-window | 3.35 | 7.96 |
| Ethnicity | Non-Hispanic/non-Latino | Random | 3.46 | — |
| Ethnicity | Non-Hispanic/non-Latino | ESI-first | 3.69 | 7.69 |
| Ethnicity | Non-Hispanic/non-Latino | Age-first | 4.23 | — |
| Ethnicity | Non-Hispanic/non-Latino | Prior-visits-first | 3.48 | 8.28 |
| Ethnicity | Non-Hispanic/non-Latino | Simple logistic | 3.95 | 12.00 |
| Ethnicity | Non-Hispanic/non-Latino | Risk-only | 3.55 | 9.91 |
| Ethnicity | Non-Hispanic/non-Latino | Risk-plus-window | 3.57 | 10.26 |
| Language | English | Random | 3.43 | — |
| Language | English | ESI-first | 3.61 | 7.12 |
| Language | English | Age-first | 3.50 | — |
| Language | English | Prior-visits-first | 3.89 | — |
| Language | English | Simple logistic | 3.95 | 12.40 |
| Language | English | Risk-only | 3.44 | 9.87 |
| Language | English | Risk-plus-window | 3.43 | 9.99 |
| Language | Mandarin | Random | 3.75 | — |
| Language | Mandarin | ESI-first | 5.63 | — |
| Language | Mandarin | Age-first | 6.83 | — |
| Language | Mandarin | Prior-visits-first | — | — |
| Language | Mandarin | Simple logistic | 3.92 | — |
| Language | Mandarin | Risk-only | 4.95 | — |
| Language | Mandarin | Risk-plus-window | 5.12 | — |
| Language | Other | Random | 3.38 | — |
| Language | Other | ESI-first | 3.01 | 0.00 |
| Language | Other | Age-first | 5.64 | — |
| Language | Other | Prior-visits-first | 3.20 | — |
| Language | Other | Simple logistic | 4.14 | — |
| Language | Other | Risk-only | 3.01 | 0.00 |
| Language | Other | Risk-plus-window | 3.01 | 0.00 |
| Language | Spanish | Random | 4.07 | — |
| Language | Spanish | ESI-first | 3.69 | — |
| Language | Spanish | Age-first | 1.66 | — |
| Language | Spanish | Prior-visits-first | 4.20 | — |
| Language | Spanish | Simple logistic | 3.35 | — |
| Language | Spanish | Risk-only | 3.99 | 12.73 |
| Language | Spanish | Risk-plus-window | 3.92 | 12.73 |
| Language | Vietnamese | Random | 2.95 | — |
| Language | Vietnamese | ESI-first | 3.88 | — |
| Language | Vietnamese | Age-first | 4.81 | — |
| Language | Vietnamese | Prior-visits-first | 1.71 | — |
| Language | Vietnamese | Simple logistic | 2.17 | — |
| Language | Vietnamese | Risk-only | 4.81 | — |
| Language | Vietnamese | Risk-plus-window | 5.12 | — |
| Interpreter need | Not required | Random | 3.50 | — |
| Interpreter need | Not required | ESI-first | 3.67 | 7.24 |
| Interpreter need | Not required | Age-first | 3.53 | — |
| Interpreter need | Not required | Prior-visits-first | 3.86 | — |
| Interpreter need | Not required | Simple logistic | 3.98 | 12.36 |
| Interpreter need | Not required | Risk-only | 3.48 | 10.13 |
| Interpreter need | Not required | Risk-plus-window | 3.48 | 10.24 |

Ten-minute reviews, thresholds targeting 5% in 2022, and zero start delay. Each rule selects from the full eligible decision group before results are summarized by subgroup; capacity is not allocated separately to subgroups. Categories follow training frequencies; age groups use recorded age. Male and unknown sex are pooled; missing sex is separate. Rows require at least 20 events and 11 encounters and may be withheld to protect other cells. Counts of 1–10, corresponding percentages, and selected cells from which they could be calculated are not shown (—). Selection uses all subgroup encounters as denominator; reach uses subgroup events. These descriptions do not establish subgroup calibration, discrimination, equitable clinical benefit, or causation.

| eTable 41. Sequential Selection by Patient Characteristics in 2024 | | | | |
| --- | --- | --- | --- | --- |
| Characteristic | Category | Rule | Selected, % | Event reach, % |
| Discharge | | | | |
| Sex | Female | Random | 3.97 | 3.28 |
| Sex | Female | ESI-first | 3.82 | 5.75 |
| Sex | Female | Age-first | 4.22 | 6.93 |
| Sex | Female | Prior-visits-first | 3.22 | 13.69 |
| Sex | Female | Simple logistic | 3.84 | 11.13 |
| Sex | Female | Risk-only | 3.42 | 18.80 |
| Sex | Female | Risk-plus-window | 3.52 | 19.34 |
| Sex | Male, non-binary or unknown sex | Random | 3.80 | 3.09 |
| Sex | Male, non-binary or unknown sex | ESI-first | 4.48 | 5.98 |
| Sex | Male, non-binary or unknown sex | Age-first | 3.90 | 5.60 |
| Sex | Male, non-binary or unknown sex | Prior-visits-first | 5.28 | 17.95 |
| Sex | Male, non-binary or unknown sex | Simple logistic | 4.69 | 12.74 |
| Sex | Male, non-binary or unknown sex | Risk-only | 4.73 | 20.08 |
| Sex | Male, non-binary or unknown sex | Risk-plus-window | 4.65 | 19.69 |
| Age | 18–39 | Random | 3.97 | — |
| Age | 18–39 | ESI-first | 3.90 | 5.98 |
| Age | 18–39 | Age-first | 0.00 | 0.00 |
| Age | 18–39 | Prior-visits-first | 3.24 | 15.06 |
| Age | 18–39 | Simple logistic | 0.82 | — |
| Age | 18–39 | Risk-only | 1.44 | 13.51 |
| Age | 18–39 | Risk-plus-window | 1.45 | 13.13 |
| Age | 40–64 | Random | 3.95 | 3.14 |
| Age | 40–64 | ESI-first | 4.05 | 6.04 |
| Age | 40–64 | Age-first | 0.00 | 0.00 |
| Age | 40–64 | Prior-visits-first | 4.70 | 16.67 |
| Age | 40–64 | Simple logistic | 3.35 | 12.08 |
| Age | 40–64 | Risk-only | 3.91 | 16.43 |
| Age | 40–64 | Risk-plus-window | 3.88 | 16.67 |
| Age | 65 or older | Random | 3.68 | — |
| Age | 65 or older | ESI-first | 4.54 | 5.60 |
| Age | 65 or older | Age-first | 17.10 | 17.05 |
| Age | 65 or older | Prior-visits-first | 4.72 | 15.27 |
| Age | 65 or older | Simple logistic | 11.30 | — |
| Age | 65 or older | Risk-only | 8.45 | 26.46 |
| Age | 65 or older | Risk-plus-window | 8.56 | 26.72 |
| Race | Asian | Random | 3.84 | — |
| Race | Asian | ESI-first | 4.71 | — |
| Race | Asian | Age-first | 5.54 | — |
| Race | Asian | Prior-visits-first | 1.38 | — |
| Race | Asian | Simple logistic | 3.63 | 9.66 |
| Race | Asian | Risk-only | 3.82 | 15.86 |
| Race | Asian | Risk-plus-window | 3.89 | 15.17 |
| Race | Black or African American | Random | 3.99 | — |
| Race | Black or African American | ESI-first | 3.47 | — |
| Race | Black or African American | Age-first | 1.40 | — |
| Race | Black or African American | Prior-visits-first | 10.71 | 20.51 |
| Race | Black or African American | Simple logistic | 6.91 | — |
| Race | Black or African American | Risk-only | 4.91 | 15.38 |
| Race | Black or African American | Risk-plus-window | 4.97 | 19.23 |
| Race | Declines to state | Random | 4.06 | — |
| Race | Declines to state | ESI-first | 4.19 | — |
| Race | Declines to state | Age-first | 2.13 | — |
| Race | Declines to state | Prior-visits-first | 2.06 | — |
| Race | Declines to state | Simple logistic | 2.06 | — |
| Race | Declines to state | Risk-only | 2.32 | — |
| Race | Declines to state | Risk-plus-window | 2.32 | — |
| Race | Multiple recorded labels | Random | 4.34 | — |
| Race | Multiple recorded labels | ESI-first | 4.90 | — |
| Race | Multiple recorded labels | Age-first | — | — |
| Race | Multiple recorded labels | Prior-visits-first | 4.34 | — |
| Race | Multiple recorded labels | Simple logistic | 3.36 | — |
| Race | Multiple recorded labels | Risk-only | 3.53 | — |
| Race | Multiple recorded labels | Risk-plus-window | 3.74 | — |
| Race | Other | Random | 3.95 | 4.78 |
| Race | Other | ESI-first | 3.58 | 6.62 |
| Race | Other | Age-first | 1.55 | 4.41 |
| Race | Other | Prior-visits-first | 3.65 | 16.54 |
| Race | Other | Simple logistic | 2.81 | 10.29 |
| Race | Other | Risk-only | 3.38 | 23.16 |
| Race | Other | Risk-plus-window | 3.30 | 21.32 |
| Race | White | Random | 3.82 | — |
| Race | White | ESI-first | 4.51 | 5.26 |
| Race | White | Age-first | 6.97 | 8.42 |
| Race | White | Prior-visits-first | 4.90 | 16.84 |
| Race | White | Simple logistic | 5.76 | 13.89 |
| Race | White | Risk-only | 4.65 | 18.53 |
| Race | White | Risk-plus-window | 4.69 | 19.37 |
| Ethnicity | Non-Hispanic/non-Latino | Random | 3.83 | 2.83 |
| Ethnicity | Non-Hispanic/non-Latino | ESI-first | 4.36 | 5.47 |
| Ethnicity | Non-Hispanic/non-Latino | Age-first | 5.58 | 7.42 |
| Ethnicity | Non-Hispanic/non-Latino | Prior-visits-first | 4.11 | 14.04 |
| Ethnicity | Non-Hispanic/non-Latino | Simple logistic | 4.98 | 11.88 |
| Ethnicity | Non-Hispanic/non-Latino | Risk-only | 4.23 | 17.41 |
| Ethnicity | Non-Hispanic/non-Latino | Risk-plus-window | 4.30 | 18.08 |
| Language | English | Random | 3.84 | — |
| Language | English | ESI-first | 4.15 | 5.61 |
| Language | English | Age-first | 3.83 | 5.09 |
| Language | English | Prior-visits-first | 4.58 | 16.09 |
| Language | English | Simple logistic | 4.23 | 11.23 |
| Language | English | Risk-only | 3.94 | 17.48 |
| Language | English | Risk-plus-window | 3.95 | 18.40 |
| Language | Spanish | Random | 4.11 | — |
| Language | Spanish | ESI-first | 3.75 | — |
| Language | Spanish | Age-first | 2.34 | 8.03 |
| Language | Spanish | Prior-visits-first | 2.30 | — |
| Language | Spanish | Simple logistic | 2.82 | — |
| Language | Spanish | Risk-only | 3.31 | 25.55 |
| Language | Spanish | Risk-plus-window | 3.32 | 22.63 |
| Interpreter need | Not required | Random | 3.83 | — |
| Interpreter need | Not required | ESI-first | 4.12 | 5.81 |
| Interpreter need | Not required | Age-first | 3.82 | 5.07 |
| Interpreter need | Not required | Prior-visits-first | 4.47 | 16.01 |
| Interpreter need | Not required | Simple logistic | 4.20 | 11.16 |
| Interpreter need | Not required | Risk-only | 3.91 | 17.59 |
| Interpreter need | Not required | Risk-plus-window | 3.93 | 18.49 |
| Interpreter need | Required | Random | — | — |
| Interpreter need | Required | ESI-first | — | 6.15 |
| Interpreter need | Required | Age-first | — | 12.29 |
| Interpreter need | Required | Prior-visits-first | 2.64 | 14.53 |
| Interpreter need | Required | Simple logistic | — | 15.64 |
| Interpreter need | Required | Risk-only | 4.31 | 28.49 |
| Interpreter need | Required | Risk-plus-window | 4.36 | 24.58 |
| Admission | | | | |
| Sex | Female | Random | 3.34 | 2.90 |
| Sex | Female | ESI-first | 3.28 | 5.40 |
| Sex | Female | Age-first | 3.98 | 5.20 |
| Sex | Female | Prior-visits-first | 3.48 | 8.40 |
| Sex | Female | Simple logistic | 3.03 | 8.40 |
| Sex | Female | Risk-only | 2.91 | 8.40 |
| Sex | Female | Risk-plus-window | 2.94 | 8.00 |
| Sex | Male, non-binary or unknown sex | Random | 3.47 | 4.16 |
| Sex | Male, non-binary or unknown sex | ESI-first | 3.73 | 6.88 |
| Sex | Male, non-binary or unknown sex | Age-first | 2.74 | 4.48 |
| Sex | Male, non-binary or unknown sex | Prior-visits-first | 3.66 | 7.04 |
| Sex | Male, non-binary or unknown sex | Simple logistic | 4.00 | 12.32 |
| Sex | Male, non-binary or unknown sex | Risk-only | 3.93 | 13.44 |
| Sex | Male, non-binary or unknown sex | Risk-plus-window | 3.97 | 13.28 |
| Age | 18–39 | Random | 3.20 | — |
| Age | 18–39 | ESI-first | 3.71 | — |
| Age | 18–39 | Age-first | 0.00 | 0.00 |
| Age | 18–39 | Prior-visits-first | 3.88 | 16.26 |
| Age | 18–39 | Simple logistic | 2.36 | 12.20 |
| Age | 18–39 | Risk-only | 2.14 | 11.38 |
| Age | 18–39 | Risk-plus-window | 2.02 | 9.76 |
| Age | 40–64 | Random | 3.48 | — |
| Age | 40–64 | ESI-first | 3.37 | — |
| Age | 40–64 | Age-first | 0.00 | 0.00 |
| Age | 40–64 | Prior-visits-first | 5.20 | 10.87 |
| Age | 40–64 | Simple logistic | 3.70 | 11.14 |
| Age | 40–64 | Risk-only | 3.06 | 10.60 |
| Age | 40–64 | Risk-plus-window | 3.13 | 10.33 |
| Age | 65 or older | Random | 3.41 | 3.23 |
| Age | 65 or older | ESI-first | 3.52 | 7.89 |
| Age | 65 or older | Age-first | 6.50 | 8.52 |
| Age | 65 or older | Prior-visits-first | 2.44 | 4.10 |
| Age | 65 or older | Simple logistic | 3.73 | 9.94 |
| Age | 65 or older | Risk-only | 4.01 | 11.51 |
| Age | 65 or older | Risk-plus-window | 4.07 | 11.51 |
| Race | Asian | Random | 3.53 | — |
| Race | Asian | ESI-first | 3.36 | — |
| Race | Asian | Age-first | 3.57 | — |
| Race | Asian | Prior-visits-first | 2.29 | 5.39 |
| Race | Asian | Simple logistic | 2.70 | 8.33 |
| Race | Asian | Risk-only | 3.66 | 14.22 |
| Race | Asian | Risk-plus-window | 3.88 | 14.22 |
| Race | Black or African American | Random | 2.81 | — |
| Race | Black or African American | ESI-first | 3.09 | — |
| Race | Black or African American | Age-first | 1.05 | — |
| Race | Black or African American | Prior-visits-first | 6.47 | 19.30 |
| Race | Black or African American | Simple logistic | 5.06 | — |
| Race | Black or African American | Risk-only | 4.08 | — |
| Race | Black or African American | Risk-plus-window | 4.01 | — |
| Race | Declines to state | Random | 3.12 | — |
| Race | Declines to state | ESI-first | 4.04 | — |
| Race | Declines to state | Age-first | — | 0.00 |
| Race | Declines to state | Prior-visits-first | 4.96 | — |
| Race | Declines to state | Simple logistic | 4.04 | — |
| Race | Declines to state | Risk-only | 4.04 | — |
| Race | Declines to state | Risk-plus-window | 4.04 | — |
| Race | Multiple recorded labels | Random | 3.29 | — |
| Race | Multiple recorded labels | ESI-first | 6.19 | — |
| Race | Multiple recorded labels | Age-first | 3.99 | — |
| Race | Multiple recorded labels | Prior-visits-first | 3.61 | — |
| Race | Multiple recorded labels | Simple logistic | 5.41 | 23.08 |
| Race | Multiple recorded labels | Risk-only | 4.38 | 21.15 |
| Race | Multiple recorded labels | Risk-plus-window | 4.38 | — |
| Race | Native Hawaiian or Other Pacific Islander | Random | — | — |
| Race | Native Hawaiian or Other Pacific Islander | ESI-first | — | — |
| Race | Native Hawaiian or Other Pacific Islander | Age-first | — | 0.00 |
| Race | Native Hawaiian or Other Pacific Islander | Prior-visits-first | — | — |
| Race | Native Hawaiian or Other Pacific Islander | Simple logistic | — | — |
| Race | Native Hawaiian or Other Pacific Islander | Risk-only | — | — |
| Race | Native Hawaiian or Other Pacific Islander | Risk-plus-window | — | — |
| Race | Other | Random | 3.09 | — |
| Race | Other | ESI-first | 3.29 | 4.15 |
| Race | Other | Age-first | 1.86 | — |
| Race | Other | Prior-visits-first | 4.09 | 9.43 |
| Race | Other | Simple logistic | 3.58 | 11.32 |
| Race | Other | Risk-only | 3.29 | 10.94 |
| Race | Other | Risk-plus-window | 3.22 | 9.81 |
| Race | White | Random | 3.63 | 3.97 |
| Race | White | ESI-first | 3.52 | 5.65 |
| Race | White | Age-first | 4.51 | 6.49 |
| Race | White | Prior-visits-first | 3.36 | 5.65 |
| Race | White | Simple logistic | 3.47 | 8.37 |
| Race | White | Risk-only | 3.17 | 9.00 |
| Race | White | Risk-plus-window | 3.20 | 9.21 |
| Ethnicity | Hispanic/Latino | Random | 3.20 | — |
| Ethnicity | Hispanic/Latino | ESI-first | 3.37 | — |
| Ethnicity | Hispanic/Latino | Age-first | 1.50 | — |
| Ethnicity | Hispanic/Latino | Prior-visits-first | — | — |
| Ethnicity | Hispanic/Latino | Simple logistic | — | — |
| Ethnicity | Hispanic/Latino | Risk-only | — | — |
| Ethnicity | Hispanic/Latino | Risk-plus-window | — | — |
| Ethnicity | Non-Hispanic/non-Latino | Random | 3.47 | — |
| Ethnicity | Non-Hispanic/non-Latino | ESI-first | 3.49 | 6.10 |
| Ethnicity | Non-Hispanic/non-Latino | Age-first | 3.96 | — |
| Ethnicity | Non-Hispanic/non-Latino | Prior-visits-first | 3.31 | 6.81 |
| Ethnicity | Non-Hispanic/non-Latino | Simple logistic | 3.50 | 10.33 |
| Ethnicity | Non-Hispanic/non-Latino | Risk-only | 3.40 | 10.80 |
| Ethnicity | Non-Hispanic/non-Latino | Risk-plus-window | 3.46 | 10.68 |
| Language | English | Random | 3.41 | 3.08 |
| Language | English | ESI-first | 3.55 | 5.93 |
| Language | English | Age-first | 3.17 | 4.45 |
| Language | English | Prior-visits-first | 3.63 | 7.07 |
| Language | English | Simple logistic | 3.55 | 10.03 |
| Language | English | Risk-only | 3.24 | 10.38 |
| Language | English | Risk-plus-window | 3.23 | 10.15 |
| Language | Mandarin | Random | 3.68 | 0.00 |
| Language | Mandarin | ESI-first | 3.33 | — |
| Language | Mandarin | Age-first | 7.18 | — |
| Language | Mandarin | Prior-visits-first | 3.15 | — |
| Language | Mandarin | Simple logistic | 1.93 | — |
| Language | Mandarin | Risk-only | 4.03 | — |
| Language | Mandarin | Risk-plus-window | 4.55 | — |
| Language | Other | Random | 4.00 | — |
| Language | Other | ESI-first | 3.82 | — |
| Language | Other | Age-first | 7.45 | — |
| Language | Other | Prior-visits-first | — | — |
| Language | Other | Simple logistic | — | — |
| Language | Other | Risk-only | 4.36 | — |
| Language | Other | Risk-plus-window | 4.55 | — |
| Language | Spanish | Random | 3.25 | — |
| Language | Spanish | ESI-first | 3.30 | — |
| Language | Spanish | Age-first | 2.01 | — |
| Language | Spanish | Prior-visits-first | 3.84 | — |
| Language | Spanish | Simple logistic | 3.89 | 14.17 |
| Language | Spanish | Risk-only | 3.70 | 13.33 |
| Language | Spanish | Risk-plus-window | 3.76 | 12.50 |
| Language | Vietnamese | Random | 4.29 | — |
| Language | Vietnamese | ESI-first | 3.39 | — |
| Language | Vietnamese | Age-first | 5.36 | — |
| Language | Vietnamese | Prior-visits-first | 3.04 | — |
| Language | Vietnamese | Simple logistic | 2.32 | — |
| Language | Vietnamese | Risk-only | 2.68 | 0.00 |
| Language | Vietnamese | Risk-plus-window | 3.21 | 0.00 |
| Interpreter need | Not required | Random | 3.41 | 2.96 |
| Interpreter need | Not required | ESI-first | 3.52 | 5.70 |
| Interpreter need | Not required | Age-first | 3.23 | 4.50 |
| Interpreter need | Not required | Prior-visits-first | 3.61 | 7.02 |
| Interpreter need | Not required | Simple logistic | 3.51 | 9.98 |
| Interpreter need | Not required | Risk-only | 3.27 | 10.31 |
| Interpreter need | Not required | Risk-plus-window | 3.28 | 10.09 |

Ten-minute reviews, thresholds targeting 5% in 2022, and zero start delay. Each rule selects from the full eligible decision group before results are summarized by subgroup; capacity is not allocated separately to subgroups. Categories follow training frequencies; age groups use recorded age. Male and unknown sex are pooled with non-binary sex; missing sex is separate. Rows require at least 20 events and 11 encounters and may be withheld to protect other cells. Counts of 1–10, corresponding percentages, and selected cells from which they could be calculated are not shown (—). Selection uses all subgroup encounters as denominator; reach uses subgroup events. These descriptions do not establish subgroup calibration, discrimination, equitable clinical benefit, or causation.

| eTable 42. Eventual Recorded Direction Changes During ED Care, 2020–2023 | | | | | | |
| --- | --- | --- | --- | --- | --- | --- |
| Year | Anchors | Later change | Change, % | Tied at anchor | Tie with later opposite | No change |
| Admission plan to discharge decision | | | | | | |
| 2020 | 19,276 | 2,524 | 13.09 | 29 | 13 | 16,723 |
| 2021 | 22,787 | 2,850 | 12.51 | 42 | 21 | 19,895 |
| 2022 | 26,656 | 3,579 | 13.43 | 44 | 20 | 23,033 |
| 2023 | 27,530 | 3,117 | 11.32 | 44 | 18 | 24,369 |
| 2020–2023 | 96,249 | 12,070 | 12.54 | 159 | 72 | 84,020 |
| Discharge decision to admission plan or handoff | | | | | | |
| 2020 | 43,588 | 517 | 1.19 | 425 | 44 | 42,646 |
| 2021 | 43,619 | 605 | 1.39 | 335 | 26 | 42,679 |
| 2022 | 46,154 | 627 | 1.36 | 332 | 31 | 45,195 |
| 2023 | 49,766 | 651 | 1.31 | 394 | 35 | 48,721 |
| 2020–2023 | 183,127 | 2,400 | 1.31 | 1,486 | 136 | 179,241 |

The first qualifying admission-plan or discharge-decision marker anchors each analysis. Opposite-direction markers must occur later and no later than departure or CDU entry. An opposite-direction marker at exactly the anchor is classified as a tie even if a later opposite marker occurs; the latter is a subset of ties. The anchor populations can overlap and differ from model-eligible time-zero groups. These are recorded changes, not verified attending overrides. ED indicates emergency department; CDU, clinical decision unit.

| eTable 43. Ranking of Individual Outcome Components in 2023 | | | | |
| --- | --- | --- | --- | --- |
| Decision group | Outcome | Known labels | Events | AUROC (95% CI) |
| Admission | Death within 7 days | — | 227 | 0.804 (0.777 to 0.830) |
| Admission | Return ending in admission | 24,401 | 562 | 0.581 (0.552 to 0.609) |
| Admission | Critical care within 24 hours | 23,024 | 342 | 0.699 (0.672 to 0.727) |
| Discharge | Death within 7 days | — | 15 | Not reported (<20 events) |
| Discharge | Return ending in admission | 48,313 | 1,050 | 0.788 (0.772 to 0.802) |
| Discharge | Critical care within 24 hours | 48,299 | 11 | Not reported (<20 events) |

Analysis using the composite risk score, without component-specific fitting. Parentheses contain 95% patient-cluster percentile confidence intervals from 2,000 resamples; at least 1,900 finite estimates were required. Fitted prediction models remained fixed. AUROC estimates were not reported with fewer than 20 events. Baseline and direct critical care were excluded from critical-care component ranking. To protect patient privacy, counts of 1–10, and selected cells from which they could be calculated, are not shown (—).

| eTable 44. Capped Windows by Recorded Decision Source | | | | | |
| --- | --- | --- | --- | --- | --- |
| Decision group | Decision source | Cap (min) | Encounters | Median (IQR), min | Mean (min) |
| Admission | Attending admission plan | 30 | 63,002 | 30.0 (30.0 to 30.0) | 27.9 |
| Admission | Simultaneous admission plan codes | 30 | 1,051 | 30.0 (30.0 to 30.0) | 28.0 |
| Discharge | Conditional discharge | 30 | 36,223 | 30.0 (28.0 to 30.0) | 27.1 |
| Discharge | Simultaneous discharge decision codes | 30 | 5,327 | 30.0 (24.0 to 30.0) | 26.2 |
| Discharge | Ready for discharge | 30 | 138,248 | 18.0 (10.0 to 30.0) | 18.2 |
| Admission | Resident admission plan | 30 | 20,626 | 30.0 (30.0 to 30.0) | 28.2 |
| Admission | Attending admission plan | 60 | 63,002 | 60.0 (48.0 to 60.0) | 50.9 |
| Admission | Simultaneous admission plan codes | 60 | 1,051 | 60.0 (49.0 to 60.0) | 50.9 |
| Discharge | Conditional discharge | 60 | 36,223 | 48.0 (28.0 to 60.0) | 43.3 |
| Discharge | Simultaneous discharge decision codes | 60 | 5,327 | 41.0 (24.0 to 60.0) | 40.2 |
| Discharge | Ready for discharge | 60 | 138,248 | 18.0 (10.0 to 30.0) | 22.4 |
| Admission | Resident admission plan | 60 | 20,626 | 60.0 (53.0 to 60.0) | 52.0 |
| Admission | Attending admission plan | 120 | 63,002 | 105.0 (48.0 to 120.0) | 84.6 |
| Admission | Simultaneous admission plan codes | 120 | 1,051 | 99.0 (49.0 to 120.0) | 83.7 |
| Discharge | Conditional discharge | 120 | 36,223 | 48.0 (28.0 to 86.0) | 58.1 |
| Discharge | Simultaneous discharge decision codes | 120 | 5,327 | 41.0 (24.0 to 74.0) | 52.2 |
| Discharge | Ready for discharge | 120 | 138,248 | 18.0 (10.0 to 30.0) | 25.1 |
| Admission | Resident admission plan | 120 | 20,626 | 109.0 (53.0 to 120.0) | 86.9 |

All 2020–2023 encounters with qualifying decisions, before outcome and temporal exclusions. These are capped review windows. Caps of 30 and 120 minutes did not apply to allocation. Simultaneous same-direction codes denote multiple qualifying markers at time zero. Status names do not independently identify the clinician entering each admission marker.

| eTable 45. Recorded Windows Before Model Exclusions in 2023 | | | | |
| --- | --- | --- | --- | --- |
| Decision group | Decision-time group | Review (min) | Encounters | Sufficient window (%) |
| Admission | Recorded decision | 5 | 24,533 | 98.0 |
| Admission | Recorded decision | 10 | 24,533 | 96.2 |
| Admission | Recorded decision | 15 | 24,533 | 93.9 |
| Discharge | Recorded decision | 5 | 48,872 | 95.0 |
| Discharge | Recorded decision | 10 | 48,872 | 84.9 |
| Discharge | Recorded decision | 15 | 48,872 | 69.7 |
| No time zero | Unknown | 5 | 5,252 | Unavailable |
| No time zero | Unknown | 10 | 5,252 | Unavailable |
| No time zero | Unknown | 15 | 5,252 | Unavailable |

All 2023 candidate encounters with the stated decision-time group, before model eligibility and known-outcome exclusions. The denominators differ from Table 2. Encounters with no usable decision time have no observed review window.

| eTable 46. Readiness and Conditional-Discharge Order Among Final Discharges | | |
| --- | --- | --- |
| Qualifying-marker order | Encounters | Readiness minus conditional discharge, median (IQR), min |
| Both, conditional discharge first | 35,403 | 29.0 (13.0 to 62.0) |
| Both, readiness first | 16,533 | −7.0 (−16.0 to −2.0) |
| Both, simultaneous | 5,317 | 0.0 (0.0 to 0.0) |
| Conditional discharge only | 492 | Not applicable |
| Neither qualifying marker | 10,948 | Not applicable |
| Readiness only | 118,107 | Not applicable |

All 186,800 candidate encounters with final discharge during 2020–2023; not the discharge-decision analysis cohort. Only qualifying markers within ED care are counted. Negative differences mean readiness preceded conditional discharge. Final disposition is used for this descriptive table only.

| eTable 47. Annual Pooling at the Main Ranking Budget in 2023 | | | |
| --- | --- | --- | --- |
| Decision group | Rule or contrast | Measure | Estimate (95% interval) |
| Admission | Random | Reach (%) | 4.8065 (4.7487 to 4.8576) |
| Admission | ESI-first | Reach (%) | 9.89 (8.69 to 11.29) |
| Admission | Risk-only | Reach (%) | 14.39 (12.25 to 16.37) |
| Admission | Risk-only minus ESI-first | Difference (pp) | 4.50 (2.20 to 6.61) |
| Discharge | Random | Reach (%) | 4.1983 (4.0806 to 4.3117) |
| Discharge | ESI-first | Reach (%) | 6.69 (5.95 to 7.48) |
| Discharge | Risk-only | Reach (%) | 25.00 (22.20 to 27.82) |
| Discharge | Risk-only minus ESI-first | Difference (pp) | 18.31 (15.56 to 21.10) |

Ten-minute reviews and 50 review-minutes per 100 encounters. Parentheses contain 95% patient-cluster percentile confidence intervals from 2,000 resamples; at least 1,900 finite estimates were required. Fitted prediction models remained fixed. The coverage diagnostic concerns these main-scenario statistics under empirical resampling and does not establish population coverage or daily uncertainty.

| eTable 48. Descriptive Full-Day and Annual Ranking Contrasts in 2023 | | | | | |
| --- | --- | --- | --- | --- | --- |
| Decision group | Allocation | Review min | Minutes per 100 | Contrast | Reach difference pp |
| Admission | Annual ranking | 10 | 100 | ESI-first minus Random | 5.78 |
| Admission | Annual ranking | 10 | 100 | Risk-only minus ESI-first | 9.60 |
| Admission | Annual ranking | 10 | 100 | Risk-only minus Random | 15.39 |
| Admission | Annual ranking | 10 | 100 | Risk plus window minus ESI-first | 9.33 |
| Admission | Annual ranking | 10 | 100 | Risk plus window minus Random | 15.12 |
| Admission | Annual ranking | 10 | 100 | Risk plus window minus Risk-only | −0.27 |
| Admission | Annual ranking | 10 | 25 | ESI-first minus Random | 4.73 |
| Admission | Annual ranking | 10 | 25 | Risk-only minus ESI-first | 1.50 |
| Admission | Annual ranking | 10 | 25 | Risk-only minus Random | 6.23 |
| Admission | Annual ranking | 10 | 25 | Risk plus window minus ESI-first | 1.41 |
| Admission | Annual ranking | 10 | 25 | Risk plus window minus Random | 6.14 |
| Admission | Annual ranking | 10 | 25 | Risk plus window minus Risk-only | −0.09 |
| Admission | Annual ranking | 10 | 50 | ESI-first minus Random | 5.08 |
| Admission | Annual ranking | 10 | 50 | Risk-only minus ESI-first | 4.50 |
| Admission | Annual ranking | 10 | 50 | Risk-only minus Random | 9.58 |
| Admission | Annual ranking | 10 | 50 | Risk plus window minus ESI-first | 4.23 |
| Admission | Annual ranking | 10 | 50 | Risk plus window minus Random | 9.31 |
| Admission | Annual ranking | 10 | 50 | Risk plus window minus Risk-only | −0.27 |
| Admission | Annual ranking | 15 | 100 | ESI-first minus Random | 5.37 |
| Admission | Annual ranking | 15 | 100 | Risk-only minus ESI-first | 5.93 |
| Admission | Annual ranking | 15 | 100 | Risk-only minus Random | 11.30 |
| Admission | Annual ranking | 15 | 100 | Risk plus window minus ESI-first | 5.66 |
| Admission | Annual ranking | 15 | 100 | Risk plus window minus Random | 11.03 |
| Admission | Annual ranking | 15 | 100 | Risk plus window minus Risk-only | −0.27 |
| Admission | Annual ranking | 15 | 25 | ESI-first minus Random | 4.65 |
| Admission | Annual ranking | 15 | 25 | Risk-only minus ESI-first | −0.01 |
| Admission | Annual ranking | 15 | 25 | Risk-only minus Random | 4.65 |
| Admission | Annual ranking | 15 | 25 | Risk plus window minus ESI-first | 0.17 |
| Admission | Annual ranking | 15 | 25 | Risk plus window minus Random | 4.83 |
| Admission | Annual ranking | 15 | 25 | Risk plus window minus Risk-only | 0.18 |
| Admission | Annual ranking | 15 | 50 | ESI-first minus Random | 4.89 |
| Admission | Annual ranking | 15 | 50 | Risk-only minus ESI-first | 2.24 |
| Admission | Annual ranking | 15 | 50 | Risk-only minus Random | 7.13 |
| Admission | Annual ranking | 15 | 50 | Risk plus window minus ESI-first | 2.33 |
| Admission | Annual ranking | 15 | 50 | Risk plus window minus Random | 7.22 |
| Admission | Annual ranking | 15 | 50 | Risk plus window minus Risk-only | 0.09 |
| Admission | Annual ranking | 5 | 100 | ESI-first minus Random | 7.24 |
| Admission | Annual ranking | 5 | 100 | Risk-only minus ESI-first | 13.12 |
| Admission | Annual ranking | 5 | 100 | Risk-only minus Random | 20.36 |
| Admission | Annual ranking | 5 | 100 | Risk plus window minus ESI-first | 13.21 |
| Admission | Annual ranking | 5 | 100 | Risk plus window minus Random | 20.45 |
| Admission | Annual ranking | 5 | 100 | Risk plus window minus Risk-only | 0.09 |
| Admission | Annual ranking | 5 | 25 | ESI-first minus Random | 5.15 |
| Admission | Annual ranking | 5 | 25 | Risk-only minus ESI-first | 4.62 |
| Admission | Annual ranking | 5 | 25 | Risk-only minus Random | 9.77 |
| Admission | Annual ranking | 5 | 25 | Risk plus window minus ESI-first | 4.53 |
| Admission | Annual ranking | 5 | 25 | Risk plus window minus Random | 9.68 |
| Admission | Annual ranking | 5 | 25 | Risk plus window minus Risk-only | −0.09 |
| Admission | Annual ranking | 5 | 50 | ESI-first minus Random | 5.85 |
| Admission | Annual ranking | 5 | 50 | Risk-only minus ESI-first | 9.91 |
| Admission | Annual ranking | 5 | 50 | Risk-only minus Random | 15.76 |
| Admission | Annual ranking | 5 | 50 | Risk plus window minus ESI-first | 9.73 |
| Admission | Annual ranking | 5 | 50 | Risk plus window minus Random | 15.58 |
| Admission | Annual ranking | 5 | 50 | Risk plus window minus Risk-only | −0.18 |
| Discharge | Annual ranking | 10 | 100 | ESI-first minus Random | 4.91 |
| Discharge | Annual ranking | 10 | 100 | Risk-only minus ESI-first | 24.14 |
| Discharge | Annual ranking | 10 | 100 | Risk-only minus Random | 29.05 |
| Discharge | Annual ranking | 10 | 100 | Risk plus window minus ESI-first | 24.14 |
| Discharge | Annual ranking | 10 | 100 | Risk plus window minus Random | 29.05 |
| Discharge | Annual ranking | 10 | 100 | Risk plus window minus Risk-only | 0.00 |
| Discharge | Annual ranking | 10 | 25 | ESI-first minus Random | 1.28 |
| Discharge | Annual ranking | 10 | 25 | Risk-only minus ESI-first | 11.79 |
| Discharge | Annual ranking | 10 | 25 | Risk-only minus Random | 13.07 |
| Discharge | Annual ranking | 10 | 25 | Risk plus window minus ESI-first | 12.45 |
| Discharge | Annual ranking | 10 | 25 | Risk plus window minus Random | 13.73 |
| Discharge | Annual ranking | 10 | 25 | Risk plus window minus Risk-only | 0.66 |
| Discharge | Annual ranking | 10 | 50 | ESI-first minus Random | 2.49 |
| Discharge | Annual ranking | 10 | 50 | Risk-only minus ESI-first | 18.31 |
| Discharge | Annual ranking | 10 | 50 | Risk-only minus Random | 20.80 |
| Discharge | Annual ranking | 10 | 50 | Risk plus window minus ESI-first | 18.03 |
| Discharge | Annual ranking | 10 | 50 | Risk plus window minus Random | 20.52 |
| Discharge | Annual ranking | 10 | 50 | Risk plus window minus Risk-only | −0.28 |
| Discharge | Annual ranking | 15 | 100 | ESI-first minus Random | 3.09 |
| Discharge | Annual ranking | 15 | 100 | Risk-only minus ESI-first | 18.72 |
| Discharge | Annual ranking | 15 | 100 | Risk-only minus Random | 21.82 |
| Discharge | Annual ranking | 15 | 100 | Risk plus window minus ESI-first | 18.26 |
| Discharge | Annual ranking | 15 | 100 | Risk plus window minus Random | 21.35 |
| Discharge | Annual ranking | 15 | 100 | Risk plus window minus Risk-only | −0.47 |
| Discharge | Annual ranking | 15 | 25 | ESI-first minus Random | 0.83 |
| Discharge | Annual ranking | 15 | 25 | Risk-only minus ESI-first | 8.32 |
| Discharge | Annual ranking | 15 | 25 | Risk-only minus Random | 9.15 |
| Discharge | Annual ranking | 15 | 25 | Risk plus window minus ESI-first | 9.35 |
| Discharge | Annual ranking | 15 | 25 | Risk plus window minus Random | 10.18 |
| Discharge | Annual ranking | 15 | 25 | Risk plus window minus Risk-only | 1.03 |
| Discharge | Annual ranking | 15 | 50 | ESI-first minus Random | 1.59 |
| Discharge | Annual ranking | 15 | 50 | Risk-only minus ESI-first | 13.25 |
| Discharge | Annual ranking | 15 | 50 | Risk-only minus Random | 14.84 |
| Discharge | Annual ranking | 15 | 50 | Risk plus window minus ESI-first | 14.10 |
| Discharge | Annual ranking | 15 | 50 | Risk plus window minus Random | 15.68 |
| Discharge | Annual ranking | 15 | 50 | Risk plus window minus Risk-only | 0.84 |
| Discharge | Annual ranking | 5 | 100 | ESI-first minus Random | 8.85 |
| Discharge | Annual ranking | 5 | 100 | Risk-only minus ESI-first | 30.38 |
| Discharge | Annual ranking | 5 | 100 | Risk-only minus Random | 39.23 |
| Discharge | Annual ranking | 5 | 100 | Risk plus window minus ESI-first | 29.73 |
| Discharge | Annual ranking | 5 | 100 | Risk plus window minus Random | 38.58 |
| Discharge | Annual ranking | 5 | 100 | Risk plus window minus Risk-only | −0.66 |
| Discharge | Annual ranking | 5 | 25 | ESI-first minus Random | 2.72 |
| Discharge | Annual ranking | 5 | 25 | Risk-only minus ESI-first | 19.49 |
| Discharge | Annual ranking | 5 | 25 | Risk-only minus Random | 22.22 |
| Discharge | Annual ranking | 5 | 25 | Risk plus window minus ESI-first | 19.40 |
| Discharge | Annual ranking | 5 | 25 | Risk plus window minus Random | 22.12 |
| Discharge | Annual ranking | 5 | 25 | Risk plus window minus Risk-only | −0.09 |
| Discharge | Annual ranking | 5 | 50 | ESI-first minus Random | 5.39 |
| Discharge | Annual ranking | 5 | 50 | Risk-only minus ESI-first | 25.74 |
| Discharge | Annual ranking | 5 | 50 | Risk-only minus Random | 31.13 |
| Discharge | Annual ranking | 5 | 50 | Risk plus window minus ESI-first | 25.84 |
| Discharge | Annual ranking | 5 | 50 | Risk plus window minus Random | 31.23 |
| Discharge | Annual ranking | 5 | 50 | Risk plus window minus Risk-only | 0.09 |
| Admission | Full-day ranking | 10 | 100 | ESI-first minus Random | 5.74 |
| Admission | Full-day ranking | 10 | 100 | Risk-only minus ESI-first | 7.67 |
| Admission | Full-day ranking | 10 | 100 | Risk-only minus Random | 13.40 |
| Admission | Full-day ranking | 10 | 100 | Risk plus window minus ESI-first | 7.58 |
| Admission | Full-day ranking | 10 | 100 | Risk plus window minus Random | 13.31 |
| Admission | Full-day ranking | 10 | 100 | Risk plus window minus Risk-only | −0.09 |
| Admission | Full-day ranking | 10 | 25 | ESI-first minus Random | 3.92 |
| Admission | Full-day ranking | 10 | 25 | Risk-only minus ESI-first | 2.15 |
| Admission | Full-day ranking | 10 | 25 | Risk-only minus Random | 6.07 |
| Admission | Full-day ranking | 10 | 25 | Risk plus window minus ESI-first | 2.06 |
| Admission | Full-day ranking | 10 | 25 | Risk plus window minus Random | 5.98 |
| Admission | Full-day ranking | 10 | 25 | Risk plus window minus Risk-only | −0.09 |
| Admission | Full-day ranking | 10 | 50 | ESI-first minus Random | 5.01 |
| Admission | Full-day ranking | 10 | 50 | Risk-only minus ESI-first | 4.58 |
| Admission | Full-day ranking | 10 | 50 | Risk-only minus Random | 9.59 |
| Admission | Full-day ranking | 10 | 50 | Risk plus window minus ESI-first | 4.85 |
| Admission | Full-day ranking | 10 | 50 | Risk plus window minus Random | 9.86 |
| Admission | Full-day ranking | 10 | 50 | Risk plus window minus Risk-only | 0.27 |
| Admission | Full-day ranking | 15 | 100 | ESI-first minus Random | 5.34 |
| Admission | Full-day ranking | 15 | 100 | Risk-only minus ESI-first | 5.50 |
| Admission | Full-day ranking | 15 | 100 | Risk-only minus Random | 10.85 |
| Admission | Full-day ranking | 15 | 100 | Risk plus window minus ESI-first | 5.23 |
| Admission | Full-day ranking | 15 | 100 | Risk plus window minus Random | 10.58 |
| Admission | Full-day ranking | 15 | 100 | Risk plus window minus Risk-only | −0.27 |
| Admission | Full-day ranking | 15 | 25 | ESI-first minus Random | 3.04 |
| Admission | Full-day ranking | 15 | 25 | Risk-only minus ESI-first | 1.15 |
| Admission | Full-day ranking | 15 | 25 | Risk-only minus Random | 4.19 |
| Admission | Full-day ranking | 15 | 25 | Risk plus window minus ESI-first | 1.24 |
| Admission | Full-day ranking | 15 | 25 | Risk plus window minus Random | 4.28 |
| Admission | Full-day ranking | 15 | 25 | Risk plus window minus Risk-only | 0.09 |
| Admission | Full-day ranking | 15 | 50 | ESI-first minus Random | 4.60 |
| Admission | Full-day ranking | 15 | 50 | Risk-only minus ESI-first | 2.44 |
| Admission | Full-day ranking | 15 | 50 | Risk-only minus Random | 7.04 |
| Admission | Full-day ranking | 15 | 50 | Risk plus window minus ESI-first | 2.26 |
| Admission | Full-day ranking | 15 | 50 | Risk plus window minus Random | 6.86 |
| Admission | Full-day ranking | 15 | 50 | Risk plus window minus Risk-only | −0.18 |
| Admission | Full-day ranking | 5 | 100 | ESI-first minus Random | 7.19 |
| Admission | Full-day ranking | 5 | 100 | Risk-only minus ESI-first | 13.06 |
| Admission | Full-day ranking | 5 | 100 | Risk-only minus Random | 20.25 |
| Admission | Full-day ranking | 5 | 100 | Risk plus window minus ESI-first | 13.06 |
| Admission | Full-day ranking | 5 | 100 | Risk plus window minus Random | 20.25 |
| Admission | Full-day ranking | 5 | 100 | Risk plus window minus Risk-only | 0.00 |
| Admission | Full-day ranking | 5 | 25 | ESI-first minus Random | 5.08 |
| Admission | Full-day ranking | 5 | 25 | Risk-only minus ESI-first | 4.79 |
| Admission | Full-day ranking | 5 | 25 | Risk-only minus Random | 9.87 |
| Admission | Full-day ranking | 5 | 25 | Risk plus window minus ESI-first | 4.88 |
| Admission | Full-day ranking | 5 | 25 | Risk plus window minus Random | 9.96 |
| Admission | Full-day ranking | 5 | 25 | Risk plus window minus Risk-only | 0.09 |
| Admission | Full-day ranking | 5 | 50 | ESI-first minus Random | 5.81 |
| Admission | Full-day ranking | 5 | 50 | Risk-only minus ESI-first | 8.05 |
| Admission | Full-day ranking | 5 | 50 | Risk-only minus Random | 13.86 |
| Admission | Full-day ranking | 5 | 50 | Risk plus window minus ESI-first | 7.96 |
| Admission | Full-day ranking | 5 | 50 | Risk plus window minus Random | 13.77 |
| Admission | Full-day ranking | 5 | 50 | Risk plus window minus Risk-only | −0.09 |
| Discharge | Full-day ranking | 10 | 100 | ESI-first minus Random | 4.92 |
| Discharge | Full-day ranking | 10 | 100 | Risk-only minus ESI-first | 23.39 |
| Discharge | Full-day ranking | 10 | 100 | Risk-only minus Random | 28.31 |
| Discharge | Full-day ranking | 10 | 100 | Risk plus window minus ESI-first | 23.01 |
| Discharge | Full-day ranking | 10 | 100 | Risk plus window minus Random | 27.94 |
| Discharge | Full-day ranking | 10 | 100 | Risk plus window minus Risk-only | −0.37 |
| Discharge | Full-day ranking | 10 | 25 | ESI-first minus Random | 1.36 |
| Discharge | Full-day ranking | 10 | 25 | Risk-only minus ESI-first | 12.74 |
| Discharge | Full-day ranking | 10 | 25 | Risk-only minus Random | 14.10 |
| Discharge | Full-day ranking | 10 | 25 | Risk plus window minus ESI-first | 12.84 |
| Discharge | Full-day ranking | 10 | 25 | Risk plus window minus Random | 14.19 |
| Discharge | Full-day ranking | 10 | 25 | Risk plus window minus Risk-only | 0.09 |
| Discharge | Full-day ranking | 10 | 50 | ESI-first minus Random | 2.50 |
| Discharge | Full-day ranking | 10 | 50 | Risk-only minus ESI-first | 16.99 |
| Discharge | Full-day ranking | 10 | 50 | Risk-only minus Random | 19.49 |
| Discharge | Full-day ranking | 10 | 50 | Risk plus window minus ESI-first | 16.99 |
| Discharge | Full-day ranking | 10 | 50 | Risk plus window minus Random | 19.49 |
| Discharge | Full-day ranking | 10 | 50 | Risk plus window minus Risk-only | 0.00 |
| Discharge | Full-day ranking | 15 | 100 | ESI-first minus Random | 3.00 |
| Discharge | Full-day ranking | 15 | 100 | Risk-only minus ESI-first | 18.07 |
| Discharge | Full-day ranking | 15 | 100 | Risk-only minus Random | 21.07 |
| Discharge | Full-day ranking | 15 | 100 | Risk plus window minus ESI-first | 18.35 |
| Discharge | Full-day ranking | 15 | 100 | Risk plus window minus Random | 21.35 |
| Discharge | Full-day ranking | 15 | 100 | Risk plus window minus Risk-only | 0.28 |
| Discharge | Full-day ranking | 15 | 25 | ESI-first minus Random | 0.87 |
| Discharge | Full-day ranking | 15 | 25 | Risk-only minus ESI-first | 9.11 |
| Discharge | Full-day ranking | 15 | 25 | Risk-only minus Random | 9.98 |
| Discharge | Full-day ranking | 15 | 25 | Risk plus window minus ESI-first | 9.58 |
| Discharge | Full-day ranking | 15 | 25 | Risk plus window minus Random | 10.45 |
| Discharge | Full-day ranking | 15 | 25 | Risk plus window minus Risk-only | 0.47 |
| Discharge | Full-day ranking | 15 | 50 | ESI-first minus Random | 1.57 |
| Discharge | Full-day ranking | 15 | 50 | Risk-only minus ESI-first | 14.30 |
| Discharge | Full-day ranking | 15 | 50 | Risk-only minus Random | 15.87 |
| Discharge | Full-day ranking | 15 | 50 | Risk plus window minus ESI-first | 14.12 |
| Discharge | Full-day ranking | 15 | 50 | Risk plus window minus Random | 15.68 |
| Discharge | Full-day ranking | 15 | 50 | Risk plus window minus Risk-only | −0.19 |
| Discharge | Full-day ranking | 5 | 100 | ESI-first minus Random | 8.73 |
| Discharge | Full-day ranking | 5 | 100 | Risk-only minus ESI-first | 30.50 |
| Discharge | Full-day ranking | 5 | 100 | Risk-only minus Random | 39.23 |
| Discharge | Full-day ranking | 5 | 100 | Risk plus window minus ESI-first | 29.94 |
| Discharge | Full-day ranking | 5 | 100 | Risk plus window minus Random | 38.67 |
| Discharge | Full-day ranking | 5 | 100 | Risk plus window minus Risk-only | −0.56 |
| Discharge | Full-day ranking | 5 | 25 | ESI-first minus Random | 2.82 |
| Discharge | Full-day ranking | 5 | 25 | Risk-only minus ESI-first | 17.90 |
| Discharge | Full-day ranking | 5 | 25 | Risk-only minus Random | 20.72 |
| Discharge | Full-day ranking | 5 | 25 | Risk plus window minus ESI-first | 17.34 |
| Discharge | Full-day ranking | 5 | 25 | Risk plus window minus Random | 20.15 |
| Discharge | Full-day ranking | 5 | 25 | Risk plus window minus Risk-only | −0.56 |
| Discharge | Full-day ranking | 5 | 50 | ESI-first minus Random | 5.53 |
| Discharge | Full-day ranking | 5 | 50 | Risk-only minus ESI-first | 24.77 |
| Discharge | Full-day ranking | 5 | 50 | Risk-only minus Random | 30.30 |
| Discharge | Full-day ranking | 5 | 50 | Risk plus window minus ESI-first | 24.11 |
| Discharge | Full-day ranking | 5 | 50 | Risk plus window minus Random | 29.64 |
| Discharge | Full-day ranking | 5 | 50 | Risk plus window minus Risk-only | −0.66 |
| Admission | Full-day ranking | 5 | 25 | Simple logistic minus Risk-only | −1.80 |
| Admission | Full-day ranking | 5 | 25 | Age first minus Risk-only | −10.24 |
| Admission | Full-day ranking | 5 | 25 | Prior visits first minus Risk-only | −3.99 |
| Admission | Full-day ranking | 5 | 50 | Simple logistic minus Risk-only | −0.72 |
| Admission | Full-day ranking | 5 | 50 | Age first minus Risk-only | −13.16 |
| Admission | Full-day ranking | 5 | 50 | Prior visits first minus Risk-only | −5.06 |
| Admission | Full-day ranking | 5 | 100 | Simple logistic minus Risk-only | −2.88 |
| Admission | Full-day ranking | 5 | 100 | Age first minus Risk-only | −19.12 |
| Admission | Full-day ranking | 5 | 100 | Prior visits first minus Risk-only | −9.35 |
| Admission | Full-day ranking | 10 | 25 | Simple logistic minus Risk-only | −1.17 |
| Admission | Full-day ranking | 10 | 25 | Age first minus Risk-only | −6.59 |
| Admission | Full-day ranking | 10 | 25 | Prior visits first minus Risk-only | −2.62 |
| Admission | Full-day ranking | 10 | 50 | Simple logistic minus Risk-only | −1.71 |
| Admission | Full-day ranking | 10 | 50 | Age first minus Risk-only | −10.06 |
| Admission | Full-day ranking | 10 | 50 | Prior visits first minus Risk-only | −3.90 |
| Admission | Full-day ranking | 10 | 100 | Simple logistic minus Risk-only | −0.54 |
| Admission | Full-day ranking | 10 | 100 | Age first minus Risk-only | −12.80 |
| Admission | Full-day ranking | 10 | 100 | Prior visits first minus Risk-only | −4.70 |
| Admission | Full-day ranking | 15 | 25 | Simple logistic minus Risk-only | −0.72 |
| Admission | Full-day ranking | 15 | 25 | Age first minus Risk-only | −4.68 |
| Admission | Full-day ranking | 15 | 25 | Prior visits first minus Risk-only | −1.81 |
| Admission | Full-day ranking | 15 | 50 | Simple logistic minus Risk-only | −0.72 |
| Admission | Full-day ranking | 15 | 50 | Age first minus Risk-only | −7.72 |
| Admission | Full-day ranking | 15 | 50 | Prior visits first minus Risk-only | −2.90 |
| Admission | Full-day ranking | 15 | 100 | Simple logistic minus Risk-only | −0.90 |
| Admission | Full-day ranking | 15 | 100 | Age first minus Risk-only | −10.64 |
| Admission | Full-day ranking | 15 | 100 | Prior visits first minus Risk-only | −4.26 |
| Admission | Annual ranking | 5 | 25 | Simple logistic minus Risk-only | −0.09 |
| Admission | Annual ranking | 5 | 25 | Age first minus Risk-only | −9.81 |
| Admission | Annual ranking | 5 | 25 | Prior visits first minus Risk-only | −3.64 |
| Admission | Annual ranking | 5 | 50 | Simple logistic minus Risk-only | −2.70 |
| Admission | Annual ranking | 5 | 50 | Age first minus Risk-only | −14.55 |
| Admission | Annual ranking | 5 | 50 | Prior visits first minus Risk-only | −7.00 |
| Admission | Annual ranking | 5 | 100 | Simple logistic minus Risk-only | −2.70 |
| Admission | Annual ranking | 5 | 100 | Age first minus Risk-only | −19.59 |
| Admission | Annual ranking | 5 | 100 | Prior visits first minus Risk-only | −9.12 |
| Admission | Annual ranking | 10 | 25 | Simple logistic minus Risk-only | −0.63 |
| Admission | Annual ranking | 10 | 25 | Age first minus Risk-only | −6.75 |
| Admission | Annual ranking | 10 | 25 | Prior visits first minus Risk-only | −1.81 |
| Admission | Annual ranking | 10 | 50 | Simple logistic minus Risk-only | −0.09 |
| Admission | Annual ranking | 10 | 50 | Age first minus Risk-only | −9.72 |
| Admission | Annual ranking | 10 | 50 | Prior visits first minus Risk-only | −3.64 |
| Admission | Annual ranking | 10 | 100 | Simple logistic minus Risk-only | −2.52 |
| Admission | Annual ranking | 10 | 100 | Age first minus Risk-only | −14.28 |
| Admission | Annual ranking | 10 | 100 | Prior visits first minus Risk-only | −6.73 |
| Admission | Annual ranking | 15 | 25 | Simple logistic minus Risk-only | −0.81 |
| Admission | Annual ranking | 15 | 25 | Age first minus Risk-only | −5.16 |
| Admission | Annual ranking | 15 | 25 | Prior visits first minus Risk-only | −1.48 |
| Admission | Annual ranking | 15 | 50 | Simple logistic minus Risk-only | −0.45 |
| Admission | Annual ranking | 15 | 50 | Age first minus Risk-only | −7.32 |
| Admission | Annual ranking | 15 | 50 | Prior visits first minus Risk-only | −2.56 |
| Admission | Annual ranking | 15 | 100 | Simple logistic minus Risk-only | −0.27 |
| Admission | Annual ranking | 15 | 100 | Age first minus Risk-only | −10.85 |
| Admission | Annual ranking | 15 | 100 | Prior visits first minus Risk-only | −4.61 |
| Discharge | Full-day ranking | 5 | 25 | Simple logistic minus Risk-only | −5.81 |
| Discharge | Full-day ranking | 5 | 25 | Age first minus Risk-only | −16.45 |
| Discharge | Full-day ranking | 5 | 25 | Prior visits first minus Risk-only | −4.76 |
| Discharge | Full-day ranking | 5 | 50 | Simple logistic minus Risk-only | −7.77 |
| Discharge | Full-day ranking | 5 | 50 | Age first minus Risk-only | −23.45 |
| Discharge | Full-day ranking | 5 | 50 | Prior visits first minus Risk-only | −8.08 |
| Discharge | Full-day ranking | 5 | 100 | Simple logistic minus Risk-only | −11.14 |
| Discharge | Full-day ranking | 5 | 100 | Age first minus Risk-only | −26.45 |
| Discharge | Full-day ranking | 5 | 100 | Prior visits first minus Risk-only | −11.65 |
| Discharge | Full-day ranking | 10 | 25 | Simple logistic minus Risk-only | −5.06 |
| Discharge | Full-day ranking | 10 | 25 | Age first minus Risk-only | −11.92 |
| Discharge | Full-day ranking | 10 | 25 | Prior visits first minus Risk-only | −5.83 |
| Discharge | Full-day ranking | 10 | 50 | Simple logistic minus Risk-only | −5.24 |
| Discharge | Full-day ranking | 10 | 50 | Age first minus Risk-only | −15.26 |
| Discharge | Full-day ranking | 10 | 50 | Prior visits first minus Risk-only | −4.37 |
| Discharge | Full-day ranking | 10 | 100 | Simple logistic minus Risk-only | −6.65 |
| Discharge | Full-day ranking | 10 | 100 | Age first minus Risk-only | −21.83 |
| Discharge | Full-day ranking | 10 | 100 | Prior visits first minus Risk-only | −6.87 |
| Discharge | Full-day ranking | 15 | 25 | Simple logistic minus Risk-only | −4.21 |
| Discharge | Full-day ranking | 15 | 25 | Age first minus Risk-only | −8.46 |
| Discharge | Full-day ranking | 15 | 25 | Prior visits first minus Risk-only | −4.97 |
| Discharge | Full-day ranking | 15 | 50 | Simple logistic minus Risk-only | −5.99 |
| Discharge | Full-day ranking | 15 | 50 | Age first minus Risk-only | −13.01 |
| Discharge | Full-day ranking | 15 | 50 | Prior visits first minus Risk-only | −7.17 |
| Discharge | Full-day ranking | 15 | 100 | Simple logistic minus Risk-only | −5.81 |
| Discharge | Full-day ranking | 15 | 100 | Age first minus Risk-only | −16.33 |
| Discharge | Full-day ranking | 15 | 100 | Prior visits first minus Risk-only | −5.56 |
| Discharge | Annual ranking | 5 | 25 | Simple logistic minus Risk-only | −8.43 |
| Discharge | Annual ranking | 5 | 25 | Age first minus Risk-only | −17.98 |
| Discharge | Annual ranking | 5 | 25 | Prior visits first minus Risk-only | −6.47 |
| Discharge | Annual ranking | 5 | 50 | Simple logistic minus Risk-only | −9.36 |
| Discharge | Annual ranking | 5 | 50 | Age first minus Risk-only | −23.65 |
| Discharge | Annual ranking | 5 | 50 | Prior visits first minus Risk-only | −7.88 |
| Discharge | Annual ranking | 5 | 100 | Simple logistic minus Risk-only | −11.99 |
| Discharge | Annual ranking | 5 | 100 | Age first minus Risk-only | −26.14 |
| Discharge | Annual ranking | 5 | 100 | Prior visits first minus Risk-only | −11.64 |
| Discharge | Annual ranking | 10 | 25 | Simple logistic minus Risk-only | −3.00 |
| Discharge | Annual ranking | 10 | 25 | Age first minus Risk-only | −10.83 |
| Discharge | Annual ranking | 10 | 25 | Prior visits first minus Risk-only | −3.89 |
| Discharge | Annual ranking | 10 | 50 | Simple logistic minus Risk-only | −7.40 |
| Discharge | Annual ranking | 10 | 50 | Age first minus Risk-only | −16.69 |
| Discharge | Annual ranking | 10 | 50 | Prior visits first minus Risk-only | −5.82 |
| Discharge | Annual ranking | 10 | 100 | Simple logistic minus Risk-only | −8.05 |
| Discharge | Annual ranking | 10 | 100 | Age first minus Risk-only | −21.90 |
| Discharge | Annual ranking | 10 | 100 | Prior visits first minus Risk-only | −6.51 |
| Discharge | Annual ranking | 15 | 25 | Simple logistic minus Risk-only | −2.06 |
| Discharge | Annual ranking | 15 | 25 | Age first minus Risk-only | −7.74 |
| Discharge | Annual ranking | 15 | 25 | Prior visits first minus Risk-only | −3.33 |
| Discharge | Annual ranking | 15 | 50 | Simple logistic minus Risk-only | −5.15 |
| Discharge | Annual ranking | 15 | 50 | Age first minus Risk-only | −12.20 |
| Discharge | Annual ranking | 15 | 50 | Prior visits first minus Risk-only | −4.96 |
| Discharge | Annual ranking | 15 | 100 | Simple logistic minus Risk-only | −6.84 |
| Discharge | Annual ranking | 15 | 100 | Age first minus Risk-only | −17.11 |
| Discharge | Annual ranking | 15 | 100 | Prior visits first minus Risk-only | −6.27 |

Paired point differences in event reach, in percentage points. No confidence intervals are shown. Event reach is the proportion of all composite-event encounters selected with sufficient recorded review time. Review attempts without enough recorded time still count against the budget. Reach does not measure events prevented. Simple logistic indicates the five-variable logistic model. ESI indicates Emergency Severity Index.

| eTable 49. Risk-Only Capacity and Selection-Order Comparisons in 2023 | | | | | | | |
| --- | --- | --- | --- | --- | --- | --- | --- |
| Decision group | Rule | Planned reviews | Reviews used | Unused reviews | Events reached | Events per review % | Event reach % |
| Admission | Constant daily capacity | 1,095 | 1,095 | 0 | 148 | 13.5 | 13.31 |
| Admission | Proportional daily quotas | 1,220 | 1,220 | 0 | 160 | 13.1 | 14.39 |
| Admission | Sequential threshold | 1,095 | 890 | 205 | 113 | 12.7 | 10.16 |
| Discharge | Constant daily capacity | 2,190 | 2,190 | 0 | 238 | 10.9 | 22.28 |
| Discharge | Proportional daily quotas | 2,415 | 2,415 | 0 | 253 | 10.5 | 23.69 |
| Discharge | Sequential threshold | 2,190 | 1,857 | 333 | 207 | 11.1 | 19.38 |

This separate benchmark used a linear-quantile risk threshold without the main sequential simulation's seeded score tiebreak. Comparison of full-day ranking with sequential Risk-only selection, using a 2022 95th-percentile threshold. Constant capacities are 3 admission and 6 discharge reviews daily. Proportional daily quotas total 1,220 and 2,415 attempts; constant annual capacities are 1,095 and 2,190. Event reach is the proportion of all composite-event encounters selected with sufficient recorded review time. Review attempts without enough recorded time still count against the budget. Reach does not measure events prevented.

| eTable 50. Risk-Only Selection by Daily Demand in 2023 | | | | | | | |
| --- | --- | --- | --- | --- | --- | --- | --- |
| Decision group | Day category | Days | Ranking reviews | Sequential reviews | Ranking events | Sequential events | Net fewer events |
| Admission | Fewer flags than capacity | 135 | 405 | 200 | 60 | 40 | 20 |
| Admission | Flags equal capacity | 84 | 252 | 252 | 25 | 25 | 0 |
| Admission | More flags than capacity | 146 | 438 | 438 | 63 | 48 | 15 |
| Discharge | Fewer flags than capacity | 141 | 846 | 513 | 76 | 58 | 18 |
| Discharge | Flags equal capacity | 52 | 312 | 312 | 41 | 41 | 0 |
| Discharge | More flags than capacity | 172 | 1,032 | 1,032 | 121 | 108 | 13 |

This separate benchmark used a linear-quantile risk threshold without the main sequential simulation's seeded score tiebreak. Comparison of constant-capacity full-day risk ranking and sequential threshold selection. Day categories compare flags with daily capacity. These are retrospective selection differences, not causal clinical effects.

| eTable 51. Recorded Windows and Outcome Association in 2023 | | | | |
| --- | --- | --- | --- | --- |
| Decision group | Cap (min) | Decision-time group | Copula rho (95% CI) | At cap (%) |
| Admission | 60 | All recorded decisions | 0.042 (0.007 to 0.076) | 69.5 |
| Admission | 30 | All recorded decisions | 0.025 (−0.018 to 0.070) | 85.3 |
| Admission | 120 | All recorded decisions | 0.032 (0.002 to 0.063) | 45.5 |
| Admission | 60 | Attending admission plan | 0.031 (−0.007 to 0.072) | 68.7 |
| Admission | 60 | Simultaneous admission plan codes | — | — |
| Admission | 60 | Resident admission plan | 0.061 (−0.008 to 0.136) | 72.2 |
| Discharge | 60 | All recorded decisions | 0.066 (0.036 to 0.097) | 14.5 |
| Discharge | 30 | All recorded decisions | 0.047 (0.015 to 0.079) | 36.2 |
| Discharge | 120 | All recorded decisions | 0.078 (0.047 to 0.110) | 5.4 |
| Discharge | 60 | Conditional discharge | 0.119 (0.061 to 0.175) | 36.5 |
| Discharge | 60 | Simultaneous discharge decision codes | 0.244 (0.123 to 0.371) | 28.7 |
| Discharge | 60 | Ready for discharge | 0.026 (−0.009 to 0.062) | 7.4 |

Parentheses contain 95% patient-cluster percentile confidence intervals from 2,000 resamples; at least 1,900 finite estimates were required. Fitted prediction models remained fixed. Gaussian copula with a binary outcome and mixed window margin, including zero and cap masses. Positive rho indicates association with longer windows, not a causal or tail-dependence parameter. Source strata with fewer than 20 events are not reported.

| eTable 52. Copula Independence Tests in 2023 | | | | |
| --- | --- | --- | --- | --- |
| Decision group | Copula rho | Bootstrap SE | Two-sided P | Holm-adjusted P |
| Admission | 0.042 | 0.018 | .02 | .02 |
| Discharge | 0.066 | 0.015 | <.001 | <.001 |

Two-sided Wald tests used bootstrap standard errors. Holm adjustment covers the two 60-minute-cap tests. Association parameters are model-dependent, not causal.

| eTable 53. Spline and Copula Descriptions of Outcome Association in 2023 | | | | | |
| --- | --- | --- | --- | --- | --- |
| Decision group | Cap (min) | Copula log likelihood gain | Spline log likelihood gain | Spline event risk at 15 min (%) | Spline event risk at cap (%) |
| Admission | 60 | 2.923 | 4.916 | 4.02 (3.05 to 5.05) | 4.79 (4.44 to 5.13) |
| Admission | 30 | 0.712 | 2.964 | 4.41 (3.09 to 5.92) | 4.65 (4.34 to 4.96) |
| Admission | 120 | 2.172 | 5.192 | 4.02 (3.41 to 4.65) | 4.81 (4.40 to 5.21) |
| Discharge | 60 | 13.971 | 72.031 | 1.36 (1.18 to 1.55) | 3.65 (3.21 to 4.10) |
| Discharge | 30 | 6.182 | 67.996 | 1.57 (1.31 to 1.84) | 2.81 (2.53 to 3.07) |
| Discharge | 120 | 19.451 | 63.126 | 1.81 (1.64 to 1.98) | 5.15 (4.28 to 6.06) |

Parentheses contain 95% confidence intervals. Both models were re-estimated within bootstrap samples. These describe fit in the 2023 data, not out-of-sample performance. Caps other than 60 minutes did not apply to allocation.

| eTable 54. Recorded Windows and Predicted Risk in 2023 | | |
| --- | --- | --- |
| Decision group | Measure | Estimate (95% CI) |
| Admission | 10-minute window: all encounters | 96.21 (95.96 to 96.44) |
| Admission | 10-minute window: event encounters | 96.13 (95.00 to 97.18) |
| Admission | 10-minute window: encounters without events | 96.21 (95.96 to 96.45) |
| Admission | 15-minute window: all encounters | 93.86 (93.56 to 94.16) |
| Admission | 15-minute window: event encounters | 93.62 (92.06 to 95.01) |
| Admission | 15-minute window: encounters without events | 93.87 (93.56 to 94.18) |
| Admission | 5-minute window: all encounters | 98.02 (97.84 to 98.20) |
| Admission | 5-minute window: event encounters | 97.84 (96.98 to 98.63) |
| Admission | 5-minute window: encounters without events | 98.03 (97.85 to 98.21) |
| Discharge | 10-minute window: all encounters | 85.37 (85.03 to 85.72) |
| Discharge | 10-minute window: event encounters | 83.99 (81.63 to 86.26) |
| Discharge | 10-minute window: encounters without events | 85.40 (85.06 to 85.74) |
| Discharge | 15-minute window: all encounters | 70.06 (69.62 to 70.48) |
| Discharge | 15-minute window: event encounters | 74.44 (71.71 to 77.13) |
| Discharge | 15-minute window: encounters without events | 69.96 (69.53 to 70.39) |
| Discharge | 5-minute window: all encounters | 95.51 (95.30 to 95.72) |
| Discharge | 5-minute window: event encounters | 91.29 (89.36 to 93.21) |
| Discharge | 5-minute window: encounters without events | 95.61 (95.39 to 95.82) |
| Admission | High-risk minus lower-risk mean window (min) | 4.52 (3.92 to 5.05) |
| Admission | High-risk minus lower-risk 10-minute reviewability (pp) | 2.33 (1.73 to 2.90) |
| Admission | High-risk minus lower-risk 15-minute reviewability (pp) | 3.85 (3.10 to 4.55) |
| Admission | High-risk minus lower-risk 5-minute reviewability (pp) | 1.16 (0.71 to 1.57) |
| Admission | Risk-window Spearman correlation | 0.17 (0.16 to 0.19) |
| Discharge | High-risk minus lower-risk mean window (min) | 8.40 (7.56 to 9.24) |
| Discharge | High-risk minus lower-risk 10-minute reviewability (pp) | 6.06 (4.99 to 7.06) |
| Discharge | High-risk minus lower-risk 15-minute reviewability (pp) | 10.88 (9.46 to 12.21) |
| Discharge | High-risk minus lower-risk 5-minute reviewability (pp) | 0.95 (0.14 to 1.66) |
| Discharge | Risk-window Spearman correlation | 0.18 (0.17 to 0.19) |

Parentheses contain 95% patient-cluster percentile confidence intervals from 2,000 resamples; at least 1,900 finite estimates were required. Fitted prediction models remained fixed. High risk denotes a score at or above the 2022 90th percentile. Differences compare high-risk with other encounters. Mean-window differences are minutes; sufficiency differences are percentage points. Spearman correlation is unitless.

| eTable 55. Case Mix by Decision-Time Group | | | | | | | | |
| --- | --- | --- | --- | --- | --- | --- | --- | --- |
| Characteristic | Category | Discharge decision | Admission plan with recorded handoff | Admission plan without recorded handoff | Handoff without earlier plan | Conflicting directions | No qualifying status | Other invalid timing |
| Age group | 18-39 | 78,899 (43.9%) | 12,829 (15.3%) | 574 (52.9%) | 3,247 (23.6%) | 257 (32.5%) | 2,895 (52.0%) | — |
| Age group | 40-64 | 65,339 (36.3%) | 28,286 (33.8%) | 226 (20.8%) | 5,219 (37.9%) | 297 (37.6%) | 1,808 (32.5%) | — |
| Age group | 65-84 | 29,633 (16.5%) | 31,855 (38.1%) | 186 (17.1%) | 4,317 (31.3%) | 180 (22.8%) | 683 (12.3%) | — |
| Age group | 85-100 | 5,927 (3.3%) | 10,624 (12.7%) | 99 (9.1%) | 992 (7.2%) | 56 (7.1%) | 177 (3.2%) | — |
| Arrival mode | Ambulance, American Medical Response | 14,668 (8.2%) | 15,010 (18.0%) | 220 (20.3%) | 1,846 (13.4%) | 88 (11.1%) | 581 (10.4%) | 11 (50.0%) |
| Arrival mode | Ambulance, Palo Alto Paramedics | 3,924 (2.2%) | 4,236 (5.1%) | 90 (8.3%) | 491 (3.6%) | 26 (3.3%) | 195 (3.5%) | — |
| Arrival mode | Helicopter, Calstar | — | 37 (0.0%) | — | — | — | — | 0 (0.0%) |
| Arrival mode | Helicopter, LifeFlight | 14 (0.0%) | 131 (0.2%) | — | 29 (0.2%) | 0 (0.0%) | — | 0 (0.0%) |
| Arrival mode | Ambulance, Bayshore | — | — | 0 (0.0%) | — | 0 (0.0%) | 0 (0.0%) | 0 (0.0%) |
| Arrival mode | Ambulance, Lifestar | 0 (0.0%) | — | 0 (0.0%) | — | 0 (0.0%) | 0 (0.0%) | 0 (0.0%) |
| Arrival mode | Police, Other | 40 (0.0%) | — | — | — | — | — | 0 (0.0%) |
| Arrival mode | Ambulance, Other Service | 191 (0.1%) | 284 (0.3%) | — | 34 (0.2%) | 0 (0.0%) | — | 0 (0.0%) |
| Arrival mode | Helicopter, Other Service | — | 28 (0.0%) | 0 (0.0%) | — | 0 (0.0%) | 0 (0.0%) | 0 (0.0%) |
| Arrival mode | Other Mode of Arrival | 23 (0.0%) | 28 (0.0%) | 0 (0.0%) | — | 0 (0.0%) | 0 (0.0%) | 0 (0.0%) |
| Arrival mode | Ambulance, Golden Gate | — | 0 (0.0%) | 0 (0.0%) | 0 (0.0%) | 0 (0.0%) | 0 (0.0%) | 0 (0.0%) |
| Arrival mode | Ambulance, Silicon Valley | 374 (0.2%) | 576 (0.7%) | — | 69 (0.5%) | — | — | 0 (0.0%) |
| Arrival mode | Ambulance, Westmed | 66 (0.0%) | 139 (0.2%) | — | 19 (0.1%) | — | — | 0 (0.0%) |
| Arrival mode | Police, California Highway Patrol | — | — | 0 (0.0%) | — | 0 (0.0%) | 0 (0.0%) | 0 (0.0%) |
| Arrival mode | Police, East Palo Alto | 17 (0.0%) | — | — | 0 (0.0%) | 0 (0.0%) | 0 (0.0%) | 0 (0.0%) |
| Arrival mode | Police, Palo Alto | 46 (0.0%) | 24 (0.0%) | — | 14 (0.1%) | — | — | 0 (0.0%) |
| Arrival mode | Police, Stanford Campus | 11 (0.0%) | — | — | — | 0 (0.0%) | — | 0 (0.0%) |
| Arrival mode | Ambulance, Rural Metro | 715 (0.4%) | 842 (1.0%) | — | 90 (0.7%) | — | 21 (0.4%) | 0 (0.0%) |
| Arrival mode | Ambulance, Royal | 475 (0.3%) | 880 (1.1%) | — | 117 (0.8%) | — | — | 0 (0.0%) |
| Arrival mode | Ambulance, Falck | 52 (0.0%) | 88 (0.1%) | — | 13 (0.1%) | — | — | 0 (0.0%) |
| Arrival mode | Ambulance, ParamedicPlus | 12 (0.0%) | 12 (0.0%) | — | — | 0 (0.0%) | — | 0 (0.0%) |
| Arrival mode | Ambulance, Norcal | 78 (0.0%) | 55 (0.1%) | 0 (0.0%) | 16 (0.1%) | 0 (0.0%) | — | 0 (0.0%) |
| Arrival mode | Ambulance, Falcon | — | 19 (0.0%) | — | — | 0 (0.0%) | — | 0 (0.0%) |
| Arrival mode | Ambulance, Protransport | 24 (0.0%) | 29 (0.0%) | 0 (0.0%) | 11 (0.1%) | 0 (0.0%) | 0 (0.0%) | 0 (0.0%) |
| Arrival mode | Ambulance, Woodside Fire Dept | 734 (0.4%) | 896 (1.1%) | 24 (2.2%) | 126 (0.9%) | — | 38 (0.7%) | 0 (0.0%) |
| Arrival mode | Ambulance | 1,386 (0.8%) | 1,785 (2.1%) | 28 (2.6%) | 259 (1.9%) | — | 60 (1.1%) | — |
| Arrival mode | Helicopter | — | 56 (0.1%) | — | 17 (0.1%) | 0 (0.0%) | 0 (0.0%) | 0 (0.0%) |
| Arrival mode | Fixed Wing | — | 22 (0.0%) | 0 (0.0%) | — | 0 (0.0%) | 0 (0.0%) | 0 (0.0%) |
| Arrival mode | Private Vehicle | — | 11 (0.0%) | 0 (0.0%) | — | 0 (0.0%) | 0 (0.0%) | 0 (0.0%) |
| Arrival mode | Air - fixed wing and roto wing | — | — | 0 (0.0%) | — | 0 (0.0%) | 0 (0.0%) | 0 (0.0%) |
| Arrival mode | TeleHealth | 106 (0.1%) | 54 (0.1%) | 0 (0.0%) | — | — | — | 0 (0.0%) |
| Arrival mode | Police | — | — | 0 (0.0%) | — | 0 (0.0%) | — | 0 (0.0%) |
| Arrival mode | Car/bus | 154,649 (86.0%) | 57,550 (68.8%) | 667 (61.5%) | 10,389 (75.4%) | 638 (80.8%) | 4,358 (78.3%) | — |
| Arrival mode | Walk | 2,072 (1.2%) | 652 (0.8%) | 14 (1.3%) | 152 (1.1%) | — | 92 (1.7%) | 0 (0.0%) |
| Arrival mode | Missing | 73 (0.0%) | 119 (0.1%) | — | 26 (0.2%) | — | 164 (2.9%) | — |
| ESI | 1 | 157 (0.1%) | 1,610 (1.9%) | 47 (4.3%) | 320 (2.3%) | — | 157 (2.8%) | — |
| ESI | 2 | 22,604 (12.6%) | 33,261 (39.8%) | 588 (54.2%) | 4,671 (33.9%) | 172 (21.8%) | 872 (15.7%) | — |
| ESI | 3 | 106,797 (59.4%) | 47,314 (56.6%) | 433 (39.9%) | 8,415 (61.1%) | 477 (60.4%) | 2,316 (41.6%) | — |
| ESI | 4 | 45,820 (25.5%) | 918 (1.1%) | — | 290 (2.1%) | 117 (14.8%) | 942 (16.9%) | 0 (0.0%) |
| ESI | 5 | 3,048 (1.7%) | 40 (0.0%) | 0 (0.0%) | 13 (0.1%) | 13 (1.6%) | 107 (1.9%) | 0 (0.0%) |
| ESI | Missing | 1,372 (0.8%) | 451 (0.5%) | — | 66 (0.5%) | — | 1,169 (21.0%) | 16 (72.7%) |
| Final disposition | Admit | 1,123 (0.6%) | 73,181 (87.5%) | 51 (4.7%) | 10,925 (79.3%) | 240 (30.4%) | 25 (0.4%) | — |
| Final disposition | Discharge | 173,895 (96.7%) | 9,156 (11.0%) | 32 (2.9%) | 2,475 (18.0%) | 546 (69.1%) | 693 (12.5%) | — |
| Final disposition | Error | 59 (0.0%) | 0 (0.0%) | — | — | 0 (0.0%) | 406 (7.3%) | — |
| Final disposition | Missing | 22 (0.0%) | 166 (0.2%) | 32 (2.9%) | 65 (0.5%) | — | 930 (16.7%) | — |
| Final disposition | Other | 4,558 (2.5%) | 282 (0.3%) | 83 (7.6%) | — | — | 2,210 (39.7%) | — |
| Final disposition | Transfer | 83 (0.0%) | 186 (0.2%) | 877 (80.8%) | 99 (0.7%) | 0 (0.0%) | 1,240 (22.3%) | 0 (0.0%) |
| Final disposition | Unmapped | 58 (0.0%) | 623 (0.7%) | — | 149 (1.1%) | — | 59 (1.1%) | 0 (0.0%) |
| Interpreter need | Missing | 152 (0.1%) | 18 (0.0%) | — | — | 0 (0.0%) | 234 (4.2%) | 0 (0.0%) |
| Interpreter need | Not required | 149,013 (82.9%) | 68,579 (82.0%) | 968 (89.2%) | 11,639 (84.5%) | 631 (79.9%) | 4,478 (80.5%) | — |
| Interpreter need | Required | 30,633 (17.0%) | 14,997 (17.9%) | — | — | 159 (20.1%) | 851 (15.3%) | — |
| Language | Albanian | — | — | 0 (0.0%) | 0 (0.0%) | 0 (0.0%) | 0 (0.0%) | 0 (0.0%) |
| Language | American Sign Language | 84 (0.0%) | 48 (0.1%) | 0 (0.0%) | 11 (0.1%) | 0 (0.0%) | — | 0 (0.0%) |
| Language | Amharic | 23 (0.0%) | 18 (0.0%) | 0 (0.0%) | — | 0 (0.0%) | — | 0 (0.0%) |
| Language | Arabic | 367 (0.2%) | 218 (0.3%) | — | 28 (0.2%) | — | 12 (0.2%) | 0 (0.0%) |
| Language | Armenian | 20 (0.0%) | 19 (0.0%) | 0 (0.0%) | — | 0 (0.0%) | — | 0 (0.0%) |
| Language | Assyrian | — | 14 (0.0%) | 0 (0.0%) | 0 (0.0%) | 0 (0.0%) | 0 (0.0%) | 0 (0.0%) |
| Language | Azerbaijani | — | — | 0 (0.0%) | — | 0 (0.0%) | 0 (0.0%) | 0 (0.0%) |
| Language | Bengali | — | — | 0 (0.0%) | — | 0 (0.0%) | 0 (0.0%) | 0 (0.0%) |
| Language | Bosnian | 12 (0.0%) | — | 0 (0.0%) | 0 (0.0%) | 0 (0.0%) | — | 0 (0.0%) |
| Language | Bulgarian | — | — | 0 (0.0%) | — | 0 (0.0%) | 0 (0.0%) | 0 (0.0%) |
| Language | Burmese | 45 (0.0%) | 89 (0.1%) | 0 (0.0%) | — | — | 0 (0.0%) | 0 (0.0%) |
| Language | Cantonese | 385 (0.2%) | 671 (0.8%) | — | 61 (0.4%) | — | — | 0 (0.0%) |
| Language | Chinese | 28 (0.0%) | 22 (0.0%) | 0 (0.0%) | — | — | 0 (0.0%) | 0 (0.0%) |
| Language | Croatian | — | — | 0 (0.0%) | 0 (0.0%) | 0 (0.0%) | — | 0 (0.0%) |
| Language | Czech | — | — | 0 (0.0%) | — | 0 (0.0%) | 0 (0.0%) | 0 (0.0%) |
| Language | Dari | 177 (0.1%) | 234 (0.3%) | 0 (0.0%) | 25 (0.2%) | — | — | 0 (0.0%) |
| Language | Deaf/Non-Sign Language | 29 (0.0%) | 42 (0.1%) | 0 (0.0%) | — | 0 (0.0%) | — | 0 (0.0%) |
| Language | Dutch | — | — | 0 (0.0%) | 0 (0.0%) | 0 (0.0%) | 0 (0.0%) | 0 (0.0%) |
| Language | English | 145,266 (80.8%) | 66,666 (79.7%) | 949 (87.5%) | 11,354 (82.4%) | 613 (77.6%) | 4,391 (78.9%) | 18 (81.8%) |
| Language | Farsi | 624 (0.3%) | 496 (0.6%) | 0 (0.0%) | 52 (0.4%) | — | 15 (0.3%) | 0 (0.0%) |
| Language | Fijian | — | 11 (0.0%) | 0 (0.0%) | 0 (0.0%) | 0 (0.0%) | 0 (0.0%) | 0 (0.0%) |
| Language | Finnish | — | 0 (0.0%) | 0 (0.0%) | 0 (0.0%) | 0 (0.0%) | 0 (0.0%) | 0 (0.0%) |
| Language | French | 48 (0.0%) | 37 (0.0%) | 0 (0.0%) | — | 0 (0.0%) | 0 (0.0%) | 0 (0.0%) |
| Language | French (Canadian) | — | 0 (0.0%) | 0 (0.0%) | 0 (0.0%) | 0 (0.0%) | 0 (0.0%) | 0 (0.0%) |
| Language | Gaelic | — | 0 (0.0%) | 0 (0.0%) | 0 (0.0%) | 0 (0.0%) | 0 (0.0%) | 0 (0.0%) |
| Language | Georgian | — | 0 (0.0%) | 0 (0.0%) | 0 (0.0%) | 0 (0.0%) | 0 (0.0%) | 0 (0.0%) |
| Language | German | 19 (0.0%) | — | 0 (0.0%) | — | 0 (0.0%) | — | 0 (0.0%) |
| Language | Greek | 14 (0.0%) | 16 (0.0%) | 0 (0.0%) | 0 (0.0%) | 0 (0.0%) | — | 0 (0.0%) |
| Language | Gujarati | 27 (0.0%) | 27 (0.0%) | — | — | 0 (0.0%) | 0 (0.0%) | 0 (0.0%) |
| Language | Haitian Creole | — | — | 0 (0.0%) | 0 (0.0%) | 0 (0.0%) | — | 0 (0.0%) |
| Language | Hebrew | 16 (0.0%) | — | 0 (0.0%) | — | — | — | 0 (0.0%) |
| Language | Hindi | 166 (0.1%) | 228 (0.3%) | — | 26 (0.2%) | — | — | 0 (0.0%) |
| Language | Hmong | — | — | 0 (0.0%) | 0 (0.0%) | 0 (0.0%) | 0 (0.0%) | 0 (0.0%) |
| Language | Hungarian | 11 (0.0%) | — | 0 (0.0%) | — | 0 (0.0%) | 0 (0.0%) | 0 (0.0%) |
| Language | Ilocano | 16 (0.0%) | 20 (0.0%) | 0 (0.0%) | — | 0 (0.0%) | — | 0 (0.0%) |
| Language | Indonesian | — | — | 0 (0.0%) | — | 0 (0.0%) | — | 0 (0.0%) |
| Language | Italian | 30 (0.0%) | 18 (0.0%) | 0 (0.0%) | — | 0 (0.0%) | 0 (0.0%) | 0 (0.0%) |
| Language | Japanese | 110 (0.1%) | 95 (0.1%) | 0 (0.0%) | 17 (0.1%) | — | — | 0 (0.0%) |
| Language | Kannada | — | — | 0 (0.0%) | — | 0 (0.0%) | 0 (0.0%) | 0 (0.0%) |
| Language | Khmer | 23 (0.0%) | 32 (0.0%) | 0 (0.0%) | — | 0 (0.0%) | 0 (0.0%) | 0 (0.0%) |
| Language | Korean | 299 (0.2%) | 306 (0.4%) | — | 34 (0.2%) | — | — | 0 (0.0%) |
| Language | Laotian | — | — | 0 (0.0%) | 0 (0.0%) | 0 (0.0%) | 0 (0.0%) | 0 (0.0%) |
| Language | Macedonian | — | — | 0 (0.0%) | 0 (0.0%) | 0 (0.0%) | 0 (0.0%) | 0 (0.0%) |
| Language | Malayalam | 0 (0.0%) | — | 0 (0.0%) | — | 0 (0.0%) | 0 (0.0%) | 0 (0.0%) |
| Language | Mam | — | 0 (0.0%) | 0 (0.0%) | 0 (0.0%) | 0 (0.0%) | 0 (0.0%) | 0 (0.0%) |
| Language | Mandarin | 1,871 (1.0%) | 1,824 (2.2%) | — | 240 (1.7%) | 16 (2.0%) | 37 (0.7%) | — |
| Language | Marathi | — | — | 0 (0.0%) | — | 0 (0.0%) | 0 (0.0%) | 0 (0.0%) |
| Language | Maya-Quiche | — | — | 0 (0.0%) | 0 (0.0%) | 0 (0.0%) | 0 (0.0%) | 0 (0.0%) |
| Language | Missing | 142 (0.1%) | 17 (0.0%) | — | — | 0 (0.0%) | 231 (4.2%) | 0 (0.0%) |
| Language | Mixteco Alto | — | — | 0 (0.0%) | — | 0 (0.0%) | 0 (0.0%) | 0 (0.0%) |
| Language | Mixteco Bajo | — | — | 0 (0.0%) | 0 (0.0%) | 0 (0.0%) | 0 (0.0%) | 0 (0.0%) |
| Language | Mongolian | — | — | 0 (0.0%) | 0 (0.0%) | 0 (0.0%) | 0 (0.0%) | 0 (0.0%) |
| Language | Nepali | 16 (0.0%) | 18 (0.0%) | 0 (0.0%) | — | 0 (0.0%) | — | 0 (0.0%) |
| Language | Non-Verbal | — | — | 0 (0.0%) | — | 0 (0.0%) | 0 (0.0%) | 0 (0.0%) |
| Language | Norwegian | — | 0 (0.0%) | 0 (0.0%) | 0 (0.0%) | 0 (0.0%) | 0 (0.0%) | 0 (0.0%) |
| Language | Oriya | 0 (0.0%) | 0 (0.0%) | 0 (0.0%) | — | 0 (0.0%) | 0 (0.0%) | 0 (0.0%) |
| Language | Other | 138 (0.1%) | 90 (0.1%) | — | 14 (0.1%) | — | — | 0 (0.0%) |
| Language | Pacific Island Languages (E.G., Samoan, Tongan) | 0 (0.0%) | — | 0 (0.0%) | 0 (0.0%) | 0 (0.0%) | 0 (0.0%) | 0 (0.0%) |
| Language | Pangasinan | 0 (0.0%) | 0 (0.0%) | 0 (0.0%) | — | 0 (0.0%) | 0 (0.0%) | 0 (0.0%) |
| Language | Pashto | — | 12 (0.0%) | 0 (0.0%) | — | 0 (0.0%) | 0 (0.0%) | 0 (0.0%) |
| Language | Polish | 11 (0.0%) | — | 0 (0.0%) | 0 (0.0%) | 0 (0.0%) | 0 (0.0%) | 0 (0.0%) |
| Language | Portuguese | 114 (0.1%) | 33 (0.0%) | — | — | 0 (0.0%) | — | 0 (0.0%) |
| Language | Portuguese (Brazilian) | 222 (0.1%) | 50 (0.1%) | 0 (0.0%) | 12 (0.1%) | — | — | 0 (0.0%) |
| Language | Portuguese (European) | 15 (0.0%) | 28 (0.0%) | 0 (0.0%) | — | 0 (0.0%) | — | 0 (0.0%) |
| Language | Punjabi (Panjabi) | 116 (0.1%) | 148 (0.2%) | 0 (0.0%) | 24 (0.2%) | — | — | 0 (0.0%) |
| Language | Romanian | 18 (0.0%) | 22 (0.0%) | 0 (0.0%) | — | 0 (0.0%) | 0 (0.0%) | 0 (0.0%) |
| Language | Russian | 505 (0.3%) | 574 (0.7%) | — | 59 (0.4%) | — | 17 (0.3%) | 0 (0.0%) |
| Language | Samar-Leyte | — | — | 0 (0.0%) | 0 (0.0%) | 0 (0.0%) | 0 (0.0%) | 0 (0.0%) |
| Language | Samoan | 30 (0.0%) | 28 (0.0%) | — | — | 0 (0.0%) | 0 (0.0%) | 0 (0.0%) |
| Language | Serbian | — | 11 (0.0%) | 0 (0.0%) | — | 0 (0.0%) | 0 (0.0%) | 0 (0.0%) |
| Language | Shanghainese | 0 (0.0%) | — | 0 (0.0%) | — | 0 (0.0%) | 0 (0.0%) | 0 (0.0%) |
| Language | Somali | — | — | 0 (0.0%) | 0 (0.0%) | 0 (0.0%) | 0 (0.0%) | 0 (0.0%) |
| Language | Spanish | 26,370 (14.7%) | 8,526 (10.2%) | 97 (8.9%) | 1,407 (10.2%) | 118 (14.9%) | 746 (13.4%) | — |
| Language | Swahili | — | — | 0 (0.0%) | 0 (0.0%) | 0 (0.0%) | 0 (0.0%) | 0 (0.0%) |
| Language | Swedish | — | — | 0 (0.0%) | 0 (0.0%) | 0 (0.0%) | 0 (0.0%) | 0 (0.0%) |
| Language | Tagalog | 205 (0.1%) | 403 (0.5%) | — | 40 (0.3%) | 0 (0.0%) | — | 0 (0.0%) |
| Language | Taiwanese | 15 (0.0%) | 15 (0.0%) | 0 (0.0%) | — | 0 (0.0%) | 0 (0.0%) | 0 (0.0%) |
| Language | Tamil | — | 12 (0.0%) | 0 (0.0%) | 0 (0.0%) | 0 (0.0%) | 0 (0.0%) | 0 (0.0%) |
| Language | Telugu | 14 (0.0%) | 15 (0.0%) | 0 (0.0%) | — | 0 (0.0%) | 0 (0.0%) | 0 (0.0%) |
| Language | Thai | 16 (0.0%) | — | — | — | 0 (0.0%) | — | 0 (0.0%) |
| Language | Tibetan | — | — | 0 (0.0%) | 0 (0.0%) | 0 (0.0%) | — | 0 (0.0%) |
| Language | Tigrinya | — | — | 0 (0.0%) | 0 (0.0%) | 0 (0.0%) | 0 (0.0%) | 0 (0.0%) |
| Language | Toisanese | — | — | 0 (0.0%) | 0 (0.0%) | — | 0 (0.0%) | 0 (0.0%) |
| Language | Tongan | 412 (0.2%) | 375 (0.4%) | — | 41 (0.3%) | — | — | — |
| Language | Traditional Chinese | — | — | 0 (0.0%) | 0 (0.0%) | 0 (0.0%) | 0 (0.0%) | 0 (0.0%) |
| Language | Triqui Alto | — | — | 0 (0.0%) | 0 (0.0%) | 0 (0.0%) | 0 (0.0%) | 0 (0.0%) |
| Language | Triqui Bajo | — | 0 (0.0%) | 0 (0.0%) | 0 (0.0%) | 0 (0.0%) | 0 (0.0%) | 0 (0.0%) |
| Language | Turkish | 64 (0.0%) | 16 (0.0%) | 0 (0.0%) | — | 0 (0.0%) | — | 0 (0.0%) |
| Language | Ukrainian | 24 (0.0%) | 33 (0.0%) | 0 (0.0%) | — | 0 (0.0%) | — | 0 (0.0%) |
| Language | Unable To Verify | 51 (0.0%) | 34 (0.0%) | 0 (0.0%) | — | 0 (0.0%) | — | 0 (0.0%) |
| Language | Urdu | 66 (0.0%) | 56 (0.1%) | 0 (0.0%) | — | 0 (0.0%) | — | 0 (0.0%) |
| Language | Uyghur | — | 0 (0.0%) | 0 (0.0%) | 0 (0.0%) | 0 (0.0%) | 0 (0.0%) | 0 (0.0%) |
| Language | Vietnamese | 1,339 (0.7%) | 1,771 (2.1%) | — | 209 (1.5%) | — | 14 (0.3%) | 0 (0.0%) |
| Language | Yoruba | 0 (0.0%) | — | 0 (0.0%) | 0 (0.0%) | 0 (0.0%) | 0 (0.0%) | 0 (0.0%) |
| Sex | Female | 99,803 (55.5%) | 42,246 (50.5%) | 682 (62.9%) | 7,094 (51.5%) | 448 (56.7%) | 3,319 (59.7%) | — |
| Sex | Male | 79,974 (44.5%) | — | — | — | — | 2,229 (40.1%) | — |
| Sex | Unknown | 21 (0.0%) | — | — | — | — | 15 (0.3%) | 0 (0.0%) |
| Arrival year | 2020 | 42,647 (23.7%) | 16,788 (20.1%) | 230 (21.2%) | 2,786 (20.2%) | — | 1,224 (22.0%) | — |
| Arrival year | 2021 | 42,908 (23.9%) | 19,772 (23.7%) | 239 (22.0%) | 3,336 (24.2%) | — | 1,539 (27.7%) | — |
| Arrival year | 2022 | 45,371 (25.2%) | 22,813 (27.3%) | 304 (28.0%) | 3,660 (26.6%) | — | 1,768 (31.8%) | — |
| Arrival year | 2023 | 48,872 (27.2%) | 24,221 (29.0%) | 312 (28.8%) | 3,993 (29.0%) | — | 1,032 (18.6%) | — |

All 2020–2023 candidates. Values are No. (%) within decision-time group, including missing categories. Final disposition does not determine decision-time group assignment; Admit includes observation and outpatient admission codes. Arrival modes are recorded electronic health record categories; ambulance categories are shown separately as recorded. Excluded groups have no assigned event rates. To protect patient privacy, counts of 1–10, and selected cells from which they could be calculated, are not shown (—).

| eTable 56. Other-Template Record Presence in 2023 | | |
| --- | --- | --- |
| Vital sign | Other template | Encounters without primary-template record |
| Diastolic blood pressure | Apheresis VS | — |
| Diastolic blood pressure | Blood Administration | — |
| Diastolic blood pressure | Complex Assessment | — |
| Diastolic blood pressure | Custom Formula Data | — |
| Diastolic blood pressure | Device Data Template | 408 |
| Diastolic blood pressure | Disposition Vitals | — |
| Diastolic blood pressure | HF Visit | — |
| Diastolic blood pressure | ICU VS | 355 |
| Diastolic blood pressure | IIC VS | 292 |
| Diastolic blood pressure | M/S VS | 388 |
| Diastolic blood pressure | MAR MINI-FLOWSHEET DATA | — |
| Diastolic blood pressure | NA Flowsheet | 402 |
| Diastolic blood pressure | Non-Violent Restraints | — |
| Diastolic blood pressure | RETIRED Infusion Center Assessment | — |
| Diastolic blood pressure | Stroke | 63 |
| Diastolic blood pressure | Violent Restraint Reporting | — |
| Diastolic blood pressure | Violent Restraints | — |
| Diastolic blood pressure | Vital Signs | 55 |
| Diastolic blood pressure | Vitals From Sheet | 16 |
| Heart rate | Advisory Flowsheet Action Data | — |
| Heart rate | Apheresis VS | — |
| Heart rate | Blood Administration | — |
| Heart rate | Custom Formula Data | — |
| Heart rate | Device Data Template | 411 |
| Heart rate | Disposition Vitals | — |
| Heart rate | HF Visit | — |
| Heart rate | ICU Intake & Output | — |
| Heart rate | ICU VS | 333 |
| Heart rate | IIC VS | 291 |
| Heart rate | M/S VS | 368 |
| Heart rate | MAR MINI-FLOWSHEET DATA | — |
| Heart rate | NA Flowsheet | 382 |
| Heart rate | Non-Violent Restraints | — |
| Heart rate | OT Short Form | — |
| Heart rate | RT Assess/ Treatment | 34 |
| Heart rate | Stroke | 61 |
| Heart rate | Violent Restraint Reporting | — |
| Heart rate | Violent Restraints | — |
| Heart rate | Vital Signs | 58 |
| Heart rate | Vitals From Sheet | 14 |
| Respiratory rate | Apheresis VS | — |
| Respiratory rate | BIPAP/CPAP | — |
| Respiratory rate | Blood Administration | — |
| Respiratory rate | Custom Formula Data | — |
| Respiratory rate | Device Data Template | 279 |
| Respiratory rate | Disposition Vitals | — |
| Respiratory rate | ICU VS | 360 |
| Respiratory rate | IIC VS | 287 |
| Respiratory rate | M/S VS | 408 |
| Respiratory rate | Mod Sedation | — |
| Respiratory rate | NA Flowsheet | 423 |
| Respiratory rate | Non-Violent Restraints | — |
| Respiratory rate | Primary Survey | 257 |
| Respiratory rate | RT Assess/ Treatment | 45 |
| Respiratory rate | Stroke | 65 |
| Respiratory rate | Ventilator | 13 |
| Respiratory rate | Violent Restraint Reporting | — |
| Respiratory rate | Violent Restraints | — |
| Respiratory rate | Vital Signs | 57 |
| Respiratory rate | Vitals From Sheet | 18 |
| Peripheral oxygen saturation | Advisory Flowsheet Action Data | — |
| Peripheral oxygen saturation | Apheresis VS | — |
| Peripheral oxygen saturation | Blood Administration | — |
| Peripheral oxygen saturation | Bronchoscopy | — |
| Peripheral oxygen saturation | Custom Formula Data | — |
| Peripheral oxygen saturation | Device Data Template | 449 |
| Peripheral oxygen saturation | Disposition Vitals | — |
| Peripheral oxygen saturation | ICU VS | 363 |
| Peripheral oxygen saturation | IIC VS | 298 |
| Peripheral oxygen saturation | M/S VS | 369 |
| Peripheral oxygen saturation | NA Flowsheet | 386 |
| Peripheral oxygen saturation | Non-Violent Restraints | — |
| Peripheral oxygen saturation | RT Assess/ Treatment | 43 |
| Peripheral oxygen saturation | Stroke | 68 |
| Peripheral oxygen saturation | Trial of Ambulation | — |
| Peripheral oxygen saturation | Ventilator | — |
| Peripheral oxygen saturation | Violent Restraint Reporting | — |
| Peripheral oxygen saturation | Violent Restraints | — |
| Peripheral oxygen saturation | Vital Signs | 59 |
| Peripheral oxygen saturation | Vitals From Sheet | 13 |
| Systolic blood pressure | Apheresis VS | — |
| Systolic blood pressure | Blood Administration | — |
| Systolic blood pressure | Complex Assessment | — |
| Systolic blood pressure | Custom Formula Data | — |
| Systolic blood pressure | Device Data Template | 408 |
| Systolic blood pressure | Disposition Vitals | — |
| Systolic blood pressure | HF Visit | — |
| Systolic blood pressure | ICU VS | 355 |
| Systolic blood pressure | IIC VS | 292 |
| Systolic blood pressure | M/S VS | 388 |
| Systolic blood pressure | MAR MINI-FLOWSHEET DATA | — |
| Systolic blood pressure | NA Flowsheet | 402 |
| Systolic blood pressure | Non-Violent Restraints | — |
| Systolic blood pressure | RETIRED Infusion Center Assessment | — |
| Systolic blood pressure | Stroke | 63 |
| Systolic blood pressure | Violent Restraint Reporting | — |
| Systolic blood pressure | Violent Restraints | — |
| Systolic blood pressure | Vital Signs | 55 |
| Systolic blood pressure | Vitals From Sheet | 16 |
| Temperature | Advanced Vitals | — |
| Temperature | Apheresis Transfusion | — |
| Temperature | Blood Administration | 16 |
| Temperature | Custom Formula Data | 12 |
| Temperature | Device Data Template | — |
| Temperature | Disposition Vitals | — |
| Temperature | ICU VS | 410 |
| Temperature | IIC VS | 391 |
| Temperature | M/S VS | 785 |
| Temperature | NA Flowsheet | 804 |
| Temperature | Non-Violent Restraints | — |
| Temperature | RETIRED Infusion Center Assessment | — |
| Temperature | Stroke | 66 |
| Temperature | Violent Restraints | — |
| Temperature | Vital Signs | 78 |
| Temperature | Vitals From Sheet | 87 |
| Temperature | Vitals/Interventions | — |

Among 72,714 eligible test encounters, counts identify a nonblank record in the named template during the exact arrival-to-decision interval without a nonblank record for that vital in the two selected templates. Counts may overlap. Presence does not establish a usable value, a confirmed unit, or clinician visibility. To protect patient privacy, counts of 1–10, and selected cells from which they could be calculated, are not shown (—).

| eTable 57. First Vital-Sign Measurements Outside Screening Bounds | | | | | |
| --- | --- | --- | --- | --- | --- |
| Year | Decision group | Vital sign | First measured values | Outside screening bounds | Outside screening bounds percent |
| 2020 | Admission | Heart rate | 15,810 | — | — |
| 2020 | Admission | Systolic blood pressure | 15,699 | 0 | 0.00 |
| 2020 | Admission | Diastolic blood pressure | 15,699 | — | — |
| 2020 | Admission | Respiratory rate | 15,746 | — | — |
| 2020 | Admission | Peripheral oxygen saturation | 15,711 | — | — |
| 2020 | Admission | Temperature | 14,878 | — | — |
| 2020 | Discharge | Heart rate | 39,531 | — | — |
| 2020 | Discharge | Systolic blood pressure | 37,476 | 0 | 0.00 |
| 2020 | Discharge | Diastolic blood pressure | 37,476 | — | — |
| 2020 | Discharge | Respiratory rate | 39,306 | — | — |
| 2020 | Discharge | Peripheral oxygen saturation | 39,393 | — | — |
| 2020 | Discharge | Temperature | 38,178 | 15 | 0.04 |
| 2021 | Admission | Heart rate | 18,495 | — | — |
| 2021 | Admission | Systolic blood pressure | 18,400 | 0 | 0.00 |
| 2021 | Admission | Diastolic blood pressure | 18,400 | — | — |
| 2021 | Admission | Respiratory rate | 18,407 | — | — |
| 2021 | Admission | Peripheral oxygen saturation | 18,386 | — | — |
| 2021 | Admission | Temperature | 17,435 | — | — |
| 2021 | Discharge | Heart rate | 38,954 | — | — |
| 2021 | Discharge | Systolic blood pressure | 36,858 | 0 | 0.00 |
| 2021 | Discharge | Diastolic blood pressure | 36,858 | — | — |
| 2021 | Discharge | Respiratory rate | 38,807 | — | — |
| 2021 | Discharge | Peripheral oxygen saturation | 38,910 | — | — |
| 2021 | Discharge | Temperature | 37,586 | 14 | 0.04 |
| 2022 | Admission | Heart rate | 21,360 | — | — |
| 2022 | Admission | Systolic blood pressure | 21,308 | — | — |
| 2022 | Admission | Diastolic blood pressure | 21,308 | 11 | 0.05 |
| 2022 | Admission | Respiratory rate | 21,245 | — | — |
| 2022 | Admission | Peripheral oxygen saturation | 21,280 | — | — |
| 2022 | Admission | Temperature | 20,173 | — | — |
| 2022 | Discharge | Heart rate | 41,486 | — | — |
| 2022 | Discharge | Systolic blood pressure | 40,527 | 0 | 0.00 |
| 2022 | Discharge | Diastolic blood pressure | 40,527 | — | — |
| 2022 | Discharge | Respiratory rate | 41,347 | — | — |
| 2022 | Discharge | Peripheral oxygen saturation | 41,457 | — | — |
| 2022 | Discharge | Temperature | 39,895 | 12 | 0.03 |
| 2023 | Admission | Heart rate | 23,584 | — | — |
| 2023 | Admission | Systolic blood pressure | 23,529 | 0 | 0.00 |
| 2023 | Admission | Diastolic blood pressure | 23,529 | — | — |
| 2023 | Admission | Respiratory rate | 23,457 | — | — |
| 2023 | Admission | Peripheral oxygen saturation | 23,483 | 0 | 0.00 |
| 2023 | Admission | Temperature | 22,686 | — | — |
| 2023 | Discharge | Heart rate | 47,211 | — | — |
| 2023 | Discharge | Systolic blood pressure | 47,197 | 0 | 0.00 |
| 2023 | Discharge | Diastolic blood pressure | 47,197 | — | — |
| 2023 | Discharge | Respiratory rate | 47,109 | — | — |
| 2023 | Discharge | Peripheral oxygen saturation | 47,112 | — | — |
| 2023 | Discharge | Temperature | 46,033 | 13 | 0.03 |

Screening ranges are defined in eMethods. Out-of-range first measurements remained missing; later measurements were not substituted. To protect patient privacy, counts of 1–10, and selected cells from which they could be calculated, are not shown (—).

| eTable 58. Encounters Without a Usable Predecision Vital Sign | | | | |
| --- | --- | --- | --- | --- |
| Year | Decision group | Encounters | No usable pre-plan vital | Percent |
| 2020 | Admission | 16,932 | 921 | 5.44 |
| 2020 | Discharge | 42,310 | 2,569 | 6.07 |
| 2021 | Admission | 19,485 | 800 | 4.11 |
| 2021 | Discharge | 40,935 | 1,804 | 4.41 |
| 2022 | Admission | 22,505 | 914 | 4.06 |
| 2022 | Discharge | 43,366 | 1,707 | 3.94 |
| 2023 | Admission | 24,401 | 636 | 2.61 |
| 2023 | Discharge | 48,313 | 974 | 2.02 |

No usable first value across the six vital signs does not necessarily mean no vital was recorded. Encounters remained in the cohort with missing-value indicators.

| eTable 59. Resident-Marker Direction and Time-Zero Direction | | |
| --- | --- | --- |
| Comparison | Resident discharge | Resident admission |
| Same as time-zero direction | 72,789 | 22,828 |
| Different from time-zero direction | 719 | 1,882 |
| No usable decision time | 639 | 203 |
| Discharge decision with later reversal | 930 | 401 |

All 2020–2023 resident-plan encounters. The time-zero decision may precede the resident marker. Different-direction percentages use same plus different as denominator, excluding encounters with no usable decision time and discharge decisions with later reversal: 0.98% for resident discharge and 7.62% for resident admission. These are not subsequent reversal rates.

| eTable 60. Resident-Discharge Markers Relative to Discharge Decisions | | | |
| --- | --- | --- | --- |
| Year | Resident discharge plan | Encounters | Minutes to decision, median (IQR) |
| 2020 | After the discharge decision | 413 | −12.0 (−64.0 to −3.0) |
| 2020 | Before the discharge decision | 14,257 | 6.0 (2.0 to 13.0) |
| 2020 | No resident discharge plan | 26,872 | Not applicable |
| 2020 | At the discharge decision time | 1,105 | 0.0 (0.0 to 0.0) |
| 2021 | After the discharge decision | 242 | −28.5 (−84.8 to −3.0) |
| 2021 | Before the discharge decision | 16,437 | 6.0 (2.0 to 15.0) |
| 2021 | No resident discharge plan | 25,190 | Not applicable |
| 2021 | At the discharge decision time | 1,039 | 0.0 (0.0 to 0.0) |
| 2022 | After the discharge decision | 256 | −30.5 (−100.0 to −5.0) |
| 2022 | Before the discharge decision | 18,556 | 7.0 (3.0 to 16.0) |
| 2022 | No resident discharge plan | 25,565 | Not applicable |
| 2022 | At the discharge decision time | 994 | 0.0 (0.0 to 0.0) |
| 2023 | After the discharge decision | 229 | −29.0 (−97.0 to −4.0) |
| 2023 | Before the discharge decision | 19,167 | 7.0 (3.0 to 17.0) |
| 2023 | No resident discharge plan | 28,452 | Not applicable |
| 2023 | At the discharge decision time | 1,024 | 0.0 (0.0 to 0.0) |
| All years | After the discharge decision | 1,140 | −20.0 (−86.2 to −4.0) |
| All years | Before the discharge decision | 68,417 | 7.0 (3.0 to 15.0) |
| All years | No resident discharge plan | 106,079 | Not applicable |
| All years | At the discharge decision time | 4,162 | 0.0 (0.0 to 0.0) |

Discharge-decision population. Negative intervals mean the resident marker came after the time-zero decision. Resident discharge never set time zero. Quantiles require at least 11 contributing encounters. To protect patient privacy, counts of 1–10, and selected cells from which they could be calculated, are not shown (—).

| eTable 61. Characteristics of Eligible Encounters and Encounters Excluded for an Unknown Composite Outcome | | |
| --- | --- | --- |
| Characteristic | Eligible encounters | Excluded: unknown composite outcome |
| Cohort |  |  |
| Encounters | 258,247 | 2,962 |
| Unique patients | 136,567 | 2,521 |
| Arrival year |  |  |
| 2020 | 59,242 (22.9) | 423 (14.3) |
| 2021 | 60,420 (23.4) | 643 (21.7) |
| 2022 | 65,871 (25.5) | 1,221 (41.2) |
| 2023 | 72,714 (28.2) | 675 (22.8) |
| Age, y |  |  |
| Median (IQR) | 50 (33–68) | 44 (30–61) |
| Available | 258,247 (100.0) | 2,962 (100.0) |
| Missing | 0 (0.0) | 0 (0.0) |
| Sex |  |  |
| Female | 139,594 (54.1) | 1,422 (48.0) |
| Male | — | — |
| Missing | 0 (0.0) | 0 (0.0) |
| Unknown | — | — |
| Race |  |  |
| American Indian or Alaska Native | 632 (0.2) | — |
| Asian | 40,228 (15.6) | 436 (14.7) |
| Black or African American | 16,114 (6.2) | 268 (9.0) |
| Multiple recorded labels | 7,527 (2.9) | 125 (4.2) |
| Native Hawaiian or Other Pacific Islander | 5,419 (2.1) | 47 (1.6) |
| Other | 84,959 (32.9) | 852 (28.8) |
| White | 99,925 (38.7) | 1,034 (34.9) |
| White, non-Hispanic | — | — |
| Declines to state | 3,055 (1.2) | — |
| Missing | 215 (0.1) | — |
| Unknown | — | — |
| Ethnicity |  |  |
| Hispanic/Latino | 75,735 (29.3) | 745 (25.2) |
| Non-Hispanic/non-Latino | 179,722 (69.6) | 2,016 (68.1) |
| Declines to state | 2,128 (0.8) | — |
| Missing | 182 (0.1) | — |
| Unknown | 480 (0.2) | — |
| Language |  |  |
| English | 207,757 (80.4) | 2,468 (83.3) |
| Farsi | 1,101 (0.4) | — |
| Mandarin | 3,635 (1.4) | 27 (0.9) |
| Spanish | 34,252 (13.3) | 313 (10.6) |
| Vietnamese | 3,062 (1.2) | — |
| Other or missing | 8,440 (3.3) | 133 (4.5) |
| Interpreter need |  |  |
| Not required | 213,328 (82.6) | 2,504 (84.5) |
| Required | 44,813 (17.4) | — |
| Missing | 106 (0.0) | — |
| ESI |  |  |
| 1 | 1,704 (0.7) | 95 (3.2) |
| 2 | 55,477 (21.5) | 319 (10.8) |
| 3 | 151,483 (58.7) | 1,460 (49.3) |
| 4 | 45,006 (17.4) | 794 (26.8) |
| 5 | — | — |
| Missing | — | — |
| Arrival mode |  |  |
| Car/bus | 207,829 (80.5) | 2,335 (78.8) |
| Walk | 2,659 (1.0) | 44 (1.5) |
| Ambulance, American Medical Response | 29,225 (11.3) | 336 (11.3) |
| Ambulance, Palo Alto Paramedics | 8,039 (3.1) | 127 (4.3) |
| Ambulance | 3,131 (1.2) | 39 (1.3) |
| Other or missing | 7,364 (2.9) | 81 (2.7) |

Encounters with a recorded discharge decision or admission plan, 2020–2023. Eligible encounters met all eligibility criteria; the comparison column contains encounters excluded because the composite outcome was unknown. Encounters excluded only by temporal eligibility criteria are not shown. Repeat visits are retained; characteristics are encounter-level summaries and patients may appear in both columns. Values are No. (%) unless stated otherwise; percentages include missing records in the denominator. Age uses available recorded values and linear-interpolated quartiles. Language and arrival-mode categories follow Table 1: the five most frequent training languages and arrival modes are shown separately, with remaining categories and missing values combined as Other or missing. Arrival modes are recorded electronic health record categories; ambulance categories are shown separately as recorded. Decision-group counts are not shown because they would allow recovery of withheld cells in other tables. To protect patient privacy, counts of 1–10, and selected cells from which they could be calculated, are not shown (—). A dash does not indicate zero or a missing record. Descriptive comparison only; no tests were performed. Abbreviations: IQR, interquartile range; ESI, Emergency Severity Index.

eFigure 1. Study Workflow and Temporal Evaluation

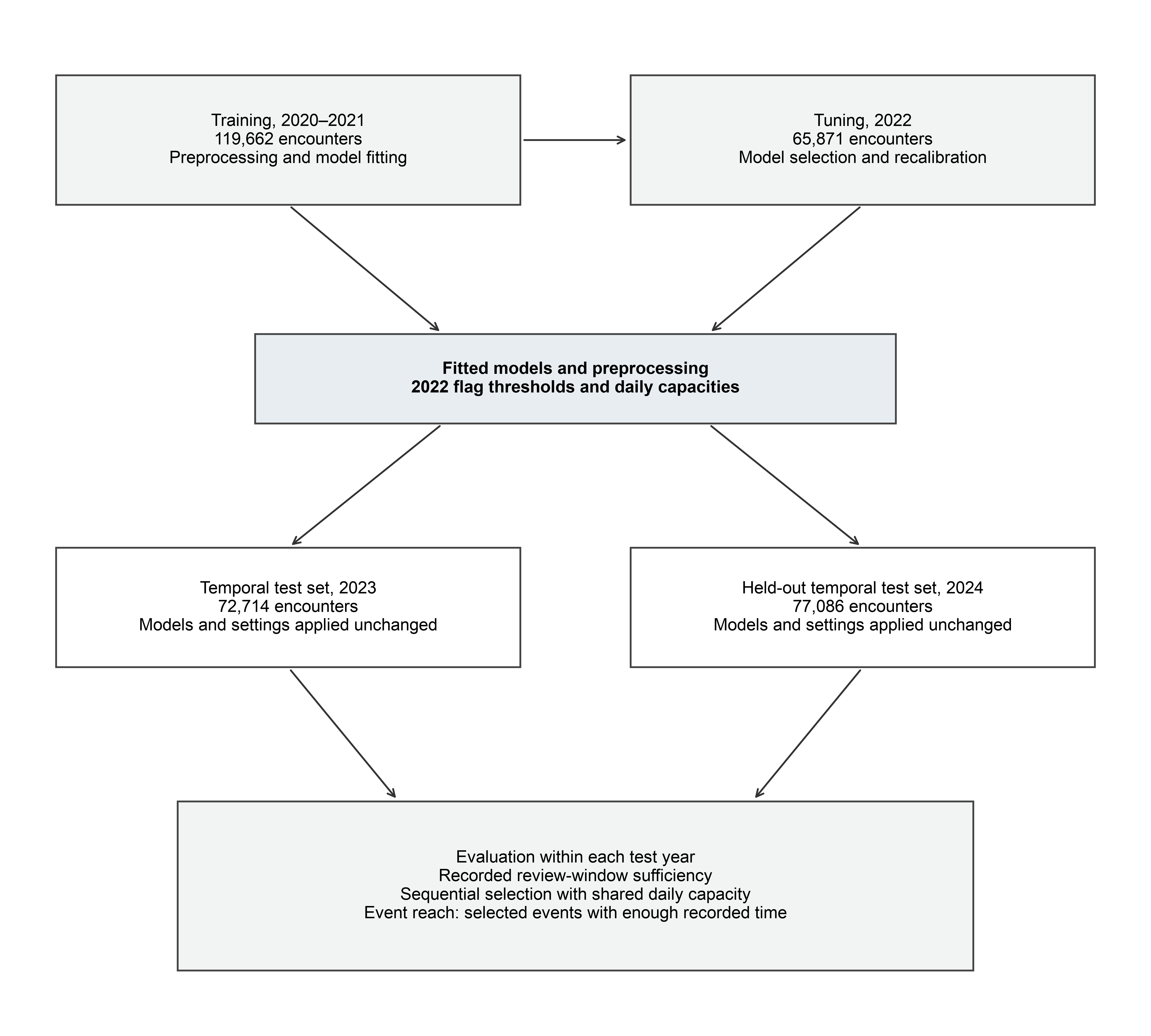

Training and tuning determine preprocessing, fitted models, recalibration, flag thresholds, and daily capacities. The 2023 temporal test set and 2024 held-out temporal test set use those fitted models and settings. Period counts are eligible encounters. Within each test year, event reach combines simulated selection with sufficient recorded time; it does not measure events prevented. Admission plans provide a retrospective contrast to discharge decisions.

eFigure 2. Predictor Missingness

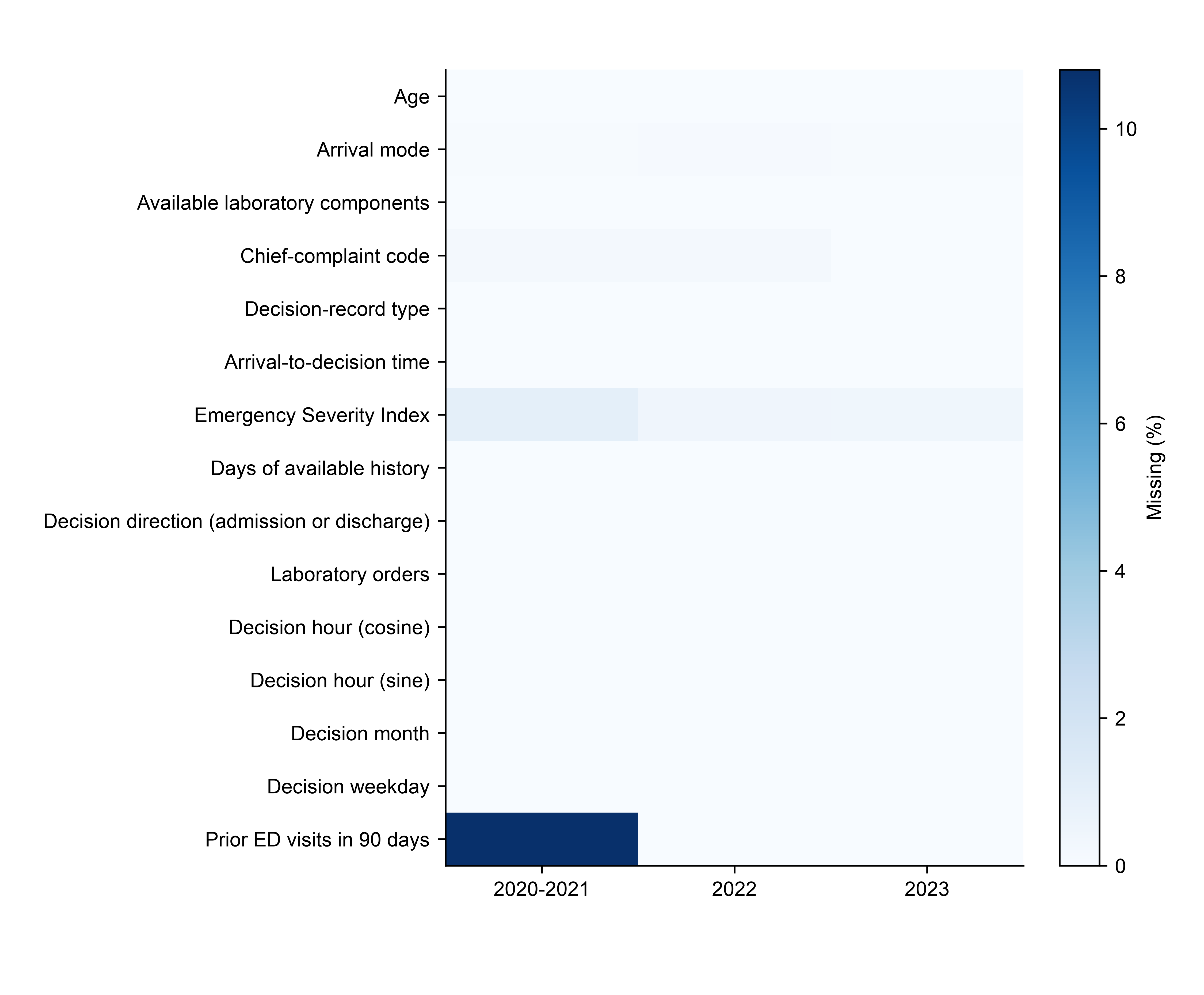

2020–2023. Percent missing among retained recorded-decision encounters, including unknown outcomes. Labels match the predictor table. The direction field is constant within each separately fitted model.

eFigure 3. Calibration in the 2023 Temporal Test Set

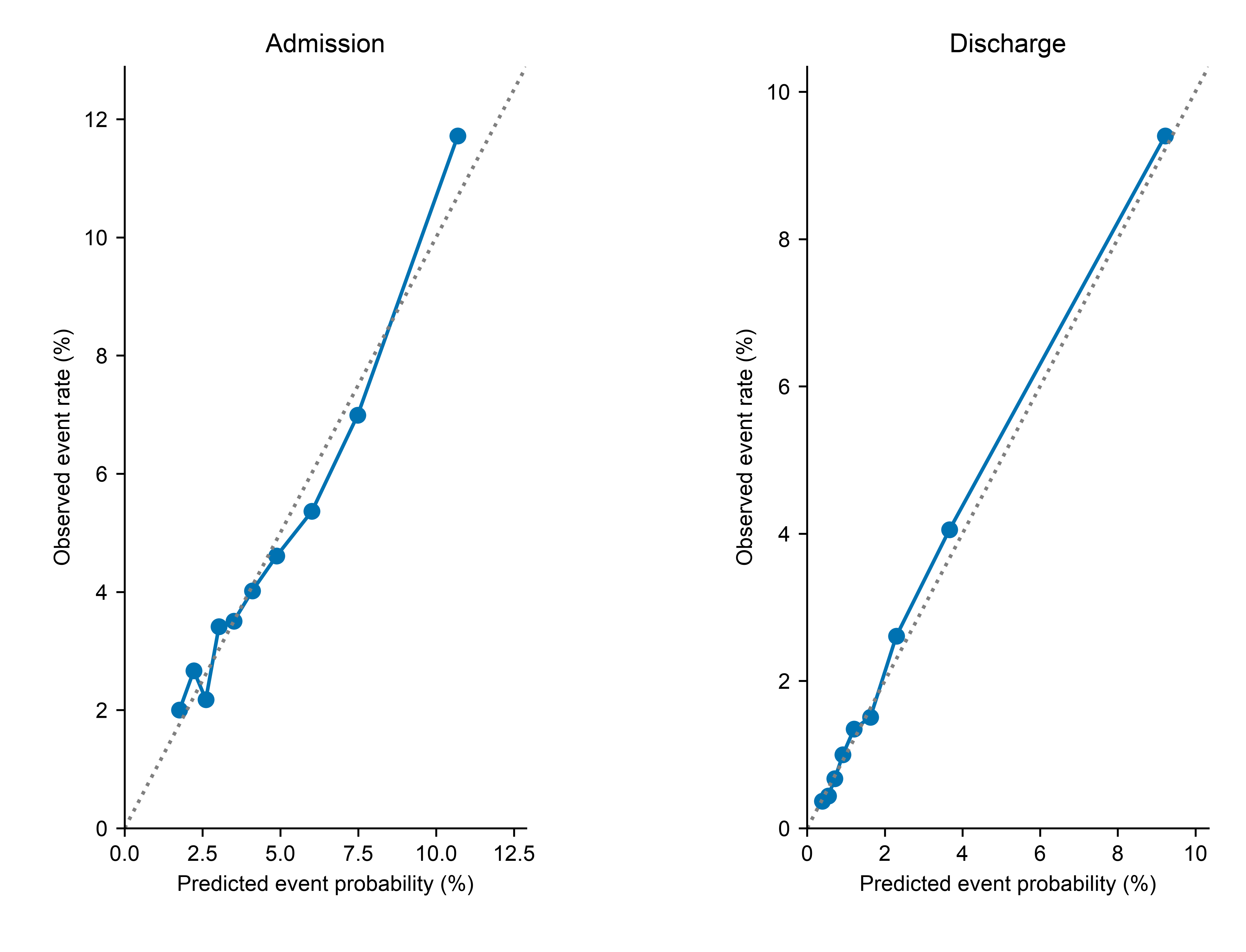

The composite comprised an adult ED return arriving within 7 days of time zero and ultimately ending in inpatient admission, qualifying critical care within 24 hours, or death within 7 days. Risk groups use 2022 recalibrated prediction deciles. The diagonal denotes equality of predicted and observed rates.

eFigure 4. Recorded Windows Sufficient for Review in 2023

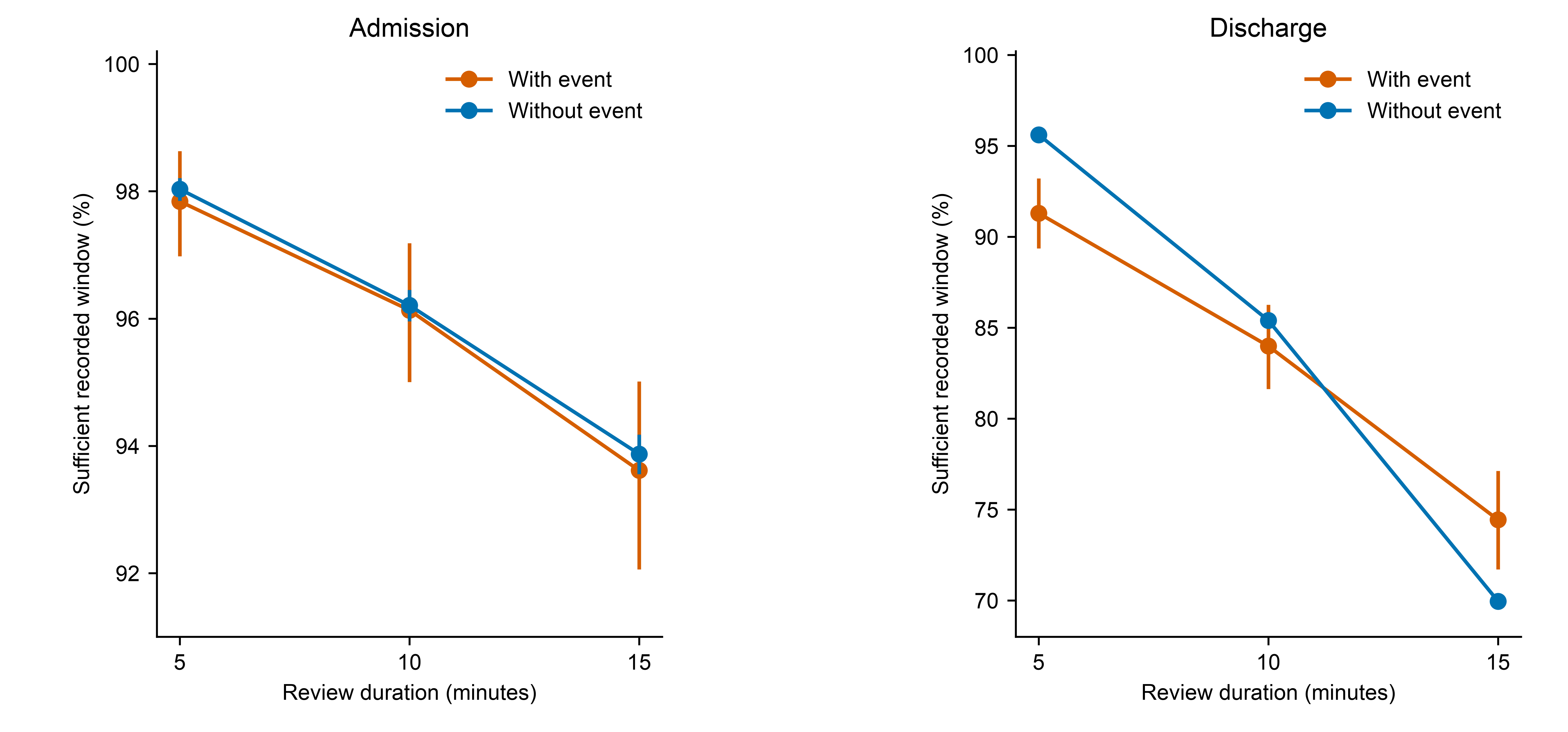

Eligible 2023 encounters with known outcomes. Points show the proportion with a recorded window at least as long as the review; bars are 95% patient-cluster intervals. Panel scales differ.

eFigure 5. Calibration in the 2024 Held-Out Temporal Test Set

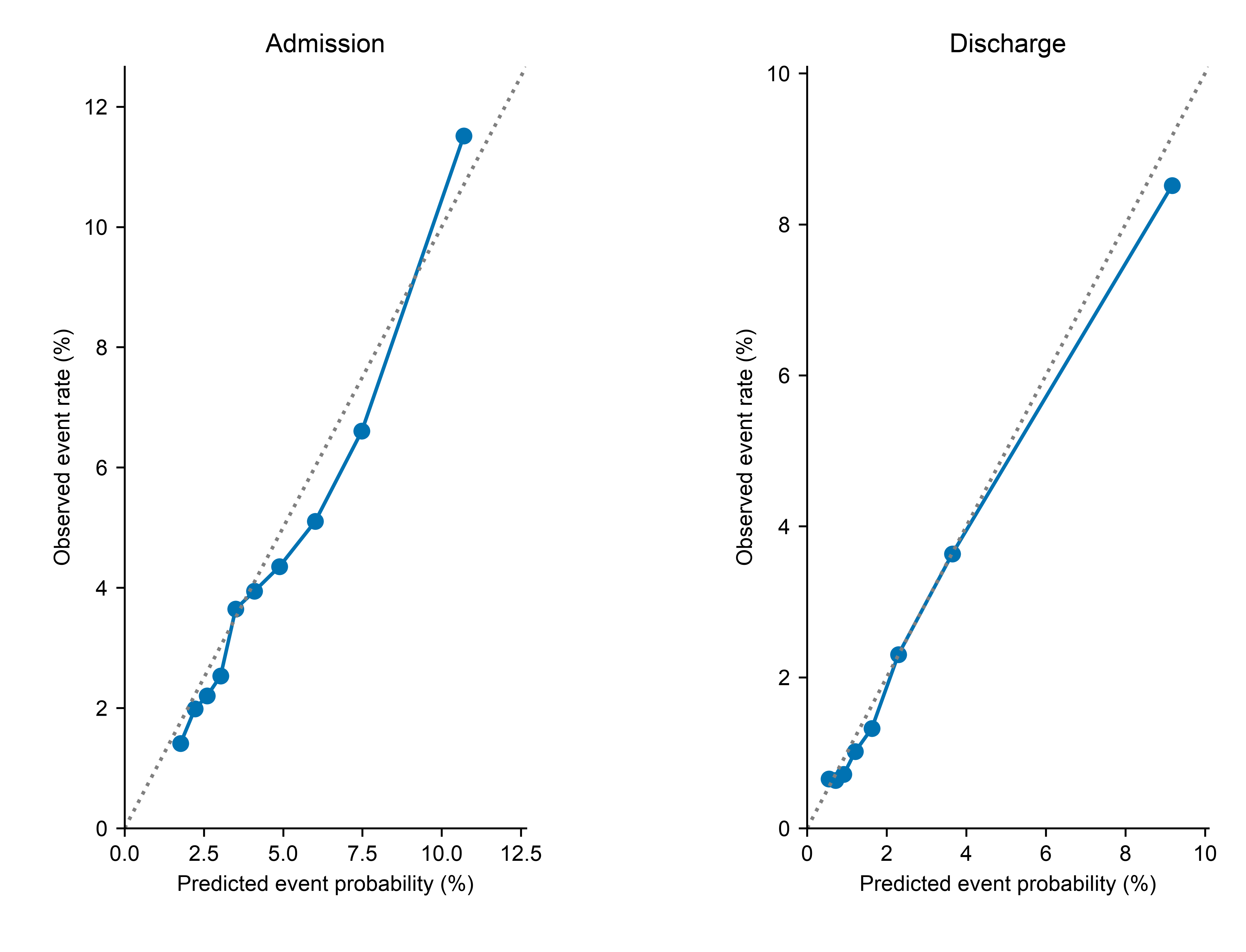

Primary risk models, with intercept recalibration estimated in 2022 and applied unchanged in 2024. Bins use 2022 prediction deciles. Points represent bins of predicted risk; connecting segments are descriptive. Bins withheld under the small-cell rule are not plotted. No bin-specific intervals were estimated. The composite comprised return admission within 7 days, qualifying critical care within 24 hours, or death within 7 days of time zero.

eFigure 6. Coverage of Full-Day and Annual Ranking Intervals in 2023

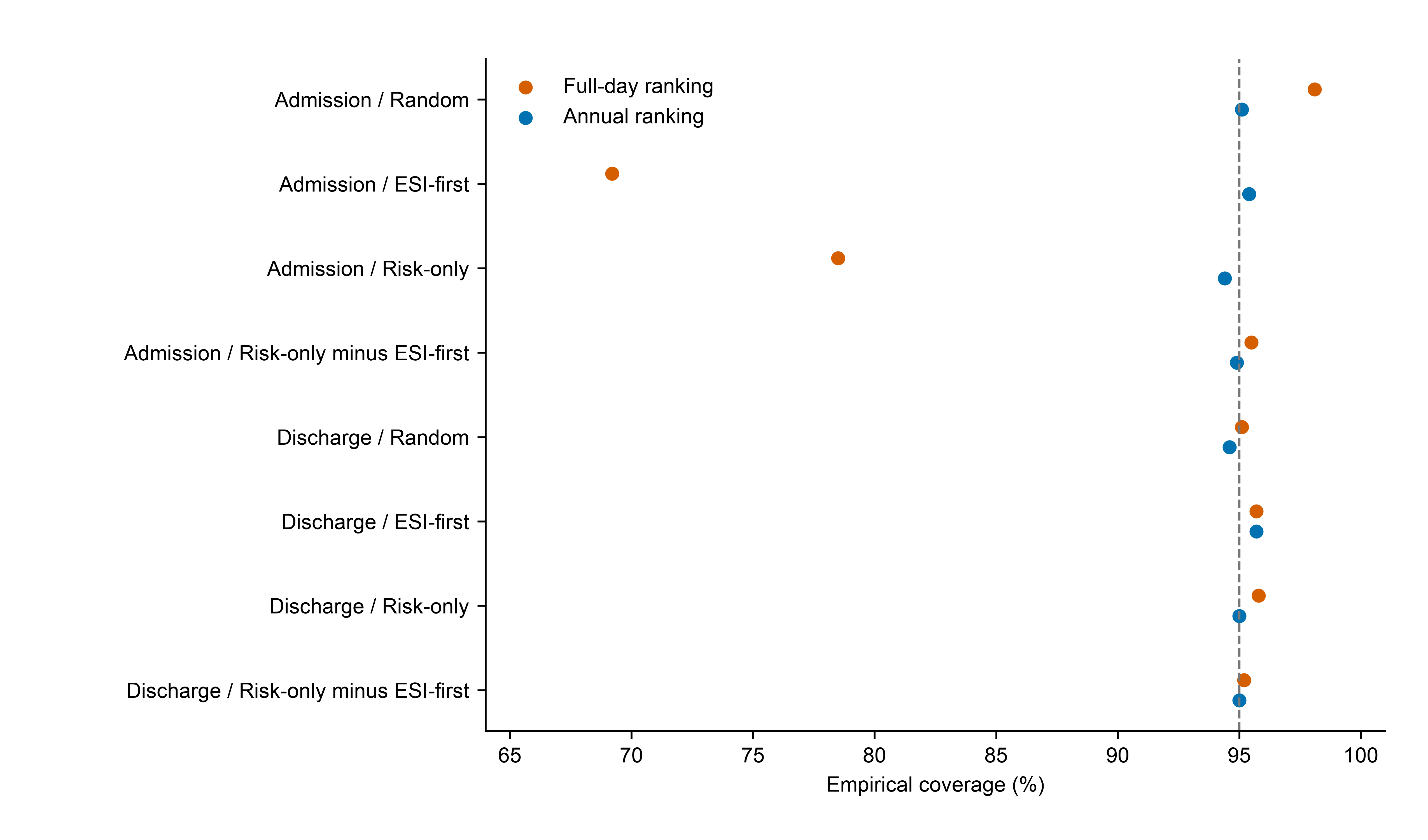

Nested diagnostic; dashed line marks nominal 95%. Empirical coverage does not establish population coverage. Values concern full-day and annual ranking, not sequential selection.

eFigure 7. Full-Day Ranking With Clinical Comparators in 2023

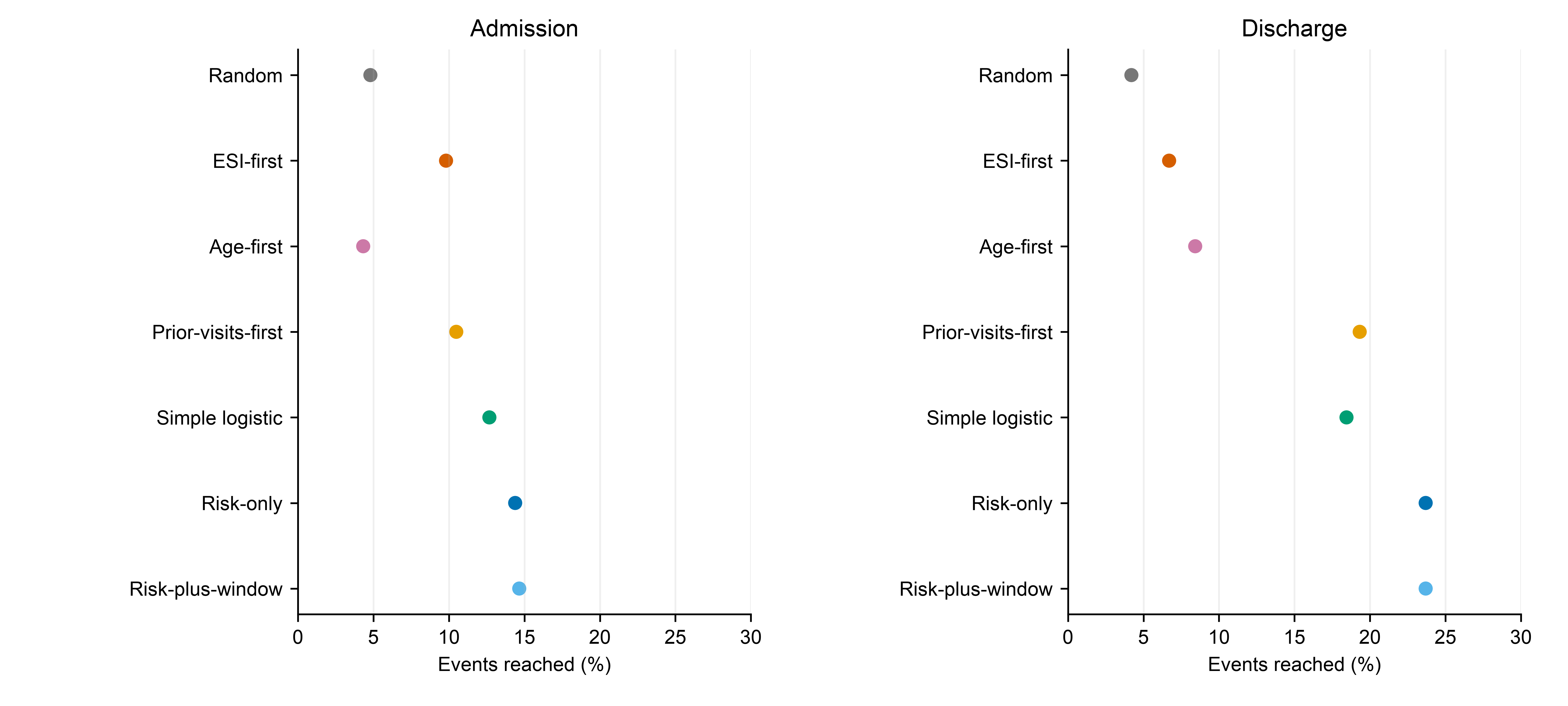

Ten-minute reviews and 50 review-minutes per 100 encounters. Full-day ranking; descriptive point estimates. Event reach is the proportion of all composite-event encounters selected with sufficient recorded review time. Review attempts without enough recorded time still count against the budget. Reach does not measure events prevented. Simple logistic indicates the five-variable logistic model. ESI indicates Emergency Severity Index.

eFigure 8. Full-Day Ranking Event Reach in 2023

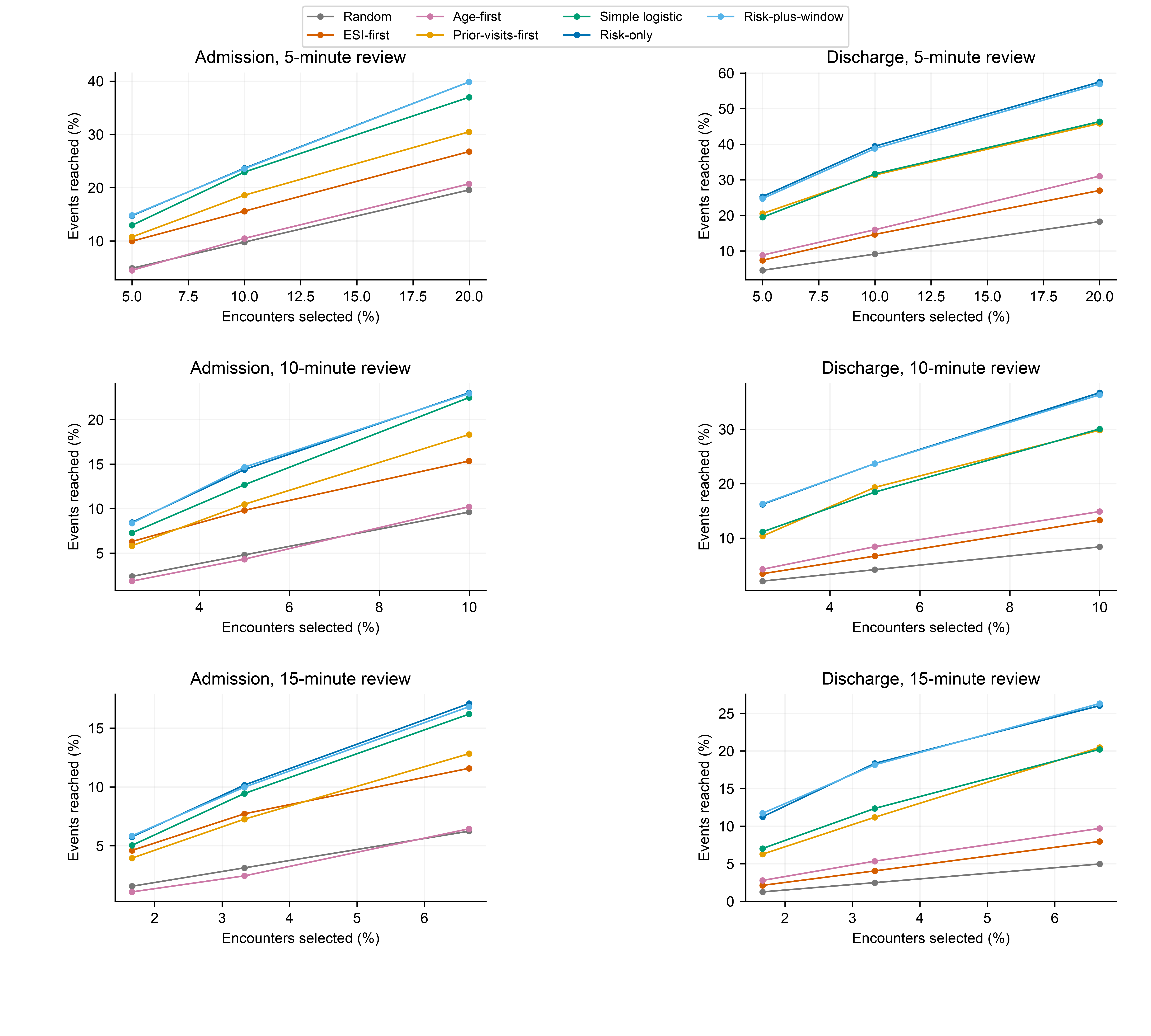

Retrospective full-day ranking benchmarks. Points are descriptive; lines connect evaluated budgets only. Event reach is the proportion of all composite-event encounters selected with sufficient recorded review time. Review attempts without enough recorded time still count against the budget. Reach does not measure events prevented. Simple logistic indicates the five-variable logistic model. ESI indicates Emergency Severity Index.

eFigure 9. Full-Day Ranking Lift in 2023

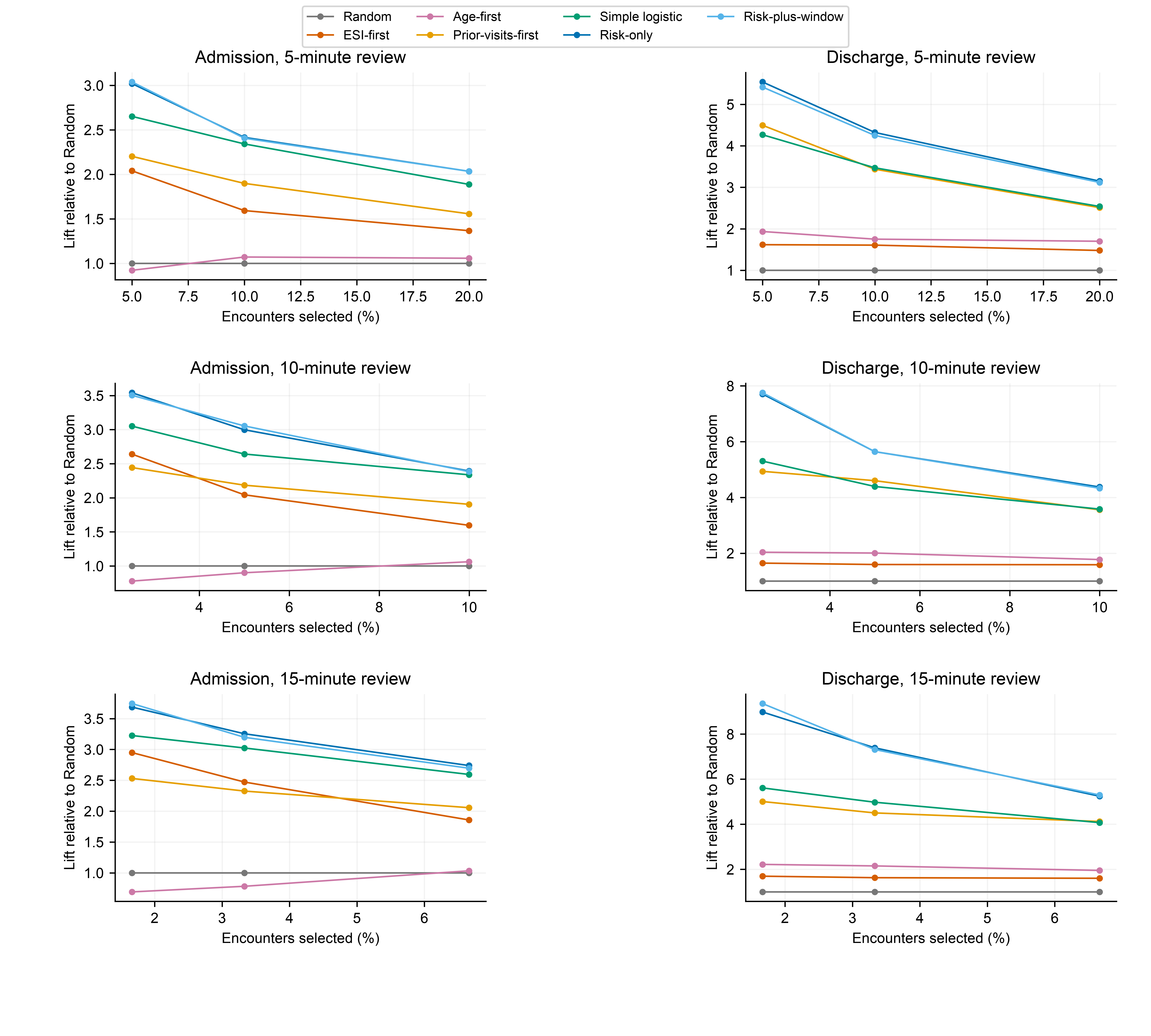

Retrospective full-day ranking benchmarks. Points are descriptive; lines connect evaluated budgets only. Event reach is the proportion of all composite-event encounters selected with sufficient recorded review time. Review attempts without enough recorded time still count against the budget. Reach does not measure events prevented. Simple logistic indicates the five-variable logistic model. ESI indicates Emergency Severity Index.

eFigure 10. Annual Ranking Event Reach in 2023

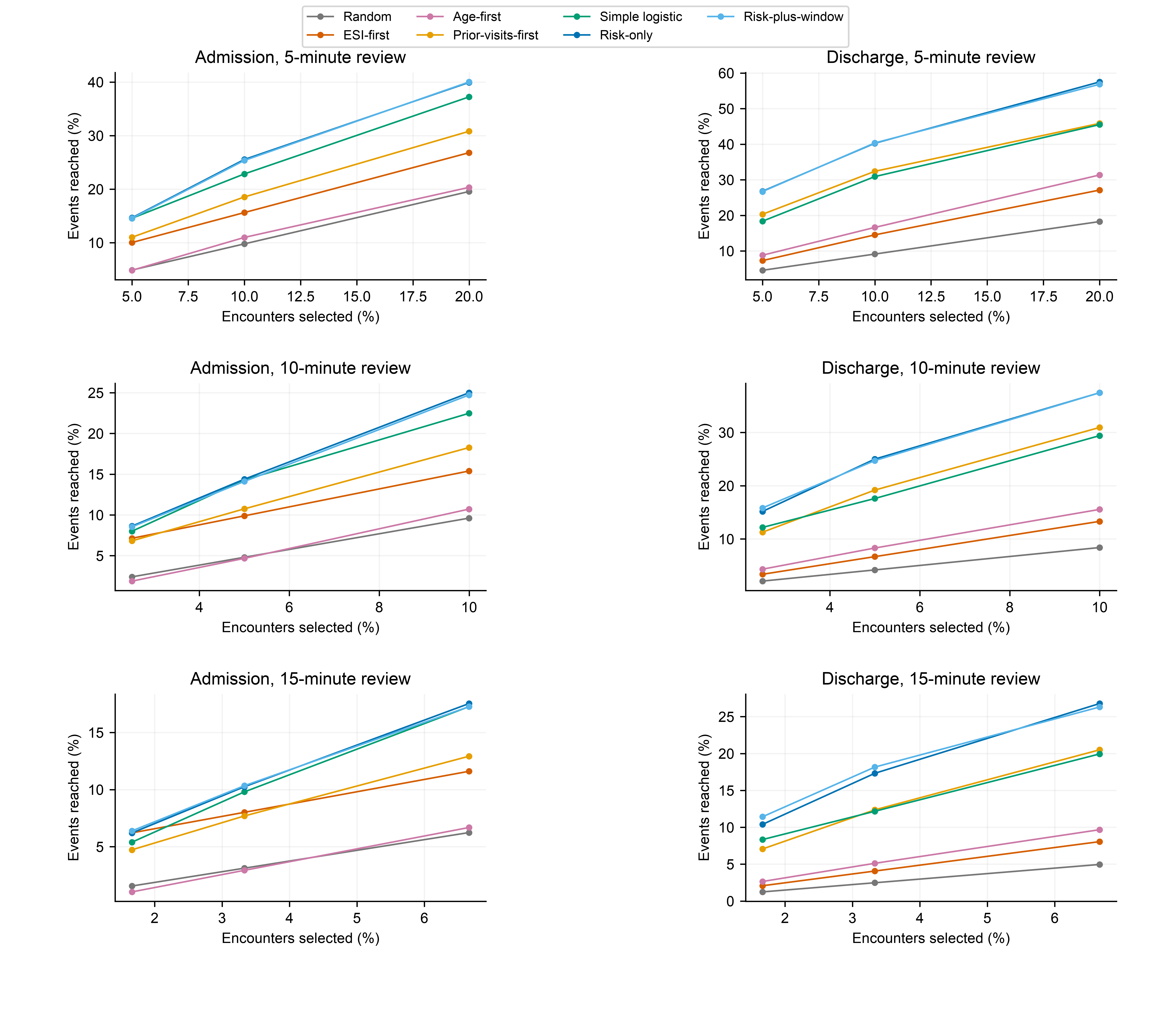

Retrospective annual ranking benchmarks. Points are descriptive; lines connect evaluated budgets only. Event reach is the proportion of all composite-event encounters selected with sufficient recorded review time. Review attempts without enough recorded time still count against the budget. Reach does not measure events prevented. Simple logistic indicates the five-variable logistic model. ESI indicates Emergency Severity Index.

eFigure 11. Annual Ranking Lift in 2023

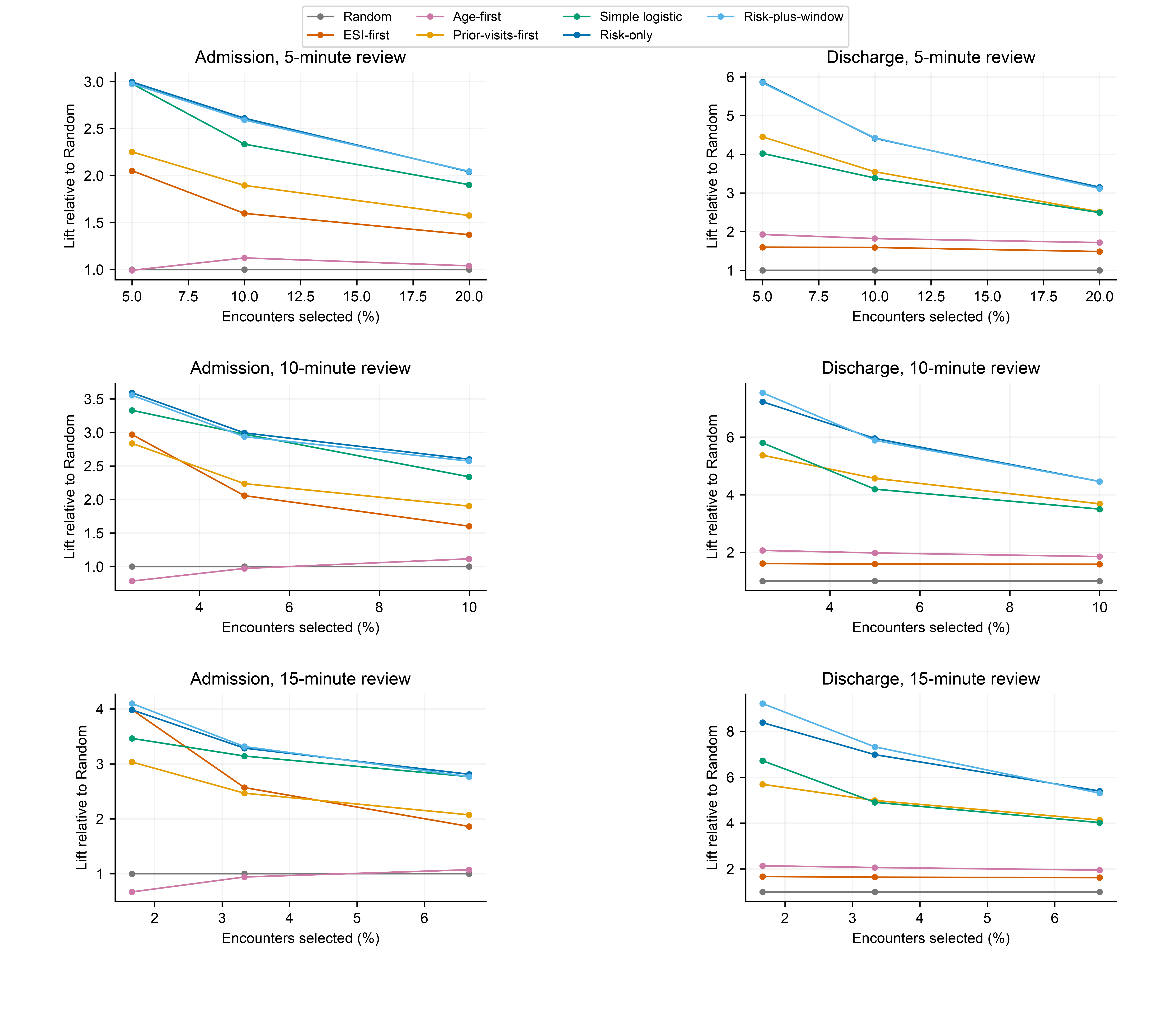

Retrospective annual ranking benchmarks. Points are descriptive; lines connect evaluated budgets only. Event reach is the proportion of all composite-event encounters selected with sufficient recorded review time. Review attempts without enough recorded time still count against the budget. Reach does not measure events prevented. Simple logistic indicates the five-variable logistic model. ESI indicates Emergency Severity Index.

eFigure 12. Recorded Windows Before Model Exclusions

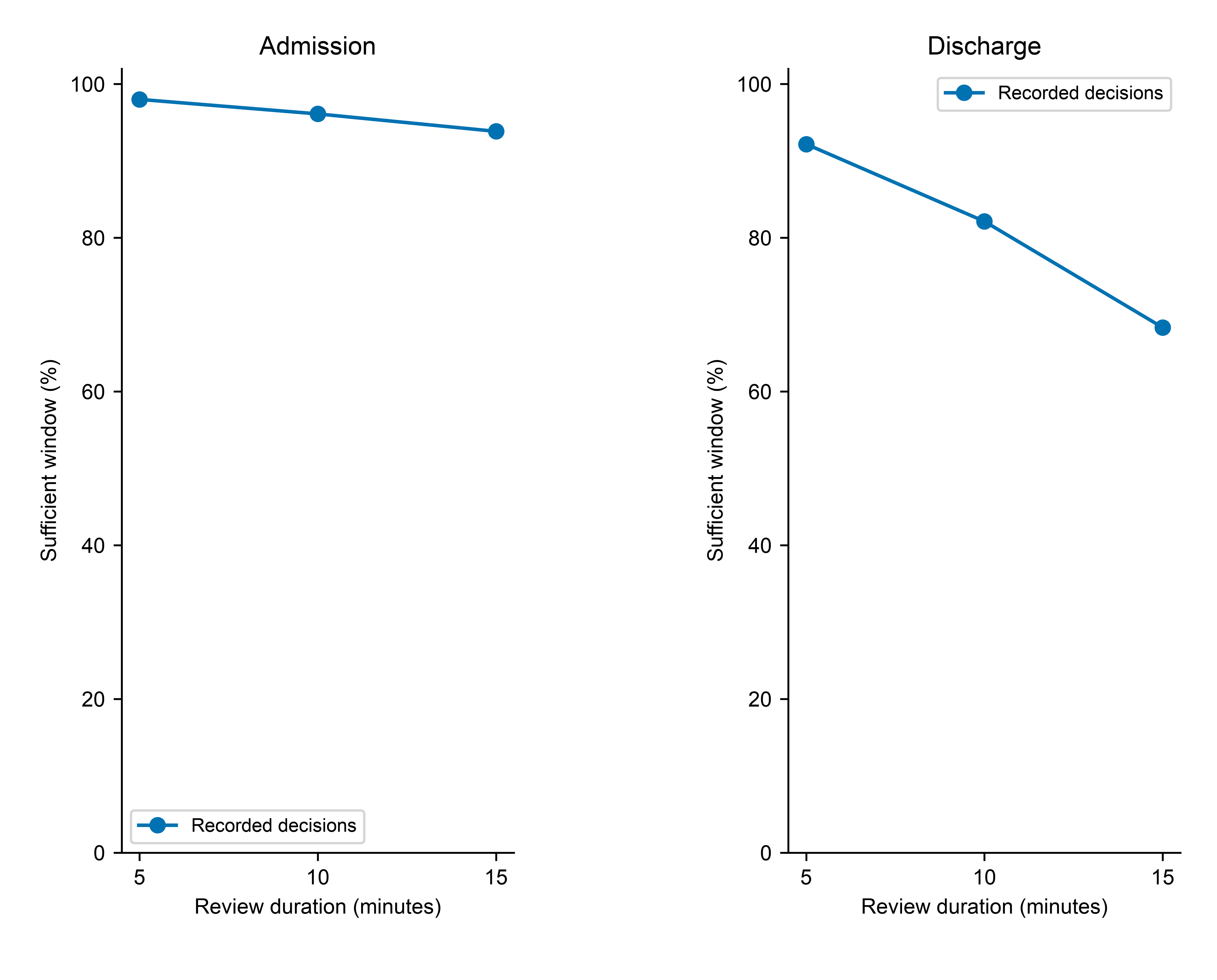

Pooled 2020–2023 recorded-decision encounters before known-outcome and temporal exclusions: 84,679 admission and 179,798 discharge encounters. These denominators differ from the model cohort.

eFigure 13. Window Length and Outcome Association in 2023

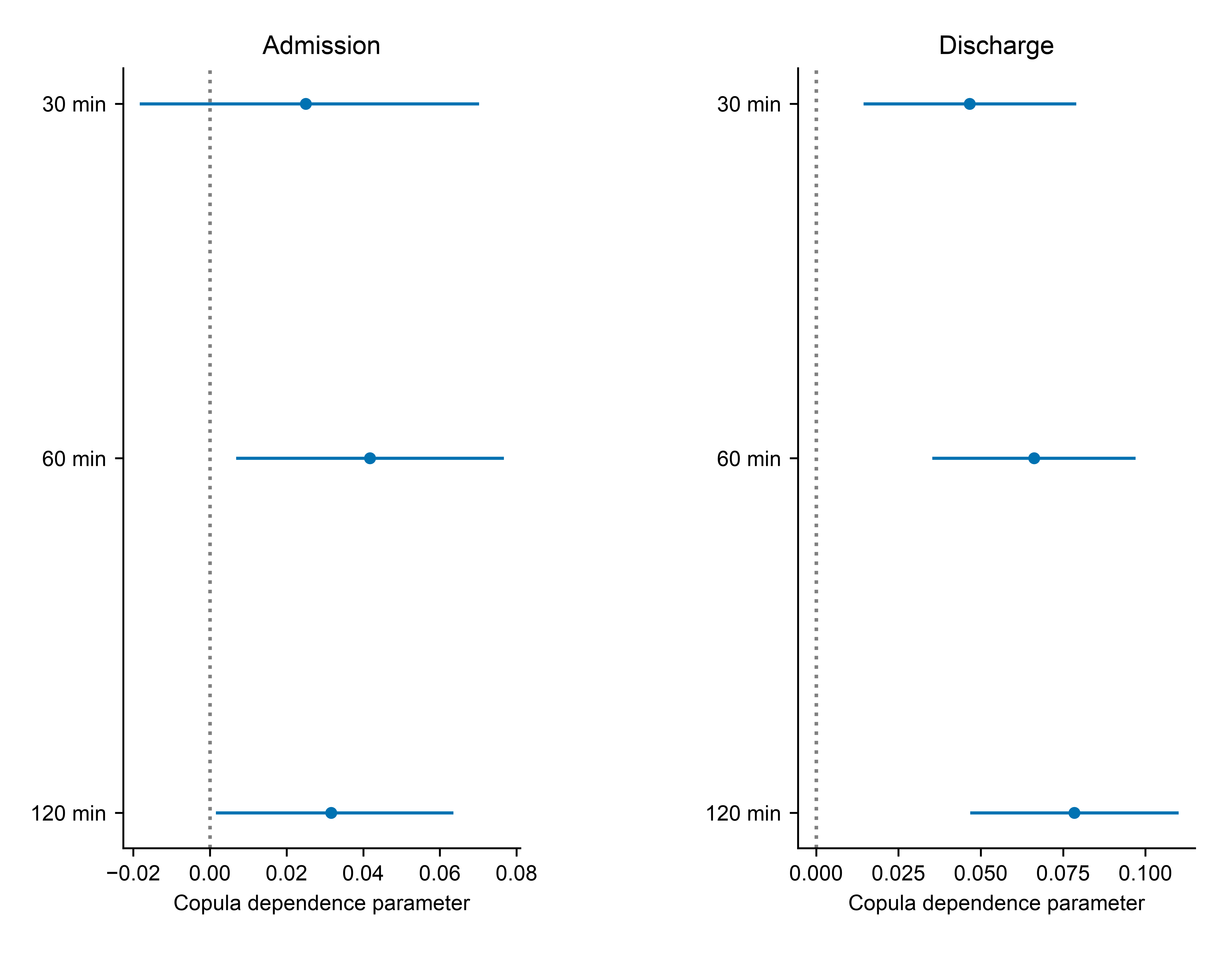

Gaussian copula estimates at 30-, 60-, and 120-minute caps. Bars are patient-cluster intervals. This is model-based association, not causation.
